# Coupling of metabolomics and exome sequencing reveals graded effects of rare damaging heterozygous variants on gene function and resulting traits and diseases

**DOI:** 10.1101/2023.10.17.23297094

**Authors:** Nora Scherer, Daniel Fässler, Oleg Borisov, Yurong Cheng, Pascal Schlosser, Matthias Wuttke, Suraj Patil, Heike Meiselbach, Fabian Telkämper, Urs Berger, Sarah Grünert, Peggy Sekula, Ulla T. Schultheiss, Yong Li, Michael Köttgen, Peter J. Oefner, Felix Knauf, Kai-Uwe Eckardt, Ines Thiele, Miriam Schmidts, Johannes Hertel, Anna Köttgen

## Abstract

Genetic studies of the metabolome can uncover enzymatic and transport processes shaping human metabolism. Using WES-based rare variant aggregation testing to detect genes associated with levels of 1,294 plasma and 1,396 urine metabolites, we discovered 235 gene-metabolite associations, many previously unreported. Validation through genetic and new computational approaches (*in silico* gene knockouts in whole-body models of human metabolism) provided orthogonal evidence that population-based studies of rare, damaging variants in the heterozygous state permit inferences usually obtained from inborn errors of metabolism. Allelic series of functional variants in transporters responsible for transcellular sulfate reabsorption (SLC13A1, SLC26A1) exhibited graded effects on plasma sulfate and human height, and pinpointed alleles that strongly increased risk for dozens of musculoskeletal traits and diseases in the population. We present a powerful approach to identify new players in incompletely characterized human metabolic reactions, and to reveal metabolic readouts of disease risk to inform disease prevention and treatment.

## Introduction

A complex interplay of thousands of enzymes and transport proteins is involved in maintaining physiological levels of intermediates and end-products of metabolism. Disturbances of their function can result in severe disease, such as those caused by inborn errors of metabolism (IEMs), or predispose to common metabolic diseases such as type 2 diabetes or gout. While the study of rare, early onset, autosomal-recessive IEMs has uncovered many metabolite-related genes, such studies are limited by the very low number of persons homozygous for the causative variants. Genome-wide association studies (GWAS) in large study populations on the other hand have revealed thousands of common genetic variants that are associated with altered metabolite levels^1–13^. However, identified loci typically contain many genes and variants are often non-coding, making it challenging to identify the causal gene.

Gene-based aggregation testing of rare, putatively damaging variants in population studies can address this challenge. Previously, such studies have focused almost exclusively on the circulating metabolome^14–20^. We have recently shown that GWAS of paired plasma and urine metabolomes not only reveal many more associations, but also enable specific insights into renal metabolite handling^2^. We therefore aimed to perform gene-based testing of the aggregate effect of rare variants on the levels of 1,294 plasma and 1,396 urine metabolites quantified from 4,737 participants of the German Chronic Kidney Disease (GCKD) study with whole-exome sequencing (WES) data, in order to identify metabolism-related genes and to understand whether the underlying rare, almost exclusively heterozygous variants permit inferences otherwise only obtained from the study of IEMs.

Patients with IEMs typically show severe symptoms that originate from an accumulation or depletion of metabolites, while heterozygous carriers of the causative variants often show milder changes of the same or related metabolic phenotypes^21^. We hypothesized that sex-specific analysis of metabolite-associated, X-chromosomal genes as well as knowledge-based, computational modeling based on sex-specific organ-resolved whole-body models (WBMs^22^; Methods) of human metabolism can inform on whether heterozygous damaging variants capture the metabolic effects of their unobserved homozygous counterparts. WBMs enable the investigation of homozygous gene defects through deterministic *in silico* knockout modeling. The resulting virtual IEMs reflect observed IEMs^22–24^. We further hypothesized that metabolite-associated rare variants identified in the GCKD study would show associations with related traits and diseases in very large population studies, and that the genetic effects would be proportional to their effects on metabolite levels if the implicated metabolites reflect the degree of the encoded proteins’ or pathways’ functional impairment and thereby are a molecular readout of disease-relevant processes. The UK Biobank (UKB), a very large population study with WES data and extensive health record linkage, permits the systematic study of the aggregated and individual effects of rare, damaging, metabolite-associated variants on a wide variety of traits and diseases.

Here, we set out to perform gene-based rare variant aggregation testing to discover genes associated with metabolite levels and to characterize their genetic architecture across the allele frequency spectrum and across plasma and urine, to validate identified genes and variants and the range of their effects through genetic and novel computational approaches based on WBMs that we make publicly available, and to identify traits and diseases for which these metabolites represent molecular readouts to aid drug development.

## Results

As summarized in **Figure 1**, rare, putatively damaging variants that qualified for gene-based testing (Methods) were identified in 16,525 genes based on WES data from 4,737 GCKD study participants (mean age 60 years, 40% women; **Supplementary Table 1**). Metabolites were quantified by non-targeted mass spectrometry and covered a wide variety of metabolic super-pathways (**Supplementary Table 2**). Genome-wide burden tests for the association between each gene and the levels of each of 1,294 plasma and 1,396 urine metabolites (781 overlapping) were carried out using two complementary approaches to select qualifying variants (QVs) for gene-based testing into “masks”. Both masks assume a loss-of-function mechanism, but account for different genetic architectures (Methods).

**Figure 1:**
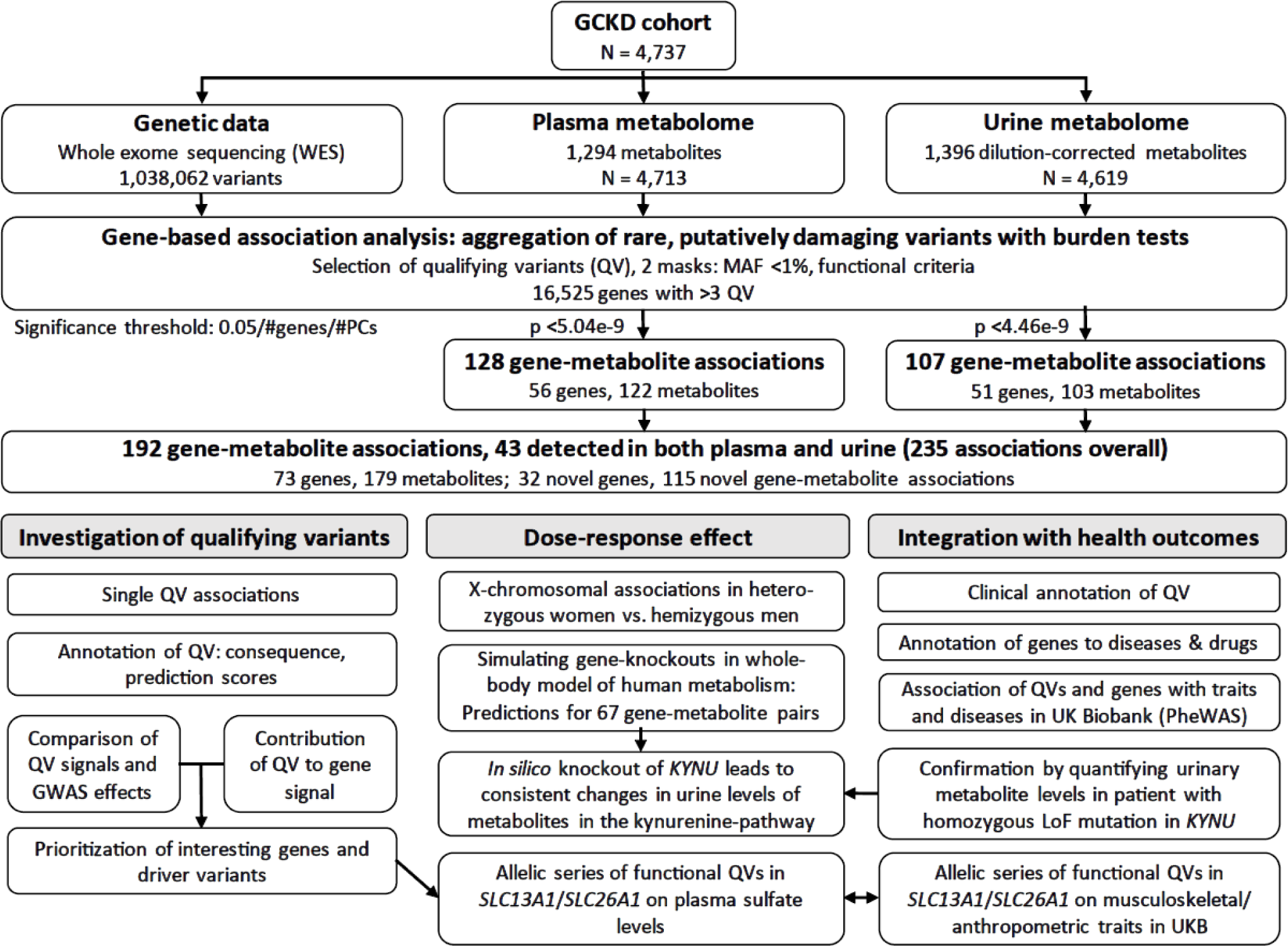
Overview of the study design. Schematic representation of the gene-based rare variant aggregation study with plasma and urine metabolite levels using whole-exome sequencing data of 4,737 participants of the GCKD study and their follow-up analyses.

### Identification and properties of 192 significant gene-metabolite associations

We identified 192 significant gene-metabolite pairs across both plasma (P-value <5.04e-9) and urine (P-value <4.46e-9), where 43 associations were detected in both (192+43 associations overall, **Figure 2a**; **Supplementary Table 3**). The significant associations involved 73 unique genes and 179 metabolites, with a comparable number of genes and metabolites identified in plasma and urine. There were 22 and 17 genes with significant associations exclusively in plasma and in urine, respectively. While the majority of associations was detected with both masks (Methods), the more inclusive mask “HI_mis” yielded more mask-specific associations than the “LoF_mis” mask (**Figure 2b**). The proportion of lipids was substantially higher among associated metabolites detected in plasma compared to urine, consistent with the absence of glomerular filtration of many lipids (**Figure 2b**). Associations detected in both plasma and urine generally affected the levels of the implicated metabolite in the same direction (**Figure 2a**).

**Figure 2:**
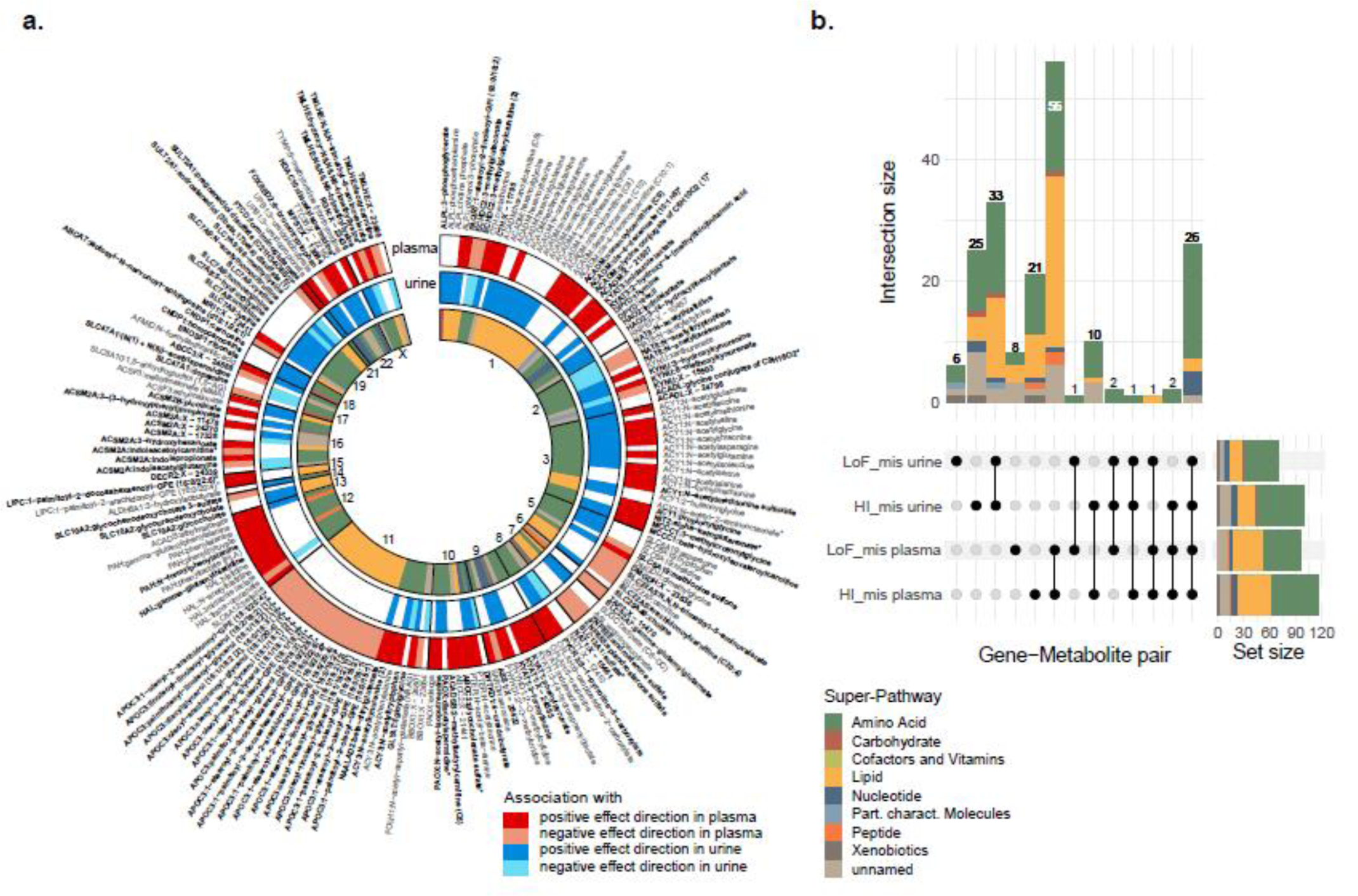
Overview of the 192 identified gene-metabolite associations across plasma and urine and their corresponding pathways. **(a)** Significant associations with plasma metabolites are shown on the outermost band (red; shading reflects effect direction), with genes ordered by chromosomal location by order of genes across the genome. Associations with urine metabolites are shown on the middle band (blue; shading reflects effect direction). Gene-metabolite associations are based on rare variant aggregation testing from both masks. The ones labeled in gray were already reported in previous rare variant studies, whereas the ones labeled in bold black are considered novel. White spaces indicate that no significant association was detected in a given matrix. For all associations detected in both matrices, effect directions are consistent. The inner band represents the super-pathway of the associated metabolite. **(b)** The UpSet plot shows the number of identified gene-metabolite associations by mask and matrix, color-coded by the respective metabolite super-pathway. The horizontal bar plot on the right represents the total number of associations identified by mask and matrix. The proportion of lipids is markedly higher among associations detected with plasma metabolites as compared to urine. The vertical bar plot on the top on the left shows the number of shared associations by mask and matrix, while the sets among which the associations are shared are indicated below each column. While the majority of associations is detected by both masks, especially the less stringent HI_mis mask provides many mask-specific findings in both plasma and urine. The group of metabolites detected in both plasma and urine is dominated by amino acids.

Comparison of our results to those from published studies that focused on rare variant aggregation testing of metabolite levels^14–20,24^ (Methods) showed that 32 of the 73 identified unique genes (44%) had not been reported as significant in any of these studies. Moreover, 115 of all 192 detected gene-metabolite associations (60%) were novel (**Supplementary Table 4**).

Of the 73 metabolite-associated genes, 8 (11%) are targets of approved or currently developed drugs, and 28 (38%) are currently known to harbor causative mutations for IEMs (**Supplementary Table 4**). In our study of middle-aged and older individuals, QVs were almost exclusively observed in the heterozygous state, as illustrated by comparisons of metabolite levels between QV carriers and non-carriers (**Supplementary Figures 1 and 2** for plasma and urine gene-metabolite pairs, respectively). Detailed annotation of each QV in both masks showed that 63 unique QVs in 15 genes and 73 unique QVs in 17 genes were listed in the ClinVar database as “pathogenic” or “pathogenic or likely pathogenic” for a corresponding monogenic disease. These observations support that gene-based aggregation of rare, heterozygous, putatively damaging variants effectively identifies gene-metabolite relationships.

### Prioritization and characteristics of driver variants

We performed a forward selection procedure^15^ to assess the contribution of individual QVs to their gene-based association signals (Methods). Plots that visualize the association P-value based on the successive aggregation of the most influential QVs in plasma (**Supplementary Figure 3**) and urine (**Supplementary Figure 4**) revealed noteworthy differences: first, each of the two masks detected some genetic associations that were not significant with the respective other mask, highlighting differences in genetic architecture (e.g., *SLC10A2* and urine glycocholate with HI_mis vs. *ABCA7* and plasma lactosyl-N-nervonoyl-sphingosine with LoF_mis). Second, some genes showed different association patterns for the same metabolite in plasma and in urine (e.g., *TMLHE* and hydroxy-N6,N6,N6-trimethyllysine). Third, histidine exemplifies a metabolite with different associated genes in plasma (*HAL*) and urine (*SLC6A19*), implicating an enzyme involved in its hepatic and blood-based breakdown and a transporter responsible for its tubular reabsorption. Fourth, the same metabolite was sometimes associated with several genes in the same matrix, which differed in terms of genetic architecture (e.g., urine diacetylspermidine with *PAOX* and *HDAC10*, or plasma N,N,N-trimethyl-5-aminovalerate with *SLC22A5* and *TMLHE*).

The inclusion of effectively neutral variants among the QVs may dilute their joint signal. We thus prioritized the variants with the strongest individual contributions to the gene-based signal that resulted in the lowest possible association P-value when aggregated for burden testing^15^ (Methods) as “driver variants”. The proteins encoded by the vast majority of identified genes are directly involved in the generation, turnover, or transport of the associated metabolite(s). It is therefore a reasonable assumption that truly functional variants are those with the strongest individual contributions to the metabolite signal. Indeed, the minimum association P-value based on driver variants only was often many orders of magnitude lower than the one obtained from all QVs, as exemplified by *DPYD* and plasma uracil (**Supplementary Figure 3**). As expected, the proportion of splice, stop-gain and frameshift variants was higher among driver QVs, whereas non-driver QVs contained a greater proportion of missense variants (Fisher’s exact test: P-value=1.3e-6, **Supplementary Figure 5a**). The median effect of driver variants on metabolite levels increased from missense over start/stop-lost, frameshift, and stop-gain to variants predicted to affect splicing (**Supplementary Figure 5b**).

Lastly, we evaluated the convergence of rare and common variant association signals by assessing whether the regions around the identified genes contained common variants significantly associated with the respective metabolite in the same matrix (Methods). We detected significant associations for 157 of the 235 (192+43) unique gene-metabolite pairs (**Supplementary Table 6**). While the absolute effect size generally increased with lower minor allele frequency, there was no relation between the absolute aggregated effect size of rare variants with the presence of a GWAS signal in the region (**Supplementary Figure 6**).

In summary, genetic architecture differs across metabolite-associated genes, and further improvements in the selection of functional variants may increase the yield of future gene discovery efforts.

### Heterozygous variant carriers inform about dose-response effects

Our identification of known IEM-causing variants such as in *CTH*, *PAH*, *SLC16A9*, and *SLC7A9* supports the notion that heterozygous QVs are functional alleles. Moreover, we had previously confirmed experimentally heterozygous sulfate-associated QVs in *SLC26A1* as loss-of-function alleles and designated the encoded protein as an important player in human sulfate homeostasis.^25^ However, experimental studies of each of the detected 2,077 QVs and 73 genes are infeasible, and IEMs are so rare that no homozygous person for a given gene may have been observed yet. We therefore used three orthogonal approaches, examination of hemizygosity, *in silico* knockout modeling, and investigation of variants prioritized through allelic series, to evaluate whether the observed metabolite-associated heterozygous variants captured similar information about a gene’s function as might be derived from homozygous damaging variants in the respective gene.

### X-chromosomal associations as readouts of variant homozygosity

Genes in the non-pseudo-autosomal region of the X chromosome offer an opportunity to study differences between heterozygous women and effectively homozygous (i.e., hemizygous) men. We therefore investigated sex differences for the two X-chromosomal genes identified in our screen, *TMLHE* and *RGN* (**Supplementary Table 7**).

Indeed, male carriers of QVs in *TMLHE* showed clearly higher urine levels of N6,N6,N6-trimethyllysine, the substrate of the encoded enzyme trimethyllysine dioxygenase, than female carriers, as well as markedly lower levels of its product hydroxy-N6,N6,N6-trimethyllysine (**Figure 3**, **Supplementary Table 7**). In plasma, male QV carriers showed 1.15 standard deviations (SD) lower levels of plasma hydroxy-N6,N6,N6-trimethyllysine as compared to non-carriers (P-value=6e-44), whereas female QV carriers only showed 0.45 SD lower metabolite levels than non-carriers (P-value=3e-4). Similar differences, albeit less pronounced, were observed for *RGN* and urine levels of the unnamed metabolite X-23436. Levels were higher in women than men, suggesting that X-23436 is a metabolite downstream of the reaction catalyzed by the encoded regucalcin (**Supplementary Table 7**). Data from the GTEx Project^26^ shows no sex differences in gene expression across tissues. Hence, sex-differential effects of QVs on metabolite levels likely represent a dose-response effect resulting from QV hetero-vs. hemizygosity.

**Figure 3:**
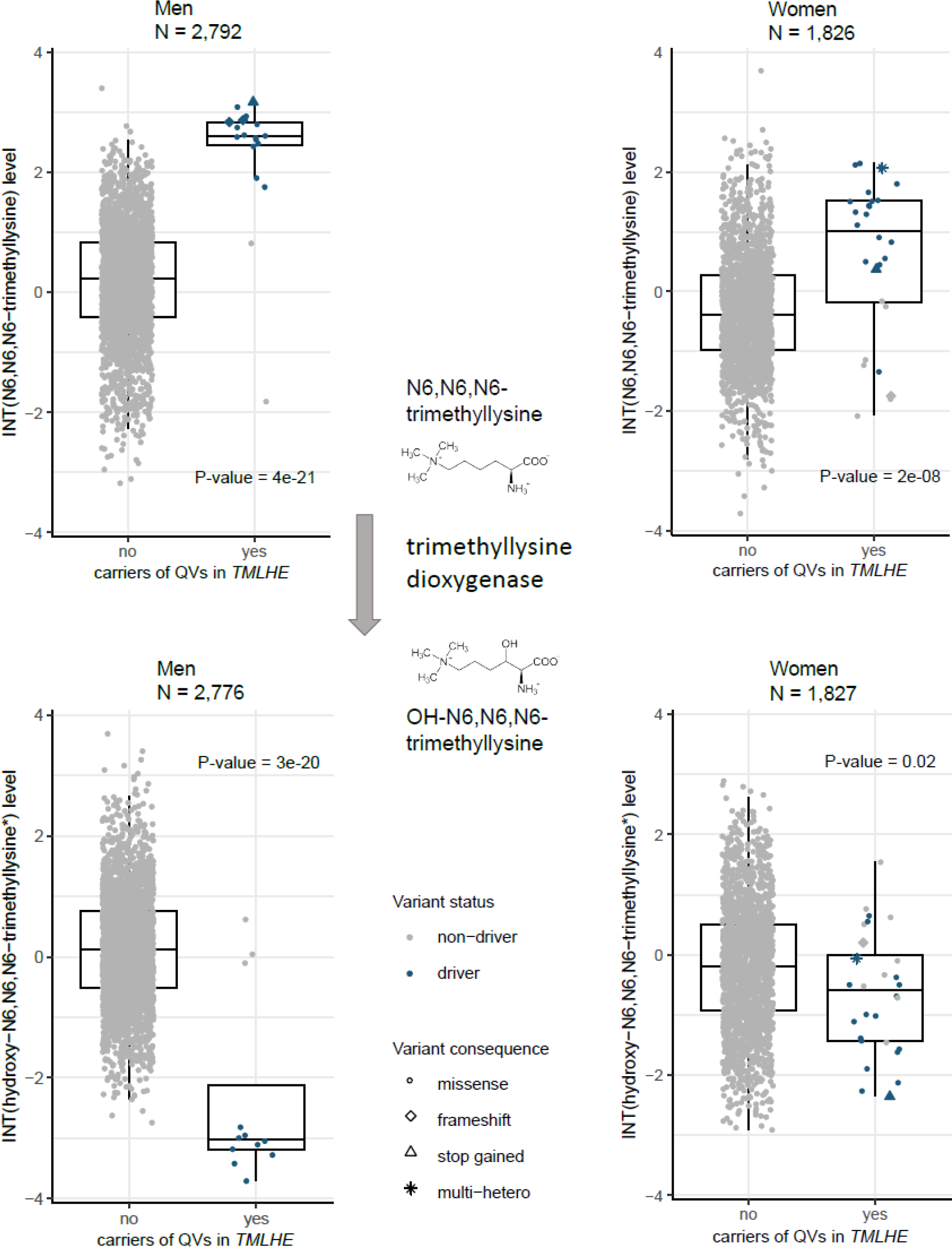
Differences in urine metabolite levels between male and female carriers of QVs in X-chromosomal *TMLHE* reflect a dose-response effect. The upper plots represent urine levels of N6,N6,N6-trimethyllysine after inverse normal transformation (y-axis) among male (left) and female (right) non-carriers and carriers of QVs in *TMLHE* based on the HI_mis mask (x-axis). Symbol color and shape indicates a variant’s driver status and consequence, respectively. The boxes range from the 25^th^ to the 75^th^ percentile of metabolite levels, the median is indicated by a line, and whiskers end at the last observed value within 1.5*(interquartile range) away from the box. Among men hemizygous for a QV in *TMLHE*, the levels of the substrate N6,N6,N6-trimethyllysine are markedly higher compared to heterozygous women, reflecting more severe impairment of encoded enzyme’s function in hemizygous men. The presented P-values correspond to the sex-specific burden tests. Metabolites’ formulas are taken from https://commons.wikimedia.org/. The lower plots represent urine levels of hydroxy-N6,N6,N6-trimethyllysine after inverse normal transformation (y-axis) across male (left) and female (right) non-carriers and carriers of QVs in *TMLHE* based on the HI_mis mask (x-axis). Because hydroxy-N6,N6,N6-trimethyllysine is the product of trimethyllysine dioxygenase, the enzyme encoded by *TMLHE*, loss-of-function QVs lead to decreased metabolite levels, more strongly among men than women.

### Virtual IEMs mirror the effects of heterozygous variants

We next investigated the implicated genes’ loss-of-function by generating virtual IEMs for 25 genes that covered 59 gene-metabolite pairs, via *in silico* knockout modeling (Methods). We compared the maximal secretion flux of the metabolite of interest into blood and/or urine between the wild-type WBM and the gene knockout WBM. Initially, the direction of the observed gene-metabolite associations was correctly predicted by virtual IEMs with an accuracy of 74.58% in the male and 77.97% in the female WBM, which is significantly better than chance (Fisher’s exact test: P-value=4.4e-03 (male), P-value=1.2e-04 (female); **Supplementary Table 8**). After model curation informed by the genome and metabolome data from the GCKD study, which included the addition of metabolites (e.g., 8-methoxykynurenate) and pathways as well as alteration of constraints (e.g., diet; details in **Supplementary Material, Supplementary Table 9**), the number of modeled gene-metabolite associations increased to 67, and accuracy to 79.1% (male; P-value=2.1e-05) and 83.58% (female; P-value=2.9e-07). These findings underline the predictive nature of the virtual IEMs for the aggregated effects of heterozygous damaging variants, and highlight opportunities to further improve WBMs by curation of the underlying knowledge base.

### Microbiome-personalized WBMs capture quantitative changes in metabolites observed for heterozygous and homozygous loss of KYNU function

Virtual IEMs only allow for qualitative prediction. To additionally study an equivalent to observed effect sizes, we introduced a second *in silico* modeling strategy as proof of principle. We successfully generated 582 microbiome-personalized^27^ WBMs (Methods), and calculated the effect size of an *in silico KYNU* knockout against the natural variation induced by the personalized microbiomes on metabolite excretion into urine (**Supplementary Table 10**). There were 16 of 242 metabolites available in both GCKD and the WBMs with modeling P-value <0.05/242, implicating them as potential biomarkers of kynureninase deficiency (**Supplementary Table 11**), mostly belonging to tryptophan metabolism and the NAD+ *de novo* synthase pathway. The *in silico* effects of these 16 biomarkers predicted their observed counterparts (Pearson correlation r=0.61 (P-value=0.013); **Figure 4a**), and highlighted large effects for 3-hydroxykynurenine, 8-methoxykynurenate, and xanthurenate. While both xanthurenate and 3-hydroxykynurenine are known biomarkers of kynureninase deficiency^28^, 8-methoxykynurenate was novel. We next measured absolute levels of these metabolites in urine samples from a patient with a homozygous loss-of-function variant causing kynureninase deficiency and her parents^29^ (Methods), and confirmed that not only xanthurenate and 3-hydroxykynurenine but also 8-methoxykynurenate constituted a biomarker of this IEM (**Figure 4b, Supplementary Figure 7**). Microbiome-personalized WBMs correctly predicted smaller changes in 8-methoxykynurenate than in its precursor xanthurenate, consistent with the absolute levels measured in the IEM patient as well as with the association statistics from aggregate variants tests in the GCKD study (**Figure 4b, Supplementary Figure 7b**). Thus, *in silico* WBM modeling faithfully captured metabolic changes observed for both, population-based heterozygous variants and an IEM caused by a homozygous *KYNU* mutation.

**Figure 4:**
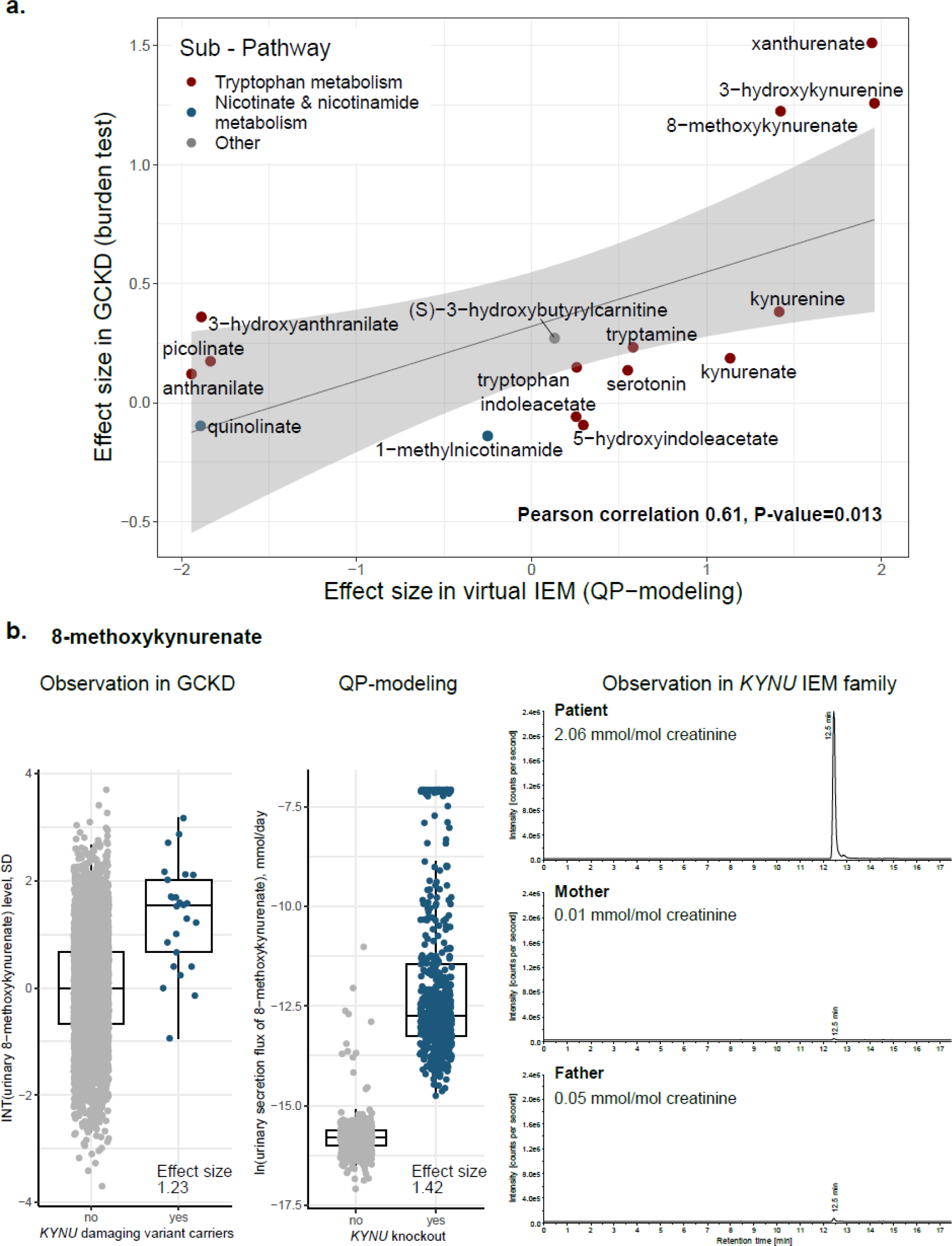
Altered metabolite levels in urine are a readout of impaired *KYNU* function: converging evidence from three approaches. **(a)** Relation between effect sizes (regression coefficients) upon *in silico* knockout of *KYNU* based on 582 microbiome-personalized WBMs (x-axis) and observed effect sizes in the GCKD study (y-axis) for 16 urine metabolites that showed significant changes upon *in silico* knockout of *KYNU*. WBM estimates are based on QP-modeling, and GCKD estimates on aggregating rare, damaging variants in *KYNU*. Symbol color represents the sub-pathway of the corresponding metabolite. The gray line is the linear regression line through the data points, the shaded gray area represents its 95% confidence interval. Simulated *in silico* effects of *KYNU* knockout are clearly correlated with the observed effects in humans (Pearson correlation r=0.61, P-value=0.013). **(b)** Three panels are shown for 8-methoxykynurenate: the left panel represents inverse-normal transformed urine levels of the metabolite (y-axis) among non-carriers and carriers of QVs in *KYNU* (x-axis). Units correspond to standard deviations. The boxes range from the 25^th^ to the 75^th^ percentile of metabolite levels, the median is indicated by a line, and whiskers end at the last observed value within 1.5*(interquartile range) away from the box. The middle panel represents the distribution of the ln-transformed secretion flux of the metabolite in mmol/day into urine (y-axis) from min-norm simulations based on 582 microbiome-personalized WBMs without and with simulated knockout of *KYNU* (x-axis). The right panel shows multiple reaction monitoring (MRM, *m/z* 220.0 → 174.1) chromatograms of the diluted urines of a child with a homozygous loss of *KYNU* function (patient), the heterozygous mother and the healthy father (maternal uniparenteral isodisomy). The signal at 12.5 min representing 8-methoxy-kynurenate is strongly enhanced in the patient sample. Chromatograms are normalized to urine creatinine concentrations; y-axes are normalized to the intensity of the signal in the patient’s chromatograms. All three independent approaches arrive at the conclusion that elevated levels of 8-methoxykynurenate in urine are a readout of impaired *KYNU* function.

### Association of metabolite-associated alleles and genes with human traits and diseases

We queried data from ∼450,000 UKB participants with WES for associations of the identified 2,077 QVs and 73 genes with thousands of quantitative and binary health outcomes that may result from disturbances of the implicated metabolites. The prefiltered UKB dataset (Methods) contained 696 QVs and 72 genes. At the gene-level, significant associations (P-value<2e-09; Methods) were identified between *APOC3* and the binary health outcome “disorders of lipoprotein metabolism and other lipidaemias” (**Supplementary Table 12**), consistent with its association with 19 plasma phosphatidylethanolamine and diacylglycerol metabolites in our study. Moreover, 13 genes showed 282 significant associations with quantitative health outcomes. These mostly arose from clinical chemistry parameters and contained many plausible and well supported examples (**Supplementary Table 12**). At the variant-level, there were 555 significant associations between a QV and a quantitative as well as two additional associations with a binary health outcome (**Supplementary Table 13**). These included well-established examples, but also less studied candidates such as an *SLC6A19* variant encoding the p.Asp173Asn substitution in the sodium-dependent neutral amino acid transporter, which was associated with lower serum creatinine and cystatin C levels and erythrocyte distribution width.

We have previously shown that the comparison of the effect of common genetic variants on plasma and urine metabolite levels can deliver specific insights into functions of the kidney^2^. In this study of rare variants, all identified genes that were associated with one or more measures of kidney function (i.e., serum creatinine or cystatin C) in the UKB encode for transport proteins that are highly expressed in the kidney^30–32:^ *SLC47A1*, *SLC6A19*, *SLC7A9*, and *SLC22A7* (**Supplementary Table 12**). The gene products of *SLC47A1*, *SLC6A19*, and *SLC7A9* are localized in the apical membrane of tubular cells^30–32^, where they are involved in the secretion of organic cations (*SLC47A1*) or tubular reabsorption of amino acids (*SLC6A19*, *SLC7A9*). Their metabolic fingerprints were almost exclusively detected in urine (**Supplementary Table 3**) and reflected the encoded proteins’ functions. For example, carriers of QVs in *SLC7A9* showed significantly higher levels of urine cystine and lysine, consistent with its function in the reabsorption of dibasic amino acids from urine. Conversely, *SLC22A7* encodes for an organic anion transporter in the basolateral membrane of tubular cells^33^. An exchange against intracellular glutamate has been reported^34^, which may contribute to the observed association with lower plasma gamma-glutamylglutamate levels among carriers of *SLC22A7* QVs compared to non-carriers (**Supplementary Table 3**). QVs in *SLC47A1* and *SLC22A7* were only associated with creatinine levels but not with cystatin C, in agreement with their known role as creatinine transporters^35^. In contrast, QVs in *SLC7A9* and *SLC6A19* showed association with lower levels of both creatinine and cystatin C^36^, suggesting that their loss-of-function is associated with better kidney function through yet unidentified mechanisms. These observations illustrate how rare damaging variants leave a specific signature in plasma and urine metabolomes that mirror exchange processes at the membranes of kidney epithelial cells and are related to kidney function.

### Allelic series: metabolites represent intermediate readouts of pathophysiological processes

Allelic series describe a dose-response relationship, in which increasingly deleterious mutations in a gene result in increasingly larger effects on a trait or a disease. We hypothesized that genetic effects on metabolite levels should manifest as allelic series if the metabolite represents a molecular readout of an underlying (patho-)physiological process. As proof of principle, we investigated plasma sulfate, because of solid evidence for causal gene-metabolite relationships: first, QVs in *SLC13A1* showed a significant aggregate effect on lower plasma sulfate levels (P-value=3E-18, lowest possible P-value=2e-25). The observed association is well supported by experimental studies establishing that the encoded Na^+^-sulfate cotransporter NaS1 (SLC13A1) reabsorbs filtered sulfate at the apical membrane of kidney tubular epithelial cells^37^. Second, we had previously confirmed experimentally that plasma sulfate-associated QVs in *SLC26A1* reduced sulfate transport capacity^25^ and confirmed a lowest possible P-value of 2e-11 for the aggregate effect of driver variants in *SLC26A1* (**Supplementary Figure 8**). The encoded sulfate transporter SAT1 localizes to basolateral membranes of tubular epithelial cells and works in series with NaS1 to mediate transcellular sulfate reabsorption (**Figure 5a**)^38,39^.

**Figure 5:**
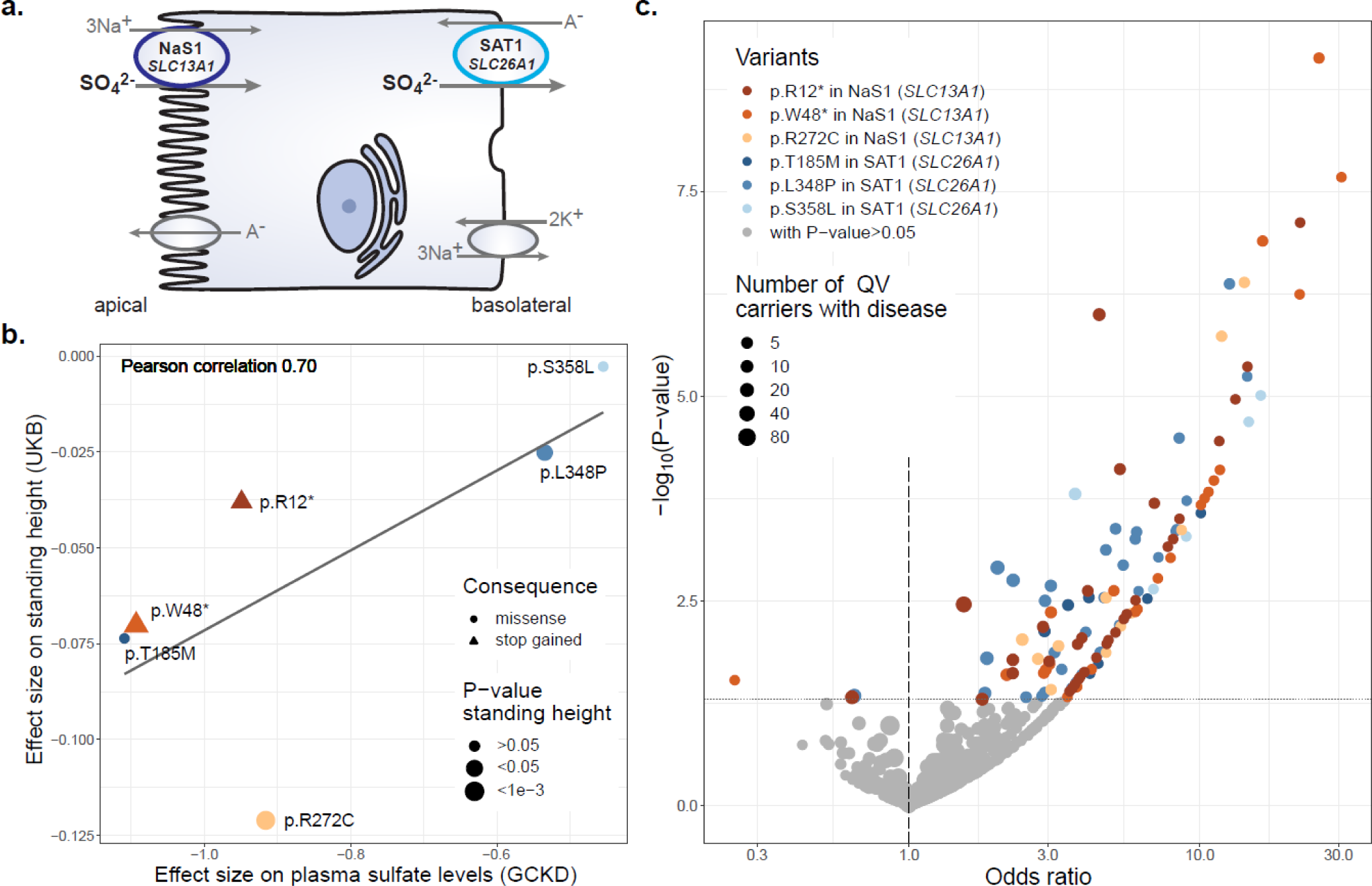
Impact of functional QVs in *SLC13A1* and *SLC26A1* on height, musculoskeletal traits and fractures support the role of plasma sulfate as intermediate readout. (a) Schematic representation of the sulfate reabsorption mechanism involving NaS1 encoded by *SLC13A1* at the apical membrane and SAT1 encoded by *SLC26A1* at the basolateral membrane of epithelial cells. (b) The scatter plot shows the relation between the effect sizes of 6 QVs on plasma sulfate levels in the GCKD study (x-axis) and on standing height in the UKB (y-axis). Effect sizes correspond to single variant association tests under additive modeling with inverse normal transformed traits. Symbol color and shape indicate the gene (shades of red: *SLC13A1*, shades of blue: *SLC26A1*) and consequence of the QV. Symbol size represents the P-value with respect to height. The gray line is the linear regression line through the data points. Variant effect sizes on plasma sulfate levels are clearly correlated with the ones on standing height (Pearson correlation r=0.70, allelic series). (c) The volcano plot shows odds ratios (x-axis) and -log_10_(P-values) (y-axis) for association of the 6 QVs with musculoskeletal diseases and fractures in the UKB, based on a Firth regression. Only clinical traits for which at least two carriers were identified are included in the plot. Symbol color indicates the QV and whether the corresponding P-value was nominally significant (P-value<0.05). Symbol size corresponds to the number of QV carriers with disease. While both increased and decreased odds of disease were observed when associations were not significant, increased odds for musculoskeletal diseases and fractures clearly dominated for significant associations.

Based on a growth retardation phenotype in slc13a1 knockout mice^40^ and the observed association between *SLC13A1* and lower sitting height in the UKB (P-value=3E-08, **Supplementary Table 12** and ^41^), we investigated relations of three functional QVs in *SLC13A1* and *SLC26A1* each with anthropometric measurements in the UKB (Methods). **Supplementary Table 14** contains traits for which at least two QVs were nominally associated (P-value<0.05). There was a clear correlation between genetic effect sizes on plasma sulfate levels in the GCKD study and both sitting and standing height in the UKB (Pearson correlation coefficients of 0.57 and 0.70, respectively; **Figure 5b**). These observations support a causal relationship between transcellular sulfate reabsorption and human height, and designate plasma sulfate as an intermediate readout. Additionally, we could observe a significant lower standing height among carriers of driver variants in one of the two genes (*SLC13A1* and *SLC26A1*) compared to non-carriers in a subsample of the GCKD study (N=3,239), where height was measured at baseline. Aggregating the effect of driver variants in *SLC13A1* provided an effect size of −0.54 (corresponding to −5.17 cm when the outcome height was not inverse normal transformed, P-value=1.6e-3, **Supplementary Figure 9a**). For *SLC26A1* we obtained even a stronger effect size of −0.73 (corresponding to −6.68 cm, P-value=1.7e-6, **Supplementary Figure 9b**).

The first patient homozygous for a loss-of-function stop gained mutation in *SLC13A1*, p.Arg12*, has just been described^42^. Besides sitting height >2 SD below the normal range, the patient featured multiple skeletal abnormalities. His fractional sulfate excretion of almost 100%, as well as earlier model-based transport studies^43^, establish this variant as a complete loss-of-function resulting in renal sulfate wasting. We found that compared to non-carriers of p.Arg12*, heterozygous carriers showed 0.95 SD lower plasma sulfate levels (GCKD, 22 carriers, P-value=9.9E-10) and 0.08 SD lower sitting height (UKB, 2,480 carriers, P-value=2.2E-07). Plasma sulfate measurements from heterozygous carriers therefore inform about phenotypes that will exhibit more extreme changes in the homozygous state.

### Functional variants of altered sulfate reabsorption increase odds of musculoskeletal diseases

Rare loss-of-function variants in *SLC13A1* and *SLC26A1* have been linked to individual musculoskeletal phenotypes through IEMs and GWAS^25,41,44,45^. We further investigated the association between the same six functional, sulfate-associated QVs in *SLC13A1* and *SLC26A1* and musculoskeletal disorders, fractures, and injuries in the UKB (Methods). There were 116 nominally significant (P-value<0.05) associations with clinical traits and diseases, 113 of which were associated with increased odds of disease (**Figure 5c**). For instance, increased odds of various fractures ranged from 1.9 for closed pertrochanteric fracture (P-value=0.016, SAT1 p.Leu348Pro) to 30.7 for closed fracture of the neck (P-value=2.1e-08, NaS1 p.Trp48*; **Supplementary Table 15**).

Lastly, we investigated UKB participants who carried more than one copy of any of the six QVs more closely. The rare allele of the missense variant p.Arg272Cys in NaS1, observed in nine heterozygous carriers in GCKD, had been prioritized because of its location in a splice region, its high impact on plasma sulfate levels, and its particularly large effect on human height (**Figure 5b**). In the UKB, we found 294 heterozygous carriers of p.Arg272Cys, four persons who carried p.Arg272Cys in NaS1 as well as SAT1 p.Leu348Pro, and a single person homozygous for p.Arg272Cys. Age- and sex-specific z-scores for human height (Methods) showed a clear dose response effect (**Figure 6a**). Interestingly, the second group of four individuals were heterozygous for loss-of-function variants in each of the two transcellular sulfate reabsorption proteins, supporting that the pathway is important for human growth. Carrier status for NaS1 p.Arg272Cys was associated with increased odds of several musculoskeletal diseases such as back pain and intervertebral disc disorders as well as fractures (**Figure 6b**). Homozygous persons were also identified for NaS1 p.Arg12* and SAT1 p.Leu348Pro, with similar findings (**Supplementary Figure 10**). Together, these findings provide convincing evidence that lower transcellular sulfate reabsorption is associated with numerous adverse musculoskeletal traits and diseases. Prioritizing variants with strong effects in allelic series for subsequent investigation in larger studies, even if the biomarker association rests on only a few heterozygous alleles, is an effective strategy to gain insight into the impact of rare damaging variants on human health.

**Figure 6:**
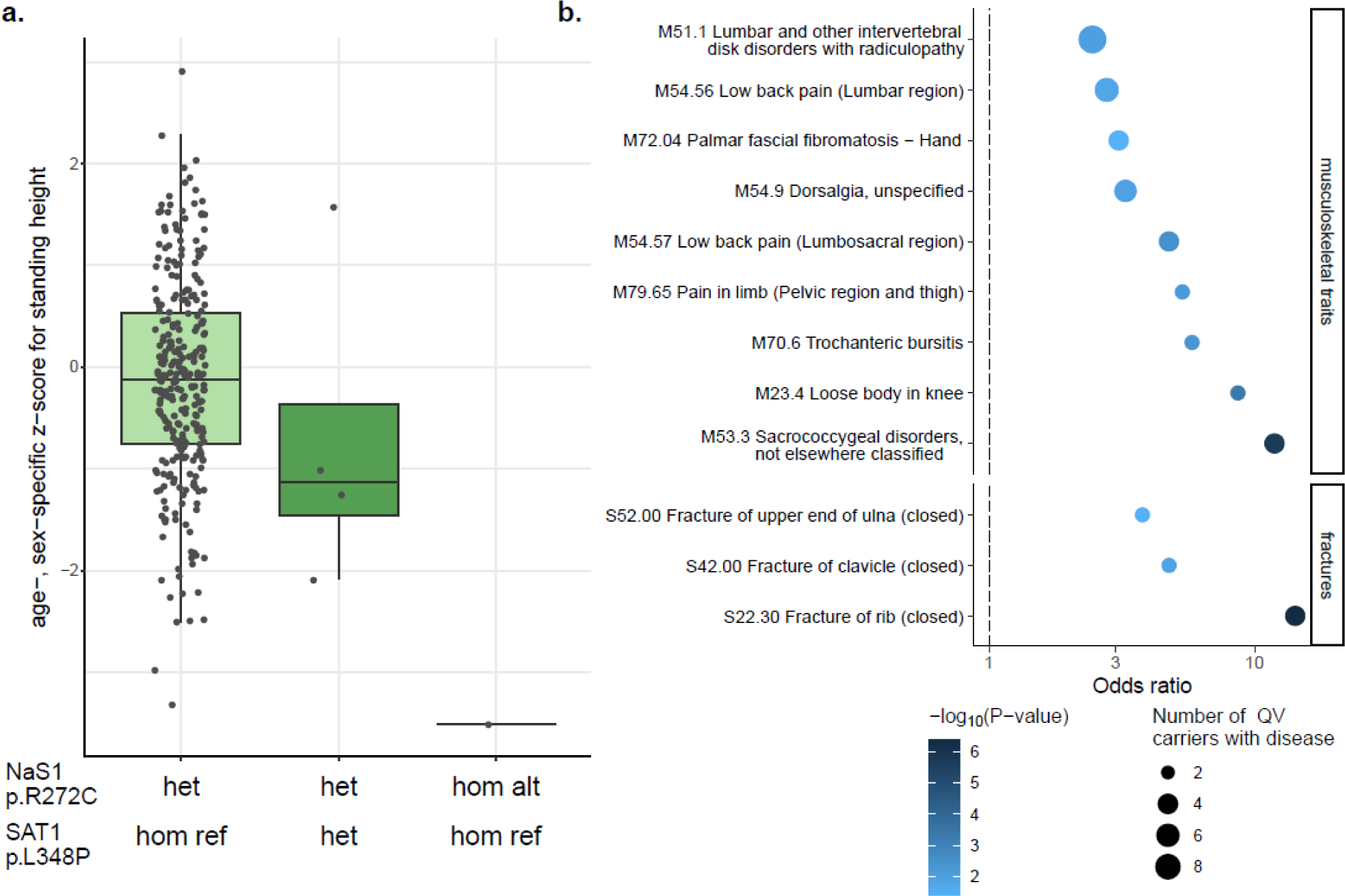
Impact of different genotypes encoding the NaS1 p.Arg272Cys substitution on height and musculoskeletal traits and fractures. **(a)** The boxplot shows differences in age- and sex-specific z-scores for standing height (y-axis) across persons heterozygous and homozygous for the p.Arg272Cys-encoding allele (x-axis). A dose-response effect is observable between heterozygous individuals (N=289, median=-0.13), individuals carrying NaS1 p.Arg272Cys as well as SAT1 p.Leu348Pro (N=4, median=-1.13), and one person homozygous for p.Arg272Cys (z-score=-3.51). **(b)** Association between the NaS1 p.Arg272Cys substitution with musculoskeletal diseases and fractures from the UKB, for which at least 2 carriers were identified (y-axis). Odds ratios (x-axis) are based on a Firth regression. The symbol color reflects the -log_10_(P-value) and the size the number of p.Arg272Cys carriers with disease. Only associations with P-value<0.05 are shown.

## Discussion

We performed a comprehensive screen of the aggregate effect of rare, putatively damaging variants on the levels of 1,294 plasma and 1,396 urine metabolites from paired specimens of 4,737 persons. Of the 192 identified gene-metabolite relationships, 115 have not yet been reported yet, and include plasma- and urine-exclusive associations that reflect organ function. We show via three computational and genetic approaches that the rare, almost exclusively heterozygous metabolite-associated variants in our study capture similar information about a gene’s function than obtained from the study of rare IEMs.

We present several lines of evidence that heterozygous variants identified in a population sample permit insights into graded effects of impaired gene function without the need to identify patients with a corresponding biallelic IEM. First, 38% of identified genes in our study are known to harbor causative mutations for autosomal recessive IEMs that often exhibit concordant changes in the implicated metabolite. This is exemplified by elevated urine cystine in cystinuria patients (MIM #220100, *SLC7A9*), elevated urine tryptophan in patients with Hartnup disease (MIM #234500, *SLC6A19*), lower plasma carnitine in patients with systemic primary carnitine deficiency (MIM #212140, *SLC22A5*), and elevated plasma histidine in patients with histidinaemia (MIM #235800, *HAL*).

Second, men exhibited significantly larger effects of rare QVs in non-pseudo-autosomal X-chromosomal genes on metabolite levels than women. This observation is consistent with male hemizygosity as an approximation of female homozygosity for a given variant, and with the known greater penetrance and severity of X-linked disorders in men as compared to women^46^.

Third, *in silico* knockout in a virtual metabolic human, i.e. the full loss-of-gene function, was predictive for both direction and magnitude of observed metabolic changes associated with variant heterozygosity. Predicted different effect sizes on metabolite levels upon *in silico* loss of *KYNU* function were also reflected in absolute urine metabolite quantification of a patient with homozygosity for a full loss-of-function *KYNU* mutation^29^. Blockage of the NAD+ *de novo* synthase pathway, as reflected in the predicted reduction of the respective metabolites’ flux upon *in silico* knockout of *KYNU*, is considered causal for the severe symptoms associated with kynureninase deficiency, or Vertebral, Cardiac, Renal and Limb Defect Syndrome^29^. Therefore, the virtual IEM is in line with the current hypothesis regarding disease etiology. Thus, deterministic, knowledge-based *in silico* modeling generates context for better biological interpretation also of heterozygous variants, while the population-based genetic screens of metabolite levels permit the identification of knowledge gaps and errors in WBMs. Our modeling pipeline for generating virtual IEMs, which we make publicly available, will constitute a valuable resource for the scientific community in particular to scrutinize genes for which no IEM has been observed.

Fourth, the presence of different causal QVs affecting a given metabolic reaction or pathway enabled the investigation of allelic series. The resulting dose-response relationships proxy a range of target inhibition, which represents highly desirable information for drug development and is relevant because enzymes and transporters are attractive drug targets. Plasma sulfate-associated functional QVs in *SLC13A1* and *SLC26A1* showed a clear dose-response effect between the degree of impaired epithelial transcellular sulfate reabsorption and lower human height. This observation is biologically plausible, because defects in genes linked to sulfate biology often result in perturbed skeletal growth and development^47^. In particular, constitutive knockouts of *slc13a1* and *slc26a1* in mice do not only cause hyposulfatemia and renal sulfate wasting^40,48^, but also general growth retardation in *slc13a1* knockout mice^40^. Interestingly, the missense variant p.Thr185Met in SAT1 exhibited the largest effect on sulfate. We have previously shown experimentally a dominant negative mechanism of this variant^25^, providing another mechanism how heterozygous variants may promote insights into an effectively full loss-of-gene-function. Moreover, our findings for the p.Arg272Cys variant in NaS1 show that even very few, heterozygous copies of a metabolite-prioritized QV can give rise to the detection of homozygous individuals and hitherto unreported disease associations in subsequent larger studies. These observations suggest that the importance of impaired transcellular epithelial sulfate transport for musculoskeletal diseases, fractures, and injuries has been underestimated previously.

Potential limitations of our study deserve discussion. First, due to the GCKD study design, it is unclear if our findings apply to persons of non-European ancestry. However, rare genetic variants that are predicted or experimentally shown to result in loss-of-function should show effects on associated metabolites and diseases regardless of genetic background. Second, burden tests assume that all aggregated QVs result in direction-consistent effects of similar size, which, if violated, results in a loss of power^49^. Because our study assumed loss-of-function as the mechanism underlying metabolic changes, we did not evaluate alternative aggregate variant tests such as SKAT^50^. SKAT is less powerful in a setting with direction-consistent effects^51^, does not provide effect sizes, and is difficult to interpret and replicate^7,52^. Third, inclusion of effectively neutral variants as QVs in a burden test can lead to an underestimation of a gene’s effect. Further methodological improvements are required in order to better predict a variant’s functional consequence, as well as for optimizing the selection and weighting of QVs to better reflect specific genetic architectures. Fourth, we analyzed non-targeted population metabolomics data. However, non-targeted metabolomics provides much broader coverage than conventional targeted screening within and across biochemical pathways^53^, thus enabling the discovery of genetic associations with previously unreported metabolites, as well as the detection of entirely new gene-metabolite relationships as observed here. Lastly, we utilized WBMs for *in silico* validation based on the steady state assumption, whereas it is conceivable that dynamic modeling may improve the predictive power of virtual IEMs. However, such modeling is computationally expensive, and adequate data for fitting dynamic models are often missing. A great advantage of the utilized constraint-based modeling is its scalability, permitting easy integration with genome-wide genetic screens.

In conclusion, the exome-wide study of rare, putative loss-of-function variants can establish causal relationships with metabolites, and highlight metabolic biomarkers that reflect the degree of impaired gene function and result in graded, adverse effects on human health.

## Online Methods

### Study design and participants

The German Chronic Kidney Disease (GCKD) study is an ongoing prospective cohort study of 5,217 participants with CKD that were enrolled from 2010 to 2012 and are under regular nephrologist care. Inclusion criteria were an age between 18 to 74 years and an eGFR between 30–60 mL/min/1.73 m^2^ or an eGFR >60 mL/min/1.73 m^2^ with an UACR >300 mg/g or with a urinary protein to creatinine ratio >500 mg/g.^54^ Biomaterial, including blood and urine, were collected at the baseline visit, processed and shipped frozen to a central biobank for storage at −80 degrees Celsius.^55^ More details on the description of the study design and participants characteristics have been published.^54,56^ The GCKD study was registered in the national registry for clinical studies (DRKS 00003971) and approved by local ethics committees of the participating institutions.^54^ All participants provided written informed consent.

### Whole-exome-sequencing and quality control

Genomic DNA was extracted from whole blood and underwent paired-end 100-bp whole-exome sequencing at Human Longevity Inc, using the IDT xGen v1 capture kit on the Illumina NovaSeq 6000 platform. More than 97% of consensus coding sequence (CCDS) release 22^57^ had at least 10x coverage. The average coverage of the CCDS was 141-fold read depth. Exomes were processed from their unaligned FASTQ state in a custom-built cloud compute platform using the Illumina DRAGEN Bio-IT Platform Germline Pipeline v3.0.7 at Astra Zeneca’s Centre for Genomics Research, including alignment of reads to the GRCh38 reference genome and variant calling.^58^

Sample level quality control included removal of samples from participants who withdrew consent, duplicated samples, those with an estimated VerifyBamID contamination level >4%^59^, samples with inconsistency between reported and genetically predicted sex, samples not having chromosomes XX or XY, samples having <94.5% of CCDS release 22 bases covered with ≥10-fold coverage^57^, related samples with kinship >0.884 (KING--kinship v2.2.3)^60^ and samples with a missing call rate >0.03. Furthermore, only samples with available high-quality DNA microarray genotype data and without outlying values (>8 SD) along any of the first 10 genetic principle components from a principal component analysis (for more details see^61^) were kept, for a final sample size of 4,779 samples.

Variant level quality control as described previously^58^ included exclusion of variants with coverage <10, heterozygous variants with a one-sided binomial exact test P-value <1e-6 for Hardy-Weinberg equilibrium, variants with a genotype quality score (GQ) <30, single nucleotide variants (SNV) with a Fisher’s strand bias score (FS) >60 and insertions and deletions (indel) with a FS >200, variants with a mapping quality score (MQ) <40, those with a quality score (QUAL) <30, variants with a read position rank sum score (RPRS) <-2, those with a mapping quality rank sum score (MQRS) <-8, variants that did not pass the DRAGEN calling algorithm filters, heterozygous genotype called variants based on an alternative allele read ratio <0.2 or >0.8, and variants with a missing call rate >10% among all remaining samples. That resulted in 1,038,062 variants across the autosomes and the X chromosome.

### Variant and gene annotation

Variants from WES were annotated using the Variant Effect Predictor (VEP) version 101^62^ with standard settings, including the canonical transcript, gene symbol and variant frequencies from the Genome Aggregation Database (gnomAD version 2.1 https://gnomad.broadinstitute.org/). VEP plugins were used to add the REVEL (version 2020-5)^63^ and CADD (version 3.0)^64^ scores. The LoFtee VEP plugin (version 2020-8)^65^ was used to downgrade loss-of-function variants. Furthermore, we added multiple *in silico* prediction scores using dbNSFP version 4.1a.^66^

For interpretation, genes were annotated for their potential function as enzymes using Uniprot (https://www.uniprot.org/)^67^ and as transporters using Gyimesi and Hediger 2022^68^.

### Metabolite identification and quantification

Metabolite levels were quantified from stored plasma and spot urine as described previously^2^. In brief, non-targeted mass spectrometry analysis was conducted at Metabolon, Inc. Metabolites were identified by automated comparison of the ion features in the experimental sample to a reference library of chemical standard. Known metabolites reported in this study were identified with the highest confidence level of identification of the Metabolomics Standards Initiative^69,70^, unless marked by an asterisk. Unnamed biochemicals of unknown structural identity were identified by virtue of their recurrent nature. For peak quantification, the area under the curve was used, followed by normalization to account for inter-day instrument variation.

### Data cleaning of quantified metabolites

Data cleaning, quality control, filtering and normalization of quantified metabolites in plasma und urine in the GCKD study has been described previously^2^. Samples and metabolites were evaluated for duplicates, missing and outlying values and metabolites with low variance were excluded. Levels of urine metabolites were normalized using the probabilistic quotient^71^ derived from 309 endogenous metabolites with <1% missing values in order to account for differences in urine dilution. After removing metabolites for which less than 300 individuals with WES data were available, the remaining 1,294 plasma and 1,396 urine metabolites (**Supplementary Table 2**) were subjected to inverse normal transformation prior to gene-based aggregation testing.

### Additional variables

Serum and urine creatinine were measured as part of standard biochemistry using an IDMS traceable enzymatic assay (Creatinine plus, Roche). Serum and urine albumin were measured using the Tina-Quant assay (Roche/Hitachi Diagnostics GmbH, Mannheim, Germany). GFR was estimated with the CKD-EPI formula^72^ from serum creatinine. UACR was calculated using the urinary albumin and creatinine measurements. Full information on WES data, covariates, and metabolites was available for 4,713 persons regarding plasma metabolites, and for 4,619 persons regarding urine metabolites. Genetic principal components were derived based on a principal component analysis as described previously.^61^

### Rare variant aggregation testing on metabolite levels

We performed burden tests to combine the effects of rare, putatively damaging variants within a gene on metabolite levels assuming a loss-of-function mechanism that results in concordant effect directions on metabolite levels^49^. The selection of high-quality QVs into masks based on their frequency and annotated properties is a state-of-the-art approach in gene-based variant aggregation studies.^73^ Annotations from the Variant Effect Predictor (VEP) version 101^62^ were used to select qualifying variants within each gene for aggregation in burden tests. Because the genetic architecture of damaging variants can vary across genes, two complementary masks for the selection of qualifying variants were defined. Both masks were restricted to contain only rare variants in canonical transcripts with a MAF of <1%. All variants that were predicted to be either high-confidence loss-of-function variants or missense variants with a MetaSVM score >0 or in-frame non-synonymous variants with a fathmm-XF-coding score >0.5 were aggregated into the first mask, termed LoF_mis. The second mask, termed HI_mis, contained all variants that were predicted either to have a high-impact consequence defined by VEP (transcript ablation, splice acceptor variant, splice donor variant, stop gained, frameshift variant, stop lost, start lost, and transcript amplification) or to be missense variants with either a REVEL score >0.5, a CADD PHRED score >20, or a M-CAP score >0.025. Only genes with a HGNC symbol, that were no read-throughs and that contained >3 qualifying variants in at least one of the masks were kept for aggregate variant testing, resulting in 16,525 analyzed genes. Burden tests were carried out as implemented in the seqMeta R-package version 1.6.7^74^, adjusting for age, sex, ln(eGFR), the first three genetic principal components as wells as serum albumin for plasma metabolites and ln(UACR) for urinary metabolites, respectively. Genotypes were coded as number of copies of the rare allele (0, 1, 2) on the autosomes and also on the X chromosome for women. For men, genotypes in the non-pseudo-autosomal region of the X chromosome were coded as (0, 2). Statistical significance was defined as nominal significance corrected for the number of tested genes and principal components that explained more than 95% of the metabolites’ variance, leading to thresholds of 0.05/16525/600=5.04e-9 in plasma and 0.05/16525/679= 4.46e-9 in urine. For significant gene-metabolite associations, single-variant association tests between each qualifying variant in the respective mask and the corresponding metabolite levels were performed under additive modeling, adjusting for the same covariates mentioned above using the seqMeta R-package version 1.6.7^74^.

### Comparison to previous rare variant association studies and to GWAS of metabolite levels

We compared our significant findings to the significant findings from eight published genetic studies of the plasma/serum or urine metabolome that focused on rare exonic variant aggregation testing and used sequencing and high-throughput metabolomics data^14–20,24^. We first assessed whether the genes identified in our study were reported as associated with any metabolite in any of the seven studies at their respective multiple-testing corrected significance threshold, after having mapped all gene names to their current version in Ensembl version 109 using https://www.ensembl.org/biomart/martview. We then ascertained for all matching i.e. previously reported genes whether they were associated with the same metabolite(s) as in our study. Metabolites were matched by biochemical name, with manual curation in case of similar names, and by HMDB ID and Compound ID for metabolites quantified at Metabolon, if available.

The presence of common variants associated with the corresponding metabolite(s) in or near the identified genes was assessed by searching for common variants (MAF >1%) within a window of ±500 kb around the gene that were significantly (P-value <5e-8) associated with the implicated metabolite. Common variant associations were based on GWAS of inverse normal transformed metabolite levels in the GCKD study (N = 4,991 for plasma, N = 4,911 for urine) using REGENIE v2.2.4^75^, based on TOPmed imputed genotypes and adjusting for age, sex and the first three genetic principal components^2^. Gene positions were based on Ensembl version 101. Conditional association analyses were not performed, because previous studies by ourselves and others have shown that the vast majority of gene-based rare variant association signals with metabolites is unaltered by conditioning on common variant genotypes.^15,24,76^

### Assessment of qualifying variant contributions and selection of driver variants

The investigation of the genetic architecture underlying gene-metabolite associations and the prioritization of QVs according to their contribution to the gene-based association signal was performed using the forward selection procedure described in Bomba *et al* 2022^15^. First, for each QV *v* the P-value *P_v_* is calculated by performing the burden test aggregating all QVs except for the variant *v*. Second, for each QV *v* the difference *Δ_v_* between *P_v_* and the total P-value of the burden test including all QVs is calculated. The more a QV *v* contributes to the gene signal, the greater the resulting *Δ_v_*. Therefore, the QVs are ranked by the magnitude of *Δ_v_*. QVs not contributing to the gene signal or even having an opposite effect can provide a negative *Δ_v_*. Finally, burden tests are performed by adding the ranked QVs one after the other until the lowest P-value is reached starting with the greatest *Δ_v_*. We thereby identified a set of QVs for each gene-metabolite association that contained only those variants that contributed most to the gene-based association signal (i.e., led to a stronger association signal) and did not contain variants that introduced noise (i.e., neutral variants or those with a small or even opposite effect on metabolite levels). The resulting set of selected variants that drove the association signal and led to the lowest possible association P-value was designated “driver variants” for the respective gene-metabolite association. Driver variants within a gene might differ for different associated metabolites, and not all driver variants necessarily represent true causative variants.

### Relation of genes and variants to clinical traits and diseases

We used different data sources to link the associated genes and qualifying variants identified in our study to clinical outcomes and diseases. Implicated genes were queried for related monogenic disorders and traits using the OMIM catalog (https://www.omim.org/; accessed on 1/6/2022) and for the presence of known IEMs using https://panelapp.genomicsengland.co.uk/panels/467/ version v3.0. Drug target status and the corresponding indication were annotated for all identified genes by querying https://platform.opentargets.org/ on 7/12/2022. Clinical significance and the corresponding trait or disease were for all qualifying variants based on ClinVar https://www.ncbi.nlm.nih.gov/clinvar/ accessed on 3/30/2022.

Furthermore, we searched for gene-level and variant-level associations of the genes and qualifying variants identified in our study with about 15,500 binary and 1,500 continuous phenotypes contained in the AstraZeneca PheWAS Portal (https://azphewas.com/; downloaded on 26/08/2022). This portal contains genetic associations identified based on whole-exome sequencing data from ∼450,000 UK Biobank (UKB) participants.^58^ Binary phenotypes with <30 cases were excluded from both gene- and variant-level analysis. At the variant-level, associations were restricted to those identified in at least 30 samples. For gene-level and variant-level associations, we only extracted the most significant collapsing model and genotype model per trait, respectively. Statistical significance was defined as P-value <2e-09^58^, and suggestive significance as P-value <1e-05.

In addition to the PheWAS Portal queries, we used WES and biomedical data of the UKB (application number 64806) to investigate allelic series of functional QVs in *SLC13A1* and *SLC26A1* with hypothesized related clinical traits and diseases. We focused on *SLC13A1* variants for which experimental validation was available or that likely result in a severe consequence (stop gained, splicing) in order to select truly functional QVs. Among these, the stop gained variant p.Arg12*, for which a complete loss-of-function has experimentally been validated^43^, the stop gained substitution p.Trp48*, for which associations with decreased serum sulfate levels^44^ and skeletal phenotypes^41^ were reported, and the missense variant encoding p.Arg272Cys, located in a splice region, were available in the UKB. For *SLC26A1,* we selected QVs for which reduced sulfate transport activity had previously been shown^25^, of which p.Leu384Pro, p.Ser358Leu, and p.Thr185Met were available in the UKB. All 6 QVs passed the “90pct10dp” QC filter, defined as at least 90% of all genotypes for a given variant, independent of variant allele zygosity, had a read depth of at least 10 (https://biobank.ndph.ox.ac.uk/ukb/ukb/docs/UKB_WES_AnalysisBestPractices.pdf).

Analyses were performed on the UKB Research Analysis Platform. Participants with all ancestries were included into analysis but excluded strongly related individuals, defined as those that were excluded from the kinship inference process and those for whom ten or more third-degree relatives were identified. After individual-level filtering, a total of N=468,292 individuals remained for association analyses. Of these, there were 10 participants who were homozygous for one of the six QVs, and 7,280 persons heterozygous for at least one of the QVs. For these persons carrying at least one of the six QVs, we determined age- and sex-specific z-scores of their quantitative anthropometric measurements, enabling interpretation of their measurements compared to non-carriers of the same age and sex. Age- and sex-specific distributions were inverse normal transformed before calculating the z-scores.

We investigated the association between each of the resulting six functional QVs with medical diagnoses defined by International Classification of Diseases version-10 (ICD-10) codes based on UKB field 41202 (primary/main diagnosis codes across hospital inpatient records). We selected musculoskeletal diseases (ICD-10 codes starting with “M”), fractures and injuries (ICD-10 codes starting with “S” and containing “fracture”, “dislocation” or “sprain” terms). The association was examined using Fisher’s exact test under a dominant model, as well as through association analysis under additive model using Firth regression, as implemented in the “brglm2” R package^77^. We included sex, age at recruitment, sex*age, and first 20 genetic principal components (UKB field 22009) as covariates in the regression model. The association with quantitative anthropometric traits was assessed after inverse normal transformation via linear regression, additive genotype modeling and adjusting for the same covariates as with binary traits.

### Set-up of the whole-body model and mapping

The utilized WBM of human metabolism was built from genomic, biochemical and physiological data that originated from the generic genome-scale reconstruction of metabolism, Recon3D^23^. The sex-specific and organ-resolved WBM covers 13,543 unique metabolic reactions and 4,140 unique metabolites. The WBM was constrained as described previously^22,24^.

Of all observed significant gene-metabolite pairs from the GCKD study, 51 genes and 71 metabolites could be mapped onto RECON3D in total. For 37 of 51 genes, their associated metabolites could be mapped, resulting in 68 unique gene-metabolite pairs. To systematically investigate the consequences of genetic perturbations of gene *G*, we first identified all reactions *R*_*G*_ = {*r*_*G*1_,…, *r*_*Gn*_} that are carried out by the corresponding encoded enzymes across all organs in the WBM^78^. We included only genes in the generation of virtual IEMs that were exclusively causal for a non-empty set of reactions (i.e., for a gene *G*, associated with reactions *R*_*G*_ = {*r*_*G*1_,…, *r*_*Gn*_}, there did not exist a gene *H*, that was associated with any reaction of *R*_*G*_), and metabolites where urine excretion reactions were defined in the WBM reconstruction. From the initial 37 genes, 25 genes and their mapped metabolites fulfilled those criteria and were selected for the generation of 25 corresponding virtual IEMs.

### In silico knockout modeling via linear programming

Following the method of Thiele et al. ^22^, the knockout simulations were based on maximizing the excretion or demand reaction of the metabolite of interest under different conditions. In every optimization step, we assume steady state (***Sv*** = ***0***), where *S* is the stoichiometric matrix (rows: metabolites; columns: reactions), and ***v*** is the vector of fluxes through each reaction, adhering to specific constraints (***v***_*l*_ ≤ ***v*** ≤ ***v***_***u***_). This procedure, known as flux balance analysis (FBA)^79^, can be written as a linear programming (LP) problem:

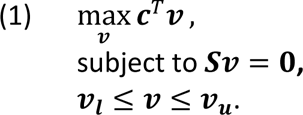

To model the impact of a gene-knockout on metabolite *M*, we maximized two key reactions: firstly, the urine excretion reaction of metabolite *M* (e.g., *EX*_*M*_), and secondly the created unbounded demand reaction (e.g., *DM*_*M*_[*bc*]), designed to reflect the accumulation of the metabolite *M* in the blood compartment. For simulating a wild-type model for gene *G*, we then solved the LP problem stated in (1), choosing the linear objective as the sum of all reactions across all organs catalyzed by the enzyme under consideration:

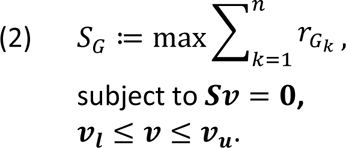

First, we checked if *S*_*G*_ > 10^−6^; a criterion implemented in the function checkIEM_WBM of the (PSCM) toolbox v.1.1^22^ for deciding whether the corresponding reactions can carry any flux, using the optimizeWBModel function of the COBRA toolbox^80^. Then, we unbound the upper bound of urine excretion, for each metabolite found to be significantly associated with gene *G*. Note that the blood demand reaction is unbounded by design. Next, we maximized the corresponding reactions of the metabolite biomarker *B*_*G*_ = {*b*_*GM*1_,…, *b*_*GMm*_}, as the LP-problem stated in (1) under the additional constraint that 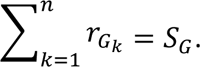 This procedure delivers two flux values – the maximal urine excretion and the maximal flux into blood given the constraint setting. Finally, to simulate the complete loss of function, we blocked all reactions in all organs catalyzed by gene *G* by setting their lower and upper bound to zero: *r*_*G*1_ = ⋯ = *r*_*Gn*_ = 0. As in the wild-type model, we then removed the upper bound of the urine excretion reaction and maximize the corresponding reactions *B*_*G*_ = {*b*_*GM*1_,…, *b*_*GMm*_}. Analogously, we derived two flux values as in the wild-type model. Subsequently, one can observe whether the knockout results in an increase, decrease or equal outcome in terms of fluxes into the blood or urine compartment for each metabolite that could be mapped in the WBM and that was found to be significantly associated with gene *G* in the GCKD cohort.

Following that paradigm, we were initially able to compute 25 virtual IEMs and modeled 59 gene-metabolite pairs in urine and blood. After curation of the male and female model, 67 gene-metabolite pairs could be computed. Curation details can be found in the **Supplementary Methods**.

LP-simulations were carried out under Windows10 using Matlab2021a (Mathworks, Inc.) as simulation environment, Ilog Cplex v10.09 (IBM, Inc.) as linear programming solver, the COBRA Toolbox v.3.4^80^, and the physiologically and stoichiometrically constrained modeling (PSCM) toolbox v.1.1^22^.

### Microbiome personalization of whole-body models

Microbiome personalized WBMs were generated by creating community models based on the genome-scale reconstructions of microbes in the AGORA1 resource^81^. Briefly, from microbe identification and relative abundance data of a metagenomic sample, the genome-scale reconstructions of the identified microbes are joined to form a microbial community that is connected via a lumen compartment, where they can exchange metabolites^82,83^. These community models can then be integrated into the WBM, personalizing the WBM according to the underlying metagenomics data^22^. Each microbial community model is connected to the WBM by connecting the microbiota lumen compartment to the large intestinal lumen of the WBM. Microbial community models (n=616) were based on publicly available metagenomics data from Yachida et al.^27^, and then embedded into the male WBM to form 616 personalized WBMs.

### In silico knockout modeling using quadratic programming

While maintaining the same conditions as outlined in (1), rather than maximizing a linear objective, we minimized a quadratic objective for each personalized WBM, as well as regulated the squared Euclidean norm of the solution vector *v*:

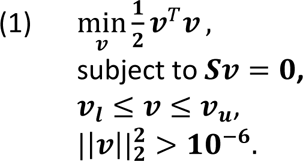

Because *f*(***v***) = ***v***^*T*^***v*** is a strictly convex function and the feasible set is convex, the solution of (3) is unique if it exists. This inherent uniqueness allows for the calculation of a unique distribution of fluxes, in contrast to a single flux maximum achieved through LP. The last condition in (3) is for regularization, where we chose the value 10^−6^, recommended in the COBRA Toolbox^80^. For each solution ***v***, we obtained the corresponding urine excretion reactions of the metabolites that were significantly associated with *KYNU* in the GCKD study. For knockout simulations, the associated reactions of *KYNU* were set to zero (*r*_*KYNU1*_ = ⋯ = *r*_*KYNUn*_ = 0) and the optimization problem stated in (3) was solved if possible. A QP-solution could be computed for 593 wild-type WBMs and for 592 knockout WBMs. For the remaining models, the QP-solver was not able to compute a solution. Considering samples for which a wild-type and knockout solution was available resulted in 582 paired wild-type / knockout WBM pairs. All urine secretion flux values were obtained from the unique QP-solution vector, including secretion fluxes for 242 metabolites covered in the GCKD urine metabolome data. The QP-simulations were carried out utilizing the high performance computing facility, called the Brain-Cluster, at the University Greifswald employing MATLAB 2019b (MathWorks, Inc.), ILOG CPLEX v10.10 (IBM, Inc.) as quadratic programming solver, and the COBRA Toolbox v.3.4^80^.

### Statistical analysis of the in silico simulation results

An extension of Fisher’s exact test for 2×3 contingency tables (Fisher-Freeman-Halton test) was used to determine significance when comparing the *in vivo* and *in silico* signs from LP-modeling. For statistical analysis of the paired 582 microbiome-personalized WBMs, we performed for each of the 242 mapped urinary metabolites a fixed effect linear regression using the ln(urine secretion flux) as response variables, the knockout status as the sole predictor (wild-type vs. knockout), and the personalized microbiome as a fixed effect. Significance of the effect of the knockout was then tested, with the significance threshold set to 0.05/242 (Bonferroni correction). Importantly, the entire variance in the regression models had two sources: 1) the knockout, 2) the microbiome personalization. Significance testing of the *in silico* regression coefficient of the knockout variable therefore delivers a test whether the knockout explains substantial amounts of variance, in comparison to the variance induced by randomly sampled microbiome communities. The *in silico* regression coefficients were then correlated with the burden-derived observed regression coefficients of gene-metabolite associations from the GCKD study, and significance was determined through the standard test for Pearson correlations.

### Absolute metabolite quantification for members of the family with the KYNU-attributed IEM

Kynurenate, 8-methoxykynurenate, xanthurenate, kynurenine and 3-hydroxykynurenine were quantified in urine samples using high performance liquid chromatography coupled to tandem mass spectrometry (HPLC/MS/MS; Exion LC and 5500+ triple quadrupole MS, AB Sciex, Framingham, MA, USA). Urine samples were diluted 1:10 with water and 10 µL of the diluted samples were injected. HPLC separation was performed at 40 °C on a Force C18 column (100 x 3.0 mm, 3 µm particles, Restek Corporation, Bellefonte, PA, USA) equipped with guard column using water (solvent A) and methanol (solvent B), both containing 0.01 vol% formic acid and 1 mM ammonium formate. The flow rate was 300 µL/min and the linear gradient profile of solvent B was as follows: 0 min 1%, 1 min 1%, 10 min 40%, 12 min 90%, then isocratic at 90% until re-equilibration. The analytes were detected using positive ion electrospray ionization (5500 V and 350 °C, nitrogen curtain and ion source gas, declustering potential 1.0 V, entrance potential 10 V) and the multiple reaction monitoring mode (nitrogen collision gas). Compound specific MS parameters are given in **Table 1**.

**Table 1.** Mass spectrometric parameters for detection and quantification of the analytes.

|  |  | Precursor ion<br>[m/z] | Product ion<br>[m/z] | Collision<br>energy [V] | Collision cell exit<br>potential [V] |
| --- | --- | --- | --- | --- | --- |
| Kynurenate | Quantifier | 190.0 | 144.1 | 29 | 10 |
|  | Qualifier | 190.0 | 116.1 | 43 | 12 |
| 8-methoxy-<br>kynurenate | Quantifier | 220.0 | 174.1 | 27 | 12 |
|  | Qualifier | 220.0 | 118.1 | 39 | 14 |
| Xanthurenate | Quantifier | 206.0 | 160.1 | 27 | 12 |
|  | Qualifier | 206.0 | 132.1 | 39 | 10 |
| Kynurenine | Quantifier | 209.0 | 94.1 | 19 | 10 |
|  | Qualifier | 209.0 | 146.1 | 29 | 10 |
| 3-hydroxy<br>kynurenine | Quantifier | 225.0 | 162.1 | 29 | 10 |
|  | Qualifier | 225.0 | 110.1 | 19 | 10 |

Quantification was based on external 4-point calibration curves covering the ranges of detected signal abundances in the samples. Quantitative results were normalized to urine creatinine concentrations (expressed as mmol/mol creatinine) before comparison between samples.

### External data sources

To look for gene expression and QTLs across tissues, we used data from the GTEx Project (https://gtexportal.org/home/). The AstraZeneca PheWAS Portal (https://azphewas.com/) was used to search for gene- and variant-level associations of detected genes and QVs.GTEx Project (https://gtexportal.org/home/): investigation of gene expression and QTLs across tissues; AstraZeneca PheWAS Portal (https://azphewas.com/): search for gene- and variant-level associations of detected genes and QVs; OMIM catalog (https://www.omim.org/): query for monogenic disorders and traits related to identified genes; Genomics England PanelApp (https://panelapp.genomicsengland.co.uk/panels/467/ version v3.0): search for known IEM related to the detected genes; Open Targets Platform (https://platform.opentargets.org/): search for drug target status and corresponding indication for identified genes; ClinVar archive (https://www.ncbi.nlm.nih.gov/clinvar/): query for clinical significance and corresponding trait/disease of detected QVs.

## Data Availability

All data produced in the present work are contained in the manuscript.

## Acknowledgements

The work of N.S., M.S., O.B., M.W., M.K., Pe.S., and A.K. was funded by German Research Foundation (DFG) project ID 431984000 (SFB 1453). N.S. and Y.L. were supported by DFG KO 3598/4-2 (to A.K.). Germany’s Excellence Strategy (CIBSS, EXC-2189, project ID 390939984) supported the work of M.K., M.S., and A.K.. The work of J.H., D.F., and A.K. was supported by DFG project ID 499552394 (SFB 1597/1). S.P. was funded by H2020 MSCA-ITN-2019 ID:860977 (TrainCKDis). Pe.S. was supported by DFG SE 2407/3-1. The work of U.T.S. was supported by the German Federal Ministry of Education and Research (BMBF) within the framework of the e:Med research and funding concept (grant 01ZX1912B). The work of Pa.S. was supported by DFG Project-ID 1050086601 (SCHL 2292/2-1). I.T. was funded by the European Research Council (ERC) under the European Union’s Horizon 2020 research and innovation programme (#757922), the Science Foundation Ireland under Grant number 12/RC/2273-P2, and a Horizon Europe grant (#101080997).

Genotyping and urine metabolomics in the GCKD study were supported by Bayer Pharma. Plasma metabolomics has received funding from the Innovative Medicines Initiative 2 Joint Undertaking (JU) under grant agreement no. 115974. The JU receives support from the European Union’s Horizon 2020 research and innovation program and the EFPIA and the JDRF. Any dissemination of results reflects only the authors’ view; the JU is not responsible for any use that may be made of the information it contains. The GCKD study was and is supported by the BMBF (FKZ 01ER 0804, 01ER 0818, 01ER 0819, 01ER 0820 and 01ER 0821) and the KfH Foundation for Preventive Medicine. Unregistered grants to support the study were provided by corporate sponsors (listed at https://gckd.org). We are grateful for the willingness of the patients to participate in the GCKD study. The enormous effort of the study personnel of the various regional centers is highly appreciated. We thank the large number of nephrologists who provide routine care for the patients and collaborate with the GCKD study. The GCKD investigators are listed in the Supplementary Note.

