## Supplementary_Materials for "Coupling of metabolomics and exome sequencing reveals graded effects of rare damaging heterozygous variants on gene function and resulting traits and diseases"

### Supplementary Methods

*Whole-body modeling*

The implicated genes' loss-of-function were investigated in virtual IEMs generated through organ-resolved sex-specific whole-body models (WBMs) based on the Virtual Metabolic Human database (VMH)[1] using a constraint-based modeling and reconstruction analysis (COBRA) approach[2]. Mapping the gene-metabolite pairs significant in the genome-wide screening onto the VMH database[3], virtual IEMs were created to explore all represented gene-metabolite pairs via *in silico* knockout modeling of the gene's function. For modeling of the male human, the WBM model version "Harvey_104b" was utilized, for modeling the female model, the WBM model version "Harvetta_104c" was employed.

### Supplementary Results

*Curation of whole model modeling based on the GCKD data*

To leverage the biological information generated by the WES-metabolite association data from the GCKD study for improving the knowledge base underlying the WBM, we performed a range of model curation steps. These curation steps ranged from adding pathways over improved mapping and checking failing simulations to altering model constraints. The following paragraphs detail all performed model curations. We performed curations for six virtual IEMs, where we could identify reasons for model failure (e.g., in the case of *DMGDH*) or where the GCKD data was instrumental in improving the knowledge base (e.g., in the case of *KYNU* and 8-methoxykynurenate).

*Modeling of 8-methoxykynurenate in the virtual IEM for kynureninase deficiency (KYNU)*

Although a known human metabolite, the metabolite 8-methoxykynurenate was not included in the initial WBMs due to limited evidence on the enzymes involved in its production. However, in the association results from the GCKD study, urine 8-methoxykynurenate was positively associated with rare, putatively damaging variants in *KYNU.* This indicates that this metabolite originates upstream of a reaction catalyzed by kynureninase. As 8-methoxykynurenate is a methylated derivative of xanthurenate, it is plausibly generated by a corresponding methylation reaction as noted in KEGG (KEGG reaction R03955; Xanthurenic acid + S-adenosyl-L-methionine <=> 8-methoxykynurenate + S-adenosyl-L-homocysteine). Interestingly, we found *ASMTL*, a gene encoding for a protein with presence of a probable catalytic S-adenosyl-L-methionine binding domain in the C-terminal region and thus a probable methyltransferase, to be negatively associated with urine 8-methoxykynurenate (P-value=5.1e-09), which barely missed the study-wide multiple-testing corrected significance threshold. On these grounds, we added 8-methoxykynurenate (C05830) along with the (hypothesized) associated methylation reaction (Xanthurenic acid + S-adenosyl-L-methionine <=> 8-Methoxykynurenate + S-adenosyl-L-homocysteine) and corresponding transport reactions to the ten organs of the WBMs (**Supplementary Table 9**), where the participating metabolites of the methylation reaction were all present. We then repeated the *in silico* knockout of *KYNU*, and successfully replicated the association of *KYNU* with higher flux of 8-methoxykynurenate into urine compared to the wild-type.

*Modeling of N-formylanthranilic acid in the virtual IEM for AFMID*

Both N-formylanthranilic acid and the *AFMID* gene were represented in the initial WBM. However, the urinary secretion of N-formylanthranilic acid could not carry flux in the initial simulations. Investigating the model setup for N-formylanthranilic acid, we found that for the transport reaction from the blood compartment to the kidney (WBM reaction name: Kidney_EX_nformanth(e)_[bc]) under the current default constraint setting (lower bound=-3.7368, upper bound=0) any flux of N-formylanthranilic into the kidney compartment was blocked. Consequently, no excretion process into urine could occur. As N-formylanthranilic acid is, however, detected in human urine, we made corresponding adjustments to the constraint setting, allowing N-formylanthranilic acid to be secreted into urine. After this adjustment, the model correctly predicted the observed association between rare, damaging variants in *AFMID* and urine N-formylanthranilic acid levels in the GCKD study. Both the initial and the curated virtual IEM correctly predicted the observed association between rare, damaging *AFMID* variants and plasma N-formylanthranilic acid levels.

*Modeling of the virtual IEM for TMLHE*

*TMLHE* is encoding for the enzyme trimethyllysine dioxygenase, which utilizes N6,N6,N6-trimethyl-L-lysine as one of its substrates. While *TMHLE* had been included in the initial version of the WBM, none of the metabolites that were associated with it in the GCKD study could be modeled. We found that in the initial WBM, N6,N6,N6-trimethyl-L-lysine was neither produced from methylated protein-bound lysine residuals, nor was it covered by dietary constraints, meaning that trimethyllysine dioxygenase reactions could not carry flux. To enable modeling, we unbound the diet constraint for N6,N6,N6-trimethyl-L-lysine[4], making N6,N6,N6-trimethyl-L-lysine available to the WBM. After this step, the virtual IEM for *TMLHE* was perfectly predicting the signs of the observed *TMLHE*-metabolite associations in the GCKD study.

*Modeling of dimethylglycine in the virtual IEM for dimethylglycine dehydrogenase deficiency (DMGDH)*

Both, dimethylglycine and the gene *DMGDH* could be mapped in the initial WBM. However, knockout of *DMGDH* had no effect on dimethylglycine blood and urine secretion fluxes in the female model, and no effect on urine secretion in the male model. Exploring the gene-protein-reaction relations in the initial WBM, we found three reactions assigned to *DMGDH* (mitochondrial dimethylglycine dehydrogenase (VMH ID: DMGDHm), N,N-dimethylglycine:electron-transfer flavoprotein oxidoreductase (VMH ID: HMR_4700), and S-adenosyl-L-methionine:sarcosine N-methyltransferase (VMH ID: HMR_4701)). To the latter two reactions, the gene *PDPR*, encoding for a regulatory subunit of the pyruvate dehydrogenase phosphatase, was assigned as well. We removed the assignment to *PDPR,* as we could not find additional evidence for *PDPR* playing a role in dimethylglycine metabolism besides a distant relation in terms of sequence similarity to *DMGDH*[5]. After removing *PDPR* as a hypothetical isozyme for the reactions HMR_4700 and HMR_4701, the virtual IEM for *DMDGH* correctly predicted the observed effect direction for dimethylglycine both in blood and urine and in both sexes.

*Modeling of the virtual IEM for KYAT1*

In the initial WBM, we were unable to map the *KYAT1* gene, although it was actually included in the model, due to an identifier discrepancy. We rectified this by adding the corresponding identifier for *KYAT1* in the VMH database (VMH gene identifier: 883), which increased the number of mapped and modeled genes to 26. Three of the metabolites associated with *KYAT1* in the GCKD study, 3-(4-hydroxyphenyl)lactate, indolelactate, and phenylpyruvate, could be mapped in the WBMs and two, 3-(4-hydroxyphenyl)lactate and phenylpyruvate, could be modeled. However, *KYAT1* knockout did not replicate the observed effects from the GCKD study, indicating that further curation of the WBMs is needed in the case of *KYAT1.*

*Modeling of hexanoylglycine in the virtual IEM for medium-chain acyl-CoA dehydrogenase deficiency (ACADM)*

In the wild-type and knockout *ACADM* models, we initially calculated maximal secretion fluxes for hexanoylglycine into urine. However, the result was consistently a maximum secretion flux of zero for all simulations. Upon exploration, we found that none of the hexanoylglycine-related reactions carried flux in the current WBM. Thus, the metabolite fails the criteria of being transported to blood and urine, and the current WBM is unable to model the *ACADM*-hexanoylglycine gene-metabolite pair. The initial flux calculations of zero were therefore without biological meaning.

### Supplementary Figures

**Supplementary Figure 1**: Plasma metabolite levels among carriers and non-carriers of QVs in significantly associated genes

Plasma metabolite levels after inverse normal transformation are shown on the y-axis, among non-carriers and carriers of QVs in both masks (LoF_mis and HI_mis) on the x-axis. Symbol color and shape indicate a variant's driver status (Methods) and consequence, respectively. Carriers of multiple heterozygous QVs are denoted by an asterisk. Orange filling of symbols denotes homozygosity for the respective QV. The boxes range from the 25$^{th}$ to the 75$^{th}$ percentile of metabolite levels, the median is indicated by a line, and whiskers end at the last observed value within 1.5*(interquartile range) away from the box.

**Supplementary Figure 2**: Urine metabolite levels among carriers and non-carriers of QVs in significantly associated genes

Urine metabolite levels after inverse normal transformation are shown on the y-axis, among non-carriers and carriers of QVs in both masks (LoF_mis and HI_mis) on the x-axis. Symbol color and shape indicate a variant's driver status (Methods) and consequence, respectively. Carriers of multiple heterozygous QVs are denoted by an asterisk. Orange filling of symbols denotes homozygosity for the respective QV. The boxes range from the 25$^{th}$ to the 75$^{th}$ percentile of metabolite levels, the median is indicated by a line, and whiskers end at the last observed value within 1.5*(interquartile range) away from the box.

**Supplementary Figure 3**: Contribution of individual QVs to their gene-based association signal with plasma metabolite levels

For each significant gene-metabolite pair in plasma (sorted by gene and metabolite's biochemical name), the symbols visualize the $-\log_{10}$(P-value) (y-axis) for the successive aggregation of the most influential QVs with respect to the forward selection procedure (Bomba et al. 2022) for both masks (LoF_mis, HI_mis). The number of QVs aggregated for burden testing is given on the x-axis. Symbol shape indicates the variant's consequence. The symbol color and size reflect the effect size and the P-value of the variant based on its single-variant association test. The gray dashed lines represent the significance threshold ($-\log_{10}$(5.04e-9)), the total $-\log_{10}$(P-value) of the aggregate variant test including all QVs in the respective gene and mask, and the $-\log_{10}$(lowest P-value) that can be reached by aggregating only the driver variants from the forward selection procedure.

**Supplementary Figure 4**: Contribution of individual QVs to their gene-based association signal with urine metabolite levels

For each significant gene-metabolite pair in urine (sorted by gene and metabolite's biochemical name), the symbols visualize the $-\log_{10}$(P-value) (y-axis) for the successive aggregation of the most influential QVs with respect to the forward selection procedure (Bomba et al. 2022) for both masks (LoF_mis, HI_mis). The number of QVs aggregated for burden testing is given on the x-axis. Symbol shape indicates the variant's consequence. The symbol color and size reflect the effect size and the P-value of the variant based on its single-variant association test. The gray dashed lines represent the significance threshold ($-\log_{10}$(4.46e-9)), the total $-\log_{10}$(P-value) of the aggregate variant test including all QVs in the respective gene and mask, and the $-\log_{10}$(lowest P-value) that can be reached by aggregating only the driver variants from the forward selection procedure.

**Supplementary Figure 5**: Driver variants show a more severe impact on metabolite levels compared to non-drivers in terms of consequence and effect size

**(a)** The bar plot represents the absolute frequency (y-axis) of each of the QVs' consequences with their proportions noted next to them, separately for driver and non-driver variants (x-axis). In case one gene was significantly associated with levels or more than one metabolite, only the QVs from the strongest gene-metabolite associations are included (for only one matrix and only one mask) to prevent counting variants multiple times. Whereas driver variants contain more splicing, stop-gain and frameshift variants, the proportion of missense variants is higher among non-driver variants (Fisher's exact test: P-value=1.3e-6).

**(b)** The swarm plot shows differences in absolute effect sizes for QVs (y-axis) across the 5 different consequence classes (x-axis). The color reflects the variant status (driver versus non-driver variant) and the horizontal lines represent the median of the absolute effect sizes separately for driver and non-driver variants. In case one gene was significantly associated with levels or more than one metabolite, only the QVs from the strongest gene-metabolite associations are included. The median among driver variants increases when ordering the consequence classes with respect to severity (missense, start/stop lost, frameshift, stop gained, splicing).

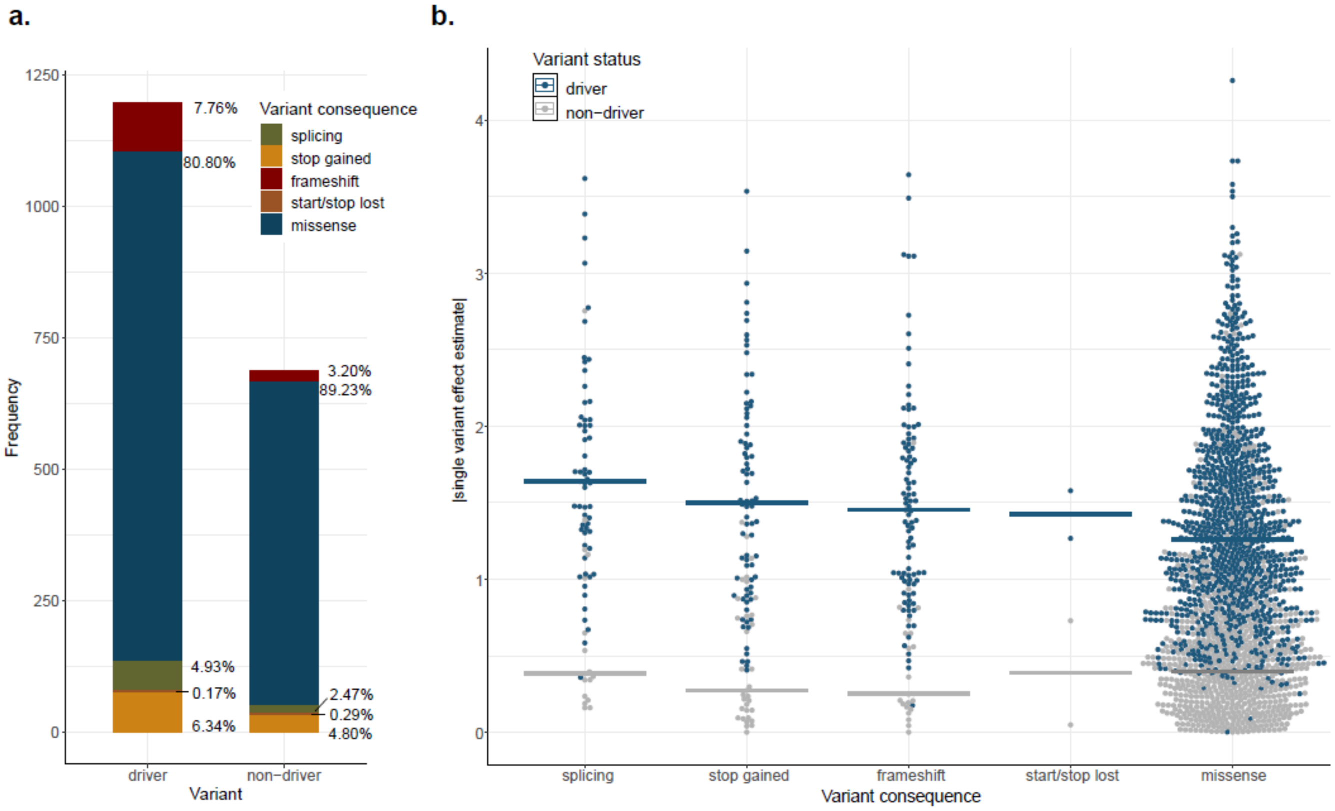

**Supplementary Figure 6**: Effect sizes of rare and common variant association signals with metabolite levels within the same locus

The scatter plot shows the absolute effect size (y-axis) of association signals with metabolite levels based on aggregating rare variants within a gene and based on common variants within the same locus (±500 kb around the gene), across different cumulative minor allele frequencies (cMAF, for aggregated rare variants) and minor allele frequencies (MAF, for common variants) (x-axis). Colors indicate whether the corresponding association signal is based on shared rare or common variants or whether it is unique to the rare variant screen. The shape represents the matrix of the corresponding metabolite. The absolute effect size tends to increase with decreasing MAF/cMAF.

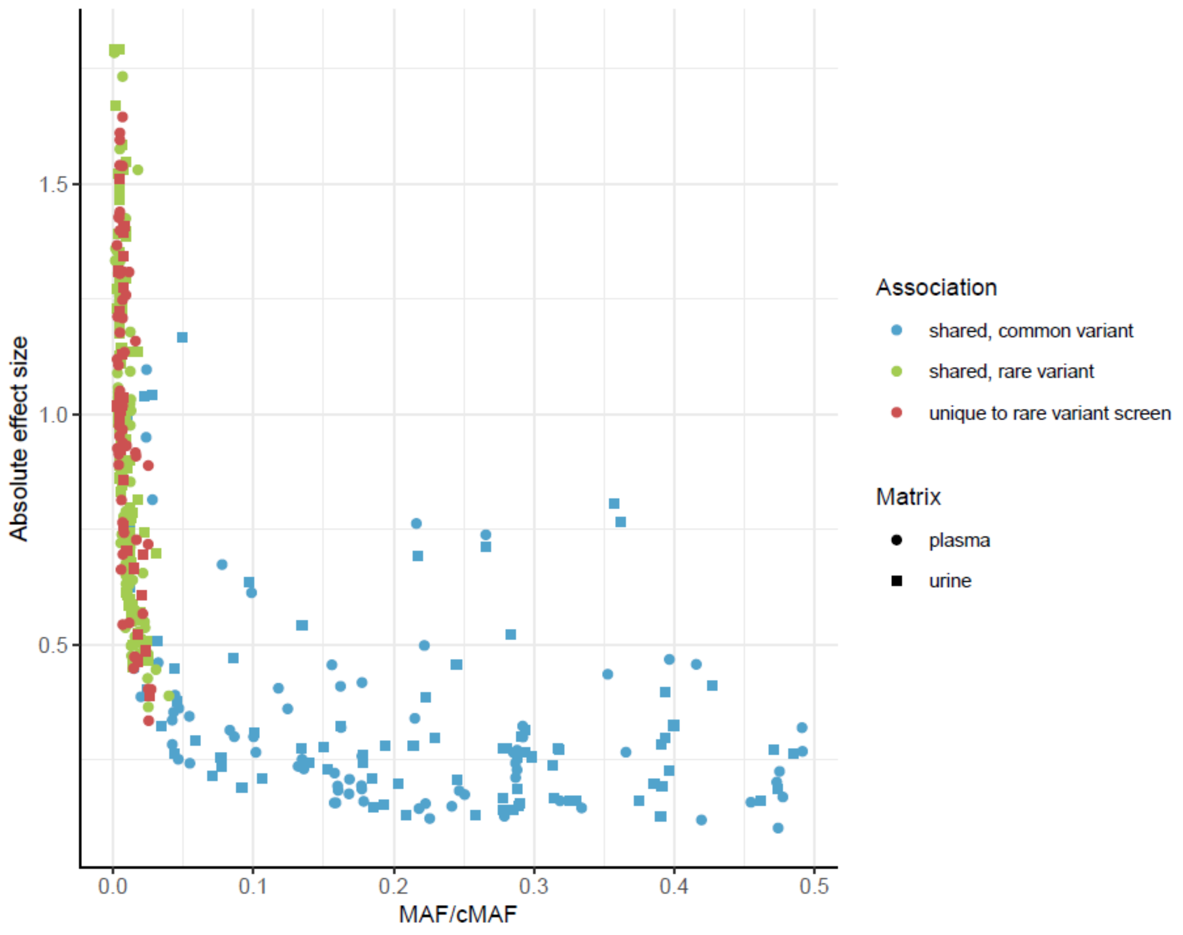

**Supplementary Figure 7**: Elevated urine levels of 3-hydroxykynurenine and xanthurenate are a readout of impaired *KYNU* function: converging evidence from three approaches

Three panels are shown for 3-hydroxykynurenine **(a)** and xanthurenate **(b)** each: the left panel represents inverse-normal transformed urine levels of the respective metabolite (y-axis) among non-carriers and carriers of QVs in *KYNU* (x-axis). Units correspond to standard deviations. The boxes range from the 25$^{th}$ to the 75$^{th}$ percentile of metabolite levels, the median is indicated by a line, and whiskers end at the last observed value within 1.5*(interquartile range) away from the box. The middle panel represents the distribution of the ln-transformed urinary secretion flux of the respective metabolite in mmol/day into urine (y-axis) from min-norm simulations based on 582 microbiome-personalized whole-body models without and with simulated knockout of *KYNU* (x-axis). The right panel shows multiple reaction monitoring (MRM, *m/z* 225.0 → 162.1, 206.0 → 160.1) chromatograms of the diluted urines of a child with a homozygous, autosomal-recessively inherited loss of *KYNU* function (patient), the mother and the father. The signals at 3.9 min (3-hydroxykynurenine) and 9.5 min (xanthurenate) are strongly enhanced in the patient sample. Chromatograms are normalized to urine creatinine concentrations; y-axes are normalized to the intensity of the signals in the patient's chromatograms. All three independent approaches arrive at the conclusion that elevated levels of 3-hydroxykynurenine and xanthurenate in urine are a readout of impaired *KYNU* function.

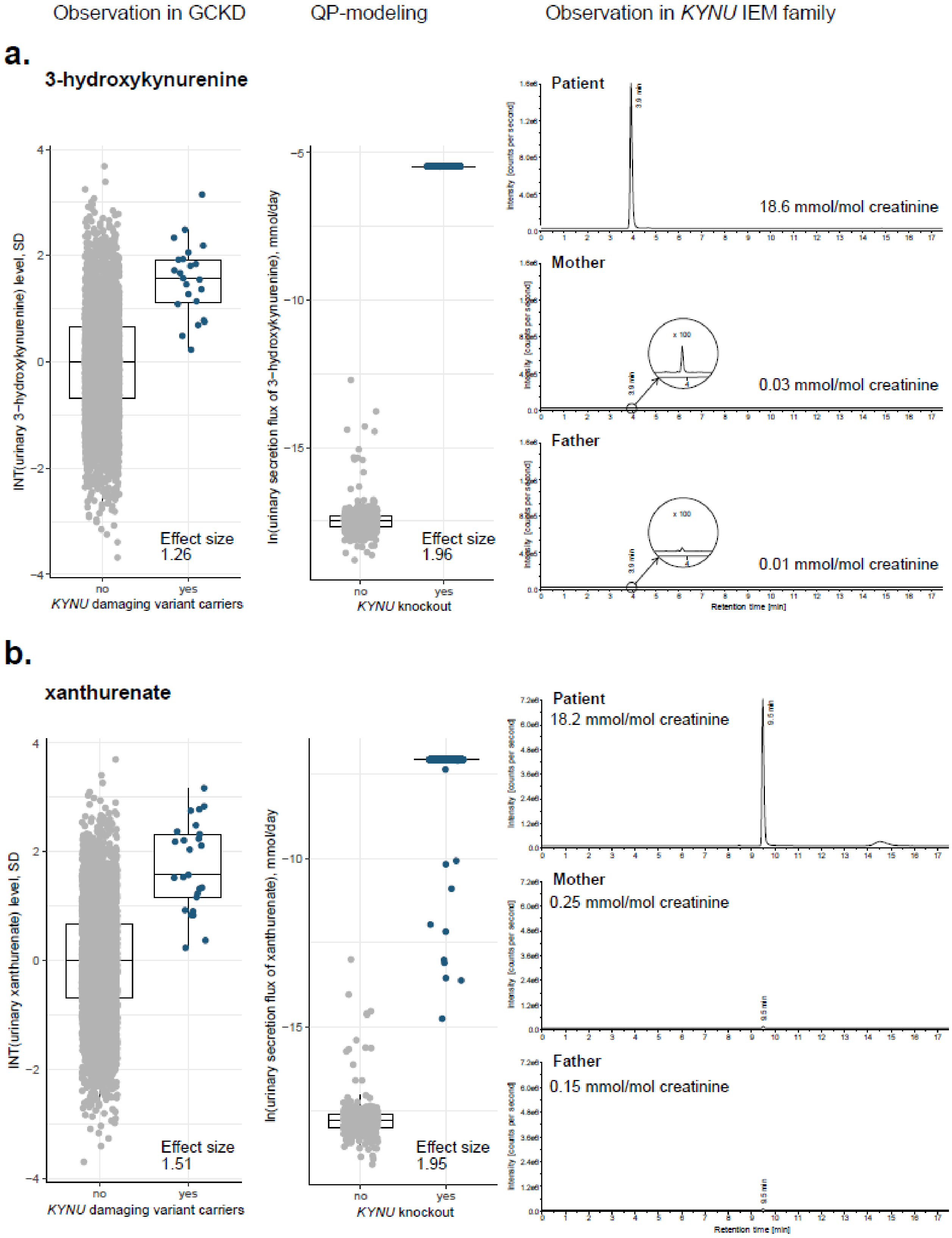
Observation in GCKD
QP-modeling
Observation in KYNU IEM family
a.
3-hydroxykynurenine
INT(urinary 3−hydroxykynurenine) level, SD
Effect size 1.26
no
yes
KYNU damaging variant carriers
ln(urinary secretion flux of 3−hydroxykynurenine), mmol/day
Effect size 1.96
KYNU knockout
Patient
18.6 mmol/mol creatinine
Mother
0.03 mmol/mol creatinine
Father
0.01 mmol/mol creatinine
Retention time [min]
b.
xanthurenate
INT(urinary xanthurenate) level, SD
Effect size 1.51
KYNU damaging variant carriers
ln(urinary secretion flux of xanthurenate), mmol/day
Effect size 1.95
KYNU knockout
Patient
18.2 mmol/mol creatinine
Mother
0.25 mmol/mol creatinine
Father
0.15 mmol/mol creatinine
Retention time [min]

**Supplementary Figure 8:** Contribution of individual QVs in *SLC26A1* to their gene-based association signal with plasma sulfate levels

The symbols visualize the -$\log_{10}$(P-value) (y-axis) with regard to plasma sulfate levels for the successive aggregation of the most influential QVs in *SLC26A1* with respect to the forward selection procedure (Bomba et al. 2022) for the mask LoF_mis. The number of QVs aggregated for burden testing is shown on the x-axis. Symbol shape indicates the variant's consequence. The symbol color and size reflect the effect size and the P-value of the variant based on its single-variant association test. The gray dashed lines represent the significance threshold (-$\log_{10}$(0.05)), the total -$\log_{10}$(P-value) of the aggregate variant test including all QVs in *SLC26A1* for the mask LoF_mis, and the -$\log_{10}$(lowest P-value) that can be reached by aggregating only the driver variants from the forward selection procedure. Summary statistics shown on the right refer to the burden tests aggregating all QVs and only driver variants. For the latter, a clear association of *SLC26A1* wit plasma sulfate levels is observed.

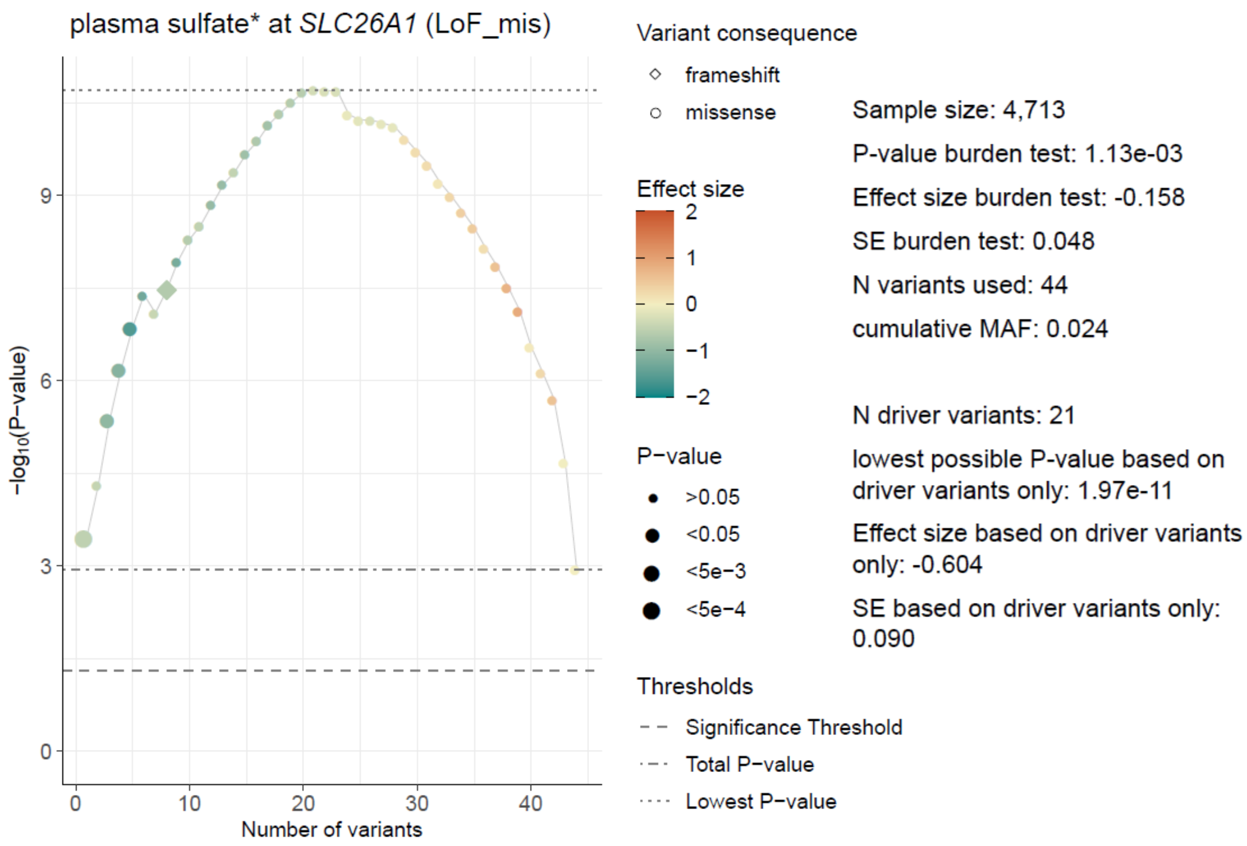

**Supplementary Figure 9**: Contribution of individual QVs in *SLC13A1* and *SLC26A1* to their gene-based association signal with height measured in the GCKD study

The symbols visualize the -$\log_{10}$(P-value) (y-axis) with regard to height measured in 3,239 participants of the GCKD study for the successive aggregation of the most influential QVs in *SLC13A1* (mask HI_mis) **(a)** and *SLC26A1* (mask LoF_mis) **(b)** with respect to the forward selection procedure (Bomba et al. 2022). The number of QVs aggregated for burden testing is shown on the x-axis. Symbol shape indicates the variant's consequence. The symbol color and size reflect the effect size and the P-value of the variant based on its single-variant association test. The gray dashed lines represent the significance threshold (-$\log_{10}$(0.05)), the total -$\log_{10}$(P-value) of the aggregate variant test including all QVs in *SLC13A1* and *SLC26A1* for the respective mask, and the -$\log_{10}$(lowest P-value) that can be reached by aggregating only the driver variants from the forward selection procedure. For both genes, a clear association with height in the GCKD study is observed when aggregating driver variants.

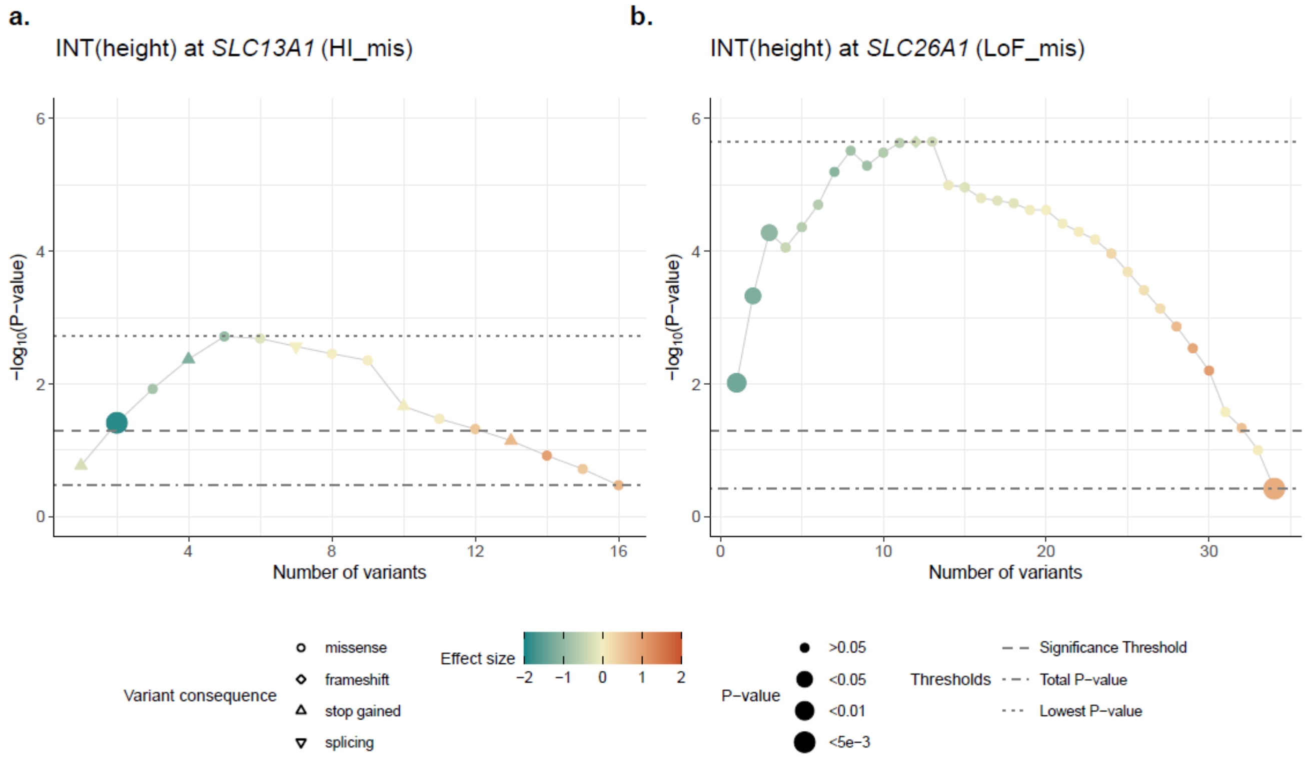

**Supplementary Figure 10**: Impact of different genotypes encoding NaS1 p.Arg12* and SAT1 p.Leu348Pro on height and musculoskeletal traits and fractures

The boxplots on the left show differences in age- and sex-specific z-scores for standing height (y-axis) across persons heterozygous and homozygous for the NaS1 p.Arg12*-encoding allele **(a)** and for the Sat1 p.Leu348Pro-encoding allele **(b)** (x-axis). Persons carrying a variant at two different DNA positions are shown in the category "het + X". For the NaS1 p.Arg12* stop gained variant, multi-heterozygous individuals who additionally carry the NaS1 p.Trp48* stop gained variant are indicated with differently shaped symbols, emphasizing that carrying two stop gained variants in NaS1 seems to lead to a more severe phenotype.

The forest plots on the right show associations between the NaS1 p.Arg12* **(c)** and SAT1 p.Leu348Pro **(d)** carrier status with those musculoskeletal diseases and fractures from the UKB, for which at least 2 carriers were identified (y-axis). Odds ratios (x-axis) are based on a Firth regression. The symbol color reflects the -$\log_{10}$(P-value), the size the number of variant carriers with disease. Only associations with P-value<0.05 are shown.

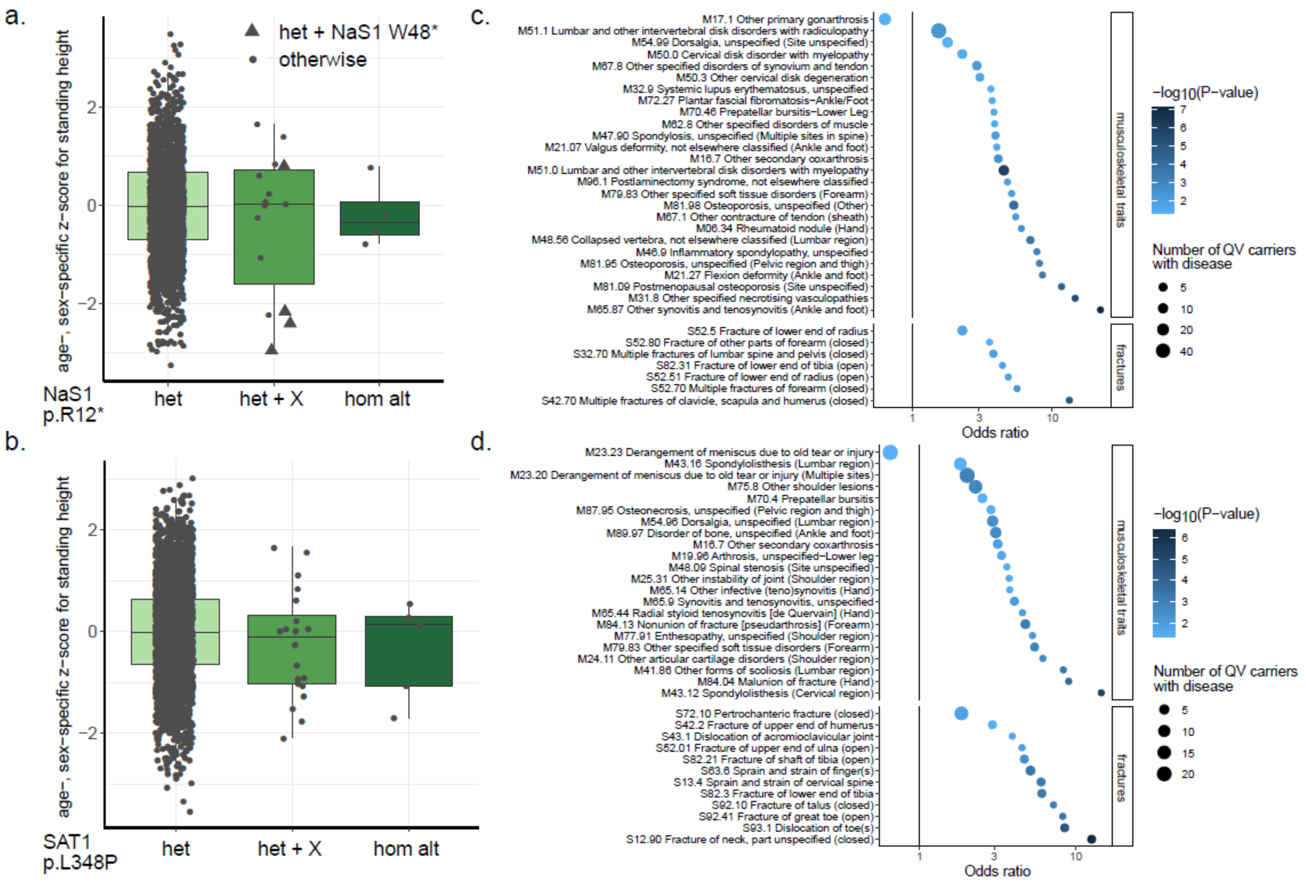

### Supplementary Acknowledgements

List of GCKD Study Investigators

A list of nephrologists currently collaborating with the GCKD study is available at http://www.gckd.org.

| | |
|---|---|
| University of Erlangen-Nürnberg | Kai-Uwe Eckardt, Heike Meiselbach, Markus Schneider, Mario Schiffer, Hans-Ulrich Prokosch, Barbara Bärthlein, Andreas Beck, André Reis, Arif B. Ekici, Susanne Becker, Ulrike Alberth-Schmidt, Anke Weigel, Sabine Marschall, Eugenia Schefler |
| University of Freiburg | Gerd Walz, Anna Köttgen, Ulla Schultheiß, Fruzsina Kotsis, Simone Meder, Erna Mitsch, Ursula Reinhard |
| RWTH Aachen University | Jürgen Floege, Turgay Saritas, Alice Groß |
| Charité, University Medicine Berlin | Elke Schaeffner, Seema Baid-Agrawal, Kerstin Theisen |
| Hannover Medical School | Hermann Haller |
| University of Heidelberg | Martin Zeier, Claudia Sommerer Mehtap Aykac |
| University of Jena | Gunter Wolf, Martin Busch, Rainer Paul |
| Ludwig-Maximilians University of München | Thomas Sitter |
| University of Würzburg | Christoph Wanner, Vera Krane, Antje Börner-Klein, Britta Bauer |
| Medical University of Innsbruck, Division of Genetic Epidemiology | Florian Kronenberg, Julia Raschenberger, Barbara Kollerits, Lukas Forer, Sebastian Schönherr, Hansi Weissensteiner |
| University of Regensburg, Institute of Functional Genomics | Peter Oefner, Wolfram Gronwald |
| Department of Medical Biometry, Informatics and Epidemiology (IMBIE), University of Bonn | Matthias Schmid, Jennifer Nadal |
