## Supplementary_Figure1 for "Coupling of metabolomics and exome sequencing reveals graded effects of rare damaging heterozygous variants on gene function and resulting traits and diseases"

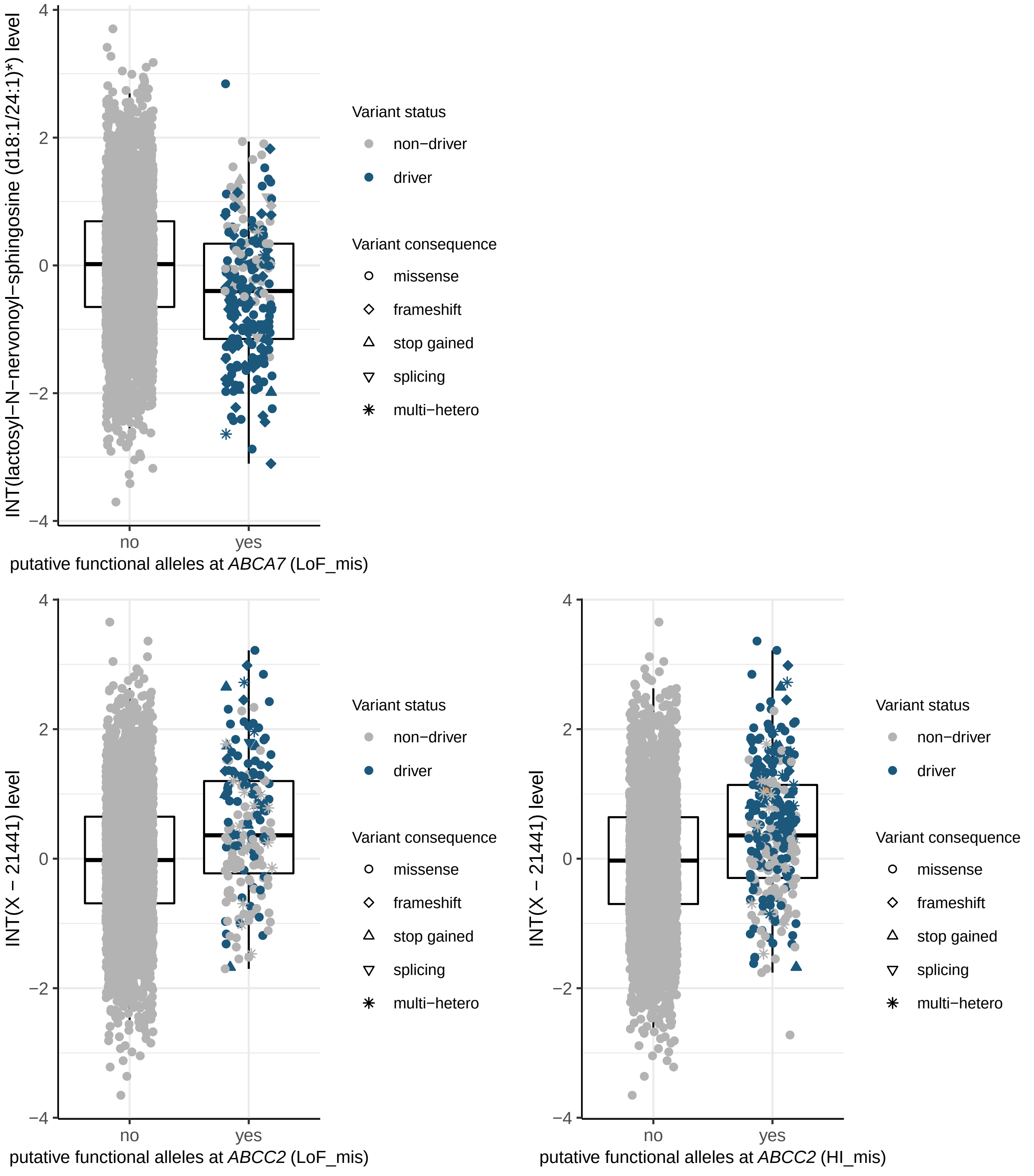

INT(lactosyl−N−nervonoyl−sphingosine (d18:1/24:1)*) level
putative functional alleles at ABCA7 (LoF_mis)
Variant status
non−driver
driver
Variant consequence
missense
frameshift
stop gained
splicing
multi−hetero
INT(X − 21441) level
putative functional alleles at ABCC2 (LoF_mis)
INT(X − 21441) level
putative functional alleles at ABCC2 (HI_mis)
no
yes

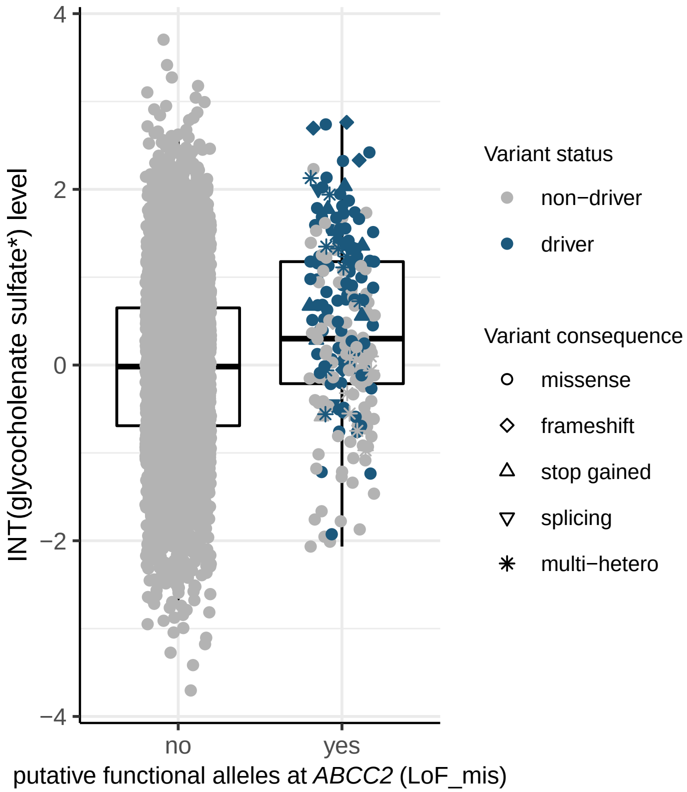
INT(glycocholenate sulfate*) level
4
2
0
−2
−4
no
yes
putative functional alleles at *ABCC2* (LoF_mis)
Variant status
non−driver
driver
Variant consequence
missense
frameshift
stop gained
splicing
multi−hetero

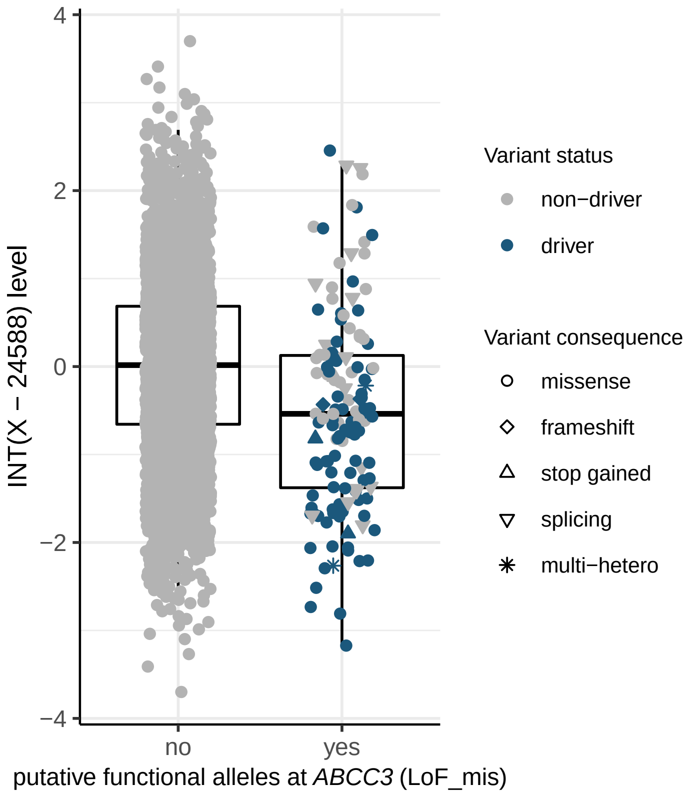
INT(X − 24588) level
4
2
0
−2
−4
no
yes
putative functional alleles at *ABCC3* (LoF_mis)
Variant status
non−driver
driver
Variant consequence
missense
frameshift
stop gained
splicing
multi−hetero

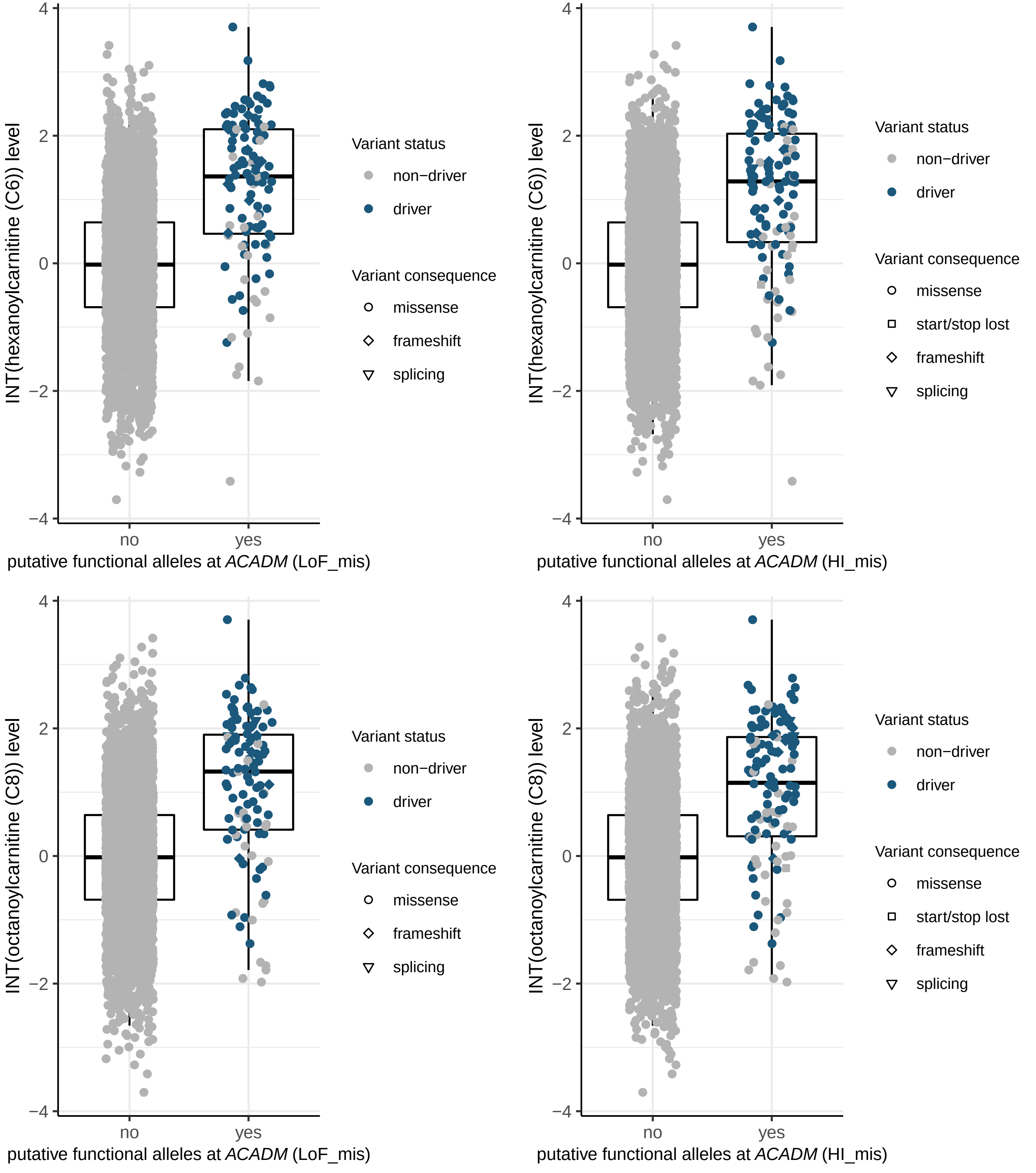
INT(hexanoylcarnitine (C6)) level
putative functional alleles at ACADM (LoF_mis)
putative functional alleles at ACADM (HI_mis)
INT(octanoylcarnitine (C8)) level
no
yes
Variant status
non−driver
driver
Variant consequence
missense
start/stop lost
frameshift
splicing

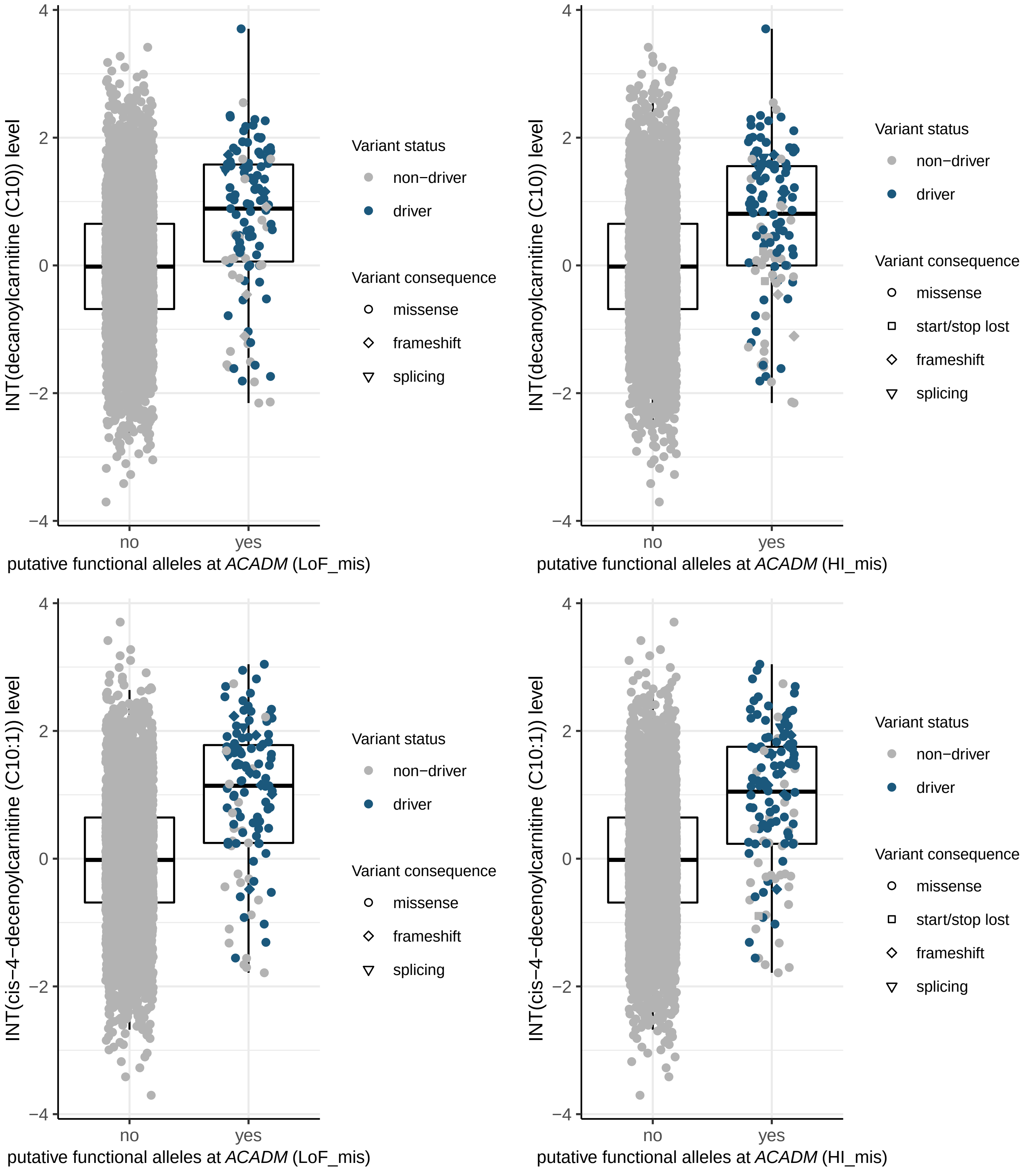
INT(decanoylcarnitine (C10)) level
4
2
0
−2
−4
no
yes
putative functional alleles at *ACADM* (LoF_mis)
Variant status
non−driver
driver
Variant consequence
missense
frameshift
splicing
INT(decanoylcarnitine (C10)) level
putative functional alleles at *ACADM* (HI_mis)
Variant status
non−driver
driver
Variant consequence
missense
start/stop lost
frameshift
splicing
INT(cis−4−decenoylcarnitine (C10:1)) level
putative functional alleles at *ACADM* (LoF_mis)
Variant status
non−driver
driver
Variant consequence
missense
frameshift
splicing
INT(cis−4−decenoylcarnitine (C10:1)) level
putative functional alleles at *ACADM* (HI_mis)
Variant status
non−driver
driver
Variant consequence
missense
start/stop lost
frameshift
splicing

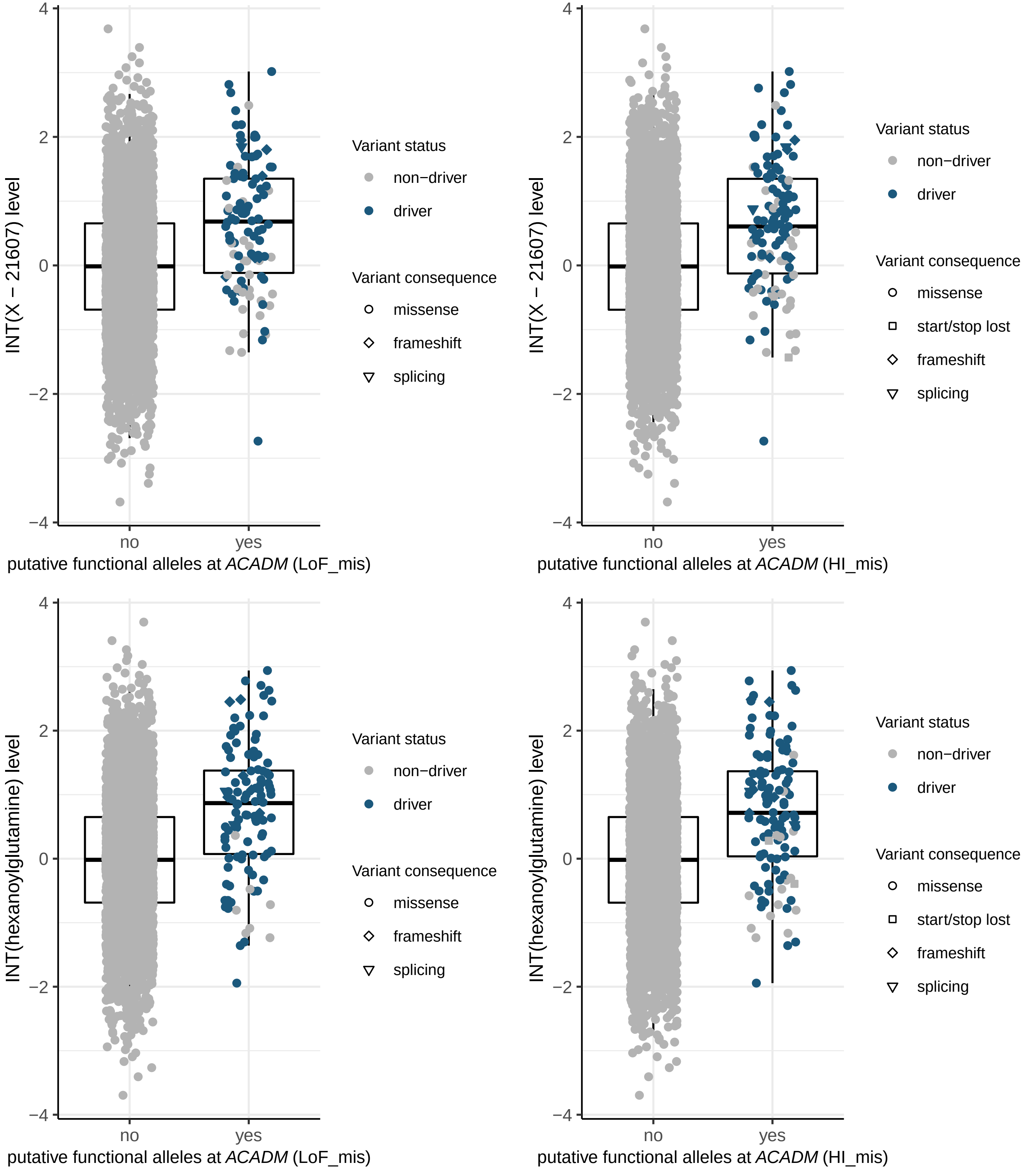

INT(X − 21607) level
putative functional alleles at ACADM (LoF_mis)
putative functional alleles at ACADM (HI_mis)
INT(hexanoylglutamine) level
no
yes
4
2
0
−2
−4
Variant status
non−driver
driver
Variant consequence
missense
start/stop lost
frameshift
splicing

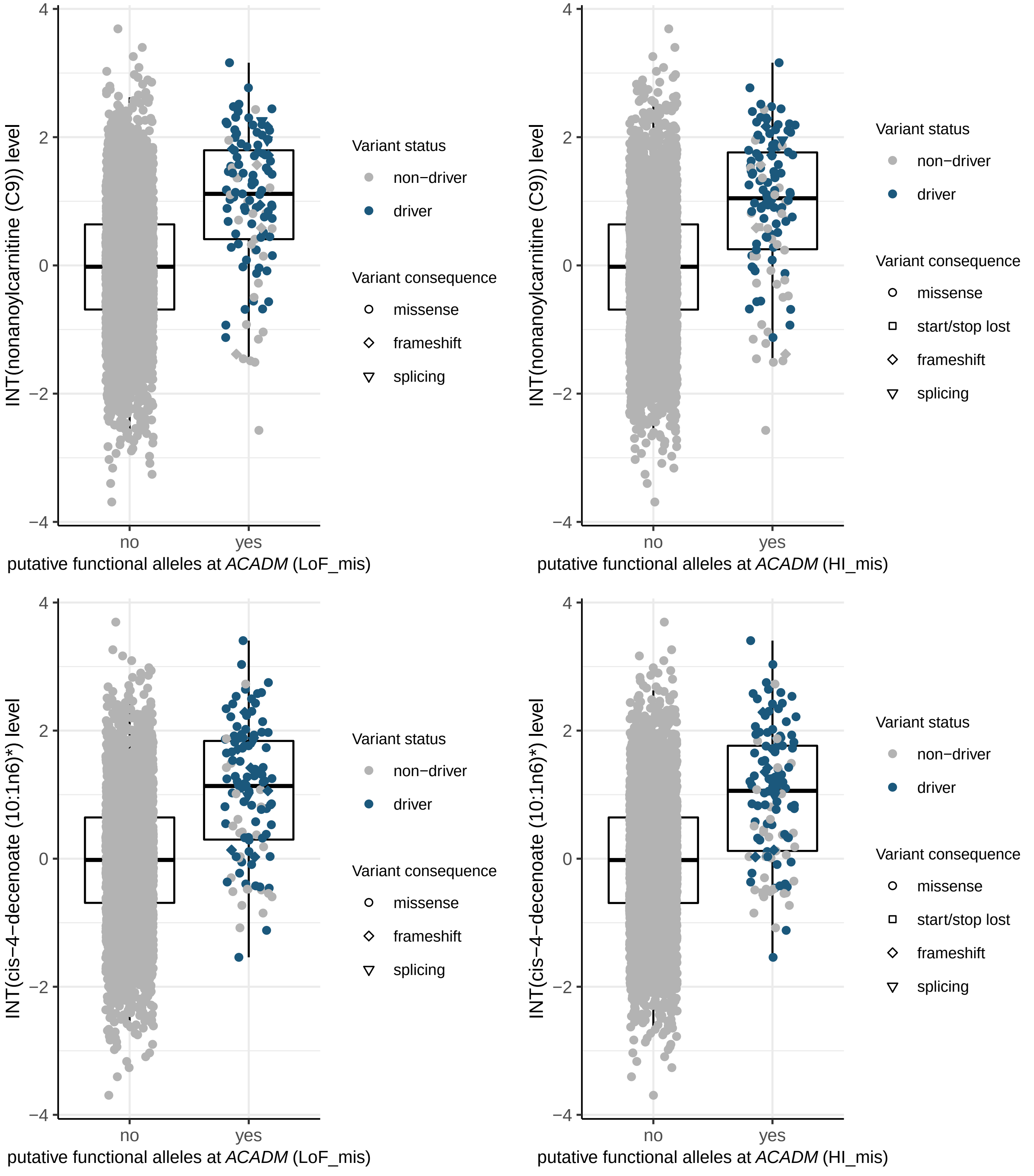

INT(nonanoylcarnitine (C9)) level
putative functional alleles at ACADM (LoF_mis)
putative functional alleles at ACADM (HI_mis)
INT(cis−4−decenoate (10:1n6)*) level
no
yes
Variant status
non−driver
driver
Variant consequence
missense
start/stop lost
frameshift
splicing

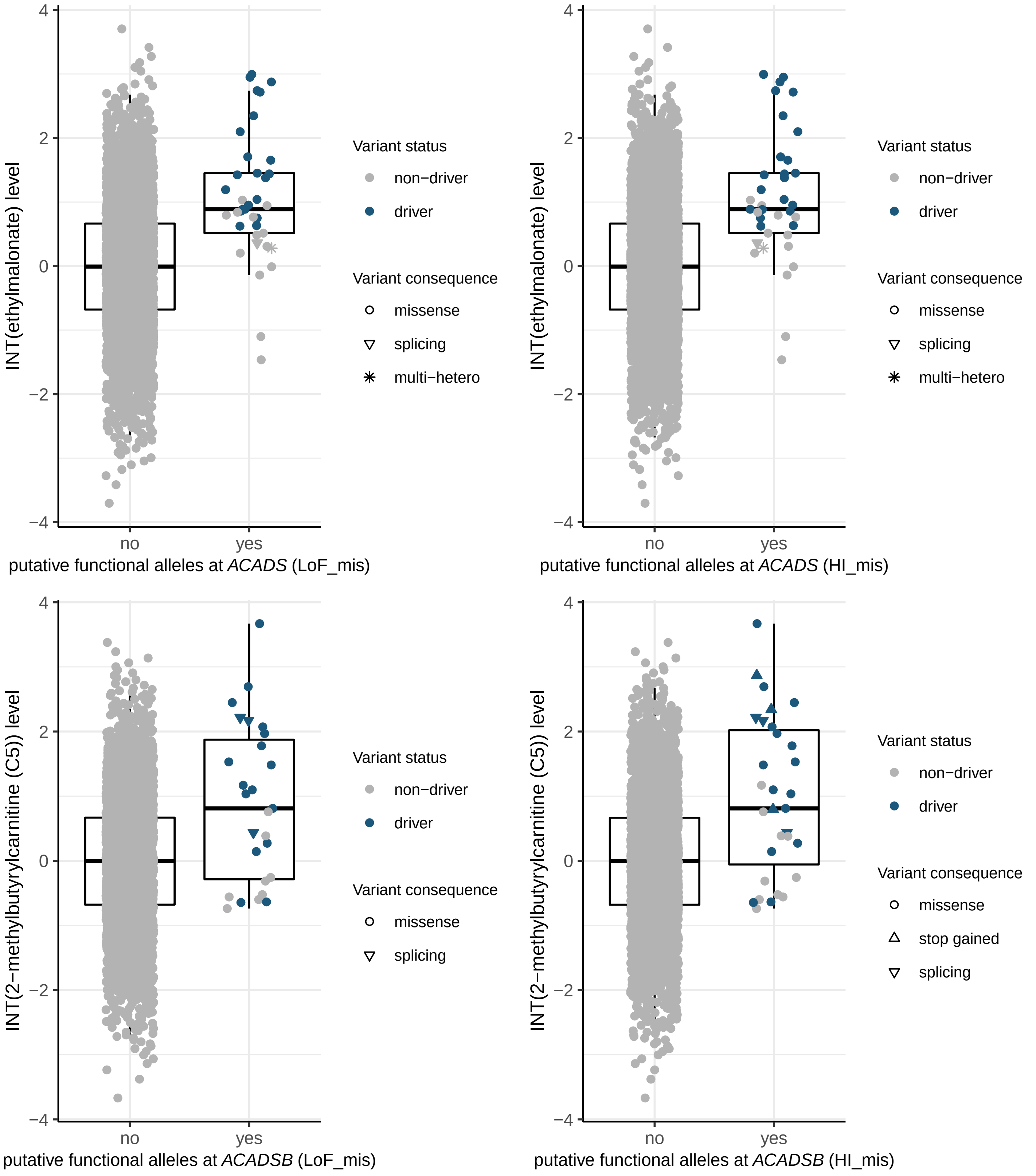
INT(ethylmalonate) level
putative functional alleles at ACADS (LoF_mis)
putative functional alleles at ACADS (HI_mis)
INT(2−methylbutyrylcarnitine (C5)) level
putative functional alleles at ACADSB (LoF_mis)
putative functional alleles at ACADSB (HI_mis)
no
yes
Variant status
non−driver
driver
Variant consequence
missense
splicing
multi−hetero
stop gained
4
2
0
−2
−4

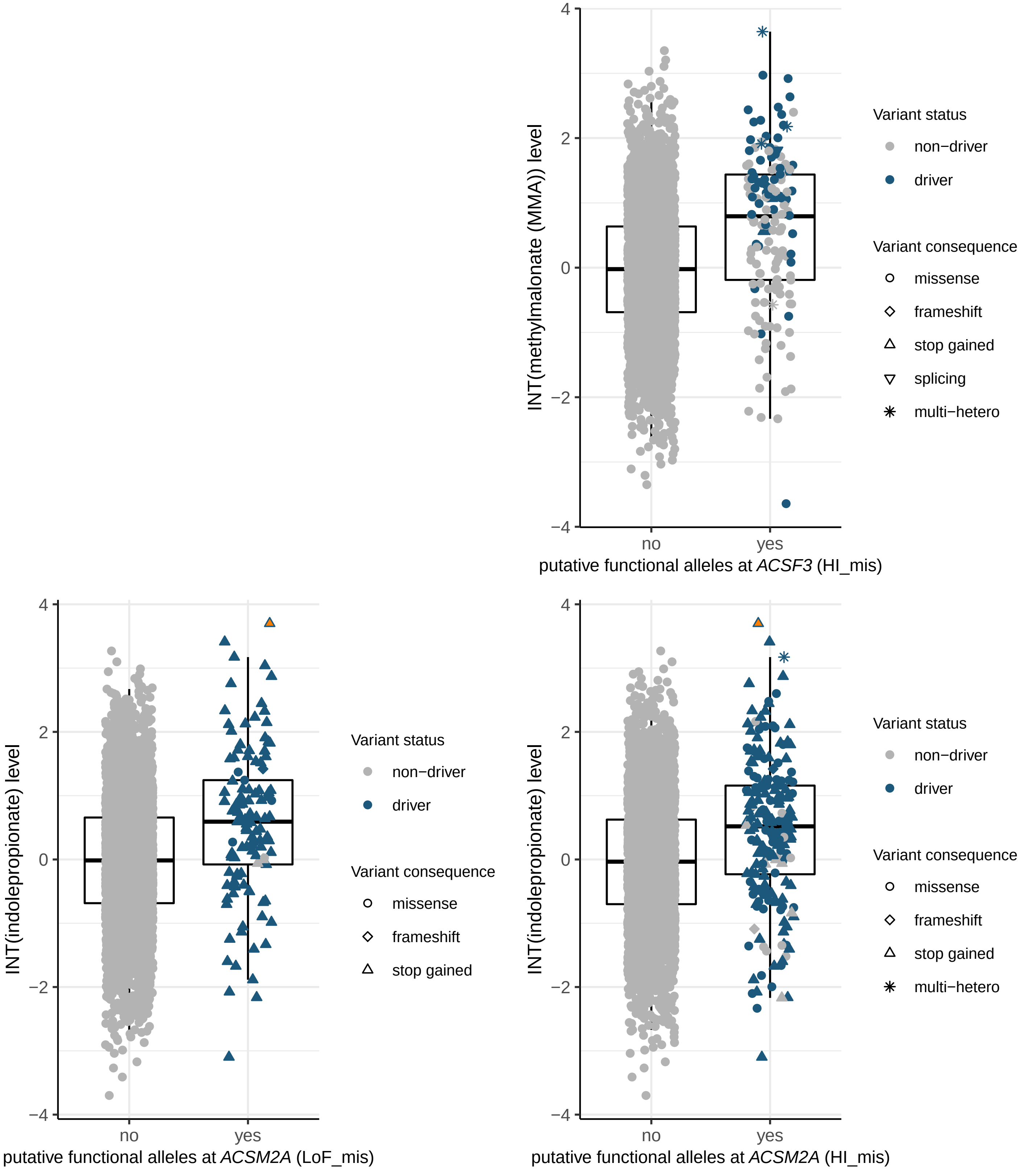
INT(methylmalonate (MMA)) level
putative functional alleles at ACSF3 (HI_mis)
no
yes
4
2
0
−2
−4
Variant status
non−driver
driver
Variant consequence
missense
frameshift
stop gained
splicing
multi−hetero
INT(indolepropionate) level
putative functional alleles at ACSM2A (LoF_mis)
Variant status
non−driver
driver
Variant consequence
missense
frameshift
stop gained
INT(indolepropionate) level
putative functional alleles at ACSM2A (HI_mis)
Variant status
non−driver
driver
Variant consequence
missense
frameshift
stop gained
multi−hetero

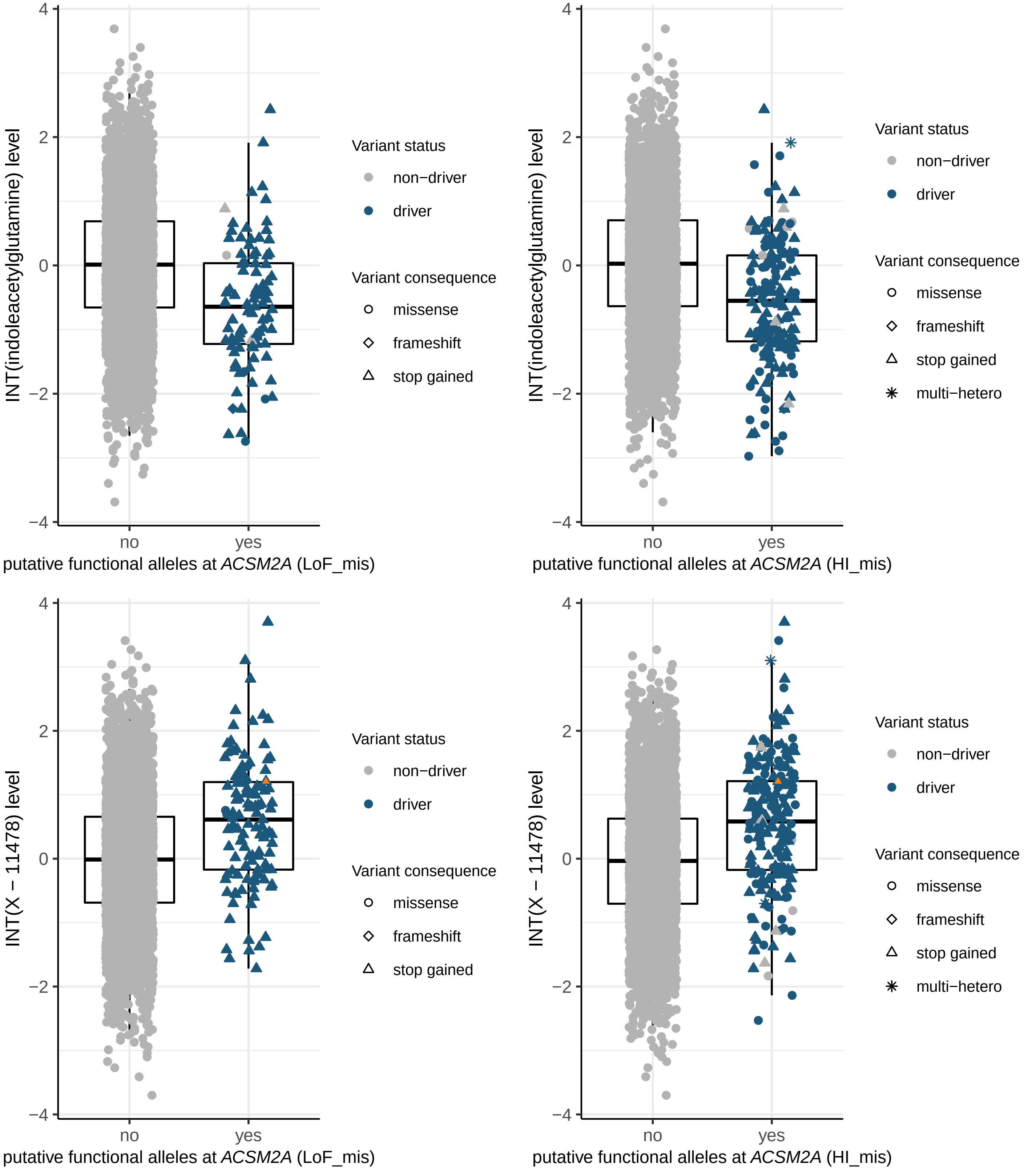
INT(indoleacetylglutamine) level
putative functional alleles at ACSM2A (LoF_mis)
putative functional alleles at ACSM2A (HI_mis)
INT(X − 11478) level
no
yes
Variant status
non−driver
driver
Variant consequence
missense
frameshift
stop gained
multi−hetero
4
2
0
−2
−4

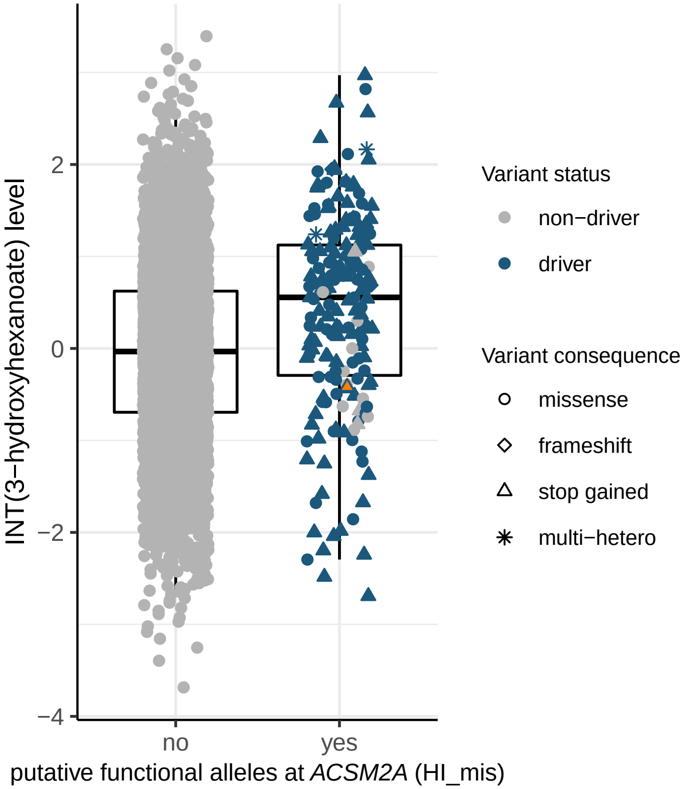
INT(3−hydroxyhexanoate) level
2
0
−2
−4
no
yes
putative functional alleles at *ACSM2A* (HI_mis)
Variant status
non−driver
driver
Variant consequence
missense
frameshift
stop gained
multi−hetero

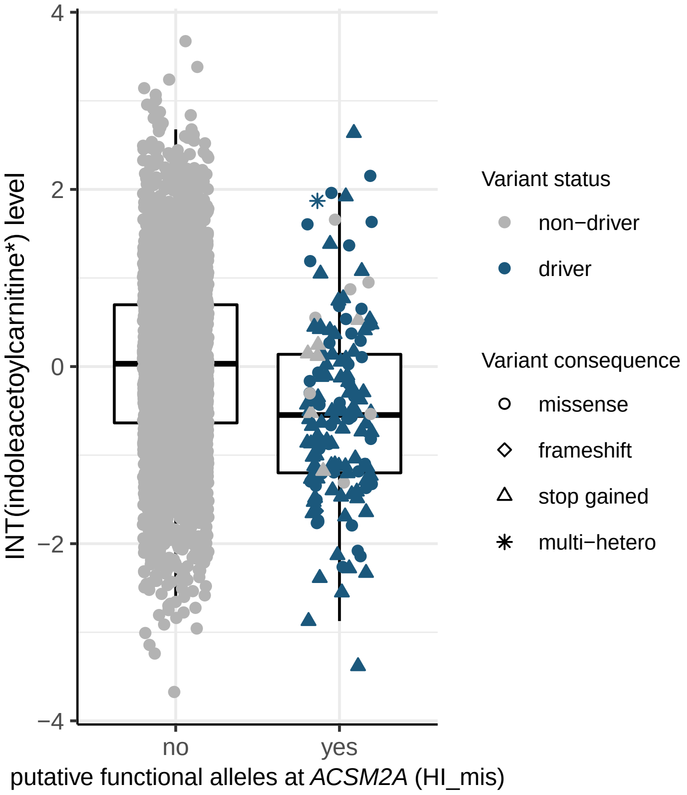
INT(indoleacetoylcarnitine*) level
4
2
0
−2
−4
no
yes
putative functional alleles at *ACSM2A* (HI_mis)
Variant status
non−driver
driver
Variant consequence
missense
frameshift
stop gained
multi−hetero

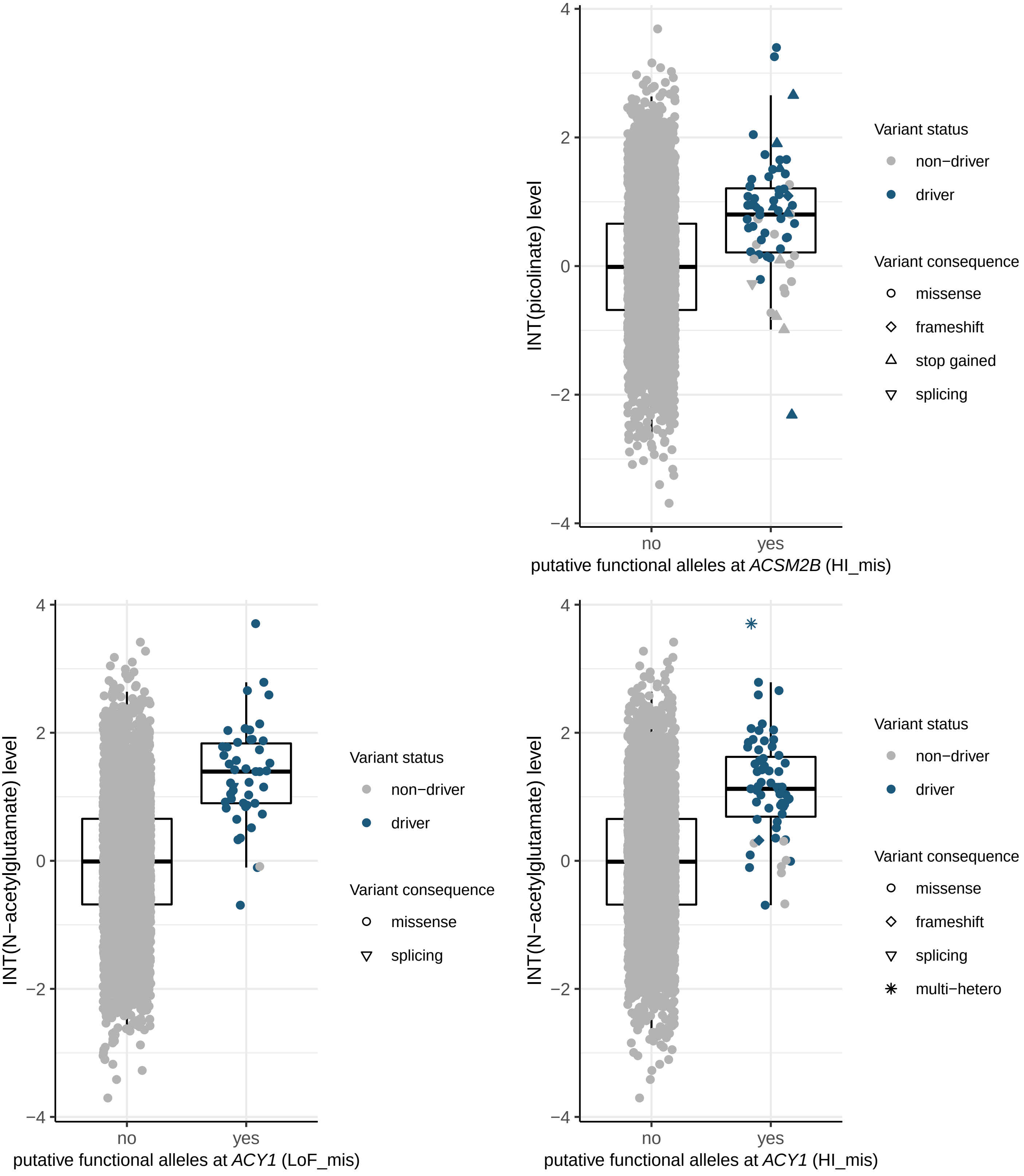
INT(picolinate) level
4
2
0
−2
−4
no
yes
putative functional alleles at ACSM2B (HI_mis)
Variant status
non−driver
driver
Variant consequence
missense
frameshift
stop gained
splicing
INT(N−acetylglutamate) level
4
2
0
−2
−4
no
yes
putative functional alleles at ACY1 (LoF_mis)
Variant status
non−driver
driver
Variant consequence
missense
splicing
INT(N−acetylglutamate) level
4
2
0
−2
−4
no
yes
putative functional alleles at ACY1 (HI_mis)
Variant status
non−driver
driver
Variant consequence
missense
frameshift
splicing
multi−hetero

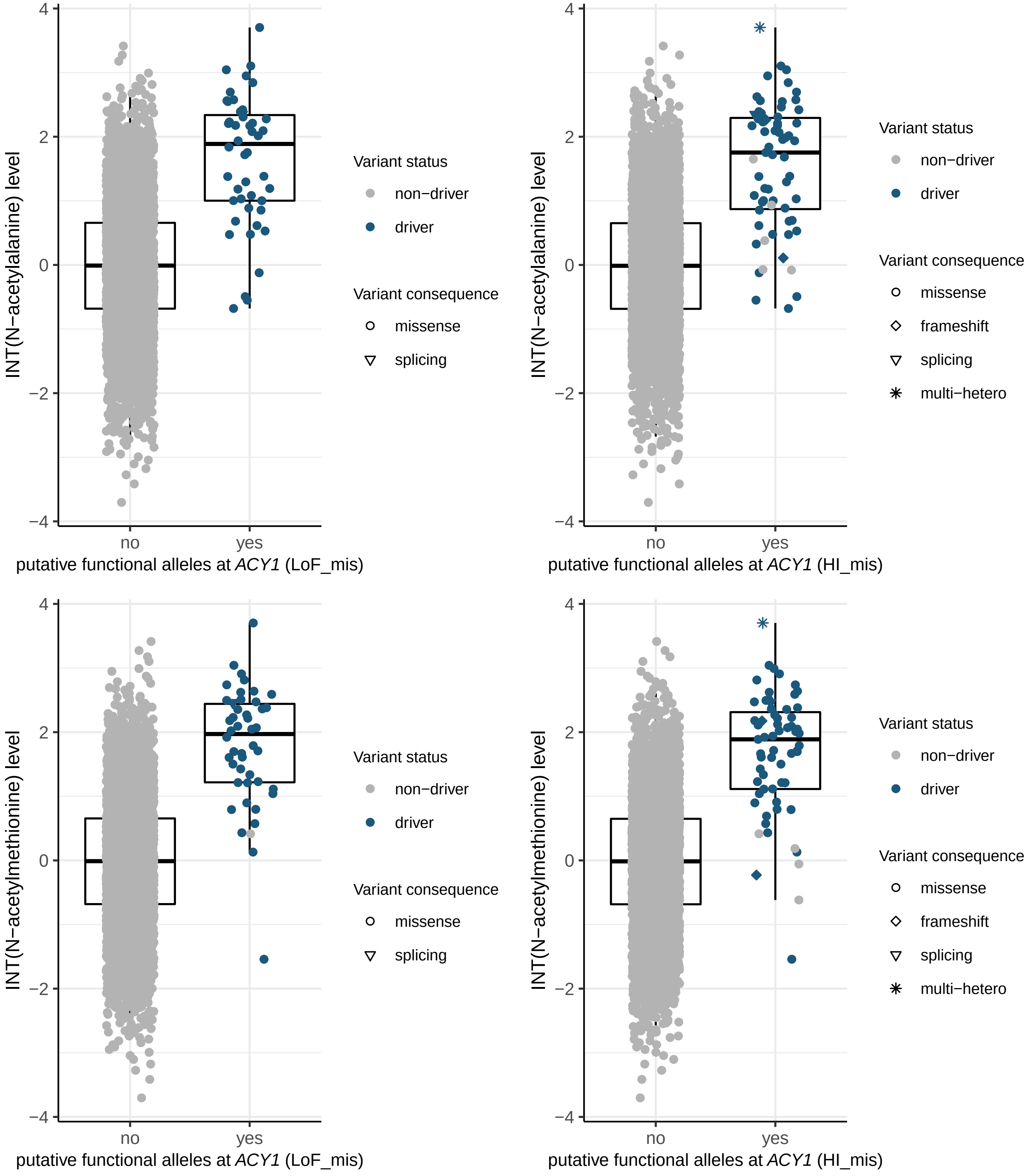
INT(N−acetylalanine) level
4
2
0
−2
−4
no
yes
putative functional alleles at *ACY1* (LoF_mis)
Variant status
non−driver
driver
Variant consequence
missense
splicing
INT(N−acetylalanine) level
4
2
0
−2
−4
no
yes
putative functional alleles at *ACY1* (HI_mis)
Variant status
non−driver
driver
Variant consequence
missense
frameshift
splicing
multi−hetero
INT(N−acetylmethionine) level
4
2
0
−2
−4
no
yes
putative functional alleles at *ACY1* (LoF_mis)
Variant status
non−driver
driver
Variant consequence
missense
splicing
INT(N−acetylmethionine) level
4
2
0
−2
−4
no
yes
putative functional alleles at *ACY1* (HI_mis)
Variant status
non−driver
driver
Variant consequence
missense
frameshift
splicing
multi−hetero

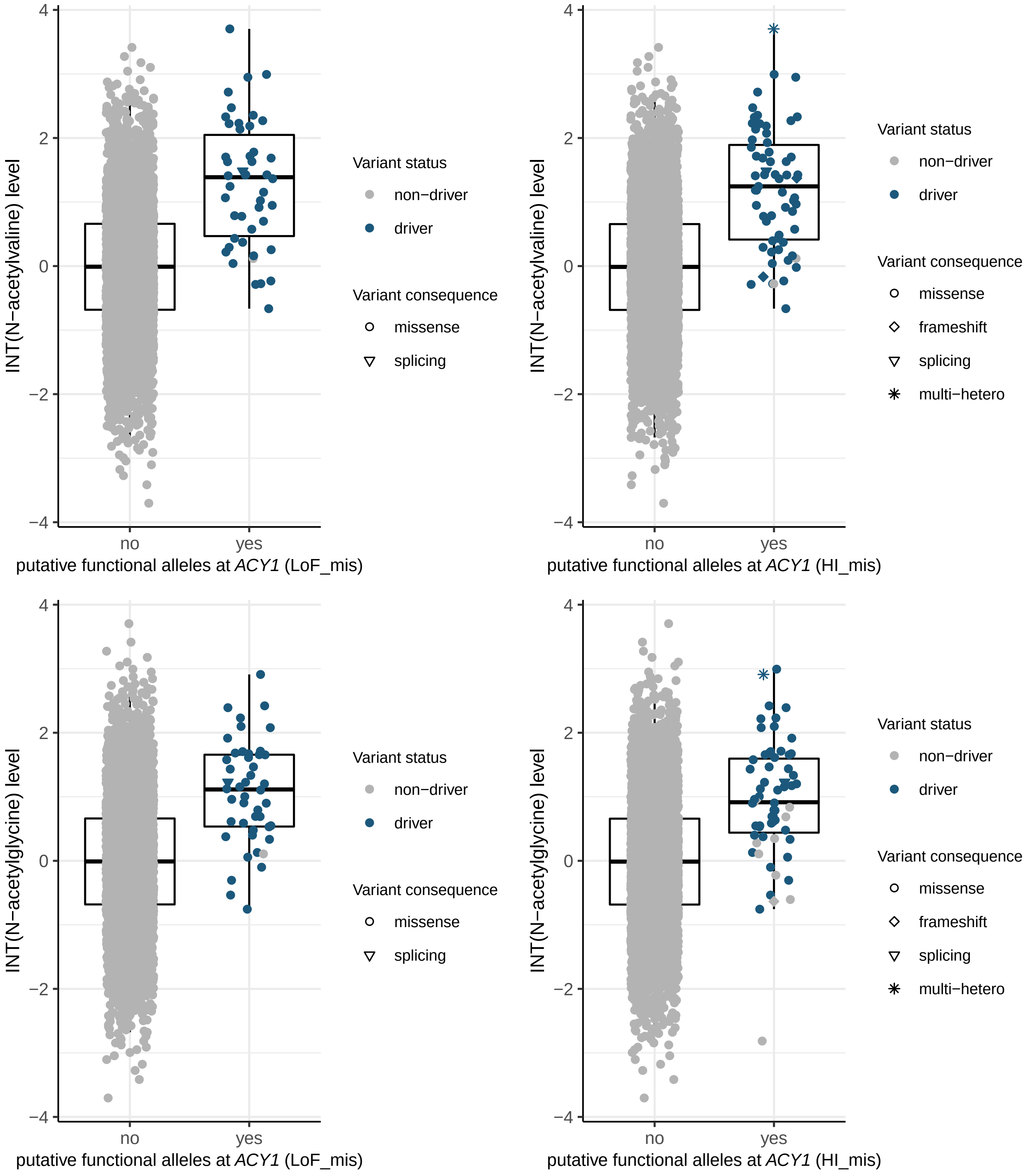

INT(N−acetylvaline) level
putative functional alleles at *ACY1* (LoF_mis)
putative functional alleles at *ACY1* (HI_mis)
INT(N−acetylglycine) level
no
yes
4
2
0
−2
−4
Variant status
non−driver
driver
Variant consequence
missense
frameshift
splicing
multi−hetero

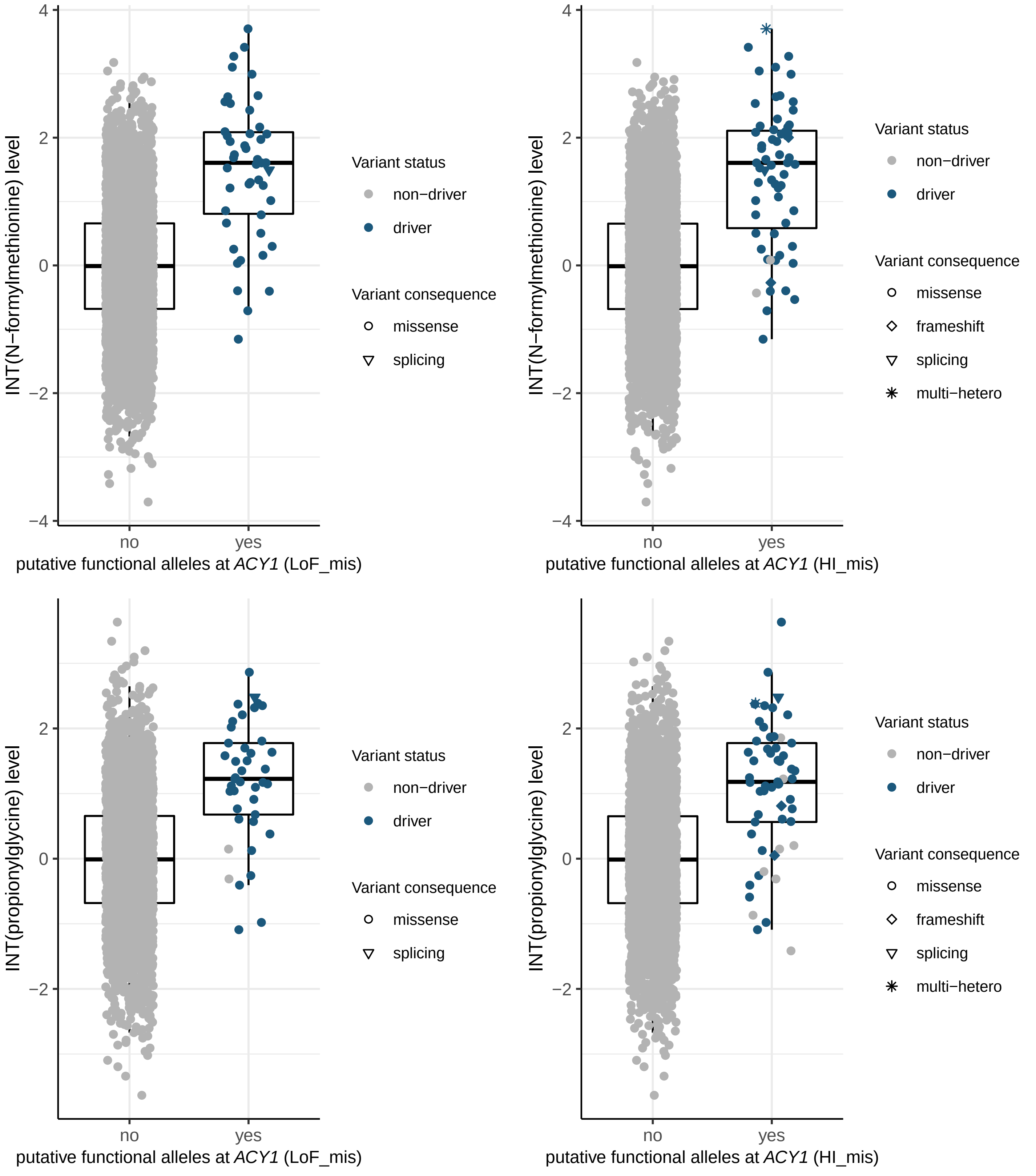

INT(N−formylmethionine) level
putative functional alleles at ACY1 (LoF_mis)
putative functional alleles at ACY1 (HI_mis)
INT(propionylglycine) level
no
yes
4
2
0
−2
−4
Variant status
non−driver
driver
Variant consequence
missense
frameshift
splicing
multi−hetero

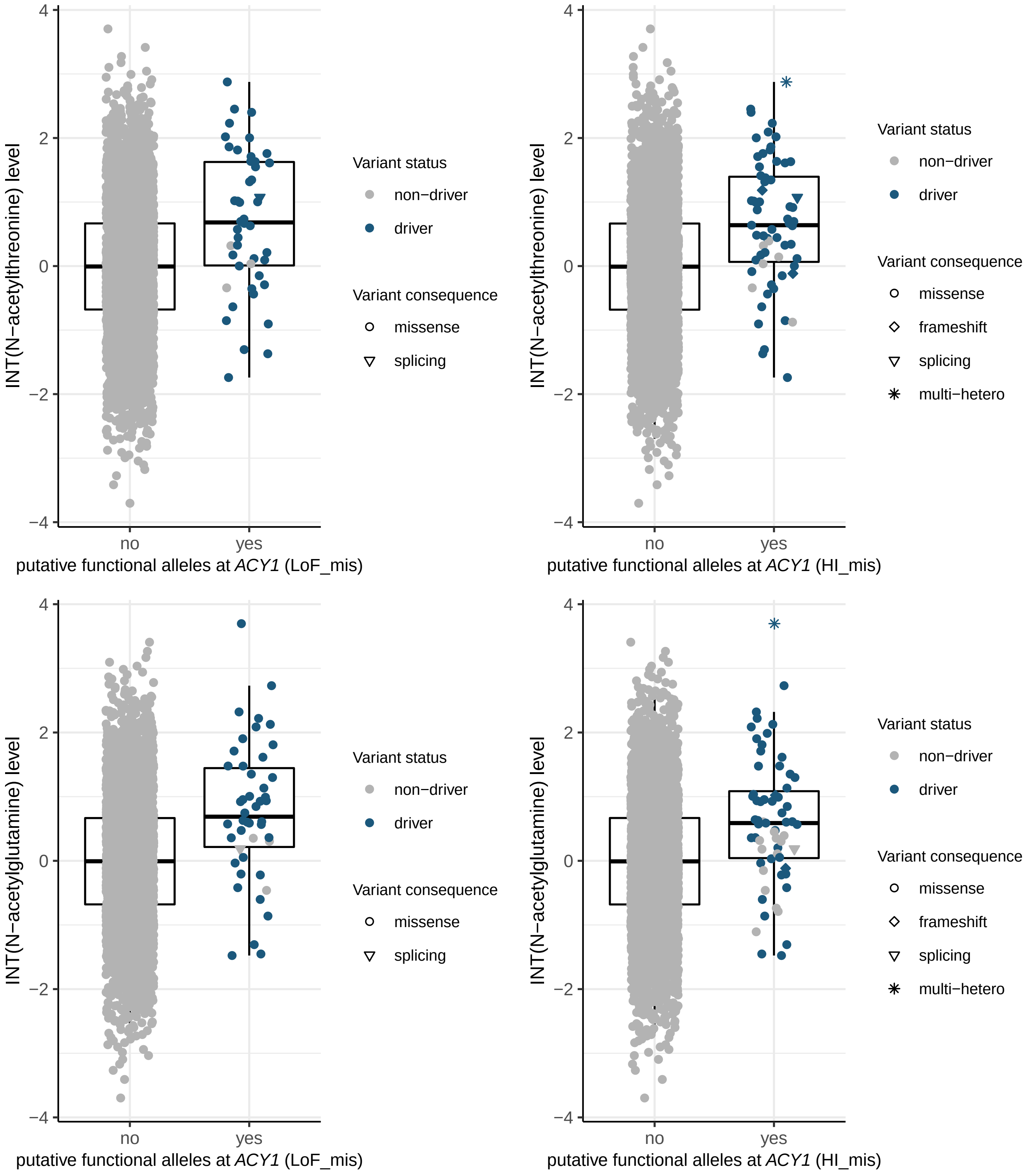

INT(N−acetylthreonine) level
putative functional alleles at ACY1 (LoF_mis)
putative functional alleles at ACY1 (HI_mis)
INT(N−acetylglutamine) level
no
yes
Variant status
non−driver
driver
Variant consequence
missense
frameshift
splicing
multi−hetero

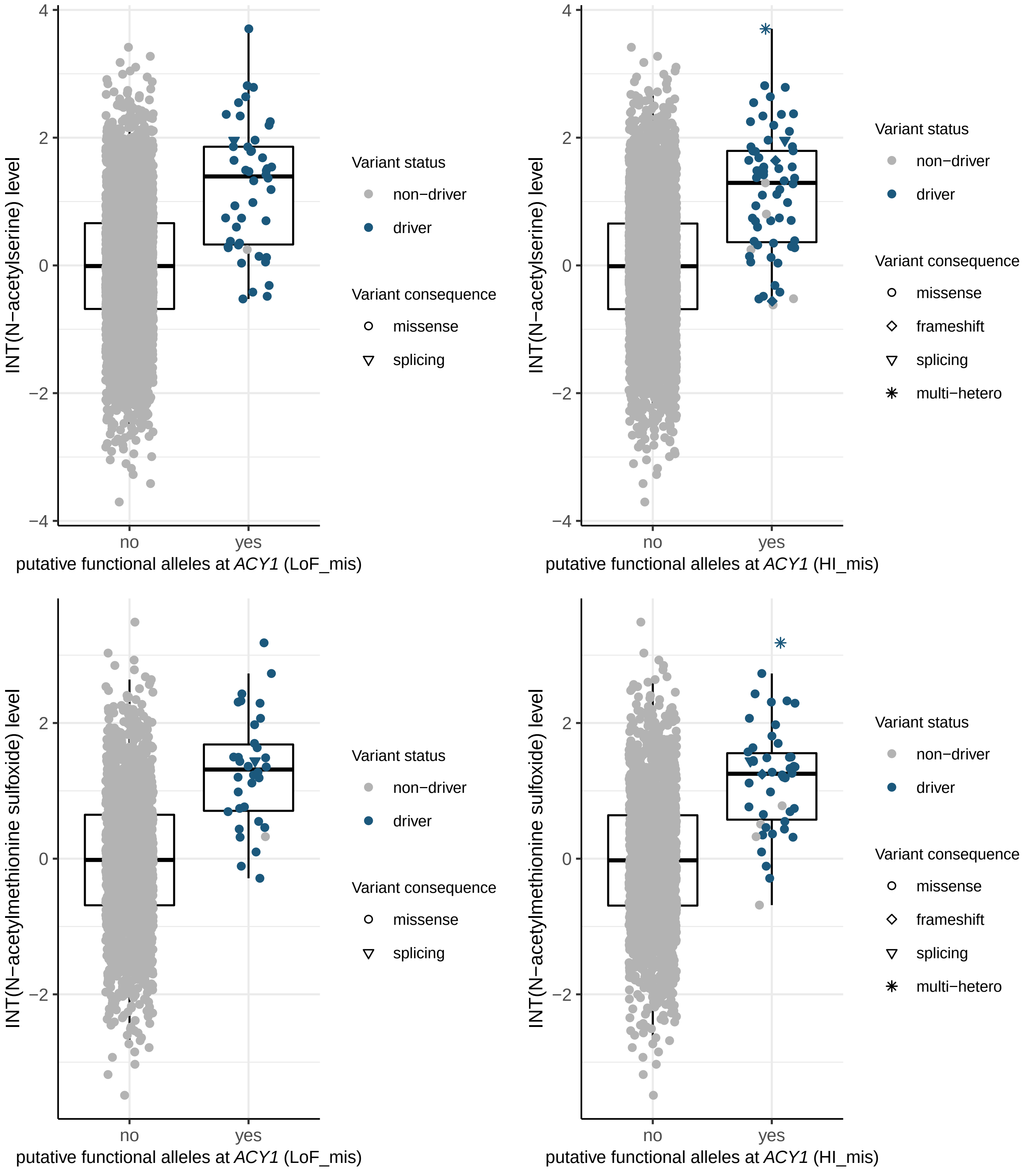

INT(N−acetylserine) level
putative functional alleles at ACY1 (LoF_mis)
putative functional alleles at ACY1 (HI_mis)
INT(N−acetylmethionine sulfoxide) level
no
yes
Variant status
non−driver
driver
Variant consequence
missense
frameshift
splicing
multi−hetero

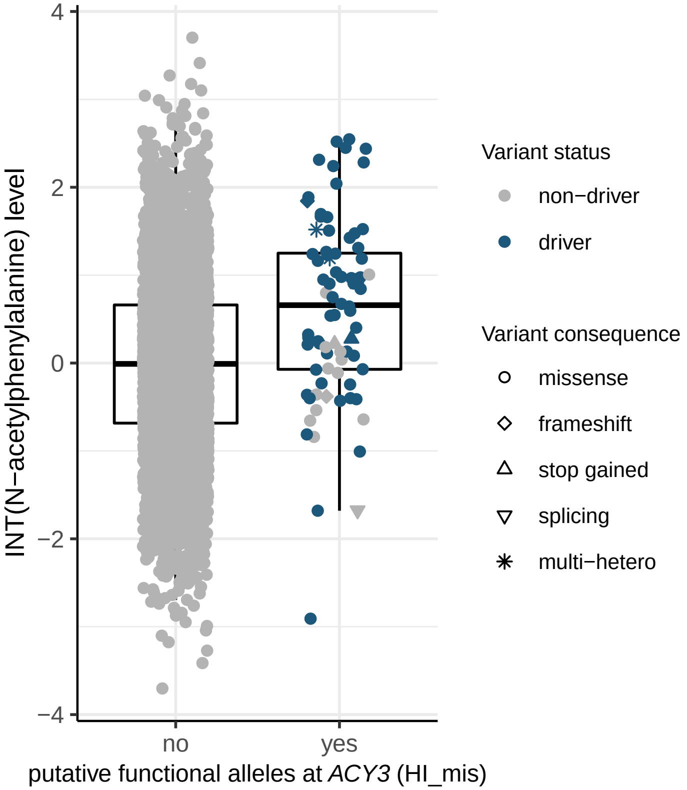
4
2
0
−2
−4
INT(N−acetylphenylalanine) level
no
yes
putative functional alleles at *ACY3* (HI_mis)
Variant status
non−driver
driver
Variant consequence
missense
frameshift
stop gained
splicing
multi−hetero

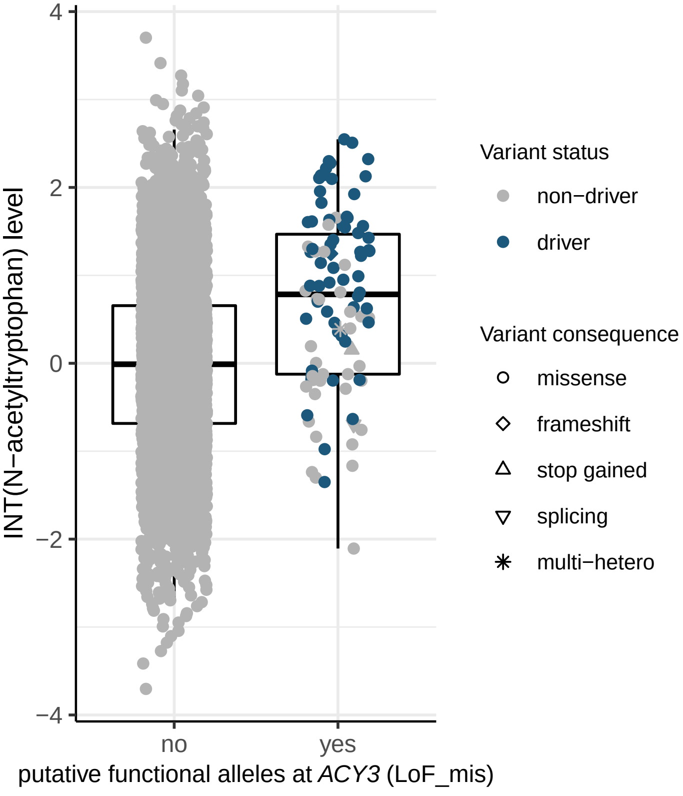
4
2
0
−2
−4
INT(N−acetyltryptophan) level
no
yes
putative functional alleles at *ACY3* (LoF_mis)
Variant status
non−driver
driver
Variant consequence
missense
frameshift
stop gained
splicing
multi−hetero

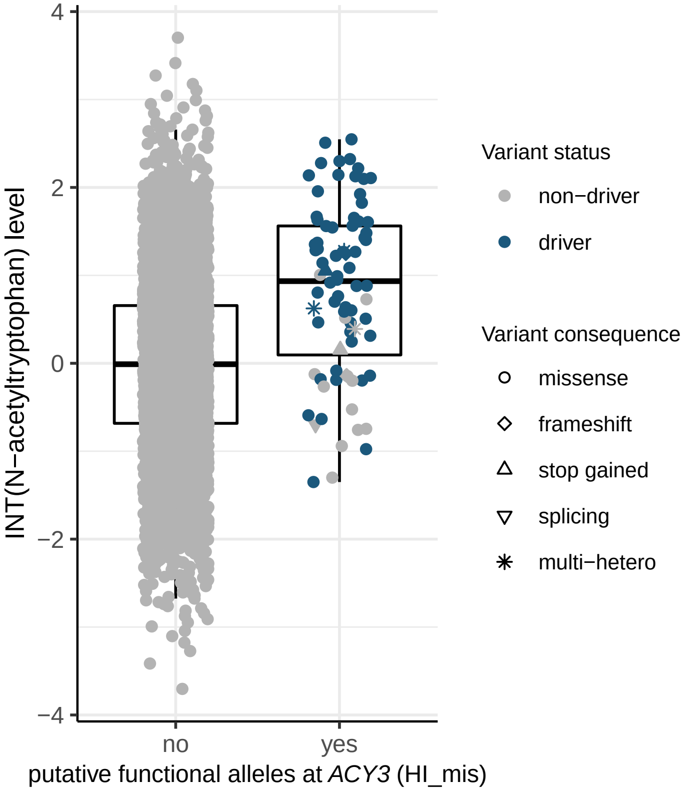
4
2
0
−2
−4
INT(N−acetyltryptophan) level
no
yes
putative functional alleles at *ACY3* (HI_mis)
Variant status
non−driver
driver
Variant consequence
missense
frameshift
stop gained
splicing
multi−hetero

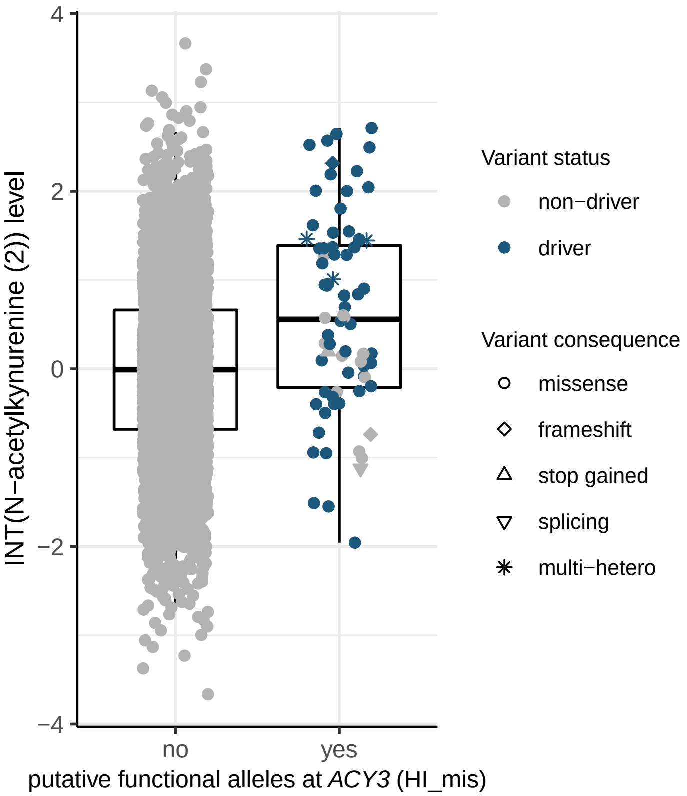

INT(N−acetylkynurenine (2)) level
4
2
0
−2
−4
no
yes
putative functional alleles at *ACY3* (HI_mis)
Variant status
non−driver
driver
Variant consequence
missense
frameshift
stop gained
splicing
multi−hetero


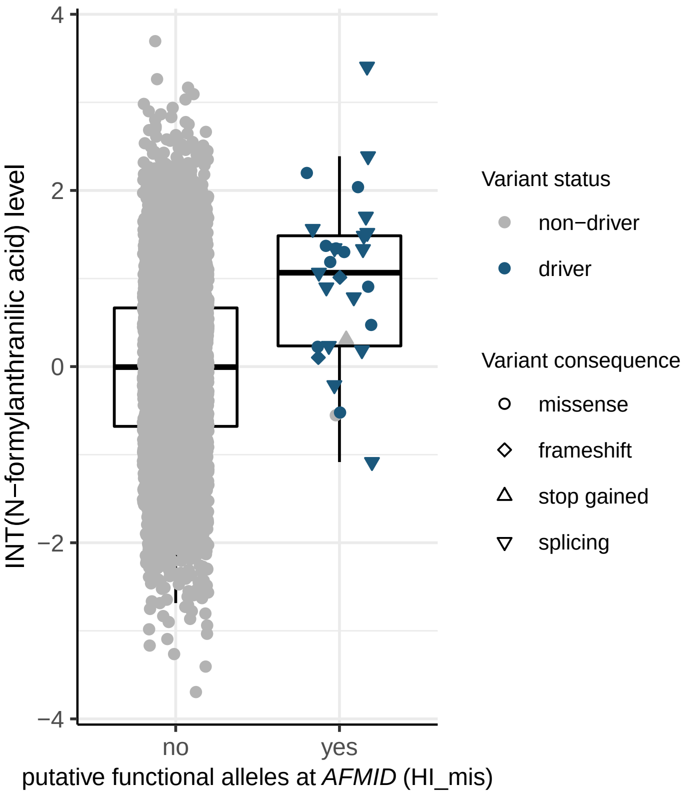

INT(N−formylanthranilic acid) level
4
2
0
−2
−4
no
yes
putative functional alleles at *AFMID* (HI_mis)
Variant status
non−driver
driver
Variant consequence
missense
frameshift
stop gained
splicing

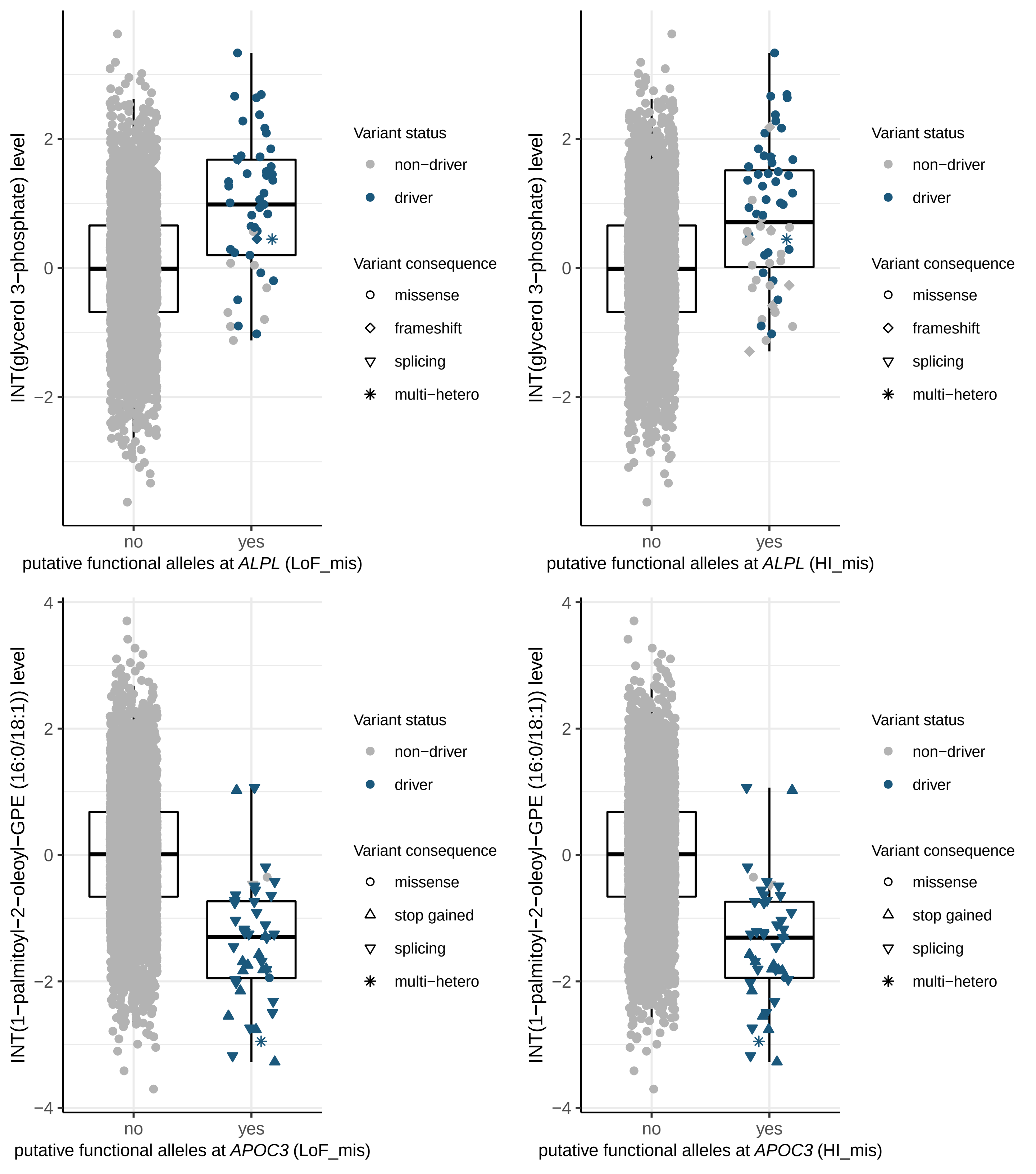
INT(glycerol 3−phosphate) level
putative functional alleles at ALPL (LoF_mis)
putative functional alleles at ALPL (HI_mis)
INT(1−palmitoyl−2−oleoyl−GPE (16:0/18:1)) level
putative functional alleles at APOC3 (LoF_mis)
putative functional alleles at APOC3 (HI_mis)
no
yes
Variant status
non−driver
driver
Variant consequence
missense
frameshift
splicing
stop gained
multi−hetero
4
2
0
−2
−4

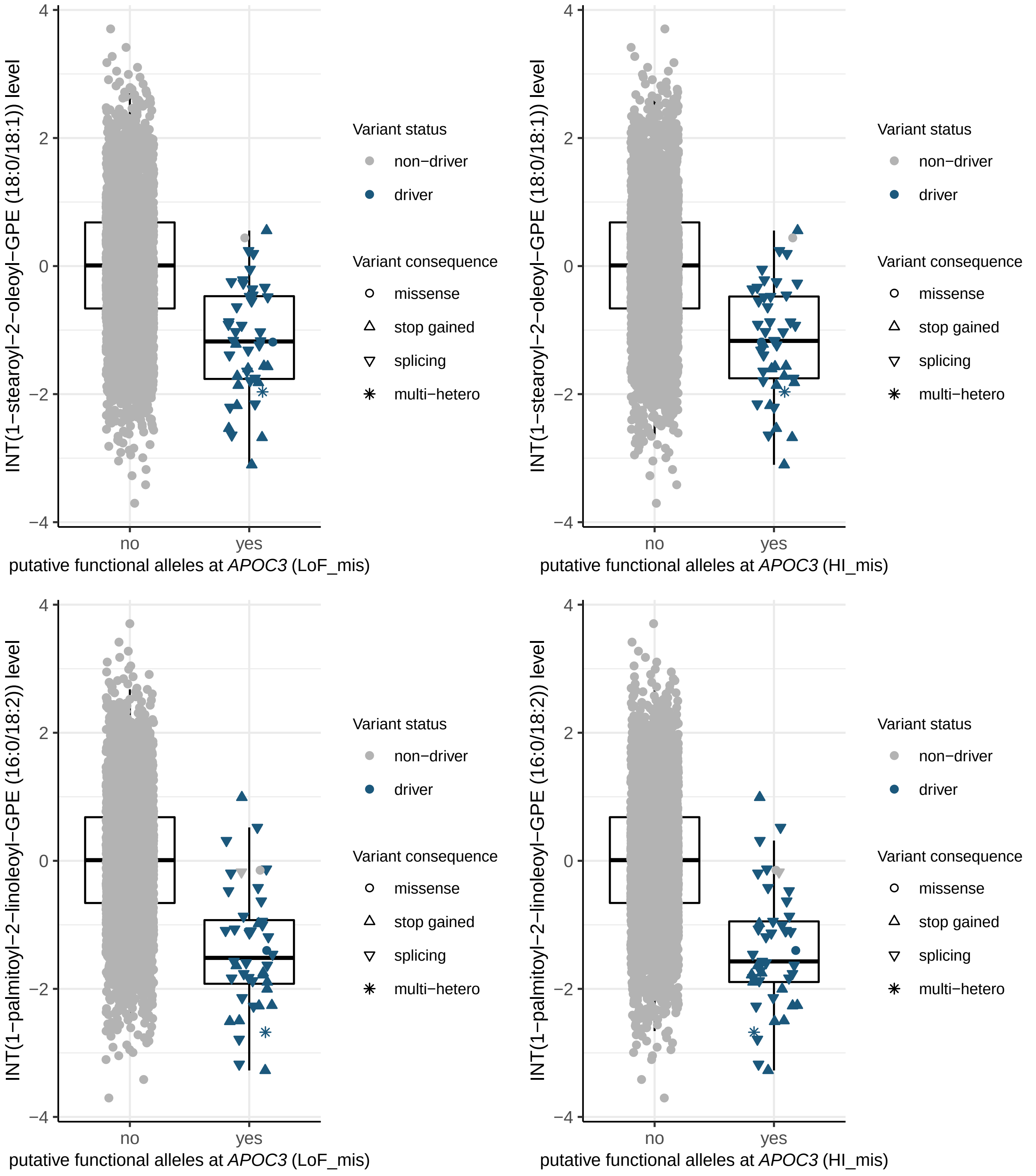

INT(1−stearoyl−2−oleoyl−GPE (18:0/18:1)) level
4
2
0
−2
−4
no
yes
putative functional alleles at *APOC3* (LoF_mis)
putative functional alleles at *APOC3* (HI_mis)
INT(1−palmitoyl−2−linoleoyl−GPE (16:0/18:2)) level
Variant status
non−driver
driver
Variant consequence
missense
stop gained
splicing
multi−hetero

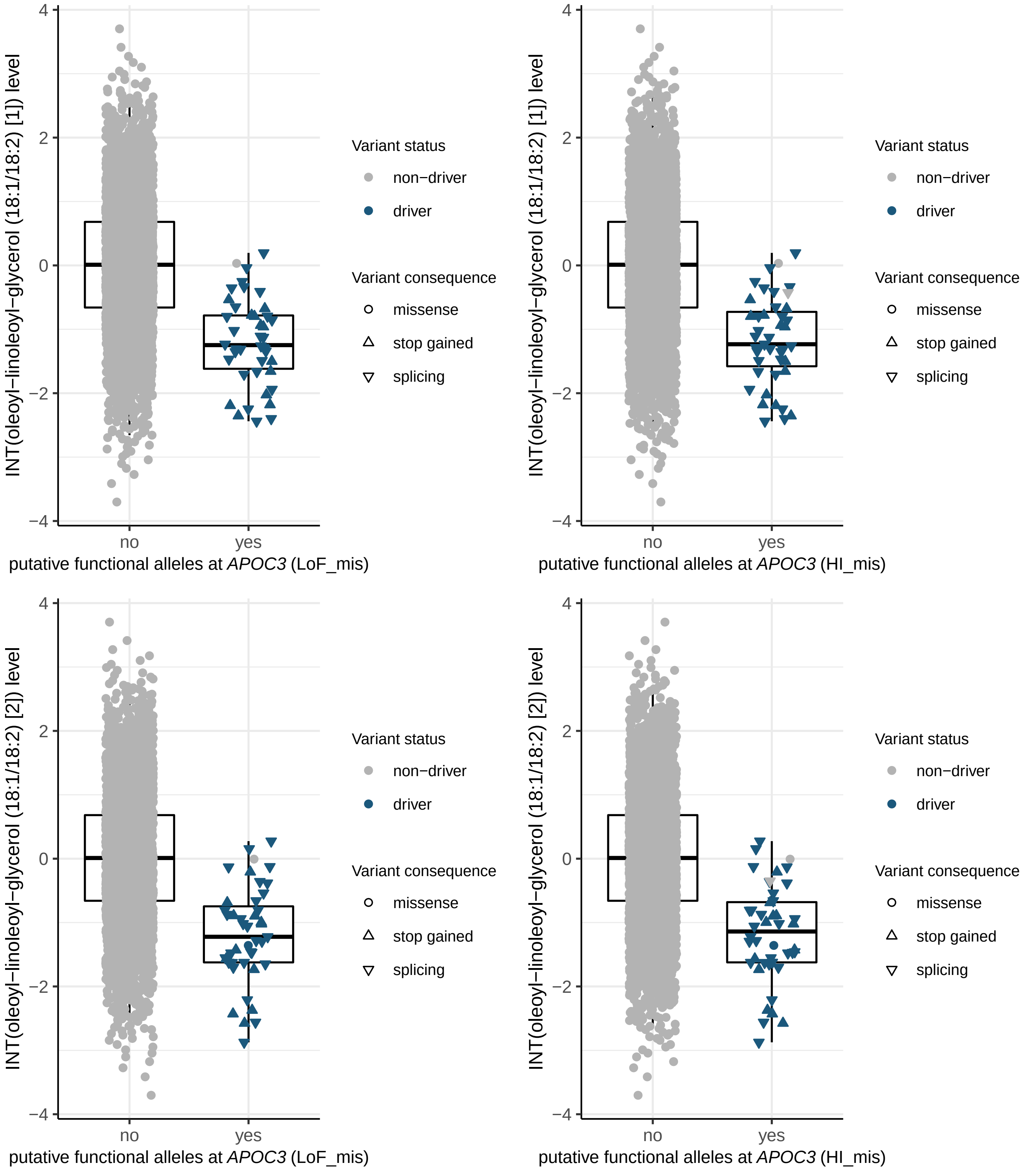

INT(oleoyl−linoleoyl−glycerol (18:1/18:2) [1]) level
4
2
0
−2
−4
no
yes
putative functional alleles at *APOC3* (LoF_mis)
Variant status
non−driver
driver
Variant consequence
missense
stop gained
splicing
INT(oleoyl−linoleoyl−glycerol (18:1/18:2) [1]) level
putative functional alleles at *APOC3* (HI_mis)
INT(oleoyl−linoleoyl−glycerol (18:1/18:2) [2]) level
putative functional alleles at *APOC3* (LoF_mis)
INT(oleoyl−linoleoyl−glycerol (18:1/18:2) [2]) level
putative functional alleles at *APOC3* (HI_mis)

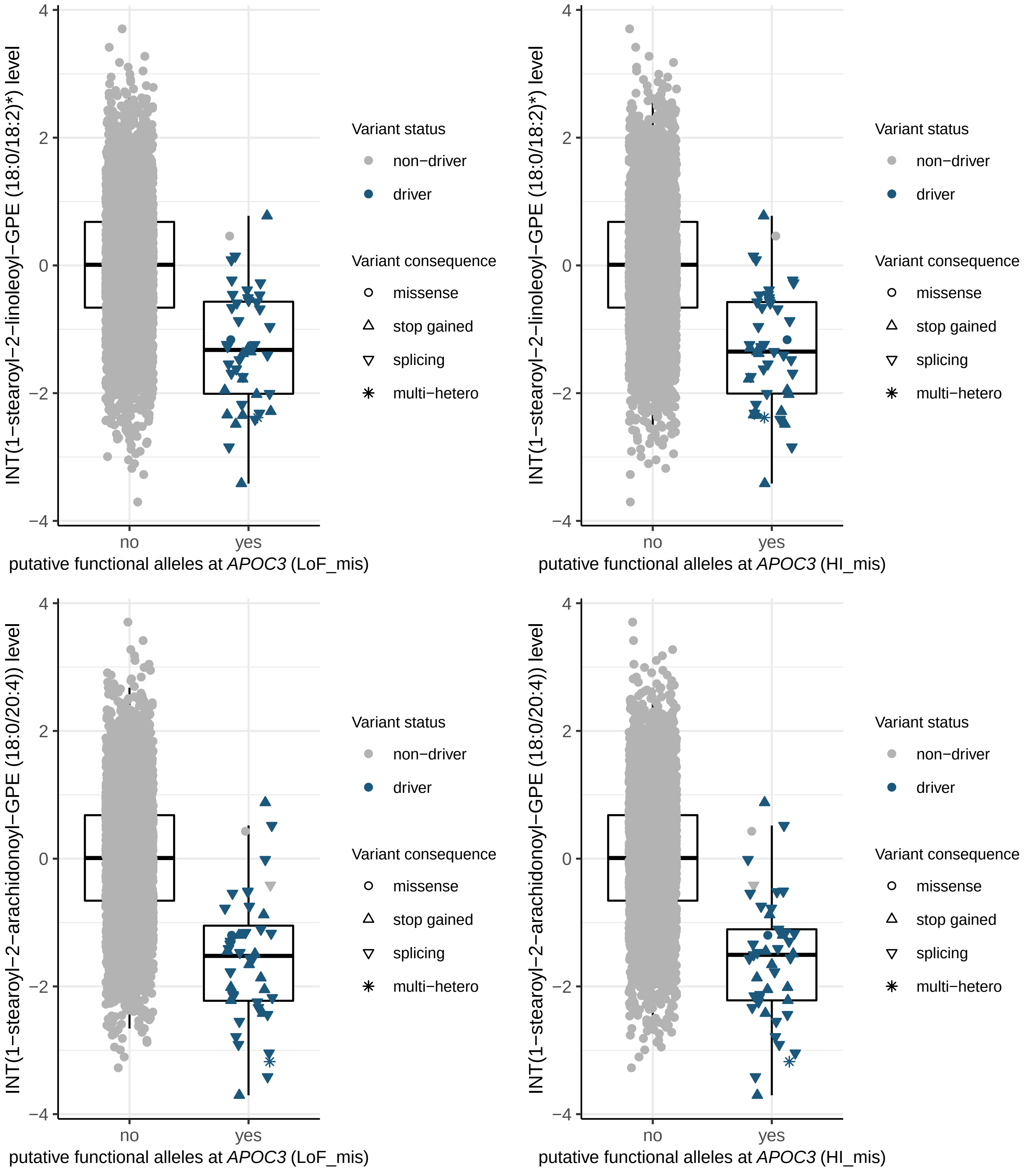

INT(1−stearoyl−2−linoleoyl−GPE (18:0/18:2)*) level
4
2
0
−2
−4
no
yes
putative functional alleles at APOC3 (LoF_mis)
Variant status
non−driver
driver
Variant consequence
missense
stop gained
splicing
multi−hetero
INT(1−stearoyl−2−linoleoyl−GPE (18:0/18:2)*) level
putative functional alleles at APOC3 (HI_mis)
INT(1−stearoyl−2−arachidonoyl−GPE (18:0/20:4)) level
putative functional alleles at APOC3 (LoF_mis)
INT(1−stearoyl−2−arachidonoyl−GPE (18:0/20:4)) level
putative functional alleles at APOC3 (HI_mis)

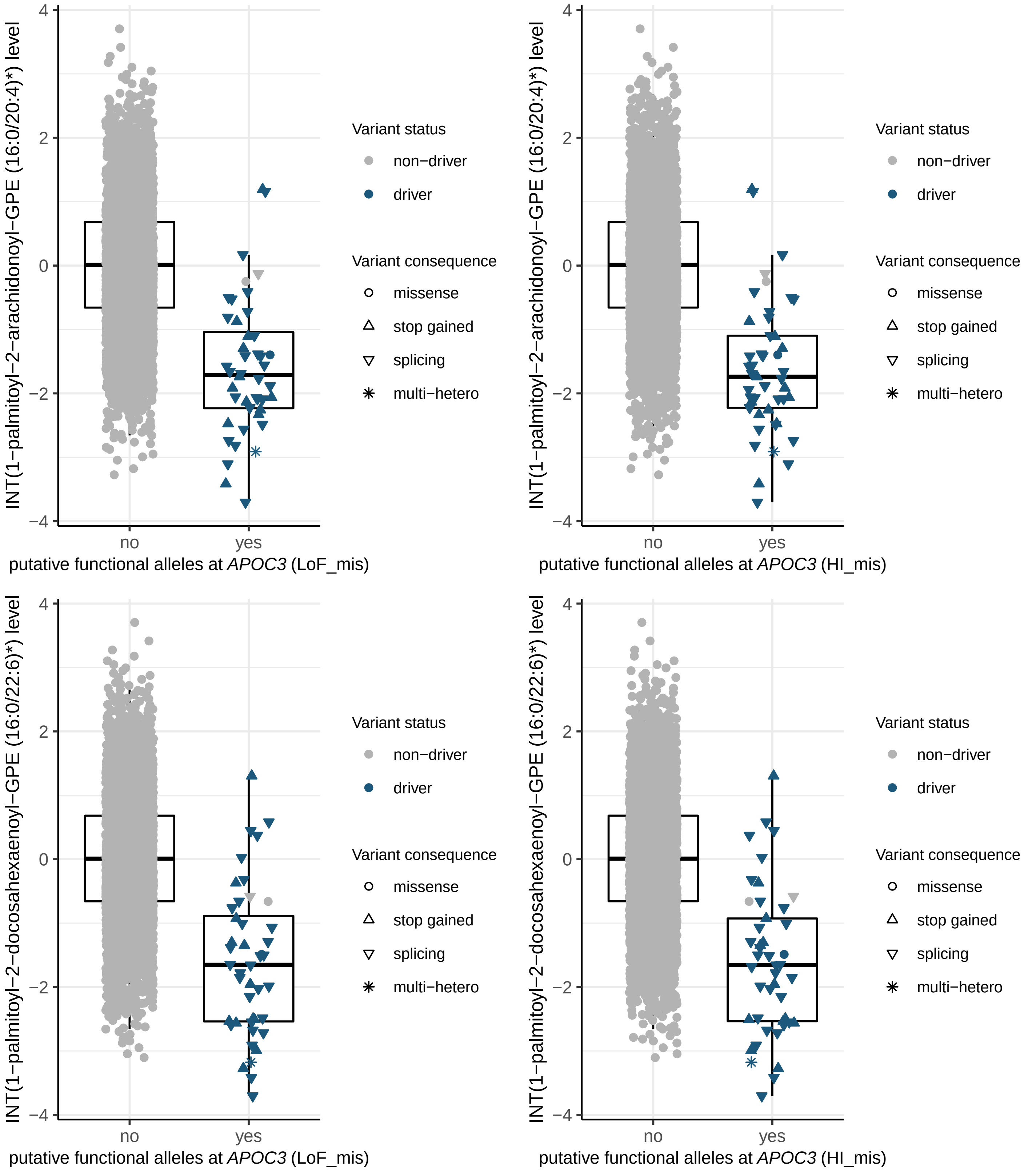
INT(1−palmitoyl−2−arachidonoyl−GPE (16:0/20:4)*) level
putative functional alleles at APOC3 (LoF_mis)
putative functional alleles at APOC3 (HI_mis)
INT(1−palmitoyl−2−docosahexaenoyl−GPE (16:0/22:6)*) level
no
yes
Variant status
non−driver
driver
Variant consequence
missense
stop gained
splicing
multi−hetero

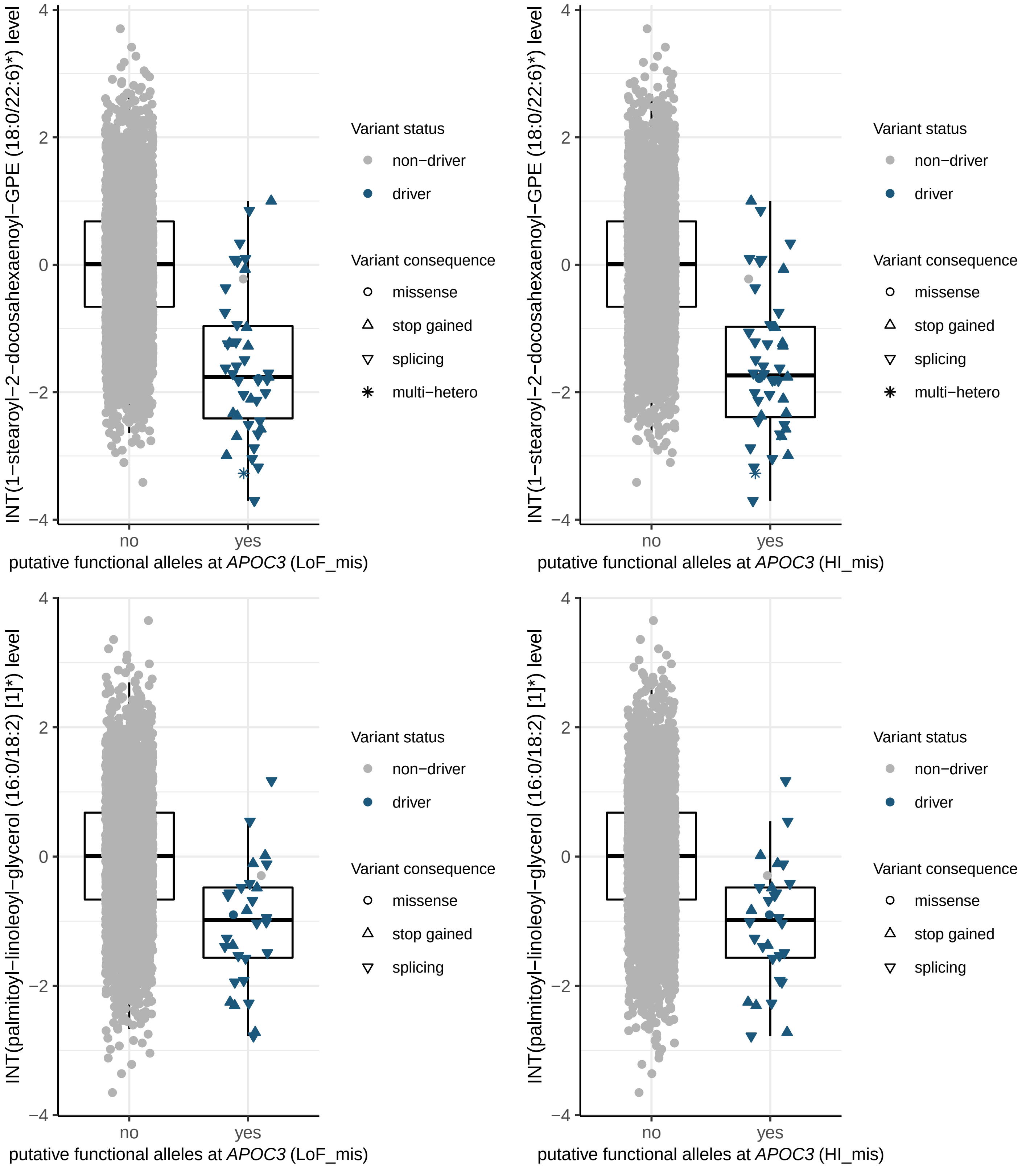
INT(1−stearoyl−2−docosahexaenoyl−GPE (18:0/22:6)*) level
INT(palmitoyl−linoleoyl−glycerol (16:0/18:2) [1]*) level
putative functional alleles at APOC3 (LoF_mis)
putative functional alleles at APOC3 (HI_mis)
no
yes
Variant status
non−driver
driver
Variant consequence
missense
stop gained
splicing
multi−hetero

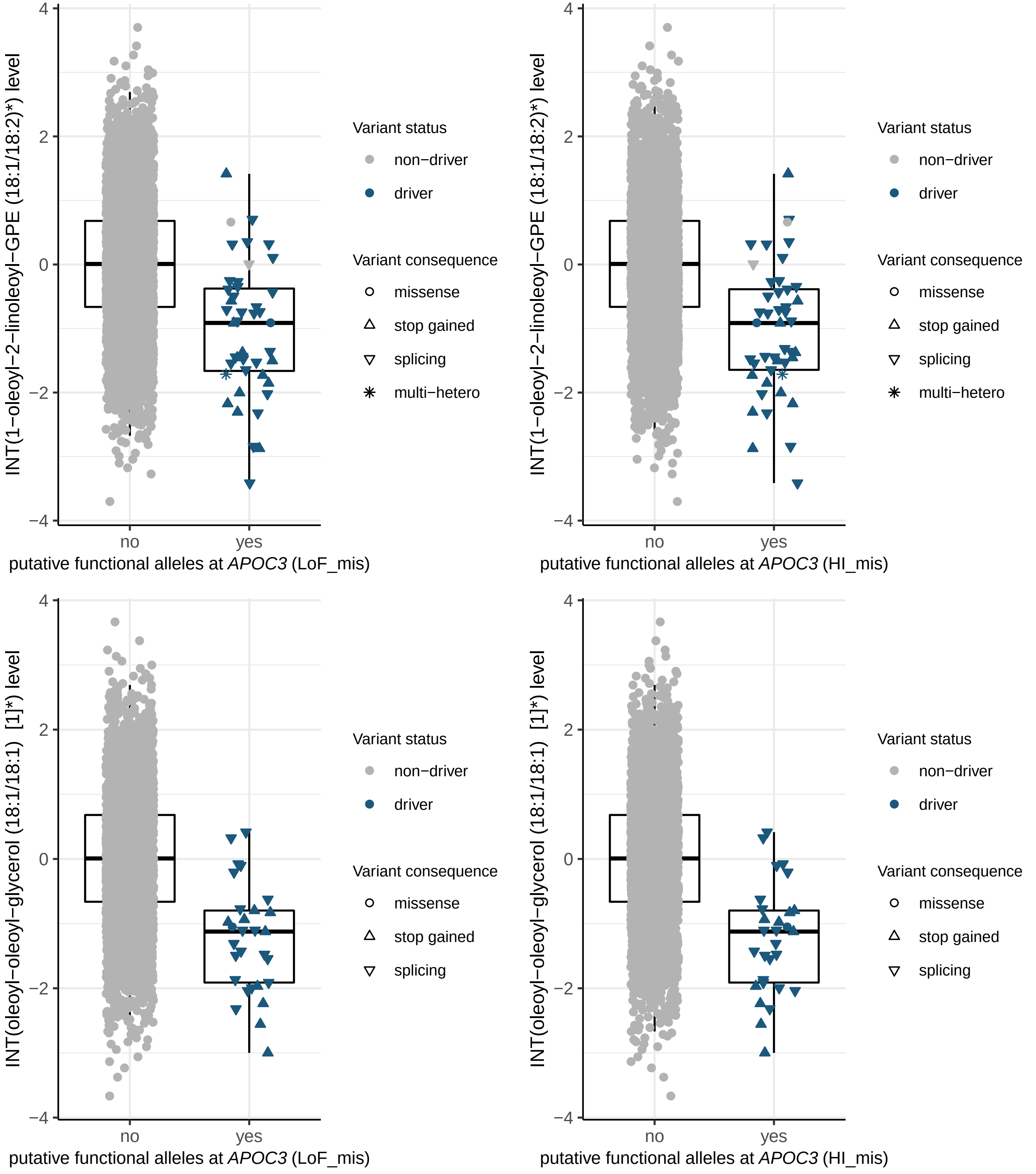

INT(1−oleoyl−2−linoleoyl−GPE (18:1/18:2)*) level
INT(oleoyl−oleoyl−glycerol (18:1/18:1) [1]*) level
4
2
0
−2
−4
no
yes
putative functional alleles at APOC3 (LoF_mis)
putative functional alleles at APOC3 (HI_mis)
Variant status
non−driver
driver
Variant consequence
missense
stop gained
splicing
multi−hetero

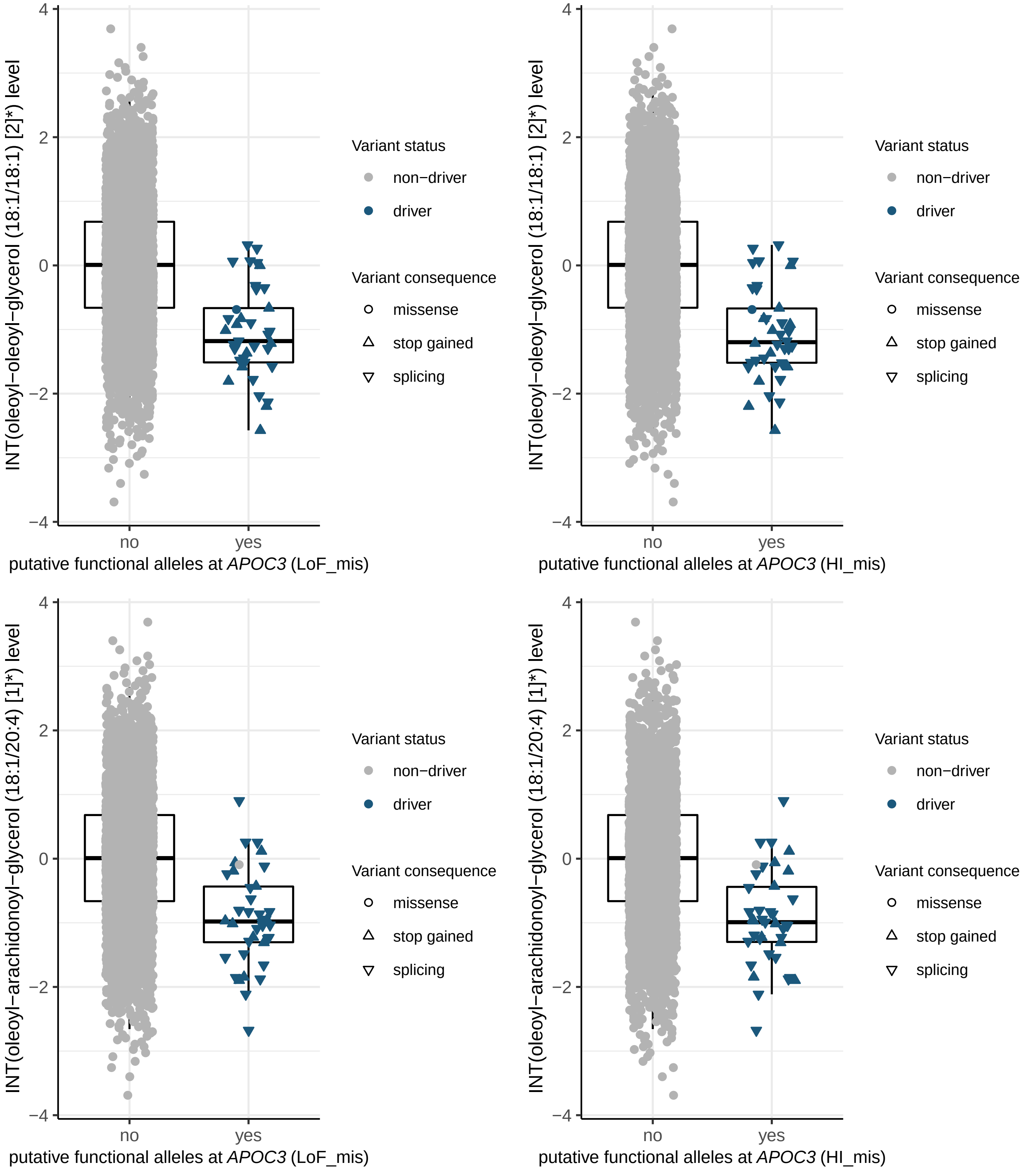
INT(oleoyl−oleoyl−glycerol (18:1/18:1) [2]*) level
4
2
0
−2
−4
no
yes
putative functional alleles at APOC3 (LoF_mis)
Variant status
non−driver
driver
Variant consequence
missense
stop gained
splicing
INT(oleoyl−oleoyl−glycerol (18:1/18:1) [2]*) level
putative functional alleles at APOC3 (HI_mis)
INT(oleoyl−arachidonoyl−glycerol (18:1/20:4) [1]*) level
putative functional alleles at APOC3 (LoF_mis)
INT(oleoyl−arachidonoyl−glycerol (18:1/20:4) [1]*) level
putative functional alleles at APOC3 (HI_mis)

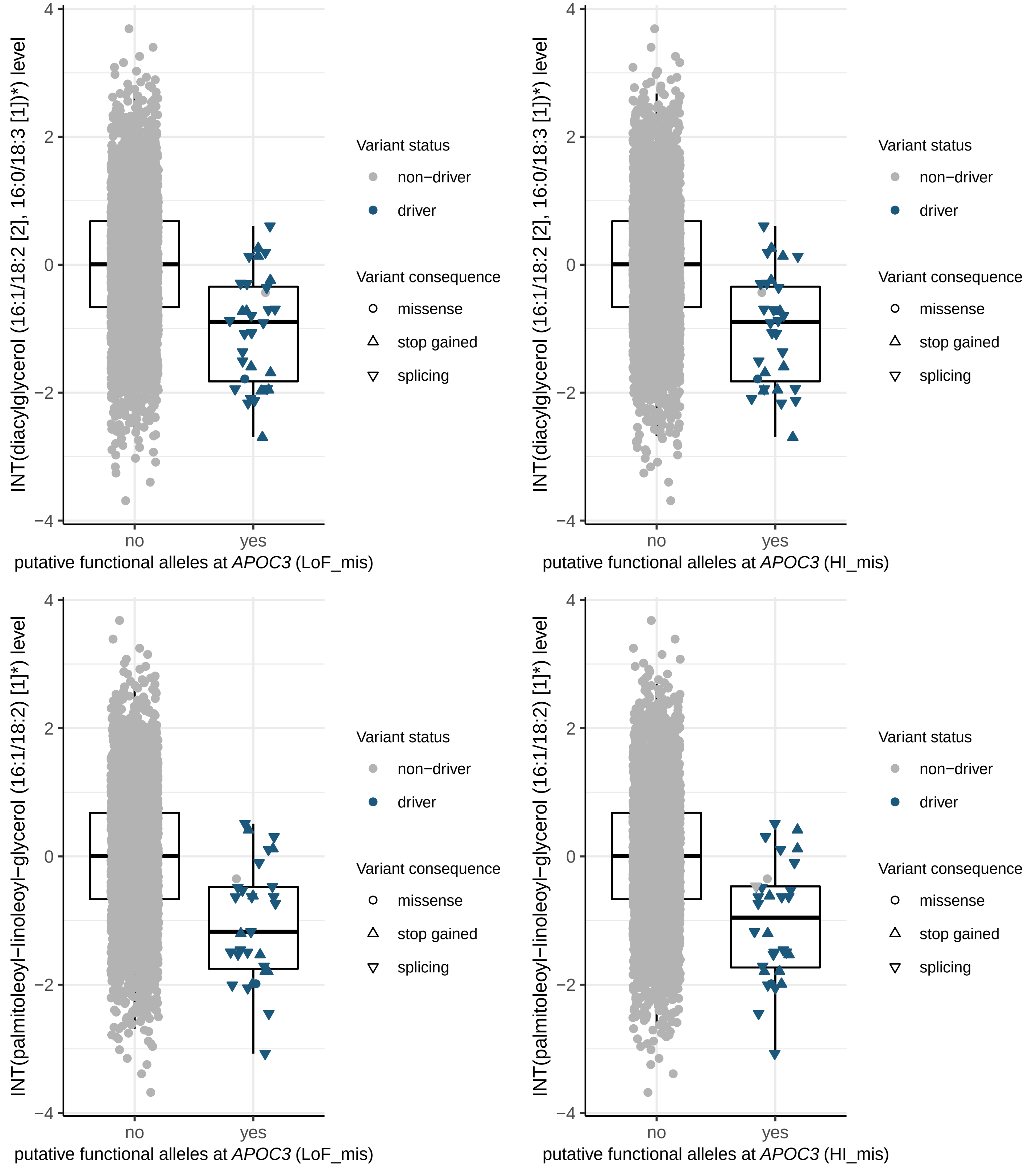
INT(diacylglycerol (16:1/18:2 [2], 16:0/18:3 [1])*) level
putative functional alleles at APOC3 (LoF_mis)
putative functional alleles at APOC3 (HI_mis)
INT(palmitoleoyl−linoleoyl−glycerol (16:1/18:2) [1]*) level
no
yes
4
2
0
−2
−4
Variant status
non−driver
driver
Variant consequence
missense
stop gained
splicing

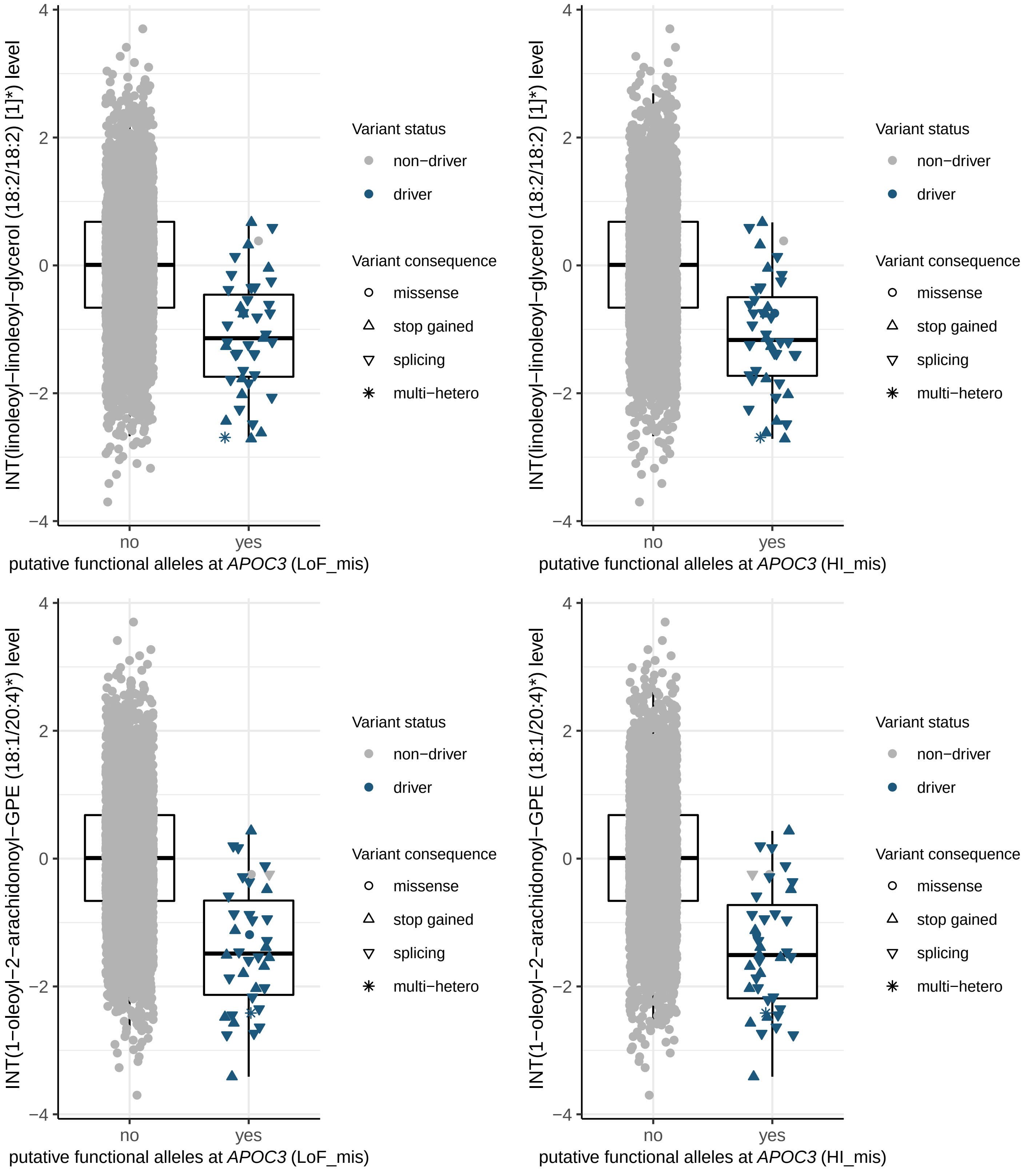

INT(linoleoyl−linoleoyl−glycerol (18:2/18:2) [1]*) level
putative functional alleles at APOC3 (LoF_mis)
putative functional alleles at APOC3 (HI_mis)
INT(1−oleoyl−2−arachidonoyl−GPE (18:1/20:4)*) level
no
yes
4
2
0
−2
−4
Variant status
non−driver
driver
Variant consequence
missense
stop gained
splicing
multi−hetero

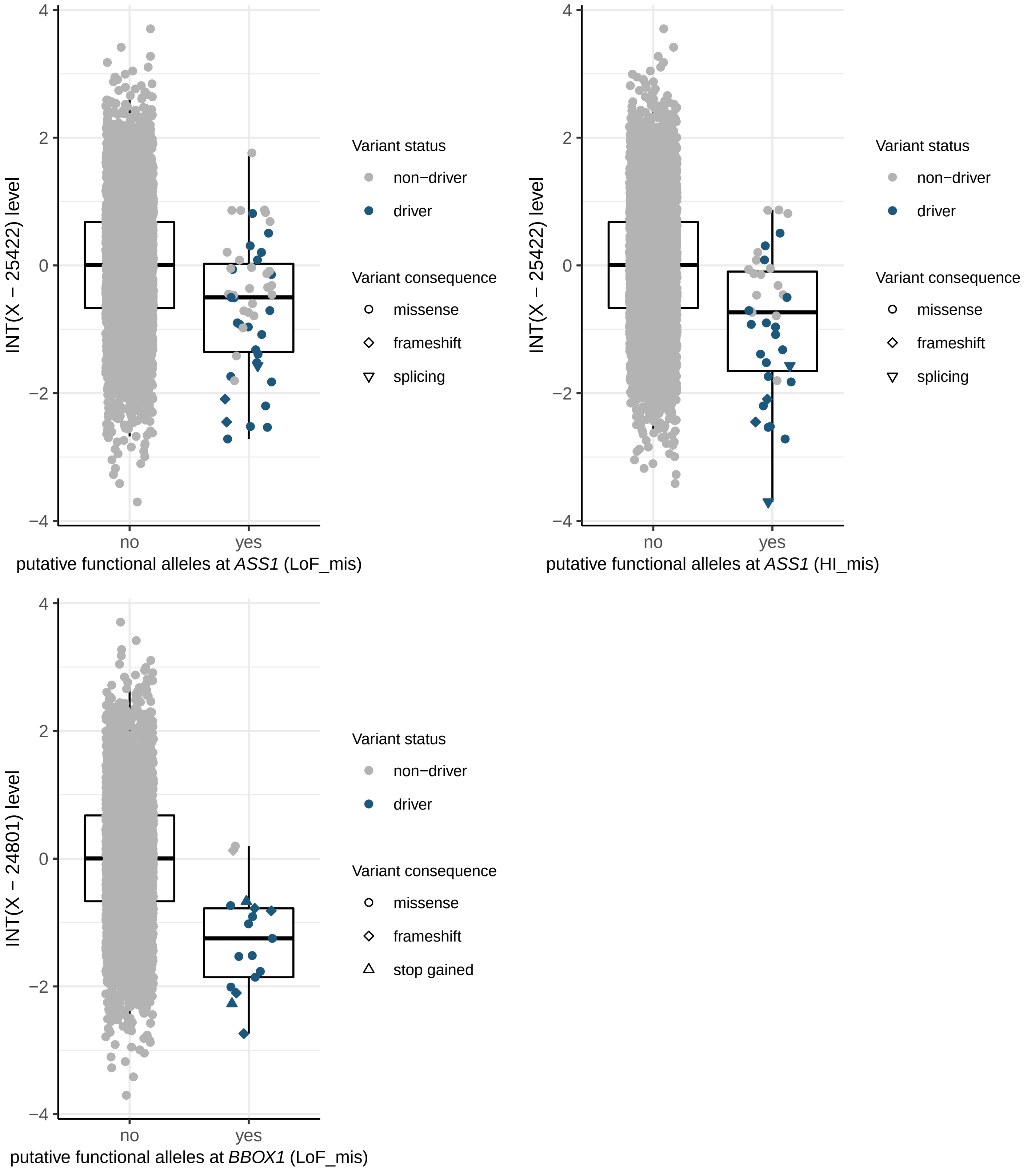

INT(X − 25422) level
4
2
0
−2
−4
no
yes
putative functional alleles at *ASS1* (LoF_mis)
Variant status
non−driver
driver
Variant consequence
missense
frameshift
splicing
INT(X − 25422) level
putative functional alleles at *ASS1* (HI_mis)
Variant status
non−driver
driver
Variant consequence
missense
frameshift
splicing
INT(X − 24801) level
putative functional alleles at *BBOX1* (LoF_mis)
Variant status
non−driver
driver
Variant consequence
missense
frameshift
stop gained

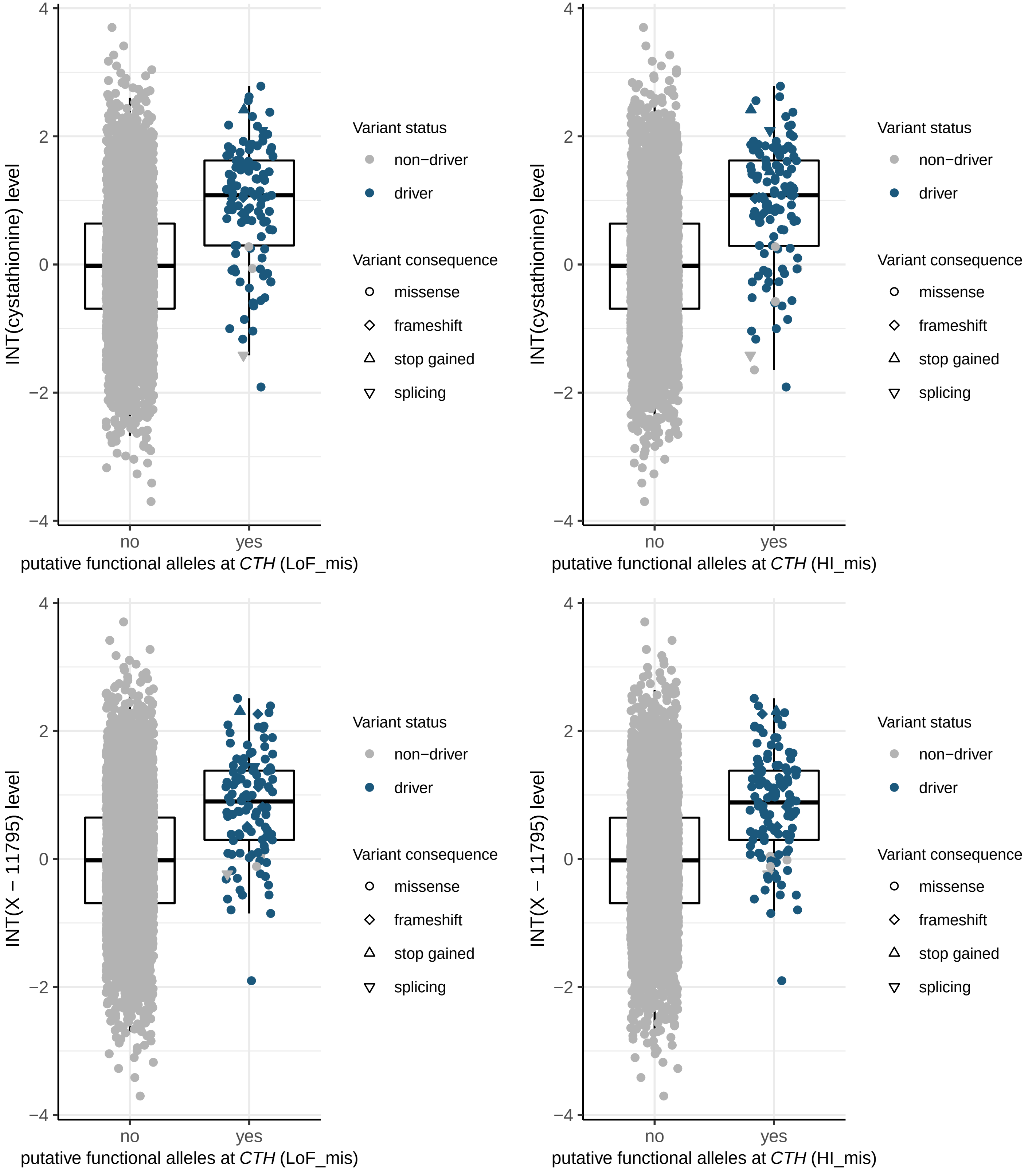

INT(cystathionine) level
putative functional alleles at *CTH* (LoF_mis)
putative functional alleles at *CTH* (HI_mis)
INT(X − 11795) level
no
yes
4
2
0
−2
−4
Variant status
non−driver
driver
Variant consequence
missense
frameshift
stop gained
splicing

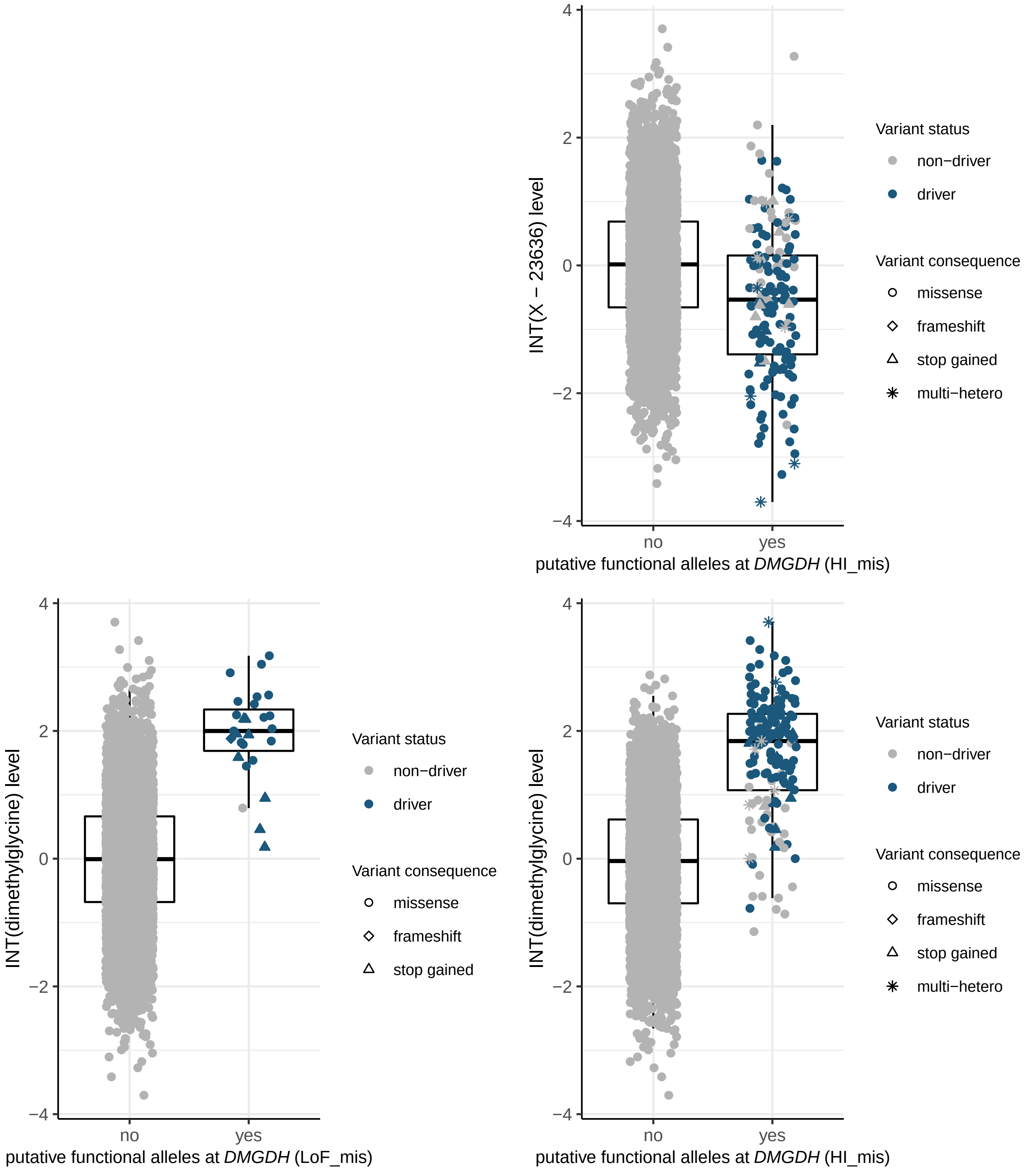

INT(X − 23636) level
4
2
0
−2
−4
no
yes
putative functional alleles at *DMGDH* (HI_mis)
Variant status
non−driver
driver
Variant consequence
missense
frameshift
stop gained
multi−hetero
INT(dimethylglycine) level
4
2
0
−2
−4
no
yes
putative functional alleles at *DMGDH* (LoF_mis)
Variant status
non−driver
driver
Variant consequence
missense
frameshift
stop gained
INT(dimethylglycine) level
4
2
0
−2
−4
no
yes
putative functional alleles at *DMGDH* (HI_mis)
Variant status
non−driver
driver
Variant consequence
missense
frameshift
stop gained
multi−hetero

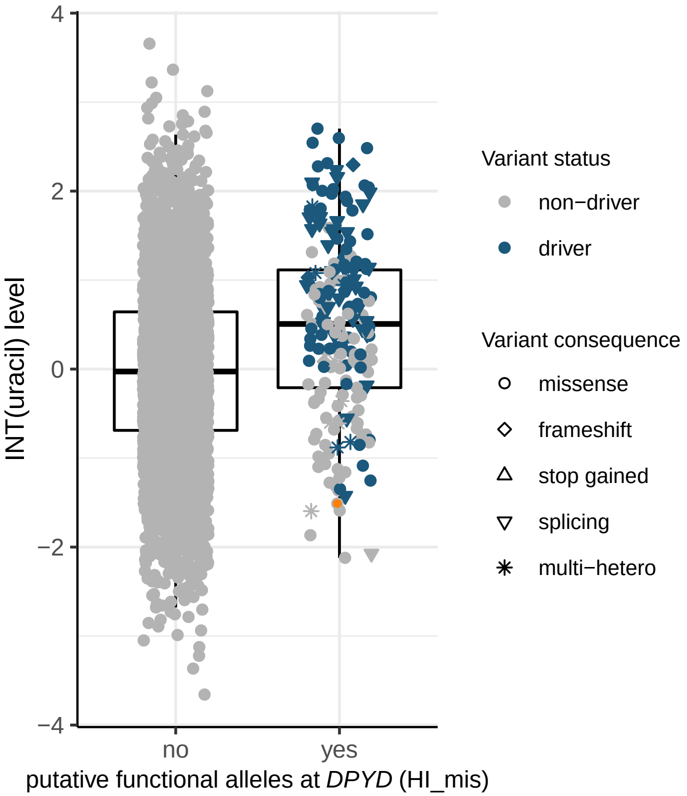

4
2
0
−2
−4
INT(uracil) level
no
yes
putative functional alleles at *DPYD* (HI_mis)
Variant status
non−driver
driver
Variant consequence
missense
frameshift
stop gained
splicing
multi−hetero


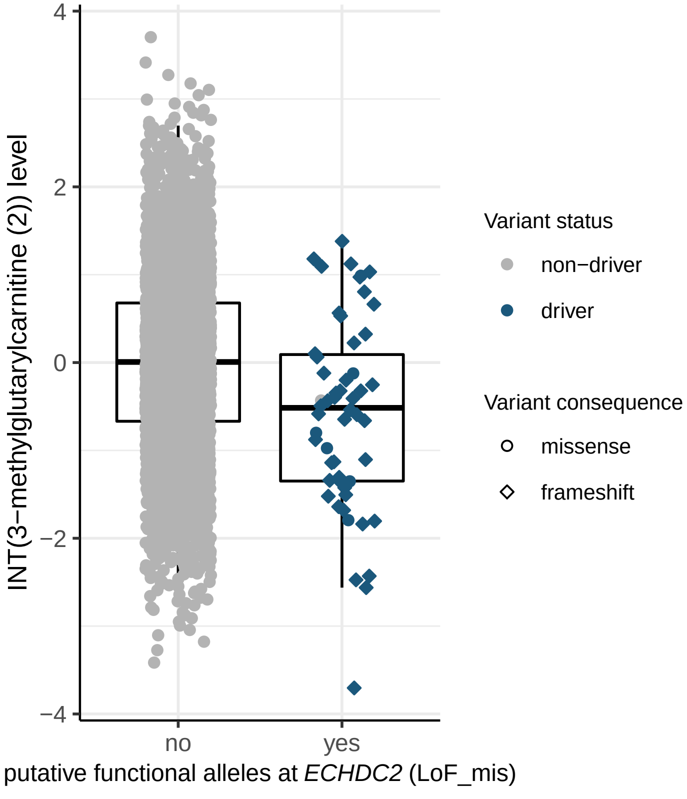

4
2
0
−2
−4
INT(3−methylglutarylcarnitine (2)) level
no
yes
putative functional alleles at *ECHDC2* (LoF_mis)
Variant status
non−driver
driver
Variant consequence
missense
frameshift

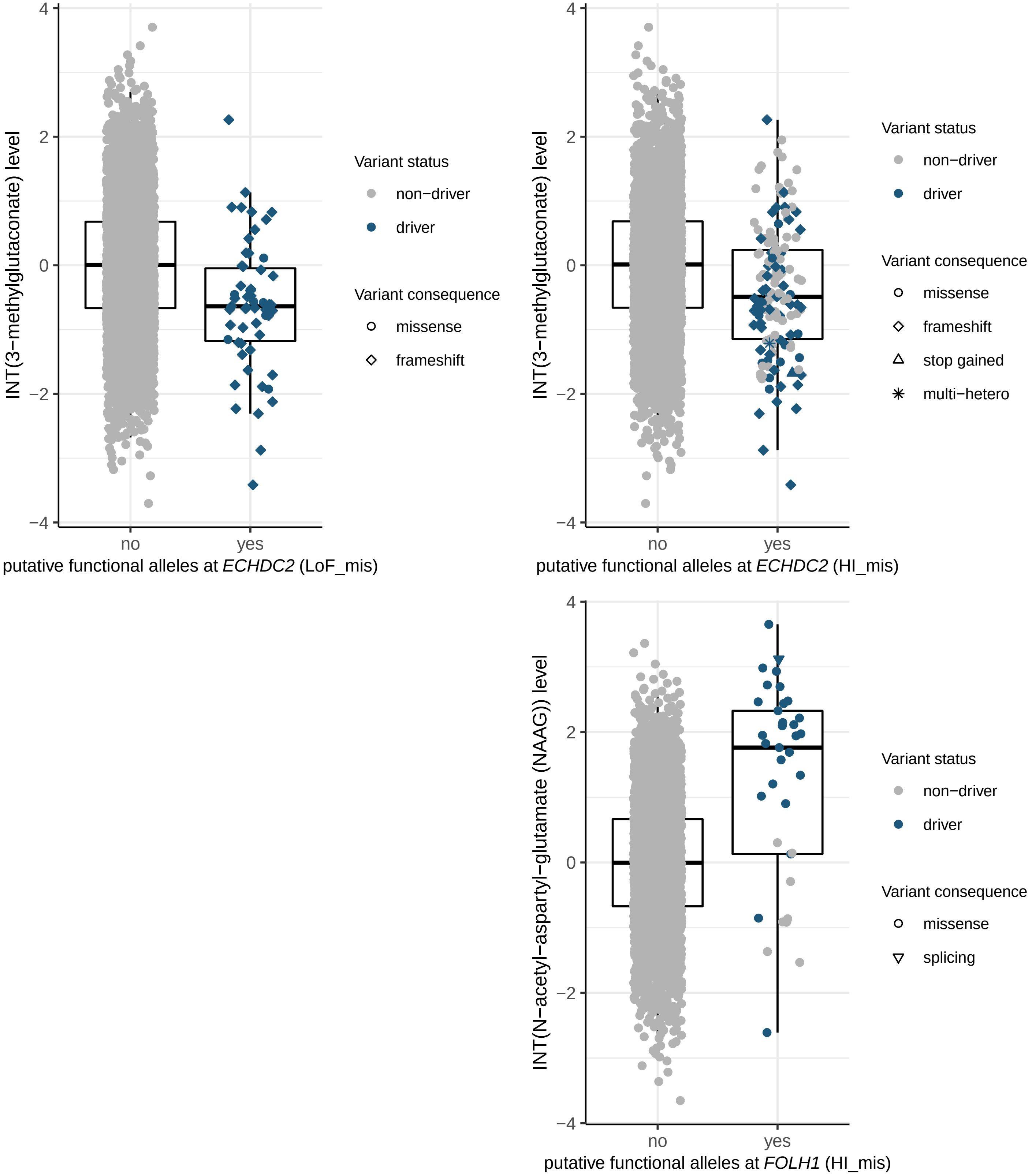
INT(3−methylglutaconate) level
4
2
0
−2
−4
no
yes
putative functional alleles at *ECHDC2* (LoF_mis)
Variant status
non−driver
driver
Variant consequence
missense
frameshift
INT(3−methylglutaconate) level
no
yes
putative functional alleles at *ECHDC2* (HI_mis)
Variant status
non−driver
driver
Variant consequence
missense
frameshift
stop gained
multi−hetero
INT(N−acetyl−aspartyl−glutamate (NAAG)) level
no
yes
putative functional alleles at *FOLH1* (HI_mis)
Variant status
non−driver
driver
Variant consequence
missense
splicing

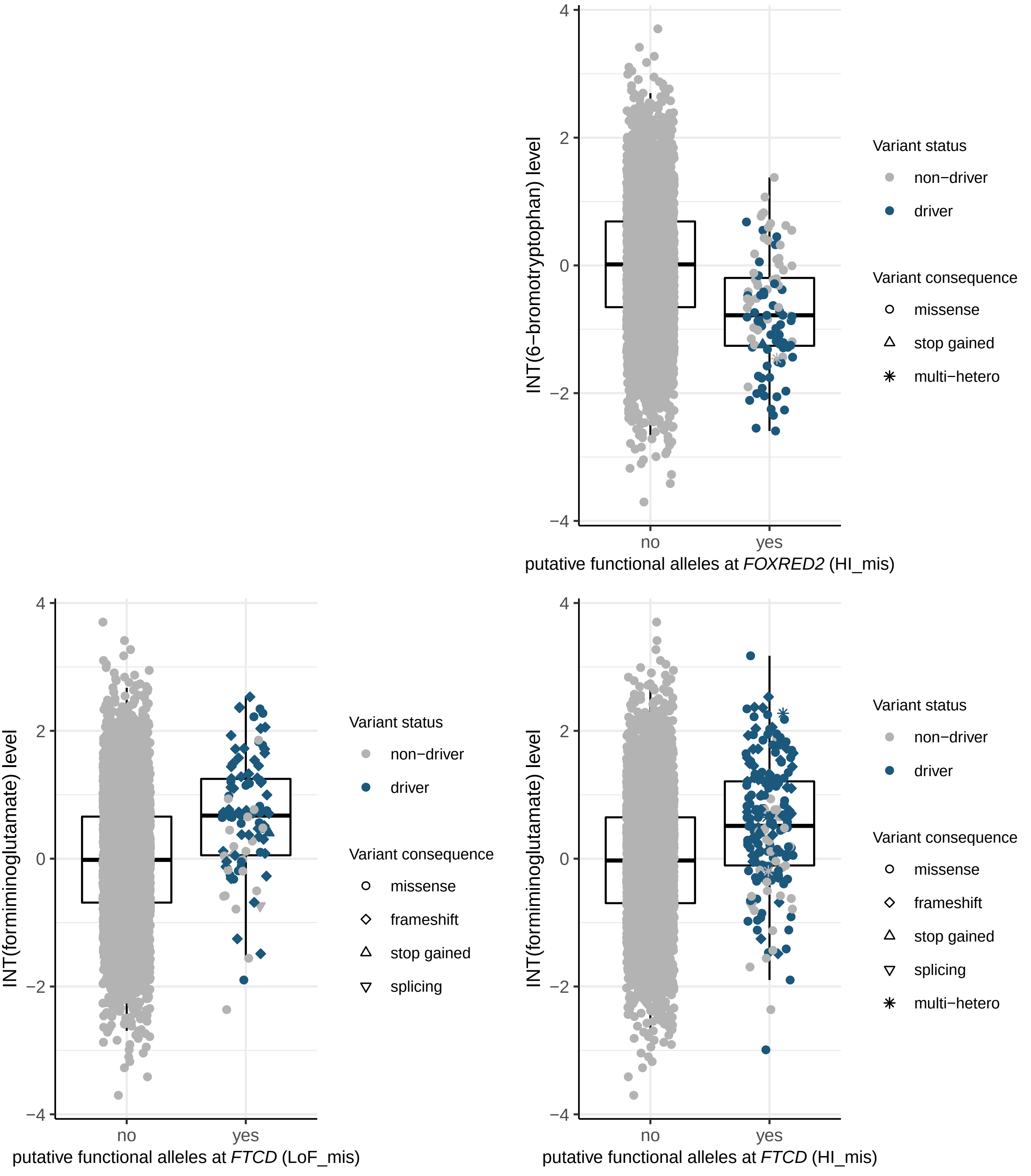

INT(6−bromotryptophan) level
putative functional alleles at FOXRED2 (HI_mis)
no
yes
Variant status
non−driver
driver
Variant consequence
missense
stop gained
multi−hetero
INT(formiminoglutamate) level
putative functional alleles at FTCD (LoF_mis)
Variant status
non−driver
driver
Variant consequence
missense
frameshift
stop gained
splicing
INT(formiminoglutamate) level
putative functional alleles at FTCD (HI_mis)
Variant status
non−driver
driver
Variant consequence
missense
frameshift
stop gained
splicing
multi−hetero

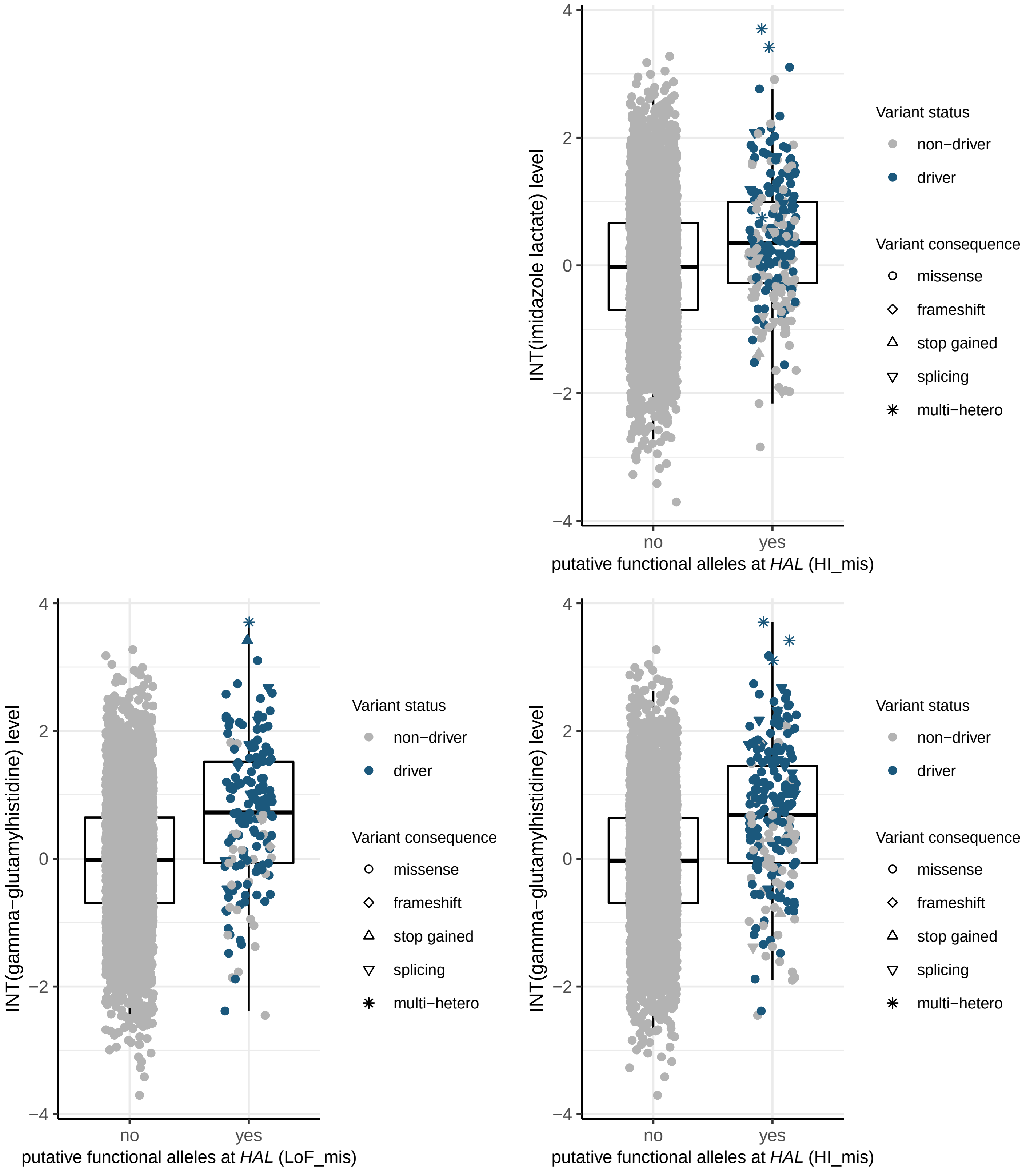

INT(imidazole lactate) level
4
2
0
−2
−4
no
yes
putative functional alleles at *HAL* (HI_mis)
Variant status
non−driver
driver
Variant consequence
missense
frameshift
stop gained
splicing
multi−hetero
INT(gamma−glutamylhistidine) level
4
2
0
−2
−4
no
yes
putative functional alleles at *HAL* (LoF_mis)
Variant status
non−driver
driver
Variant consequence
missense
frameshift
stop gained
splicing
multi−hetero
INT(gamma−glutamylhistidine) level
4
2
0
−2
−4
no
yes
putative functional alleles at *HAL* (HI_mis)
Variant status
non−driver
driver
Variant consequence
missense
frameshift
stop gained
splicing
multi−hetero

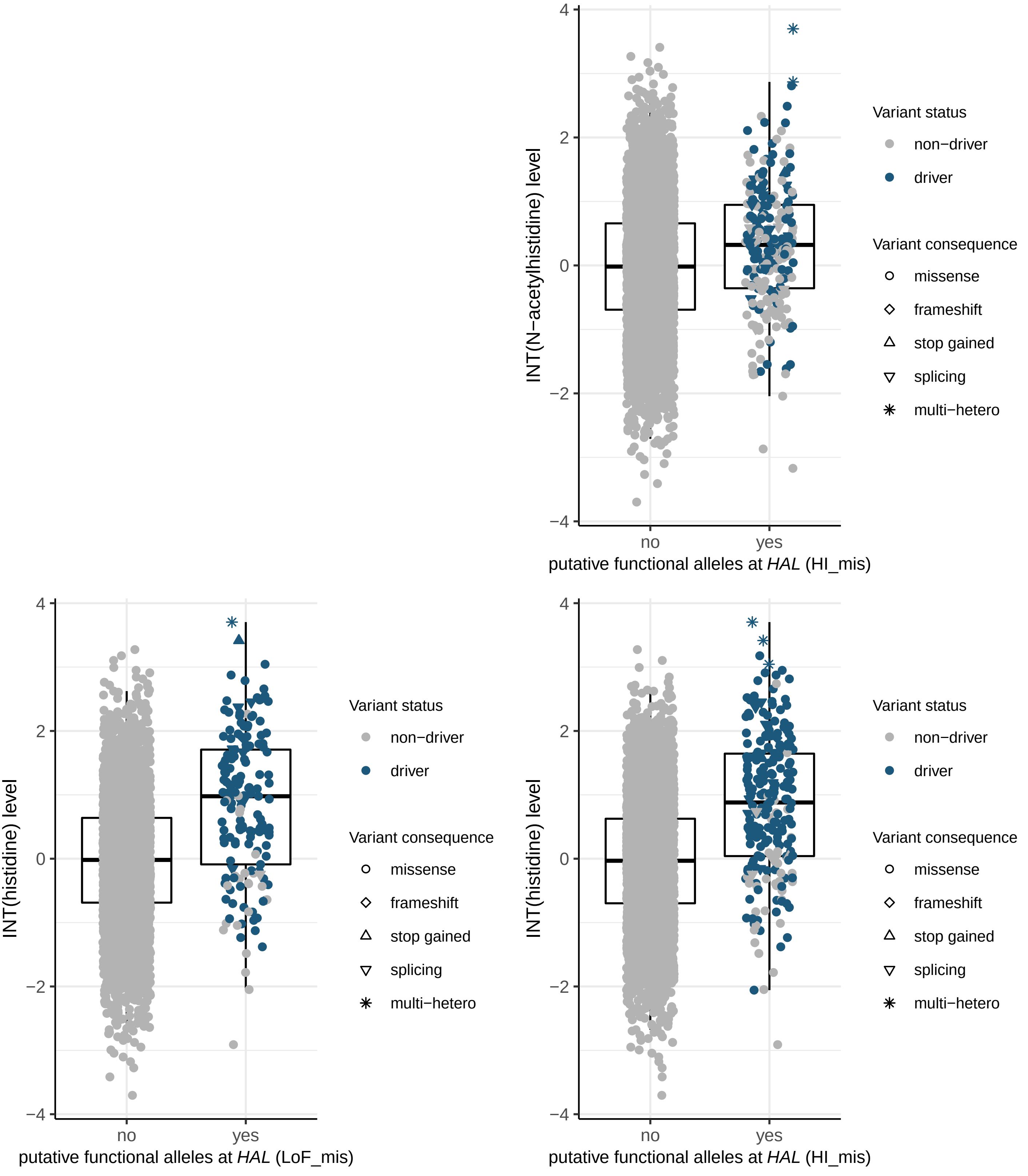
INT(N−acetylhistidine) level
putative functional alleles at HAL (HI_mis)
no
yes
4
2
0
−2
−4
Variant status
non−driver
driver
Variant consequence
missense
frameshift
stop gained
splicing
multi−hetero
INT(histidine) level
putative functional alleles at HAL (LoF_mis)
INT(histidine) level
putative functional alleles at HAL (HI_mis)

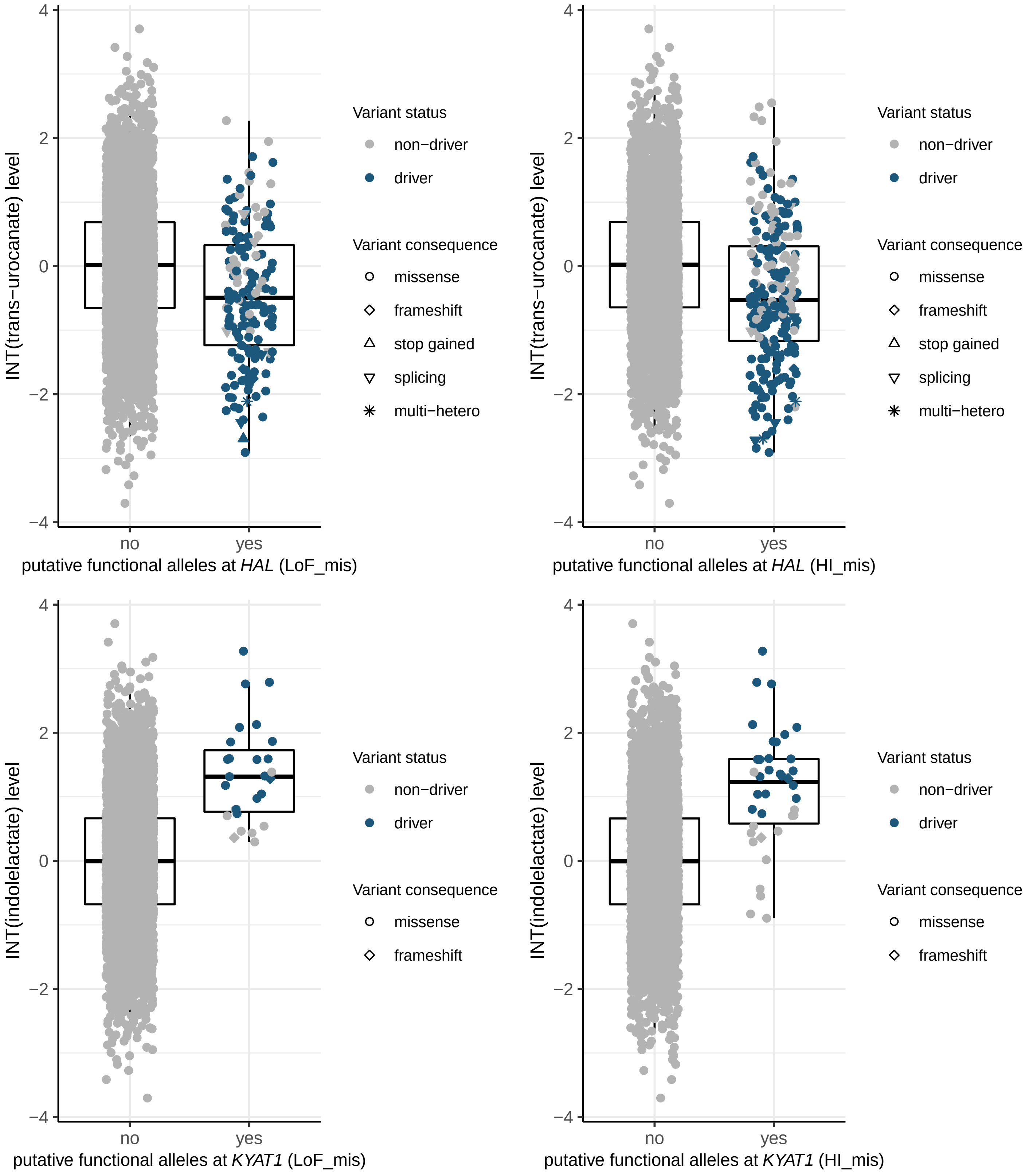
INT(trans−urocanate) level
putative functional alleles at HAL (LoF_mis)
putative functional alleles at HAL (HI_mis)
INT(indolelactate) level
putative functional alleles at KYAT1 (LoF_mis)
putative functional alleles at KYAT1 (HI_mis)
no
yes
4
2
0
−2
−4
Variant status
non−driver
driver
Variant consequence
missense
frameshift
stop gained
splicing
multi−hetero

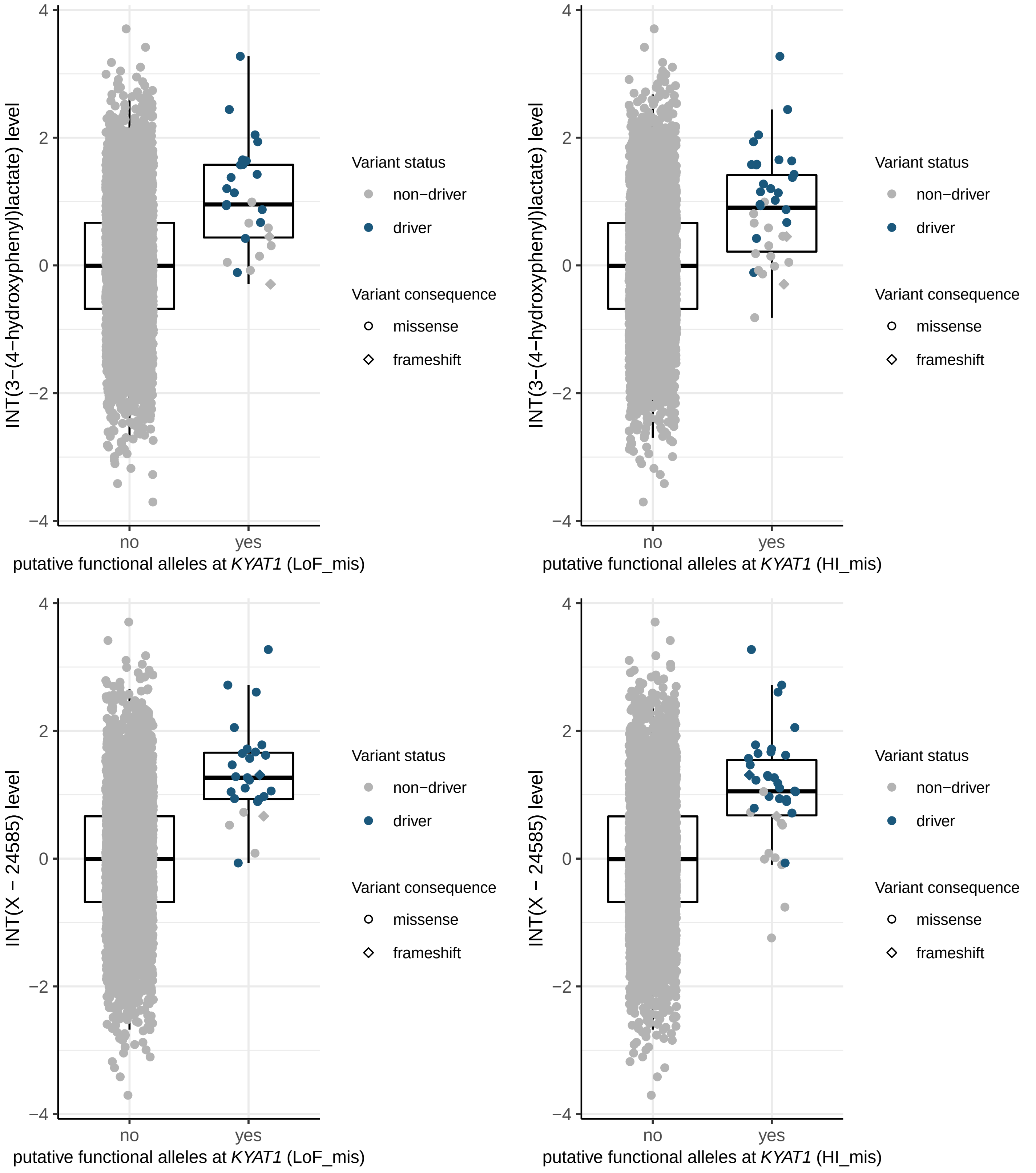

INT(3−(4−hydroxyphenyl)lactate) level
putative functional alleles at KYAT1 (LoF_mis)
putative functional alleles at KYAT1 (HI_mis)
INT(X − 24585) level
no
yes
Variant status
non−driver
driver
Variant consequence
missense
frameshift

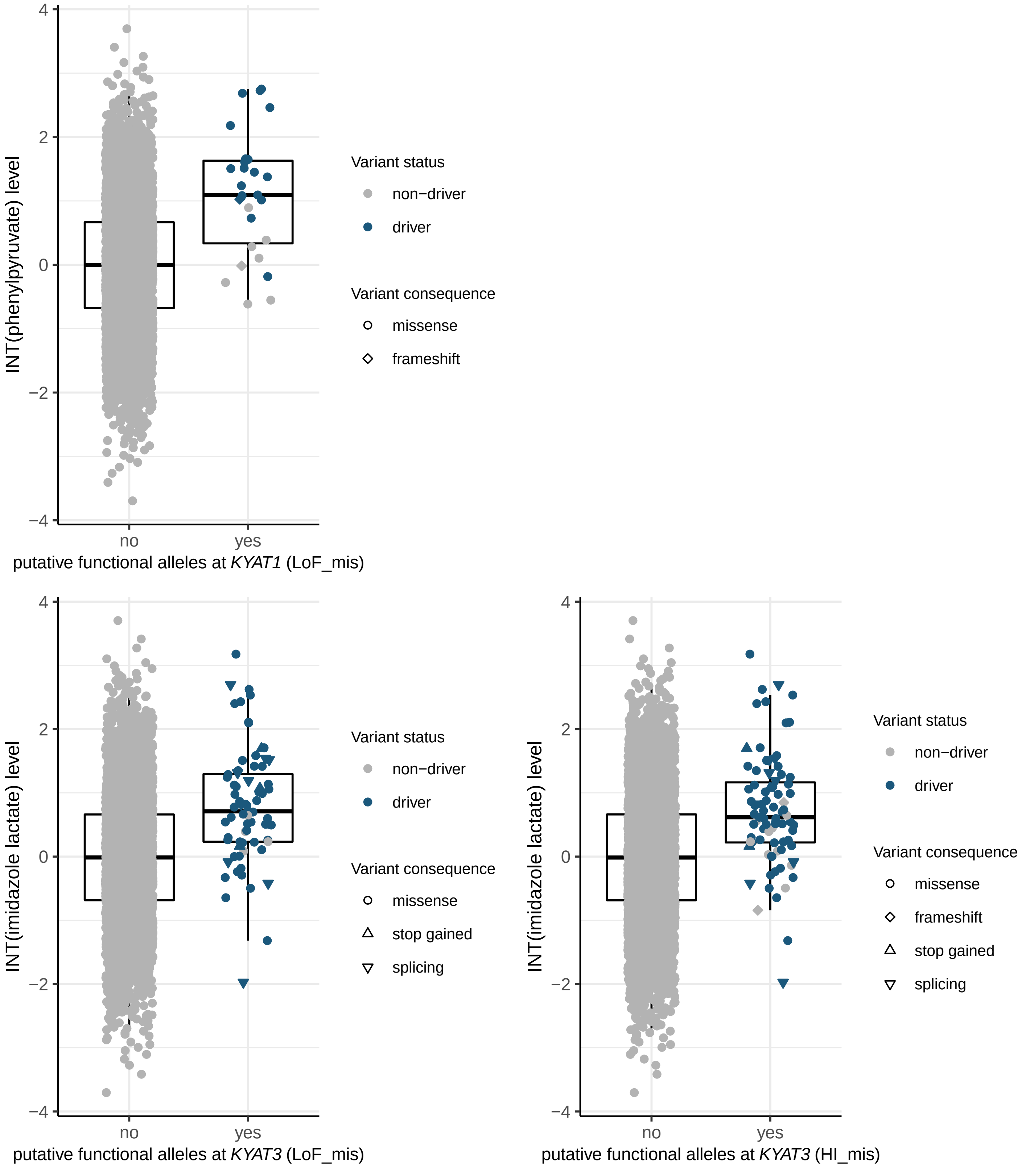
INT(phenylpyruvate) level
putative functional alleles at KYAT1 (LoF_mis)
no
yes
4
2
0
−2
−4
Variant status
non−driver
driver
Variant consequence
missense
frameshift
INT(imidazole lactate) level
putative functional alleles at KYAT3 (LoF_mis)
Variant status
non−driver
driver
Variant consequence
missense
stop gained
splicing
INT(imidazole lactate) level
putative functional alleles at KYAT3 (HI_mis)
Variant status
non−driver
driver
Variant consequence
missense
frameshift
stop gained
splicing

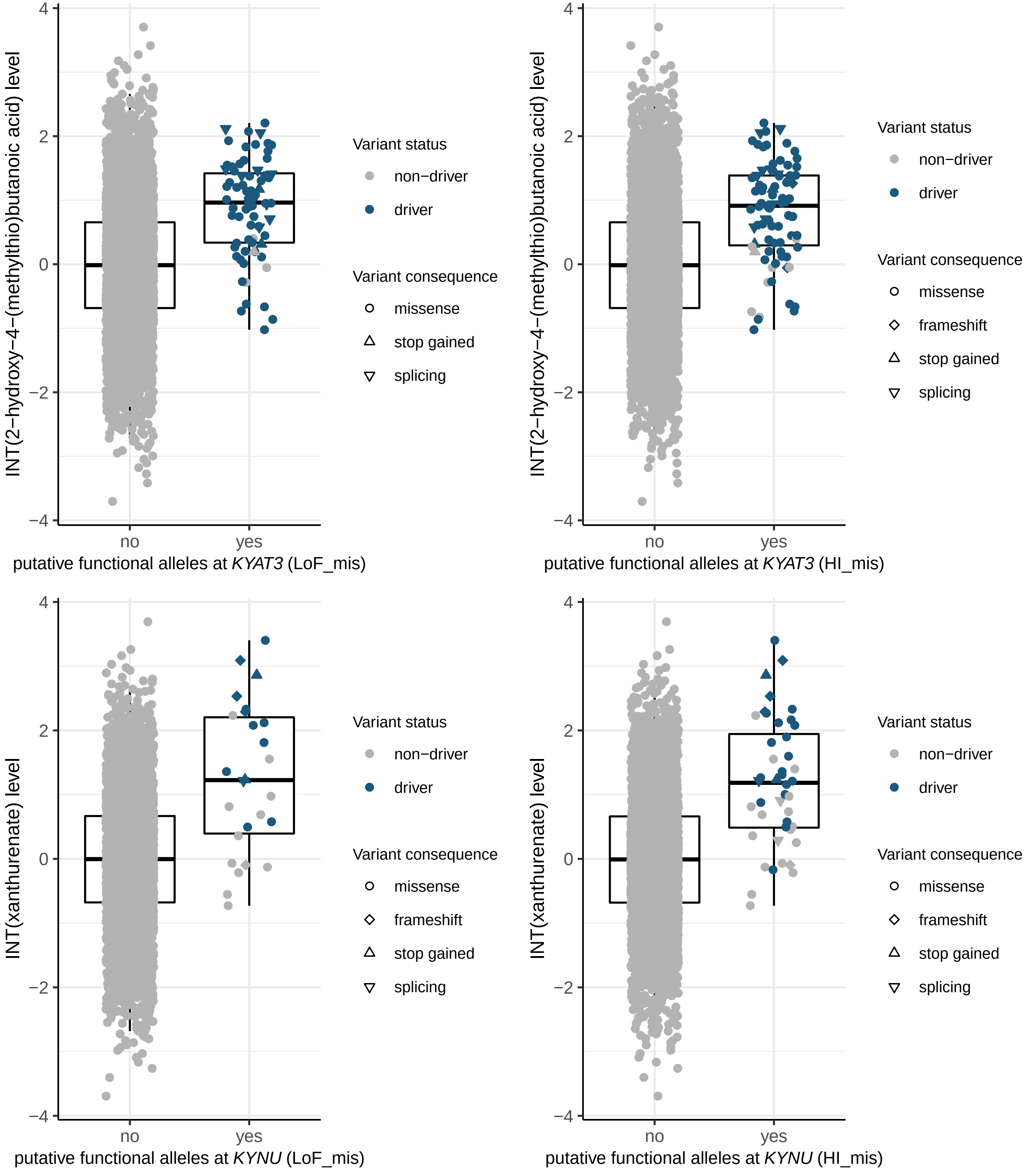
INT(2−hydroxy−4−(methylthio)butanoic acid) level
putative functional alleles at KYAT3 (LoF_mis)
putative functional alleles at KYAT3 (HI_mis)
INT(xanthurenate) level
putative functional alleles at KYNU (LoF_mis)
putative functional alleles at KYNU (HI_mis)
no
yes
4
2
0
−2
−4
Variant status
non−driver
driver
Variant consequence
missense
frameshift
stop gained
splicing

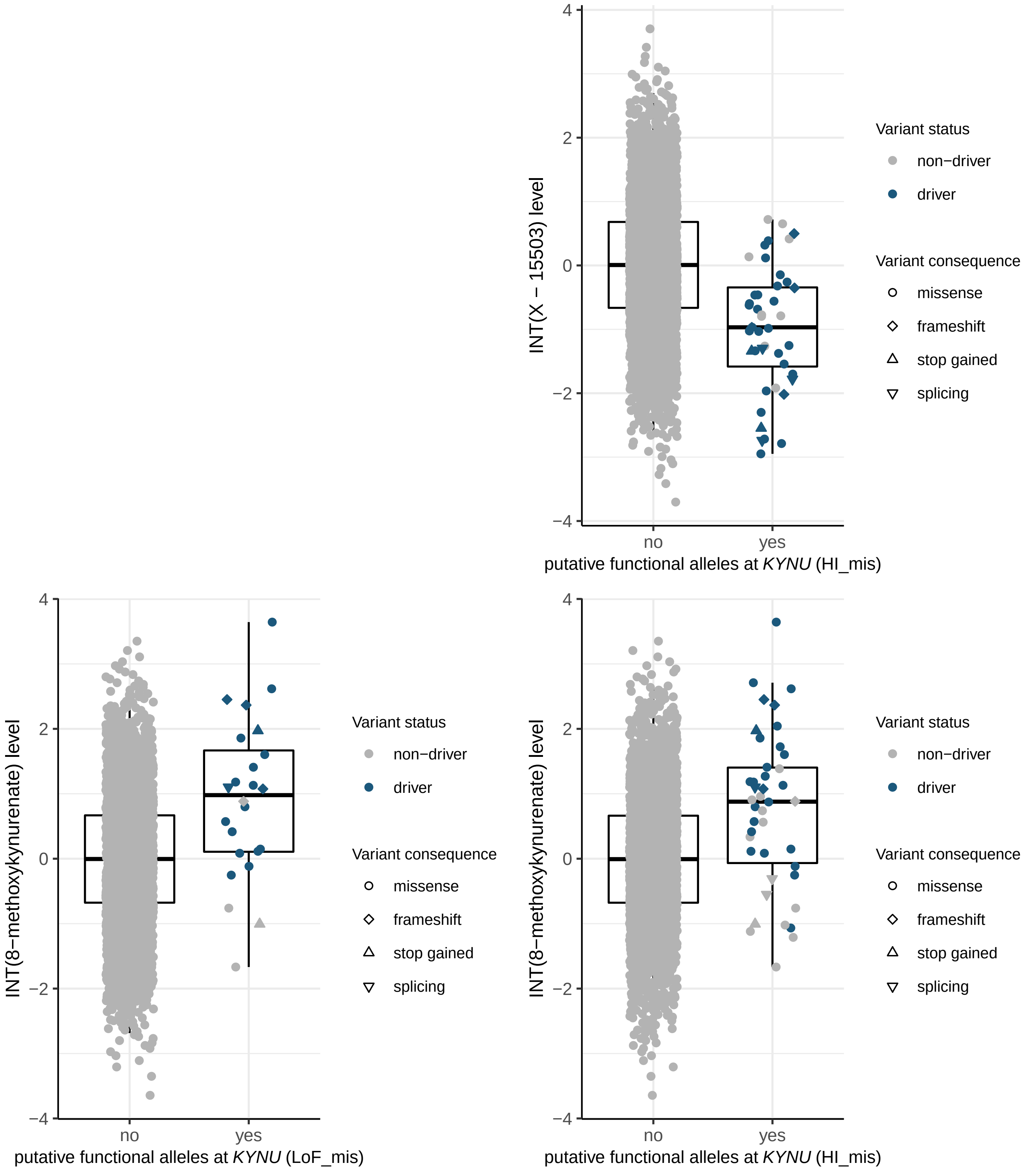

INT(X − 15503) level
putative functional alleles at KYNU (HI_mis)
INT(8−methoxykynurenate) level
putative functional alleles at KYNU (LoF_mis)
putative functional alleles at KYNU (HI_mis)
no
yes
Variant status
non−driver
driver
Variant consequence
missense
frameshift
stop gained
splicing

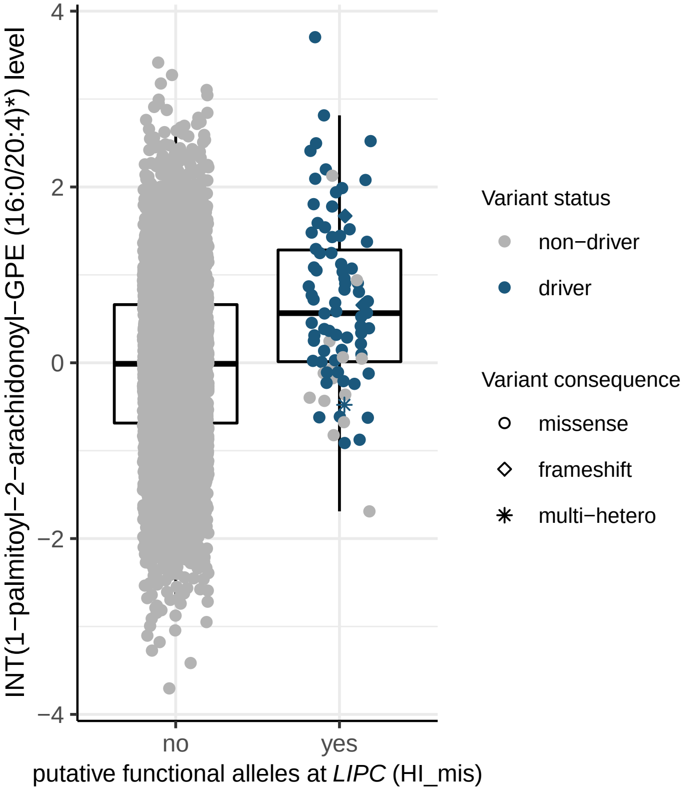
INT(1−palmitoyl−2−arachidonoyl−GPE (16:0/20:4)*) level
4
2
0
−2
−4
no
yes
putative functional alleles at LIPC (HI_mis)
Variant status
non−driver
driver
Variant consequence
missense
frameshift
multi−hetero

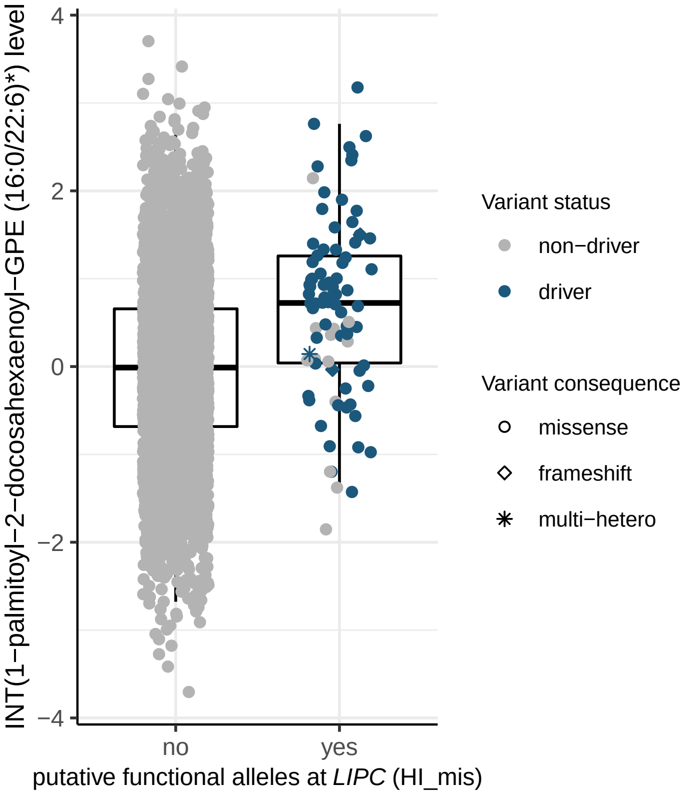
INT(1−palmitoyl−2−docosahexaenoyl−GPE (16:0/22:6)*) level
4
2
0
−2
−4
no
yes
putative functional alleles at LIPC (HI_mis)
Variant status
non−driver
driver
Variant consequence
missense
frameshift
multi−hetero

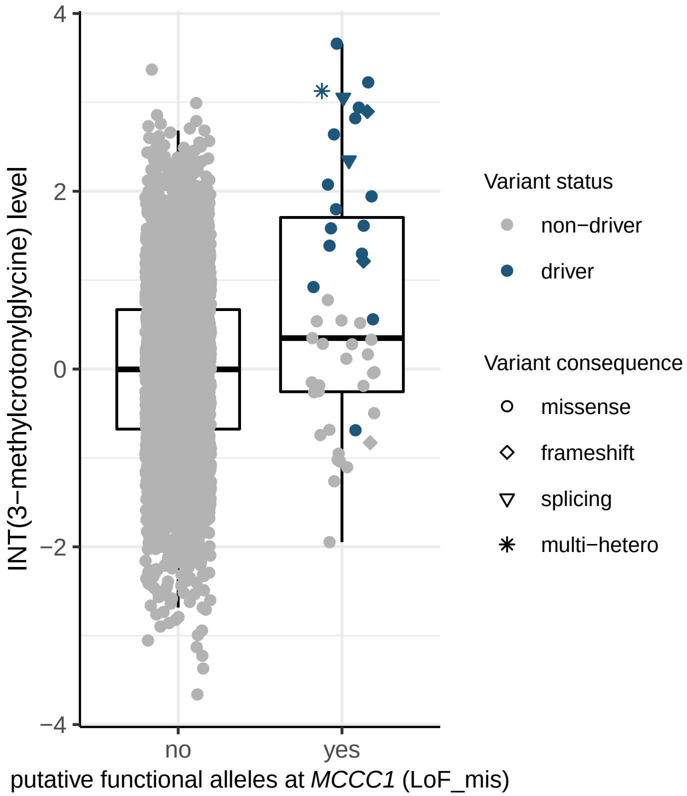
INT(3−methylcrotonylglycine) level
4
2
0
−2
−4
no
yes
putative functional alleles at MCCC1 (LoF_mis)
Variant status
non−driver
driver
Variant consequence
missense
frameshift
splicing
multi−hetero

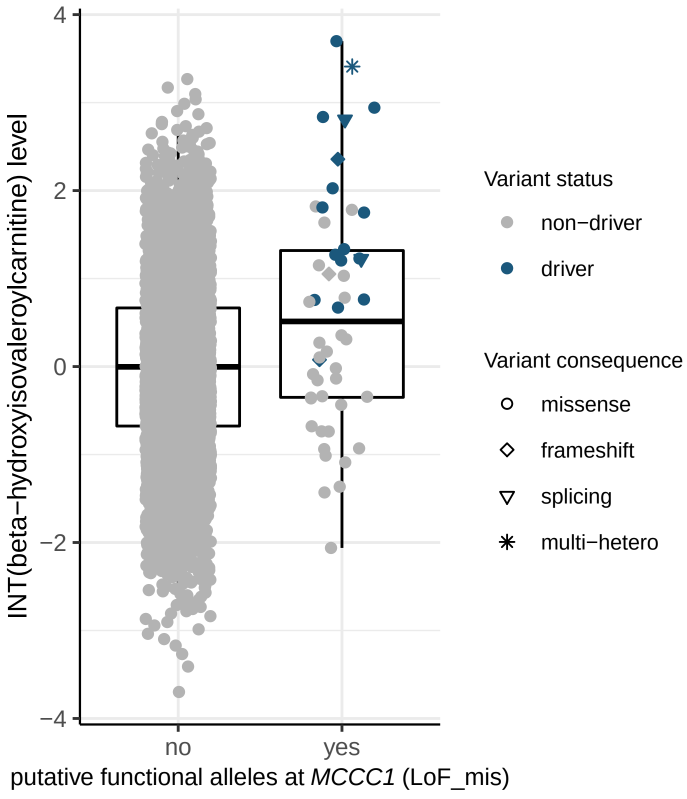
INT(beta−hydroxyisovaleroylcarnitine) level
4
2
0
−2
−4
no
yes
putative functional alleles at MCCC1 (LoF_mis)
Variant status
non−driver
driver
Variant consequence
missense
frameshift
splicing
multi−hetero

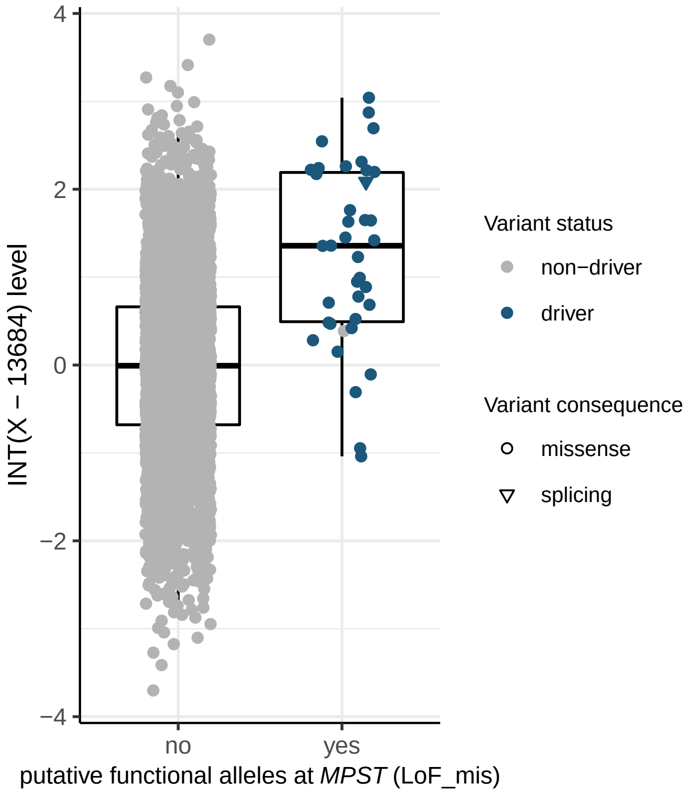

INT(X − 13684) level
4
2
0
−2
−4
no
yes
putative functional alleles at MPST (LoF_mis)
Variant status
non−driver
driver
Variant consequence
missense
splicing


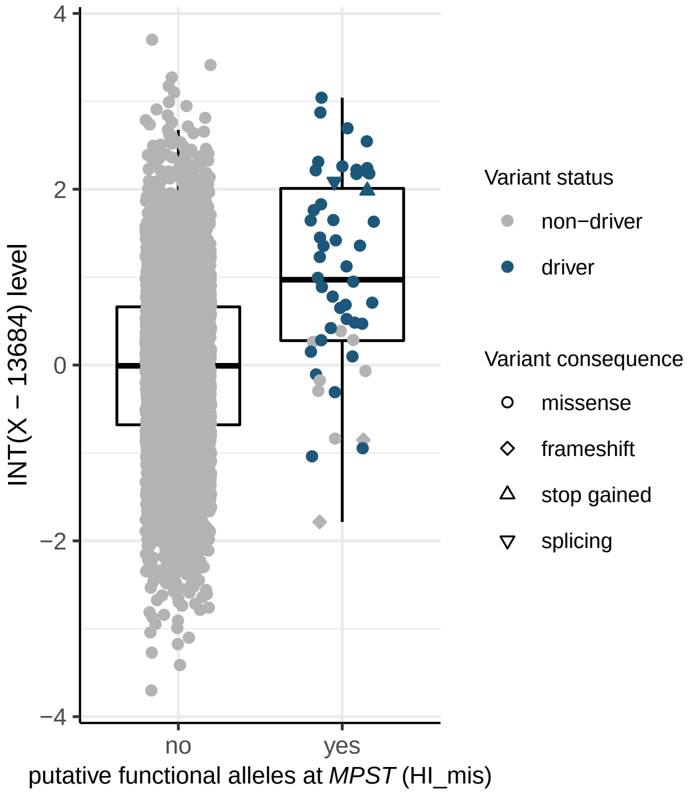

INT(X − 13684) level
4
2
0
−2
−4
no
yes
putative functional alleles at MPST (HI_mis)
Variant status
non−driver
driver
Variant consequence
missense
frameshift
stop gained
splicing


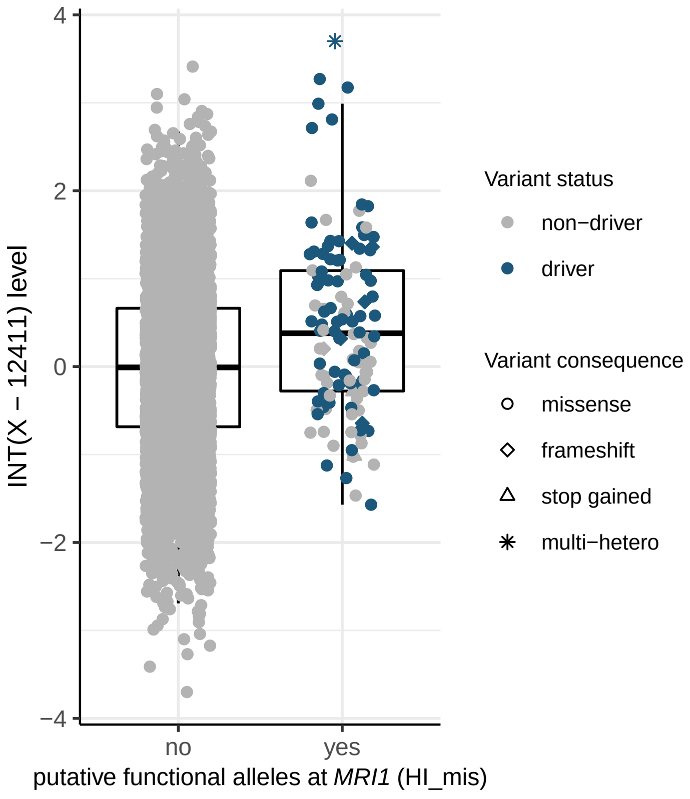

INT(X − 12411) level
4
2
0
−2
−4
no
yes
putative functional alleles at MRI1 (HI_mis)
Variant status
non−driver
driver
Variant consequence
missense
frameshift
stop gained
multi−hetero

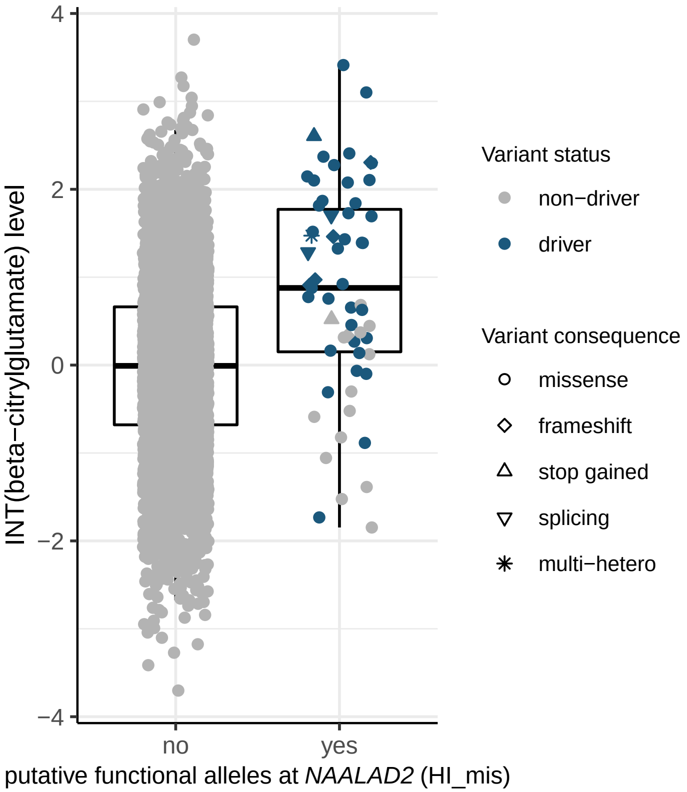
INT(beta−citrylglutamate) level
4
2
0
−2
−4
no
yes
putative functional alleles at *NAALAD2* (HI_mis)
Variant status
non−driver
driver
Variant consequence
missense
frameshift
stop gained
splicing
multi−hetero

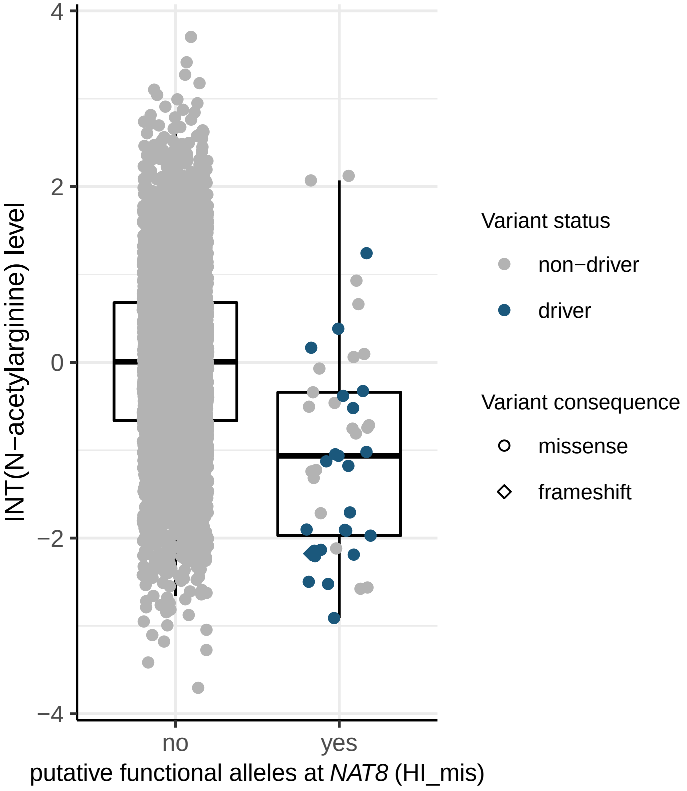
INT(N−acetylarginine) level
4
2
0
−2
−4
no
yes
putative functional alleles at *NAT8* (HI_mis)
Variant status
non−driver
driver
Variant consequence
missense
frameshift

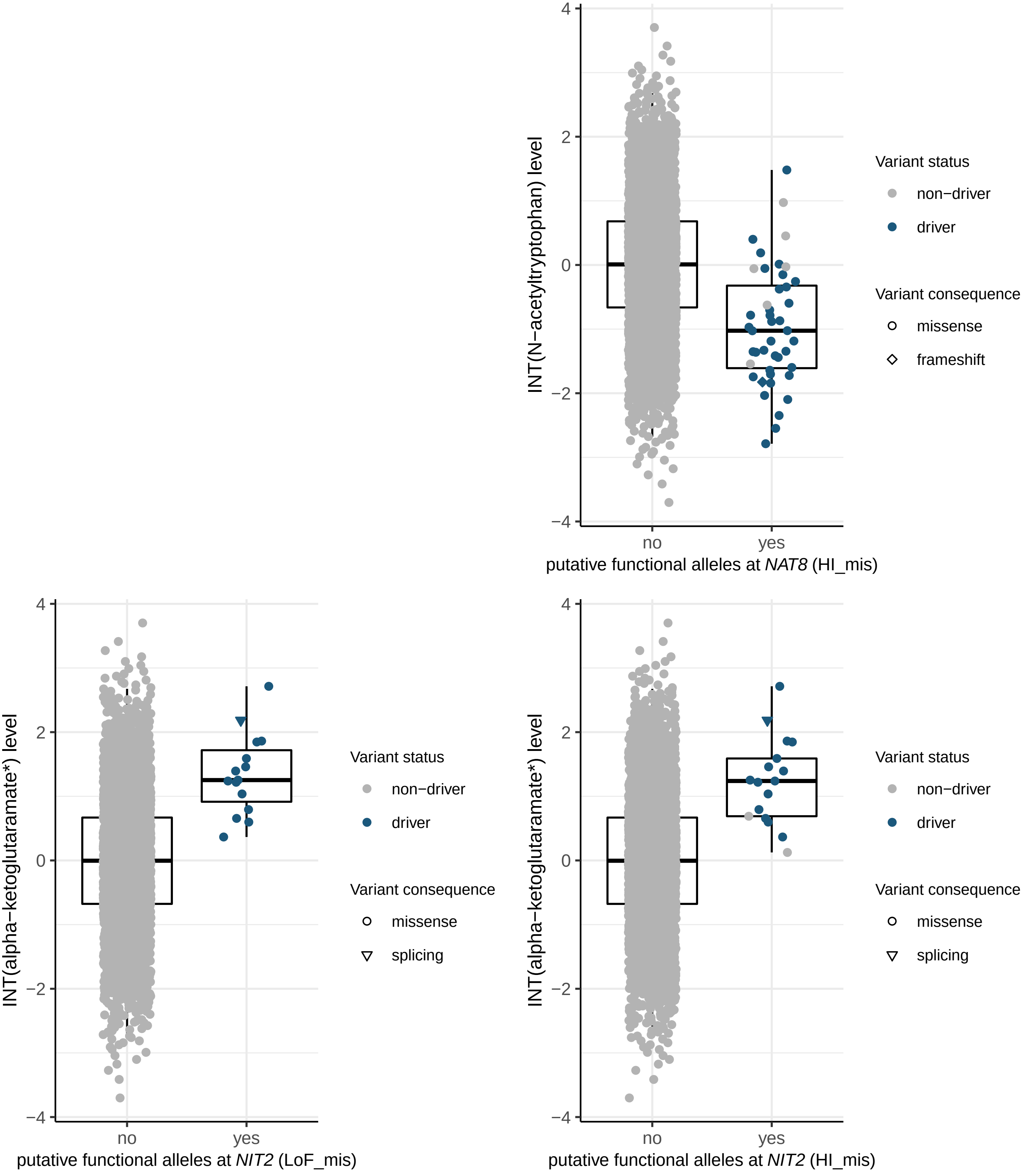

INT(N−acetyltryptophan) level
putative functional alleles at NAT8 (HI_mis)
Variant status
non−driver
driver
Variant consequence
missense
frameshift
INT(alpha−ketoglutaramate*) level
putative functional alleles at NIT2 (LoF_mis)
Variant status
non−driver
driver
Variant consequence
missense
splicing
INT(alpha−ketoglutaramate*) level
putative functional alleles at NIT2 (HI_mis)
Variant status
non−driver
driver
Variant consequence
missense
splicing
no
yes

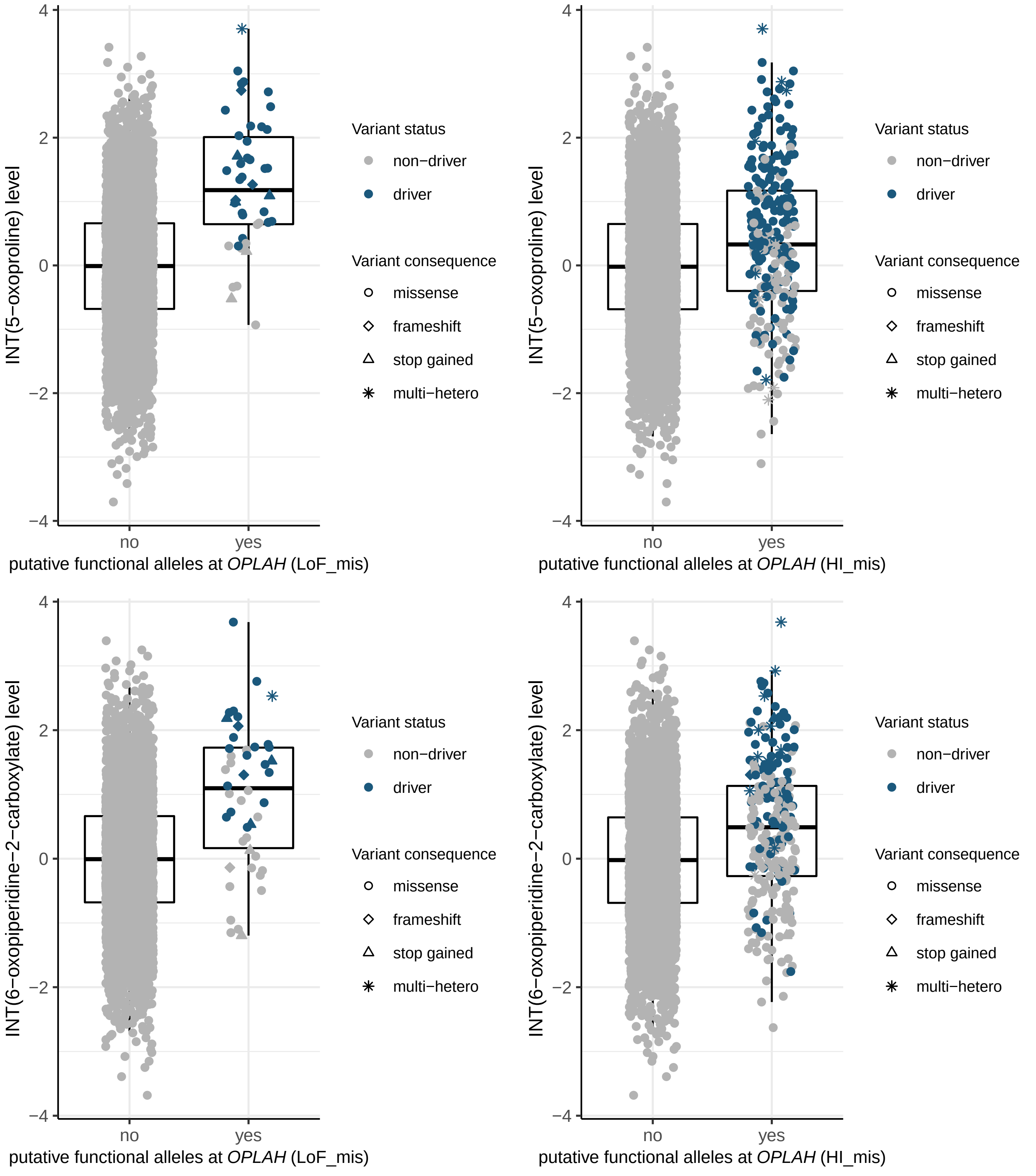

INT(5−oxoproline) level
putative functional alleles at OPLAH (LoF_mis)
INT(5−oxoproline) level
putative functional alleles at OPLAH (HI_mis)
INT(6−oxopiperidine−2−carboxylate) level
putative functional alleles at OPLAH (LoF_mis)
INT(6−oxopiperidine−2−carboxylate) level
putative functional alleles at OPLAH (HI_mis)
no
yes
Variant status
non−driver
driver
Variant consequence
missense
frameshift
stop gained
multi−hetero

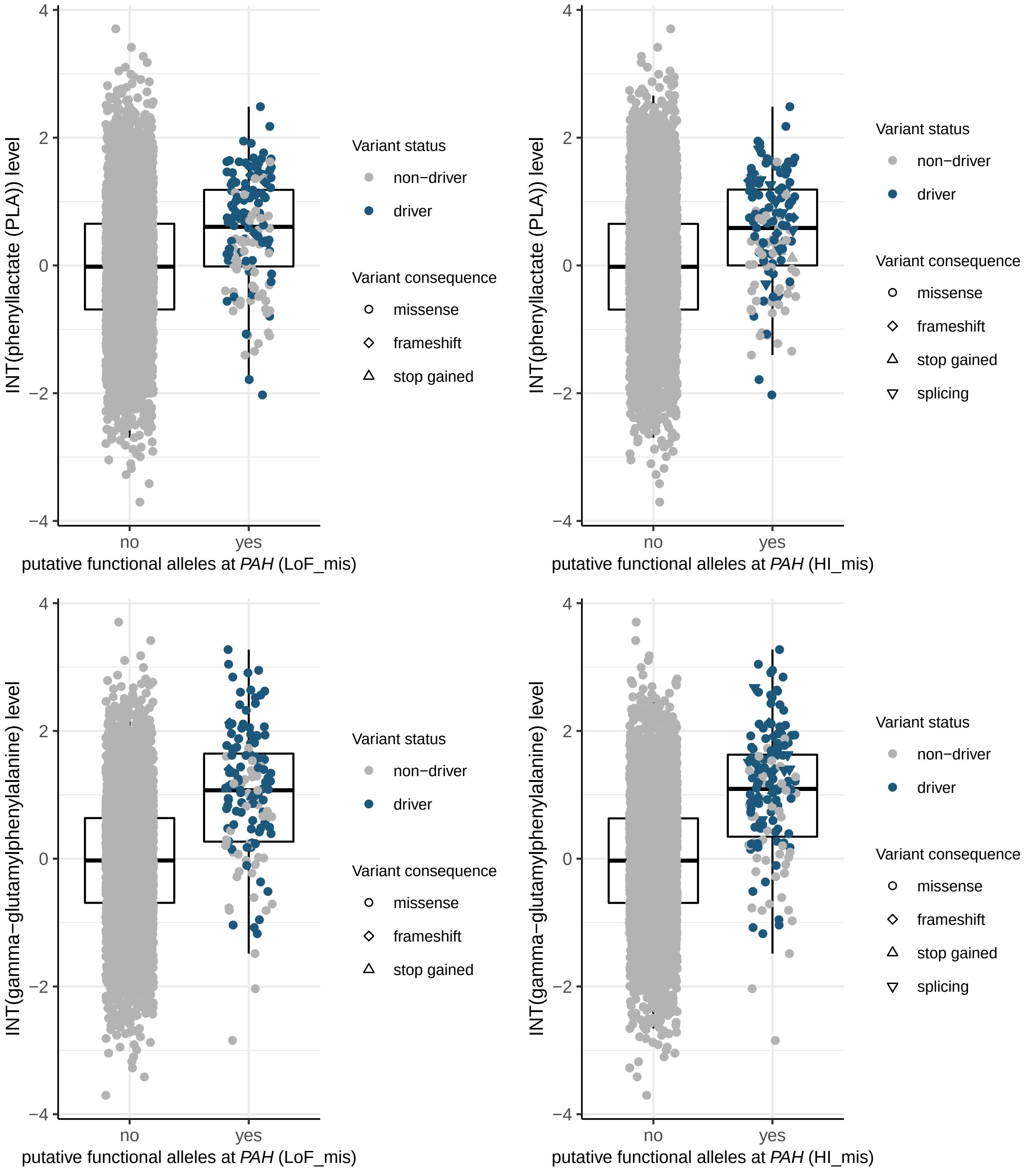

INT(phenyllactate (PLA)) level
4
2
0
−2
−4
no
yes
putative functional alleles at PAH (LoF_mis)
Variant status
non−driver
driver
Variant consequence
missense
frameshift
stop gained
INT(phenyllactate (PLA)) level
putative functional alleles at PAH (HI_mis)
Variant status
non−driver
driver
Variant consequence
missense
frameshift
stop gained
splicing
INT(gamma−glutamylphenylalanine) level
putative functional alleles at PAH (LoF_mis)
Variant status
non−driver
driver
Variant consequence
missense
frameshift
stop gained
INT(gamma−glutamylphenylalanine) level
putative functional alleles at PAH (HI_mis)
Variant status
non−driver
driver
Variant consequence
missense
frameshift
stop gained
splicing

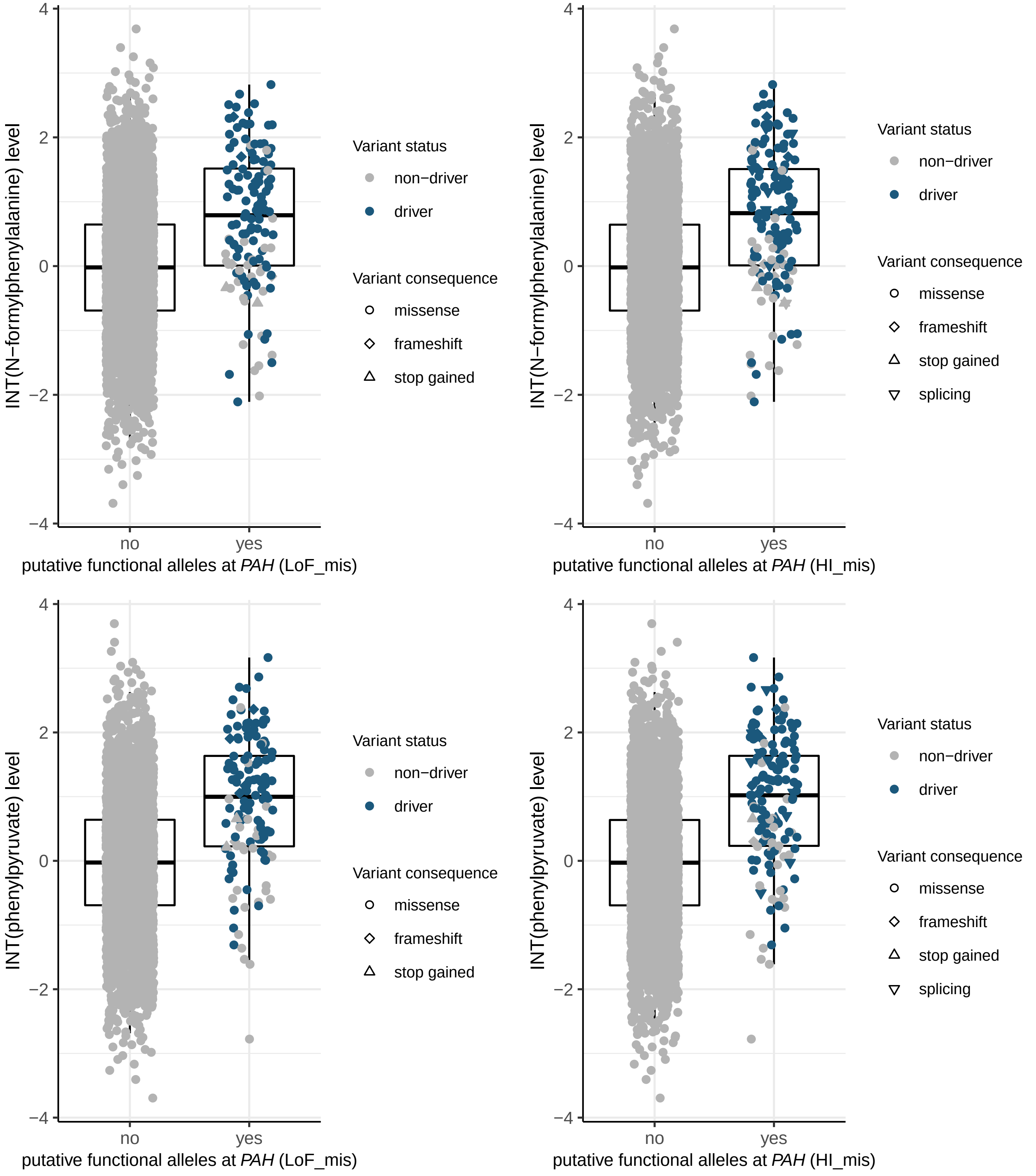

INT(N−formylphenylalanine) level
putative functional alleles at PAH (LoF_mis)
putative functional alleles at PAH (HI_mis)
INT(phenylpyruvate) level
no
yes
4
2
0
−2
−4
Variant status
non−driver
driver
Variant consequence
missense
frameshift
stop gained
splicing

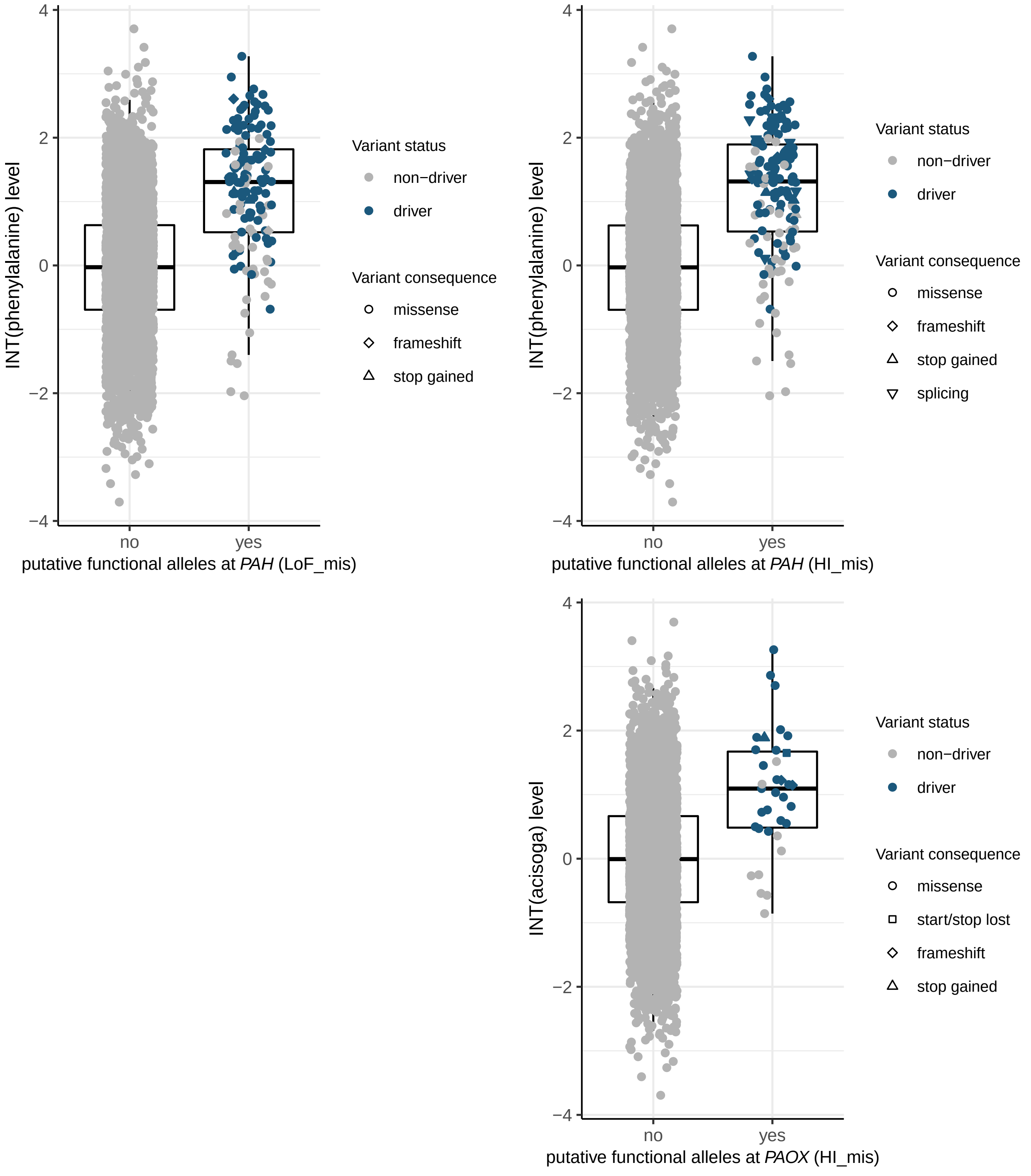

4
2
0
−2
−4
INT(phenylalanine) level
no
yes
putative functional alleles at *PAH* (LoF_mis)
Variant status
non−driver
driver
Variant consequence
missense
frameshift
stop gained
INT(phenylalanine) level
putative functional alleles at *PAH* (HI_mis)
Variant status
non−driver
driver
Variant consequence
missense
frameshift
stop gained
splicing
INT(acisoga) level
putative functional alleles at *PAOX* (HI_mis)
Variant status
non−driver
driver
Variant consequence
missense
start/stop lost
frameshift
stop gained

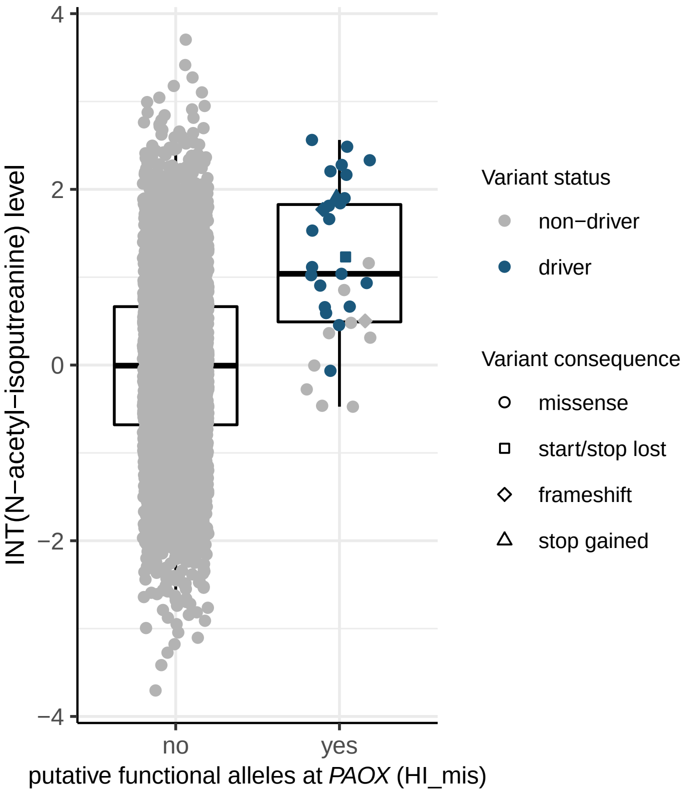

INT(N−acetyl−isoputreanine) level
putative functional alleles at *PAOX* (HI_mis)
no
yes
Variant status
non−driver
driver
Variant consequence
missense
start/stop lost
frameshift
stop gained


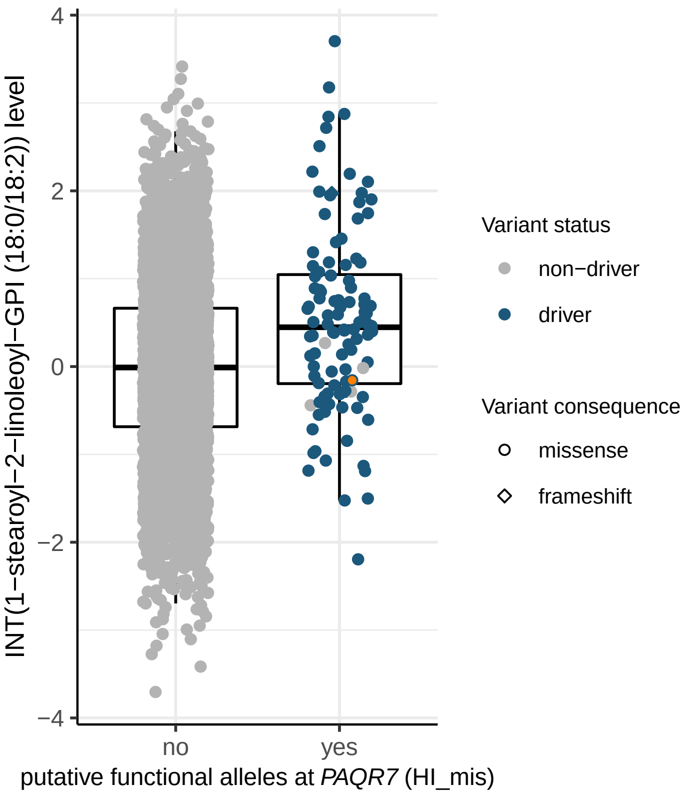

INT(1−stearoyl−2−linoleoyl−GPI (18:0/18:2)) level
putative functional alleles at *PAQR7* (HI_mis)
no
yes
Variant status
non−driver
driver
Variant consequence
missense
frameshift

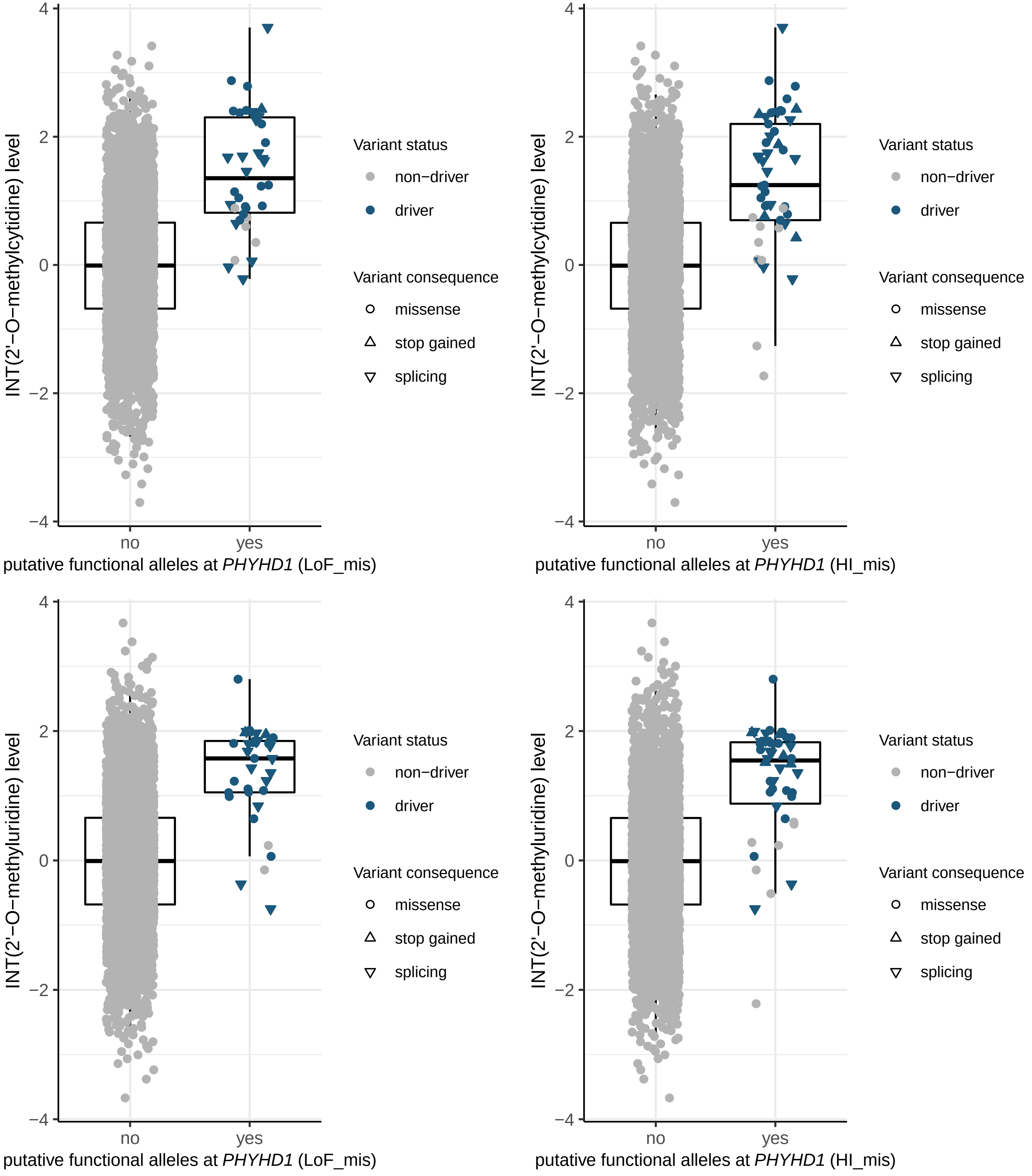

INT(2'−O−methylcytidine) level
putative functional alleles at *PHYHD1* (LoF_mis)
putative functional alleles at *PHYHD1* (HI_mis)
INT(2'−O−methyluridine) level
no
yes
Variant status
non−driver
driver
Variant consequence
missense
stop gained
splicing

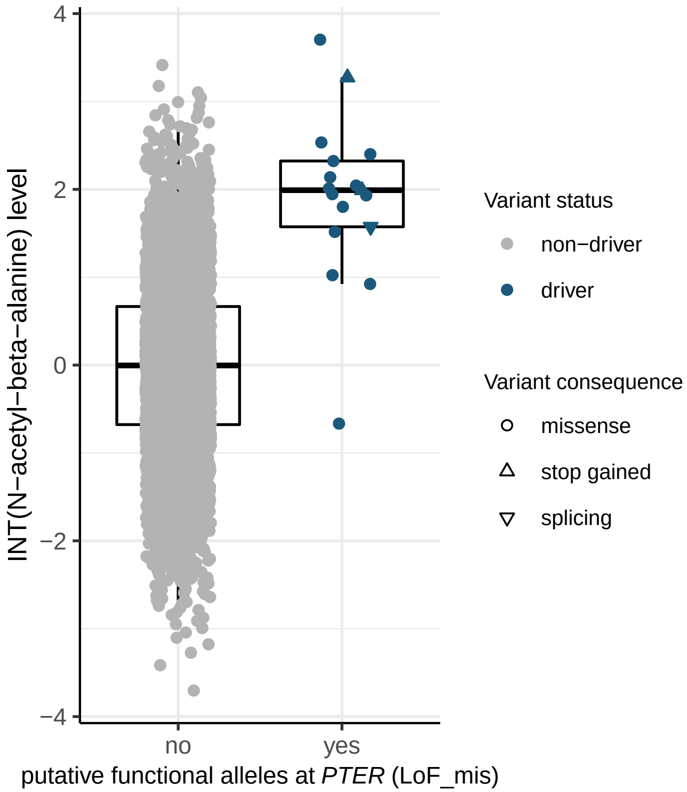

INT(N−acetyl−beta−alanine) level
4
2
0
−2
−4
no
yes
putative functional alleles at PTER (LoF_mis)
Variant status
non−driver
driver
Variant consequence
missense
stop gained
splicing


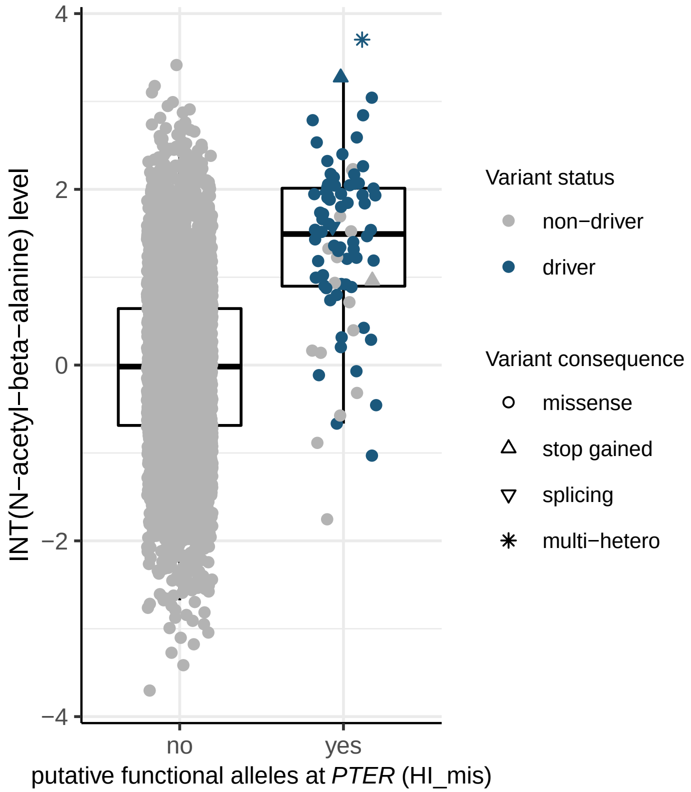

INT(N−acetyl−beta−alanine) level
4
2
0
−2
−4
no
yes
putative functional alleles at PTER (HI_mis)
Variant status
non−driver
driver
Variant consequence
missense
stop gained
splicing
multi−hetero


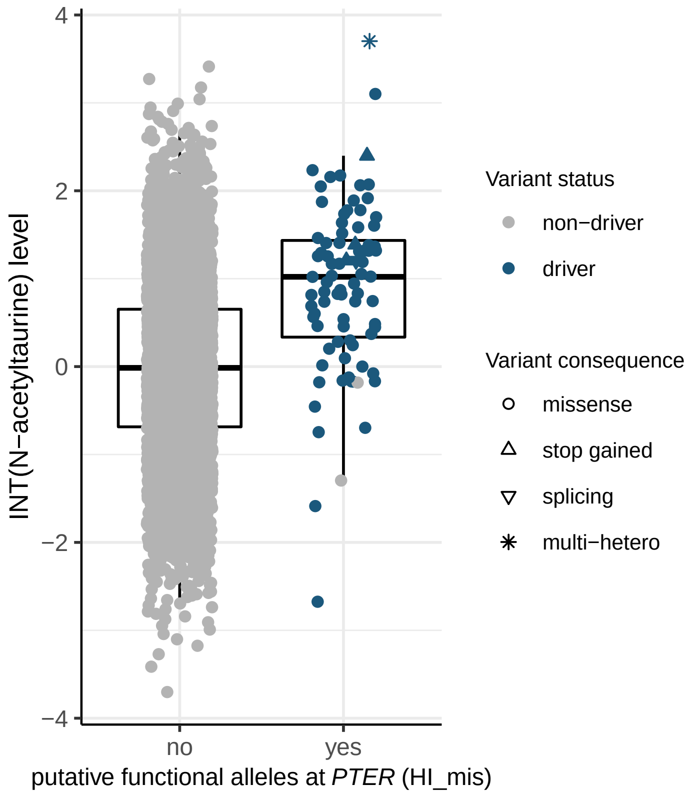

INT(N−acetyltaurine) level
4
2
0
−2
−4
no
yes
putative functional alleles at PTER (HI_mis)
Variant status
non−driver
driver
Variant consequence
missense
stop gained
splicing
multi−hetero

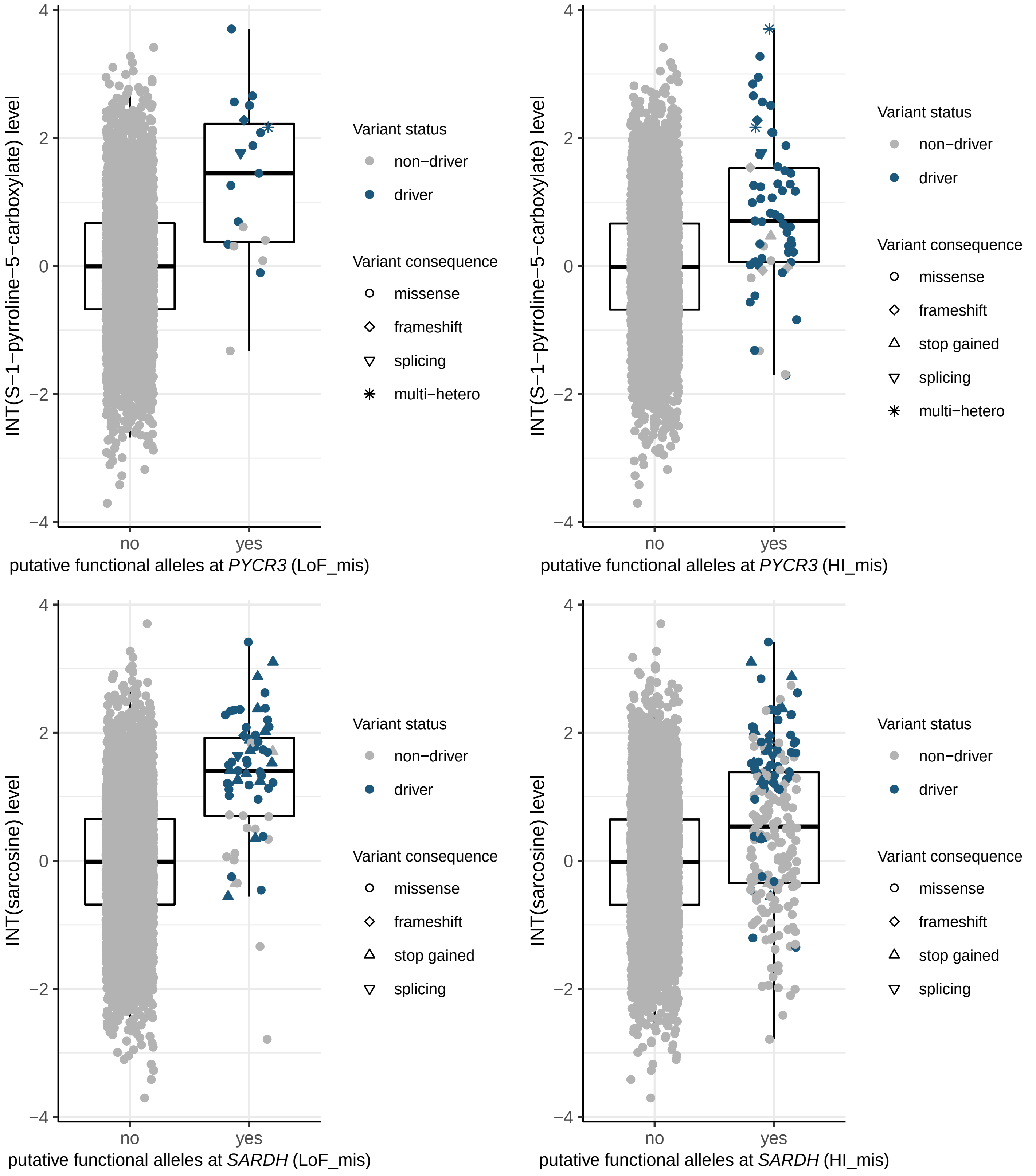
INT(S−1−pyrroline−5−carboxylate) level
4
2
0
−2
−4
no
yes
putative functional alleles at PYCR3 (LoF_mis)
Variant status
non−driver
driver
Variant consequence
missense
frameshift
splicing
multi−hetero
INT(S−1−pyrroline−5−carboxylate) level
putative functional alleles at PYCR3 (HI_mis)
Variant status
non−driver
driver
Variant consequence
missense
frameshift
stop gained
splicing
multi−hetero
INT(sarcosine) level
putative functional alleles at SARDH (LoF_mis)
Variant status
non−driver
driver
Variant consequence
missense
frameshift
stop gained
splicing
INT(sarcosine) level
putative functional alleles at SARDH (HI_mis)
Variant status
non−driver
driver
Variant consequence
missense
frameshift
stop gained
splicing

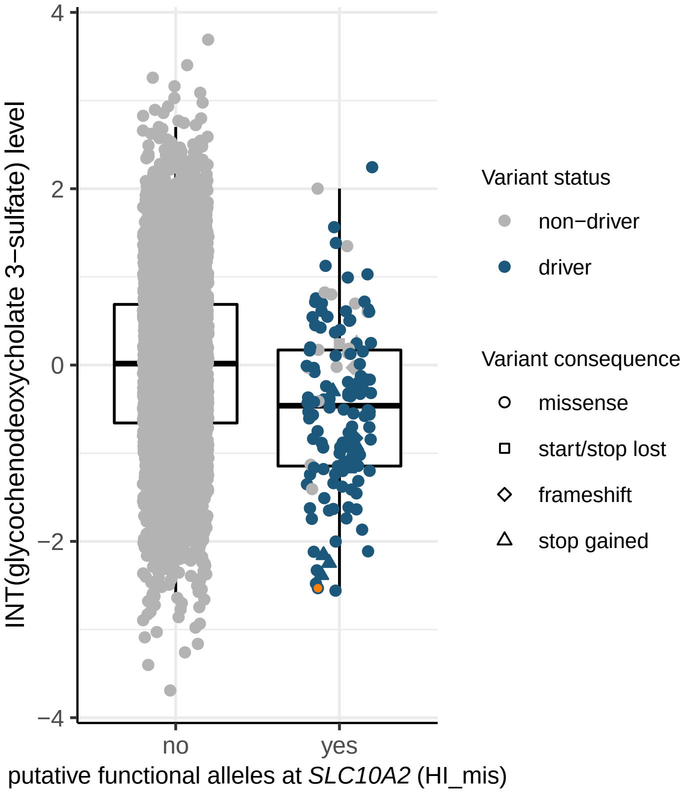

4
2
0
−2
−4
INT(glycochenodeoxycholate 3−sulfate) level
no
yes
putative functional alleles at SLC10A2 (HI_mis)
Variant status
non−driver
driver
Variant consequence
missense
start/stop lost
frameshift
stop gained


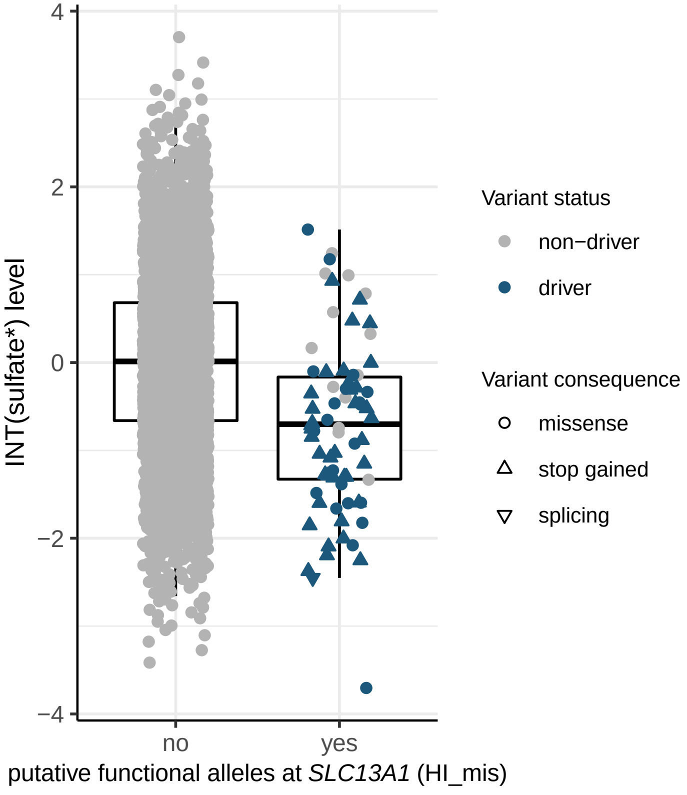

4
2
0
−2
−4
INT(sulfate*) level
no
yes
putative functional alleles at SLC13A1 (HI_mis)
Variant status
non−driver
driver
Variant consequence
missense
stop gained
splicing

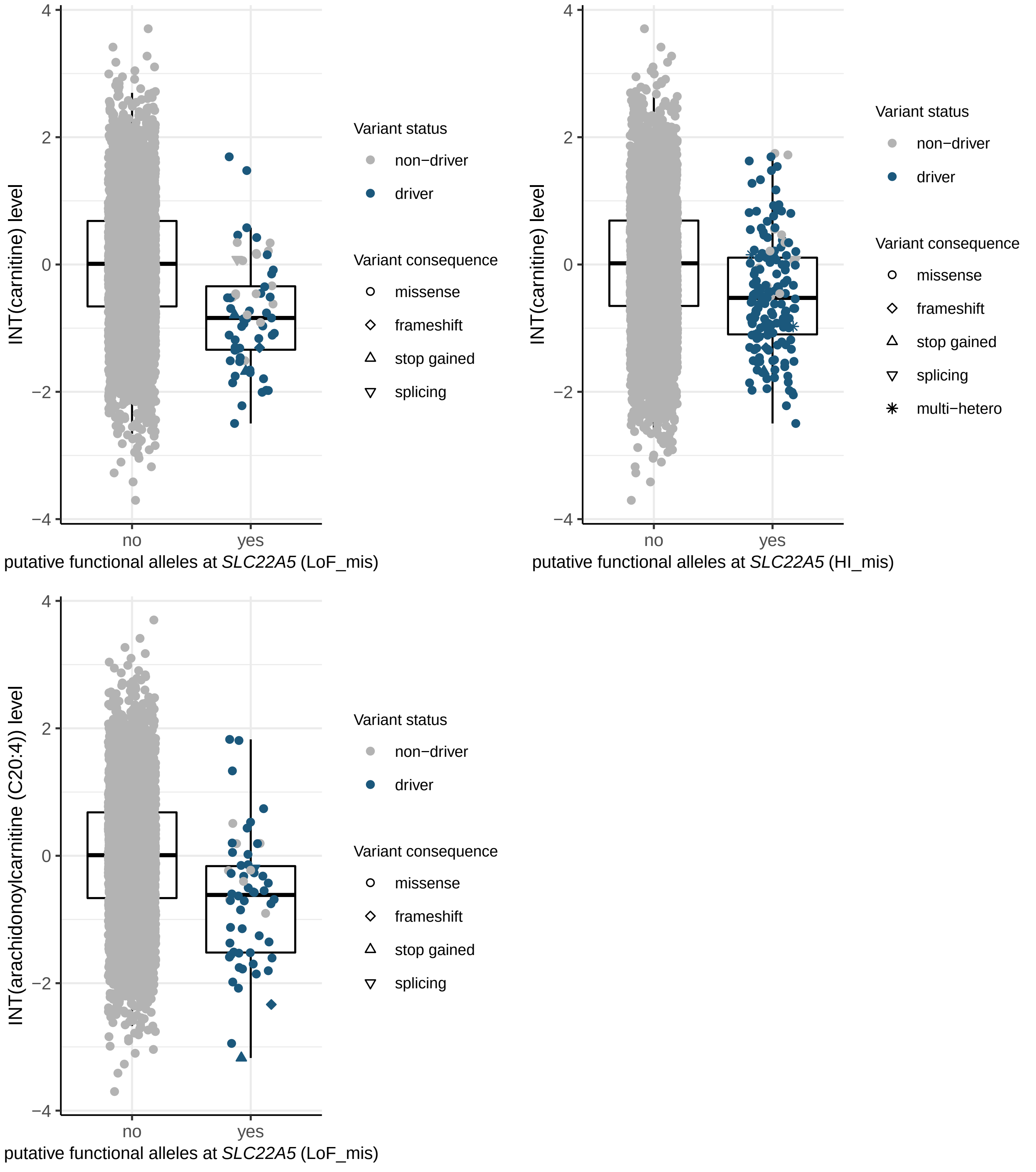

INT(carnitine) level
putative functional alleles at SLC22A5 (LoF_mis)
Variant status
non−driver
driver
Variant consequence
missense
frameshift
stop gained
splicing
INT(carnitine) level
putative functional alleles at SLC22A5 (HI_mis)
Variant status
non−driver
driver
Variant consequence
missense
frameshift
stop gained
splicing
multi−hetero
INT(arachidonoylcarnitine (C20:4)) level
putative functional alleles at SLC22A5 (LoF_mis)
Variant status
non−driver
driver
Variant consequence
missense
frameshift
stop gained
splicing

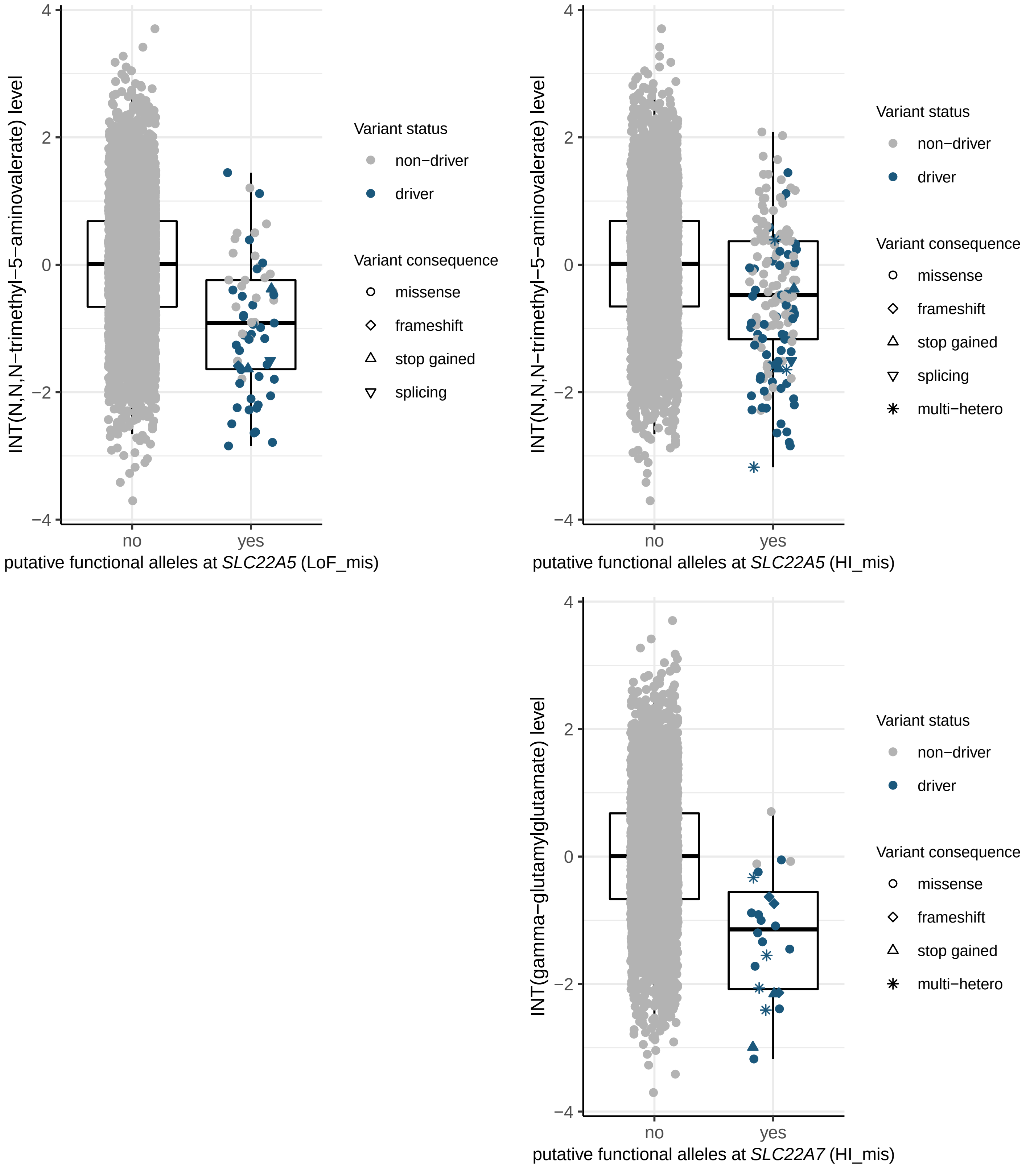
INT(N,N,N−trimethyl−5−aminovalerate) level
4
2
0
−2
−4
no
yes
putative functional alleles at *SLC22A5* (LoF_mis)
Variant status
non−driver
driver
Variant consequence
missense
frameshift
stop gained
splicing
INT(N,N,N−trimethyl−5−aminovalerate) level
4
2
0
−2
−4
no
yes
putative functional alleles at *SLC22A5* (HI_mis)
Variant status
non−driver
driver
Variant consequence
missense
frameshift
stop gained
splicing
multi−hetero
INT(gamma−glutamylglutamate) level
4
2
0
−2
−4
no
yes
putative functional alleles at *SLC22A7* (HI_mis)
Variant status
non−driver
driver
Variant consequence
missense
frameshift
stop gained
multi−hetero

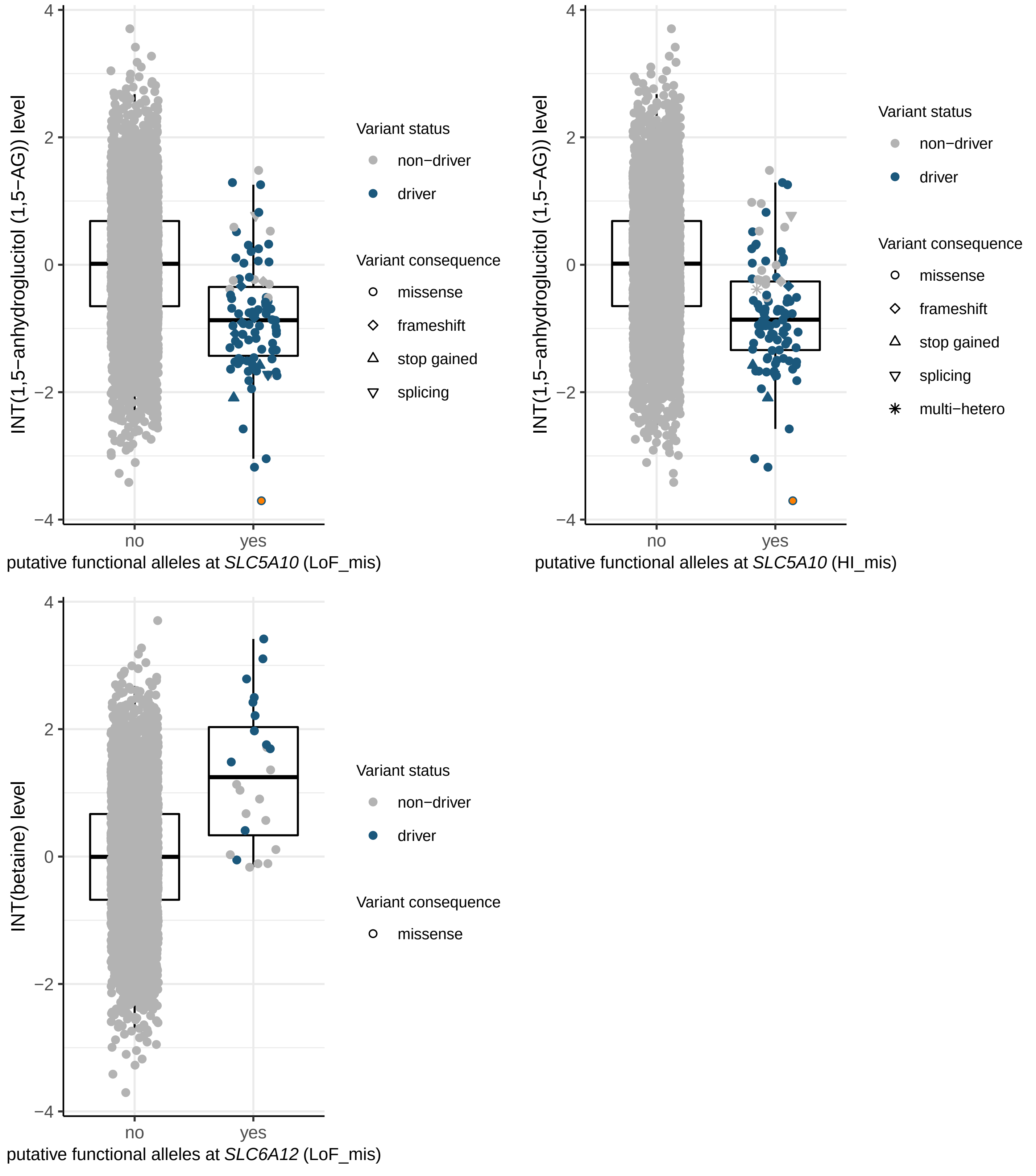

INT(1,5−anhydroglucitol (1,5−AG)) level
putative functional alleles at SLC5A10 (LoF_mis)
Variant status
non−driver
driver
Variant consequence
missense
frameshift
stop gained
splicing
INT(1,5−anhydroglucitol (1,5−AG)) level
putative functional alleles at SLC5A10 (HI_mis)
Variant status
non−driver
driver
Variant consequence
missense
frameshift
stop gained
splicing
multi−hetero
INT(betaine) level
putative functional alleles at SLC6A12 (LoF_mis)
Variant status
non−driver
driver
Variant consequence
missense
no
yes
4
2
0
−2
−4

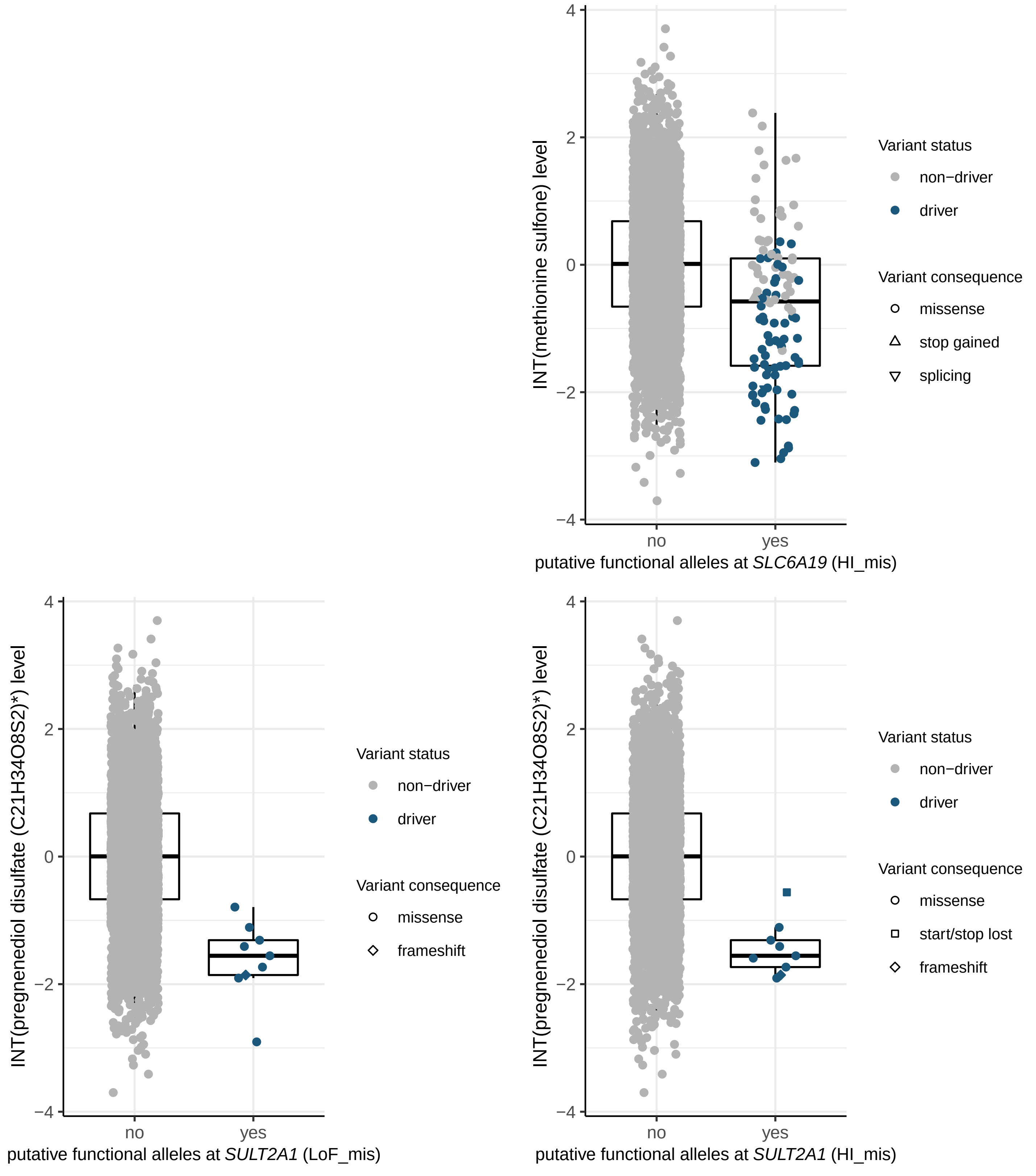

INT(methionine sulfone) level
4
2
0
−2
−4
no
yes
putative functional alleles at *SLC6A19* (HI_mis)
Variant status
non−driver
driver
Variant consequence
missense
stop gained
splicing
INT(pregnenediol disulfate (C21H34O8S2)*) level
4
2
0
−2
−4
no
yes
putative functional alleles at *SULT2A1* (LoF_mis)
Variant status
non−driver
driver
Variant consequence
missense
frameshift
INT(pregnenediol disulfate (C21H34O8S2)*) level
4
2
0
−2
−4
no
yes
putative functional alleles at *SULT2A1* (HI_mis)
Variant status
non−driver
driver
Variant consequence
missense
start/stop lost
frameshift

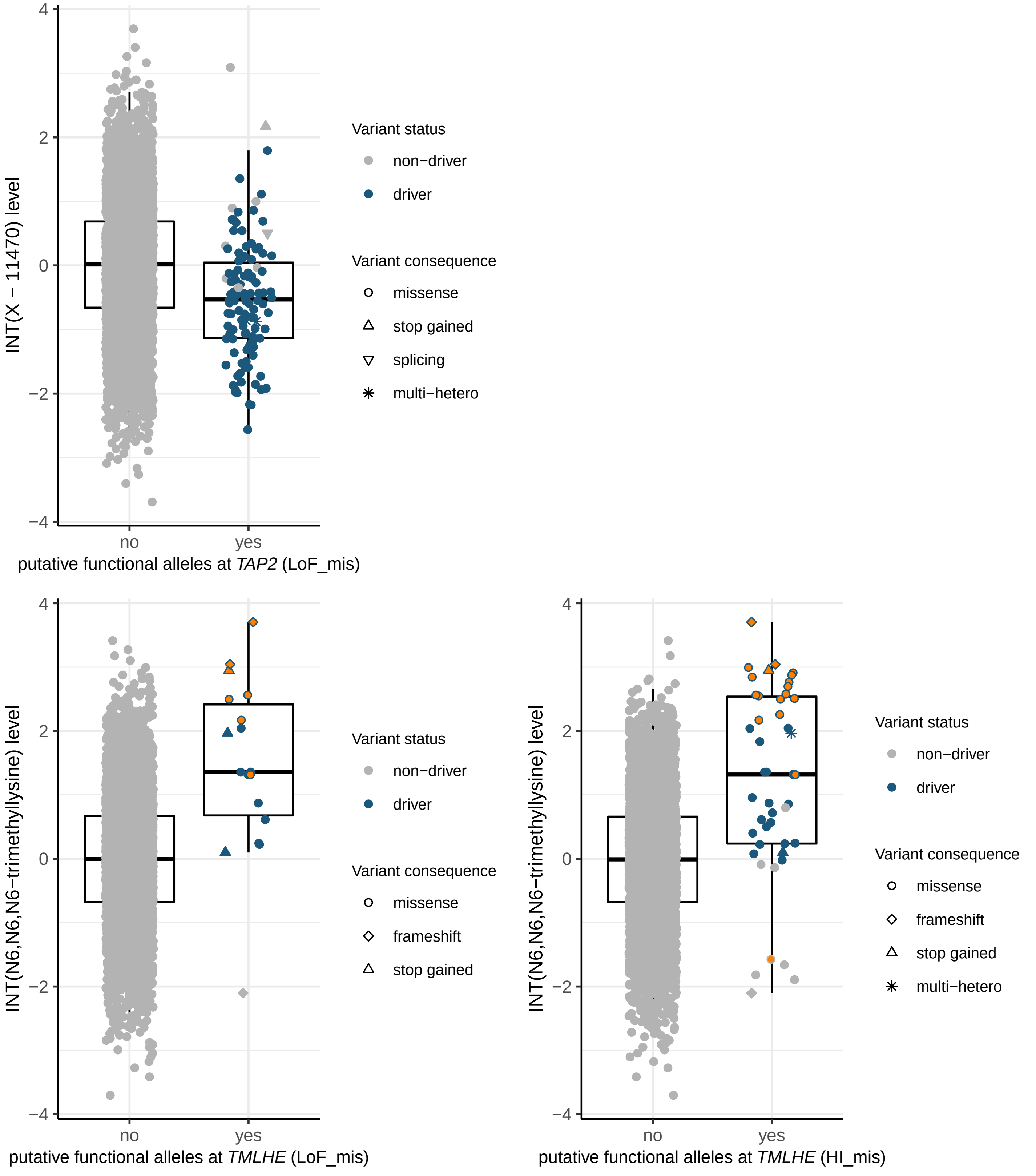

INT(X − 11470) level
4
2
0
−2
−4
no
yes
putative functional alleles at *TAP2* (LoF_mis)
Variant status
non−driver
driver
Variant consequence
missense
stop gained
splicing
multi−hetero
INT(N6,N6,N6−trimethyllysine) level
putative functional alleles at *TMLHE* (LoF_mis)
Variant status
non−driver
driver
Variant consequence
missense
frameshift
stop gained
INT(N6,N6,N6−trimethyllysine) level
putative functional alleles at *TMLHE* (HI_mis)
Variant status
non−driver
driver
Variant consequence
missense
frameshift
stop gained
multi−hetero

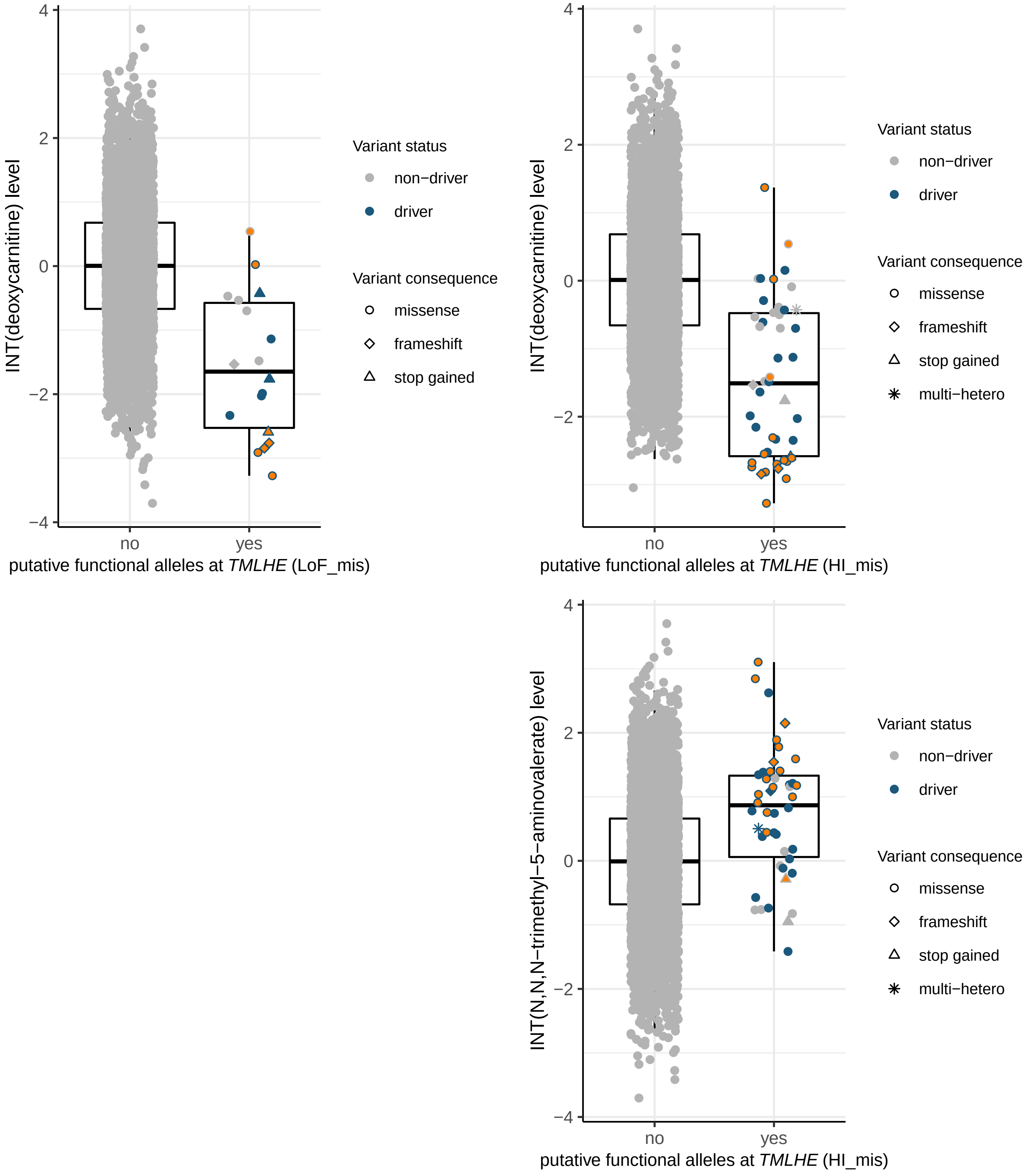
INT(deoxycarnitine) level
4
2
0
−2
−4
no
yes
putative functional alleles at TMLHE (LoF_mis)
Variant status
non−driver
driver
Variant consequence
missense
frameshift
stop gained
INT(deoxycarnitine) level
putative functional alleles at TMLHE (HI_mis)
multi−hetero
INT(N,N,N−trimethyl−5−aminovalerate) level
putative functional alleles at TMLHE (HI_mis)

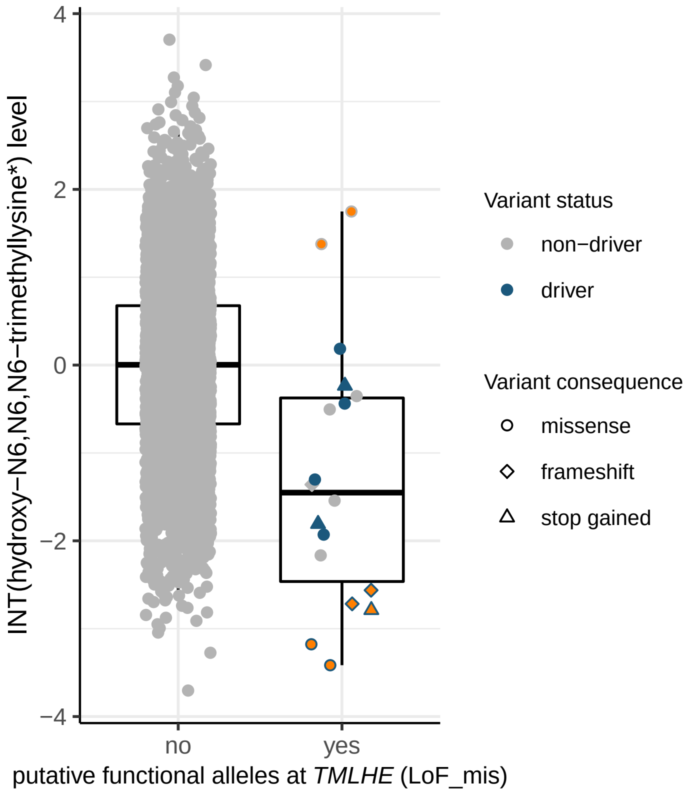

INT(hydroxy−N6,N6,N6−trimethyllysine*) level
4
2
0
−2
−4
no
yes
putative functional alleles at TMLHE (LoF_mis)
Variant status
non−driver
driver
Variant consequence
missense
frameshift
stop gained


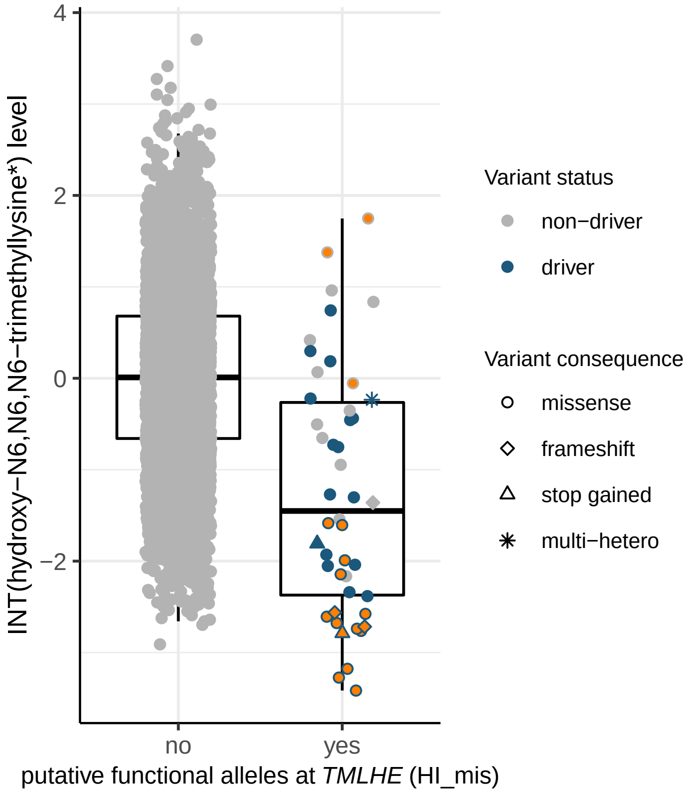

INT(hydroxy−N6,N6,N6−trimethyllysine*) level
4
2
0
−2
no
yes
putative functional alleles at TMLHE (HI_mis)
Variant status
non−driver
driver
Variant consequence
missense
frameshift
stop gained
multi−hetero


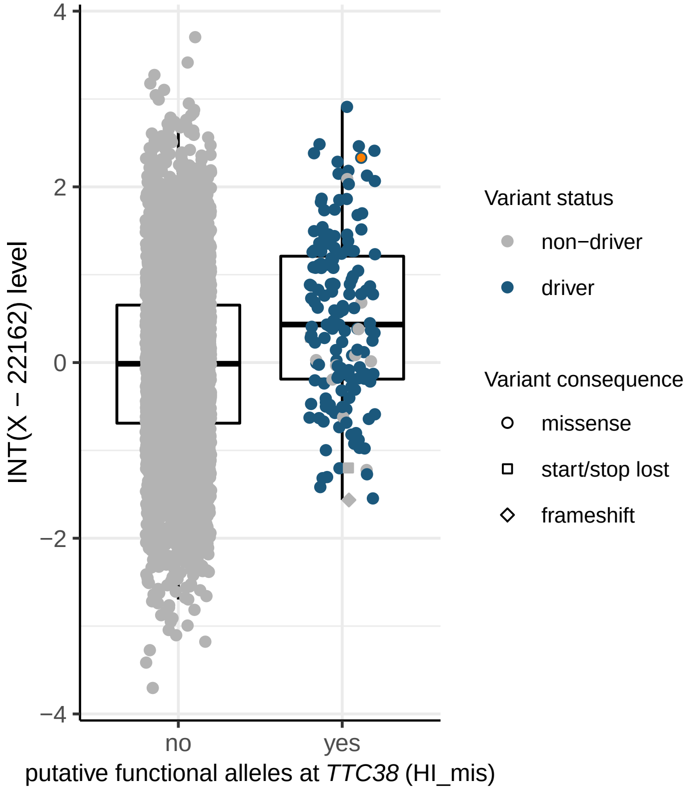

INT(X − 22162) level
4
2
0
−2
−4
no
yes
putative functional alleles at TTC38 (HI_mis)
Variant status
non−driver
driver
Variant consequence
missense
start/stop lost
frameshift

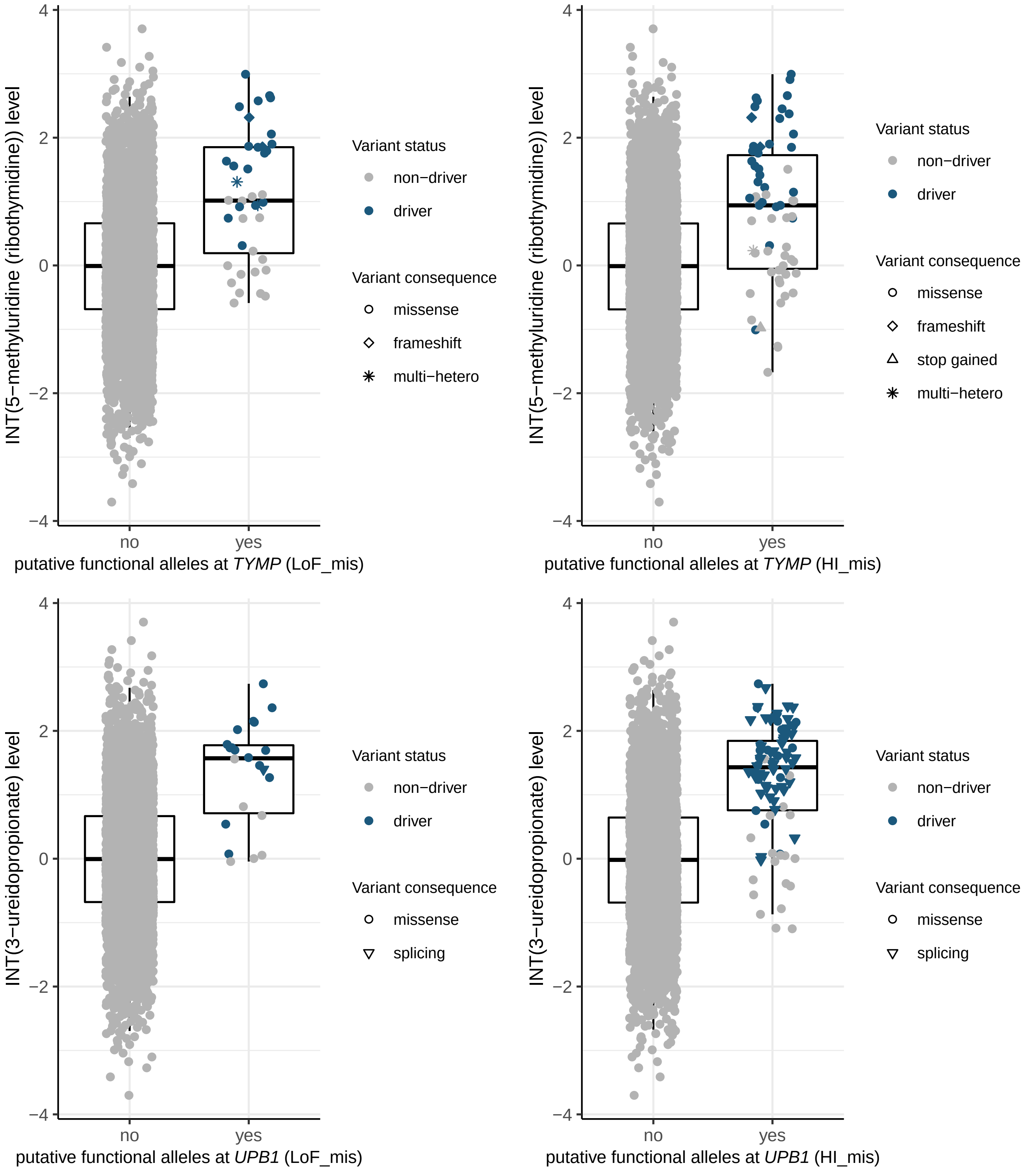

INT(5−methyluridine (ribothymidine)) level
4
2
0
−2
−4
no
yes
putative functional alleles at TYMP (LoF_mis)
Variant status
non−driver
driver
Variant consequence
missense
frameshift
multi−hetero
INT(5−methyluridine (ribothymidine)) level
putative functional alleles at TYMP (HI_mis)
Variant status
non−driver
driver
Variant consequence
missense
frameshift
stop gained
multi−hetero
INT(3−ureidopropionate) level
putative functional alleles at UPB1 (LoF_mis)
Variant status
non−driver
driver
Variant consequence
missense
splicing
INT(3−ureidopropionate) level
putative functional alleles at UPB1 (HI_mis)
Variant status
non−driver
driver
Variant consequence
missense
splicing

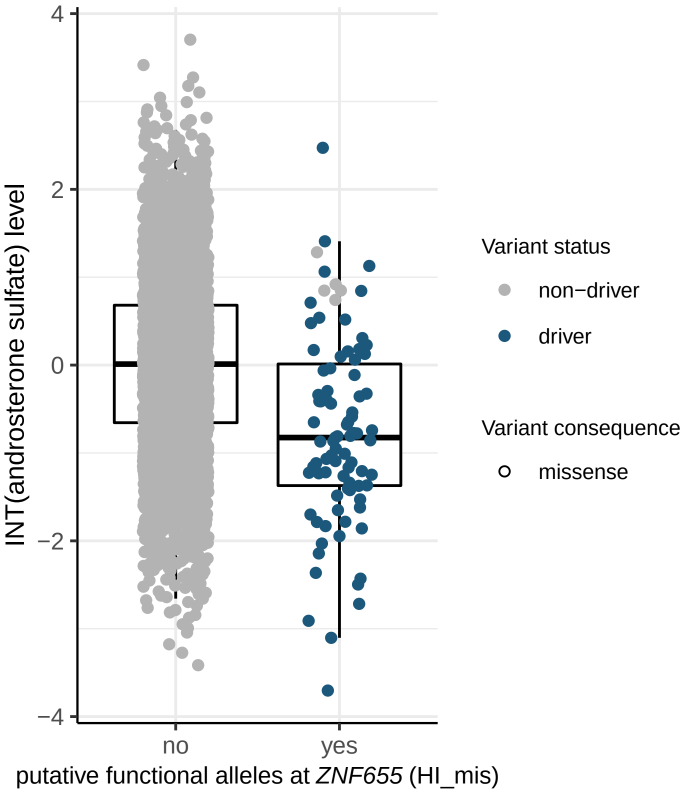

INT(androsterone sulfate) level
4
2
0
−2
−4
no
yes
putative functional alleles at *ZNF655* (HI_mis)
Variant status
non−driver
driver
Variant consequence
missense


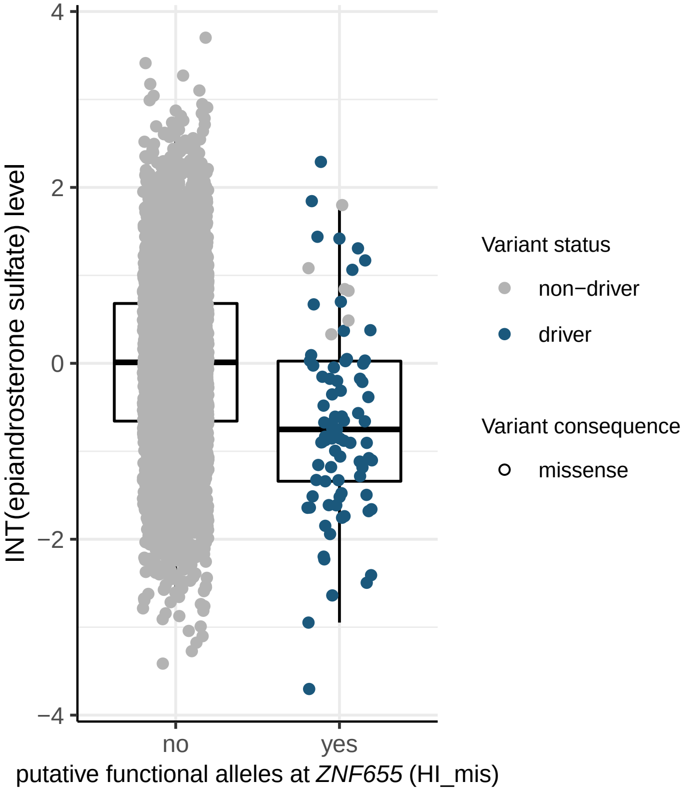

INT(epiandrosterone sulfate) level
4
2
0
−2
−4
no
yes
putative functional alleles at *ZNF655* (HI_mis)
Variant status
non−driver
driver
Variant consequence
missense
