## Supplementary_Figure2 for "Coupling of metabolomics and exome sequencing reveals graded effects of rare damaging heterozygous variants on gene function and resulting traits and diseases"

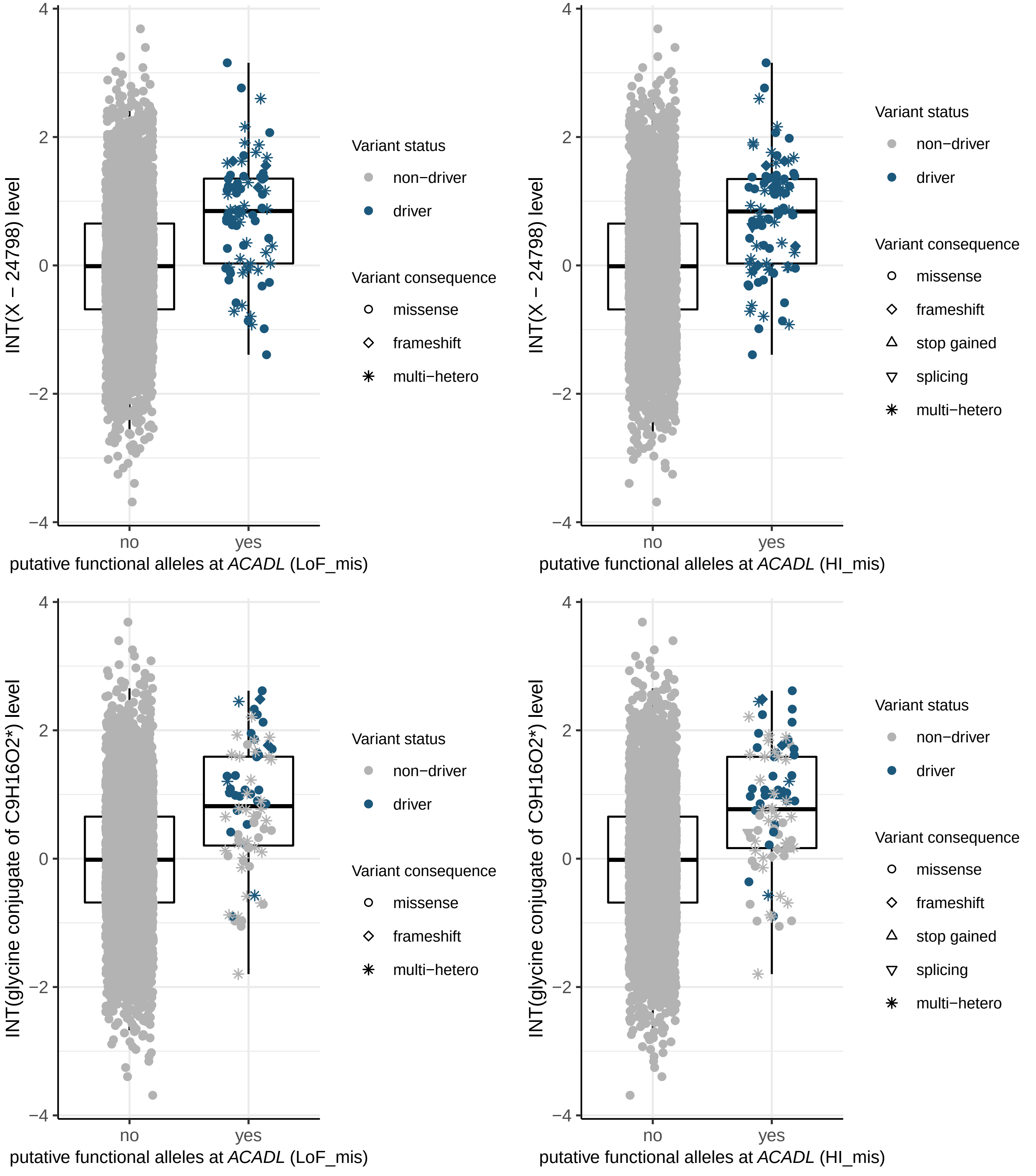
INT(X − 24798) level
putative functional alleles at ACADL (LoF_mis)
putative functional alleles at ACADL (HI_mis)
INT(glycine conjugate of C9H16O2*) level
no
yes
Variant status
non−driver
driver
Variant consequence
missense
frameshift
stop gained
splicing
multi−hetero

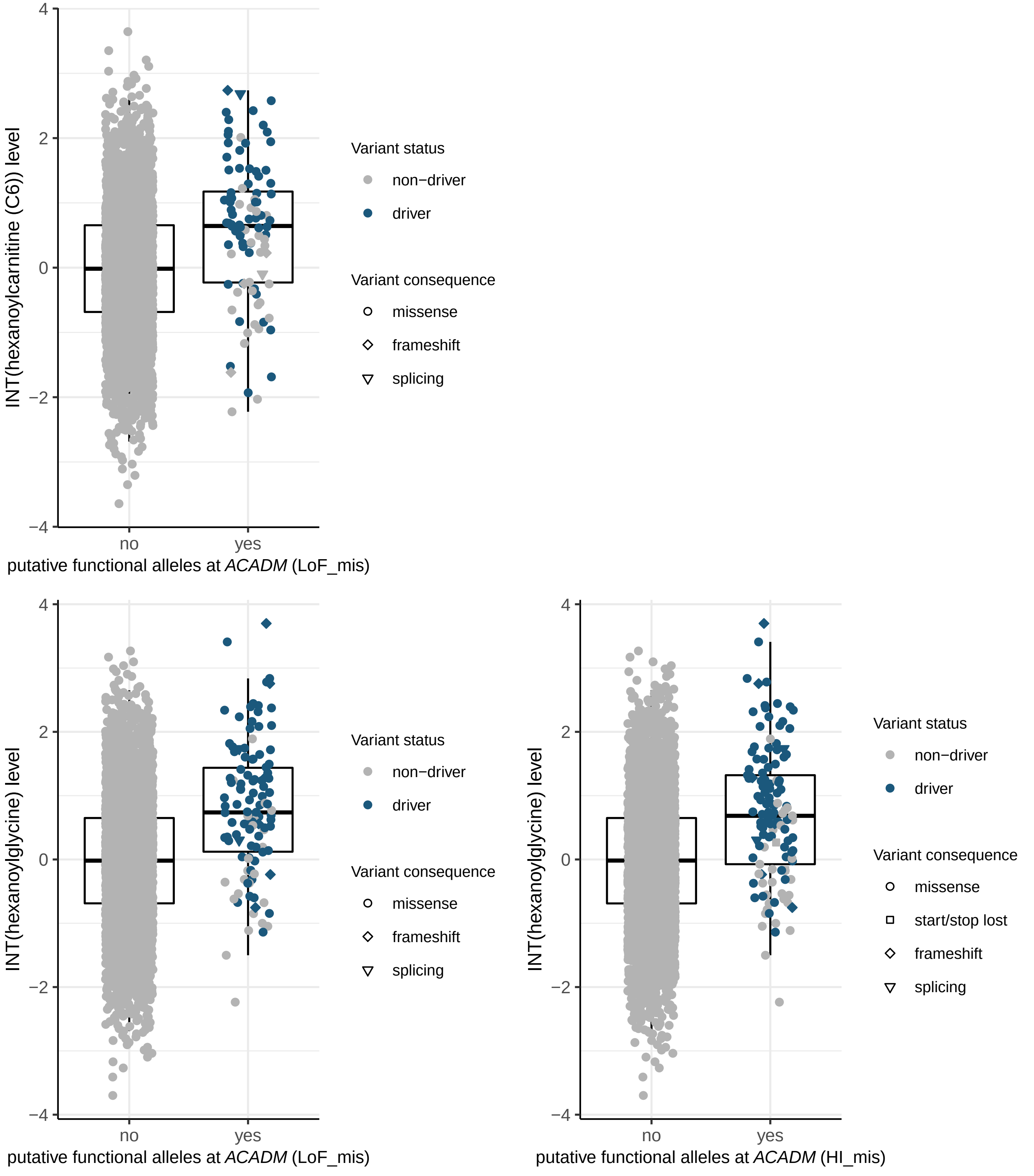

INT(hexanoylcarnitine (C6)) level
putative functional alleles at *ACADM* (LoF_mis)
no
yes
4
2
0
−2
−4
Variant status
non−driver
driver
Variant consequence
missense
frameshift
splicing
INT(hexanoylglycine) level
putative functional alleles at *ACADM* (LoF_mis)
Variant status
non−driver
driver
Variant consequence
missense
frameshift
splicing
INT(hexanoylglycine) level
putative functional alleles at *ACADM* (HI_mis)
Variant status
non−driver
driver
Variant consequence
missense
start/stop lost
frameshift
splicing

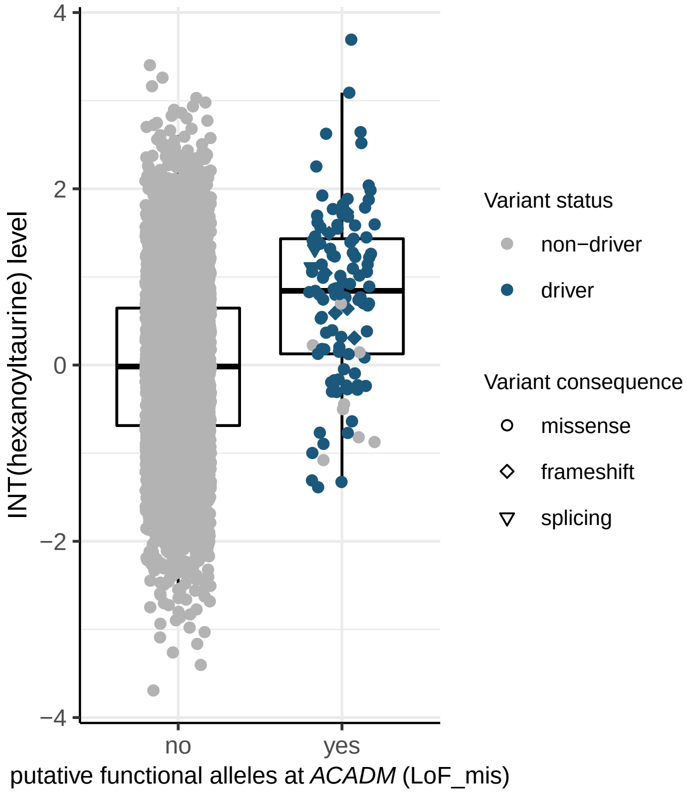
4
2
0
−2
−4
INT(hexanoyltaurine) level
no
yes
putative functional alleles at *ACADM* (LoF_mis)
Variant status
non−driver
driver
Variant consequence
missense
frameshift
splicing

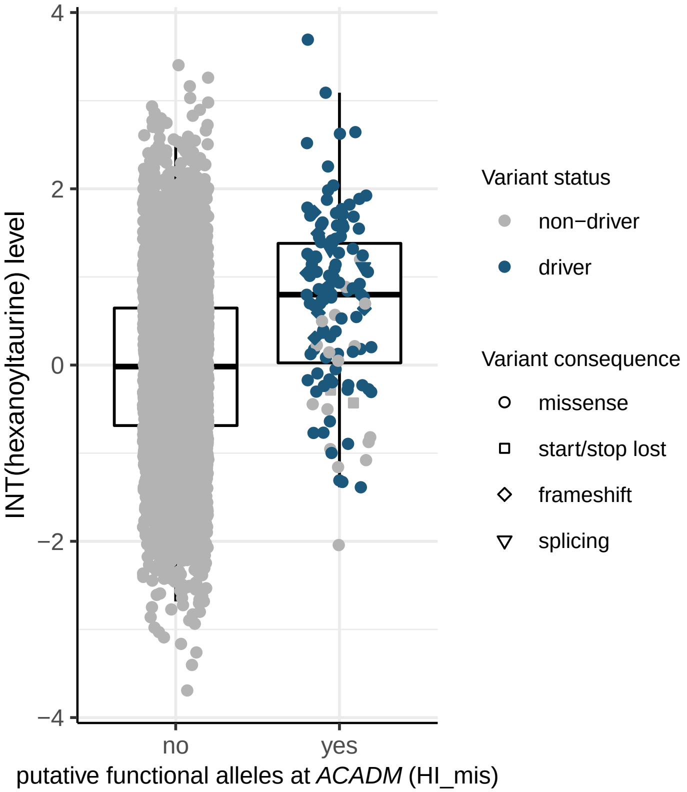
4
2
0
−2
−4
INT(hexanoyltaurine) level
no
yes
putative functional alleles at *ACADM* (HI_mis)
Variant status
non−driver
driver
Variant consequence
missense
start/stop lost
frameshift
splicing

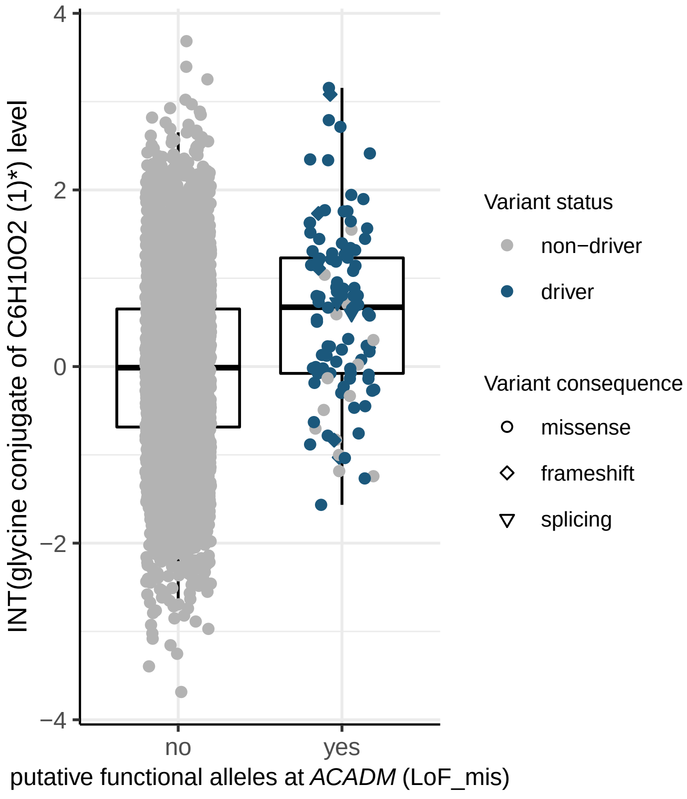
4
2
0
−2
−4
INT(glycine conjugate of C6H10O2 (1)*) level
no
yes
putative functional alleles at *ACADM* (LoF_mis)
Variant status
non−driver
driver
Variant consequence
missense
frameshift
splicing

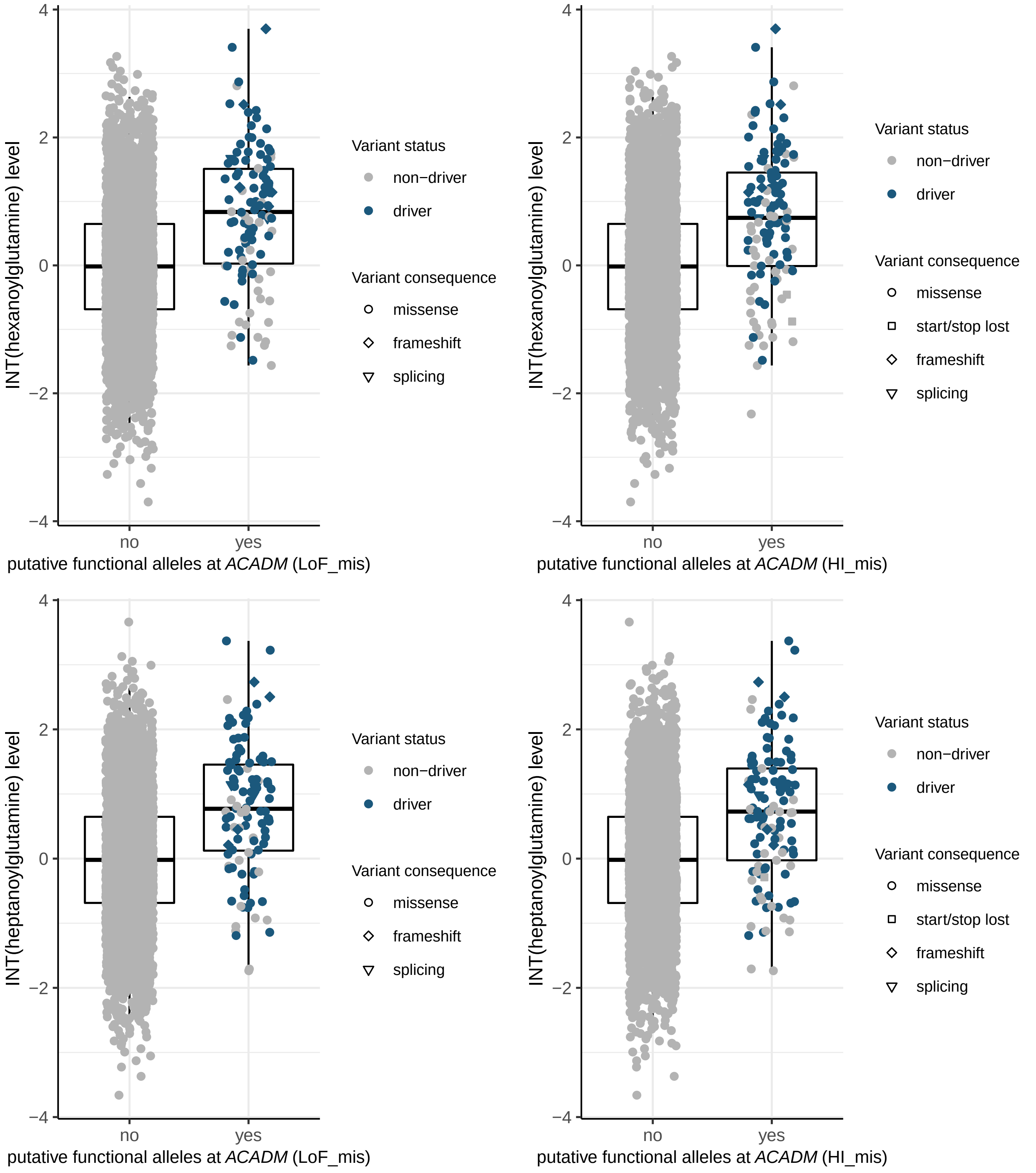
INT(hexanoylglutamine) level
putative functional alleles at ACADM (LoF_mis)
putative functional alleles at ACADM (HI_mis)
INT(heptanoylglutamine) level
no
yes
4
2
0
−2
−4
Variant status
non−driver
driver
Variant consequence
missense
start/stop lost
frameshift
splicing

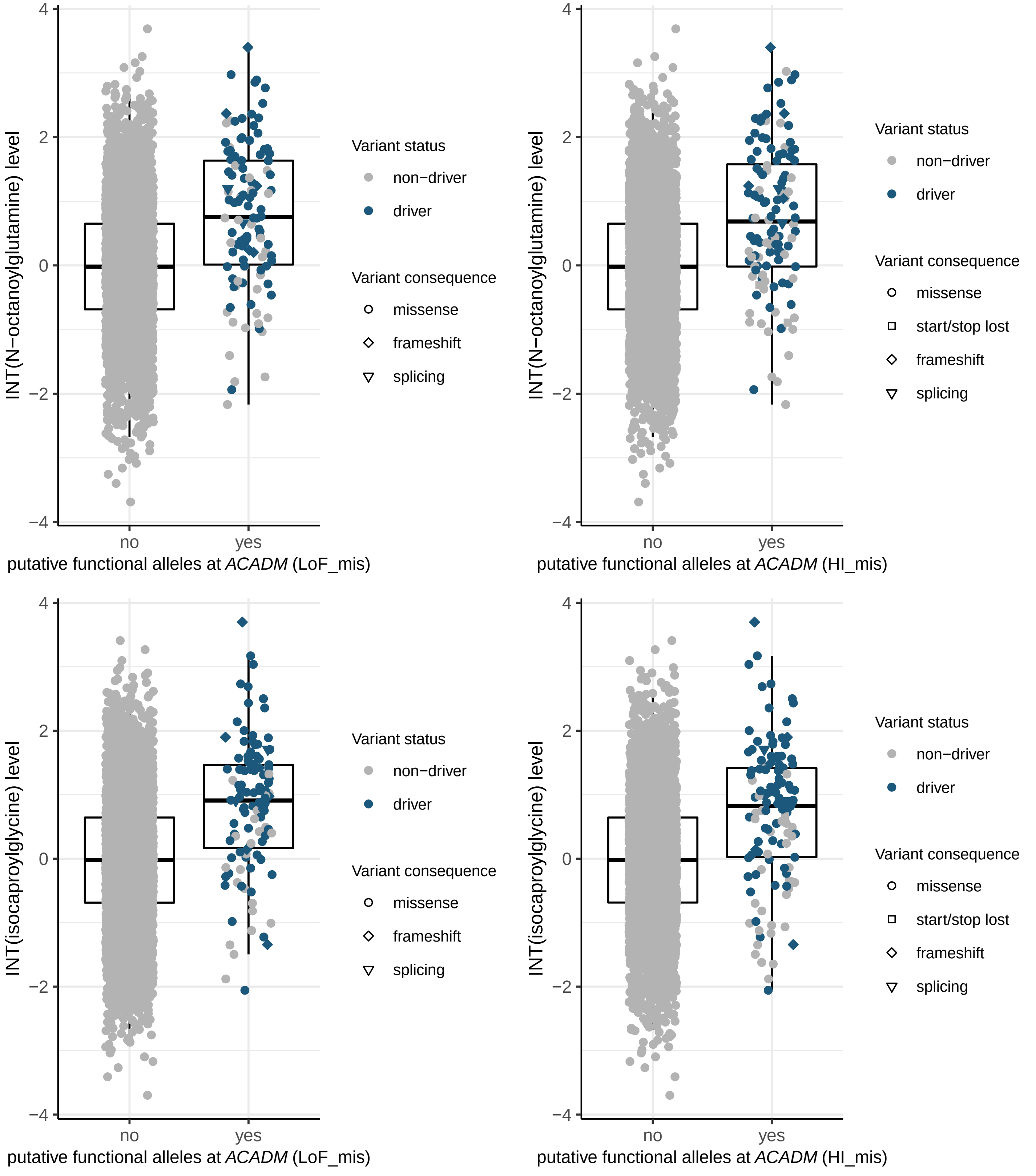
INT(N−octanoylglutamine) level
putative functional alleles at ACADM (LoF_mis)
putative functional alleles at ACADM (HI_mis)
INT(isocaproylglycine) level
no
yes
Variant status
non−driver
driver
Variant consequence
missense
start/stop lost
frameshift
splicing

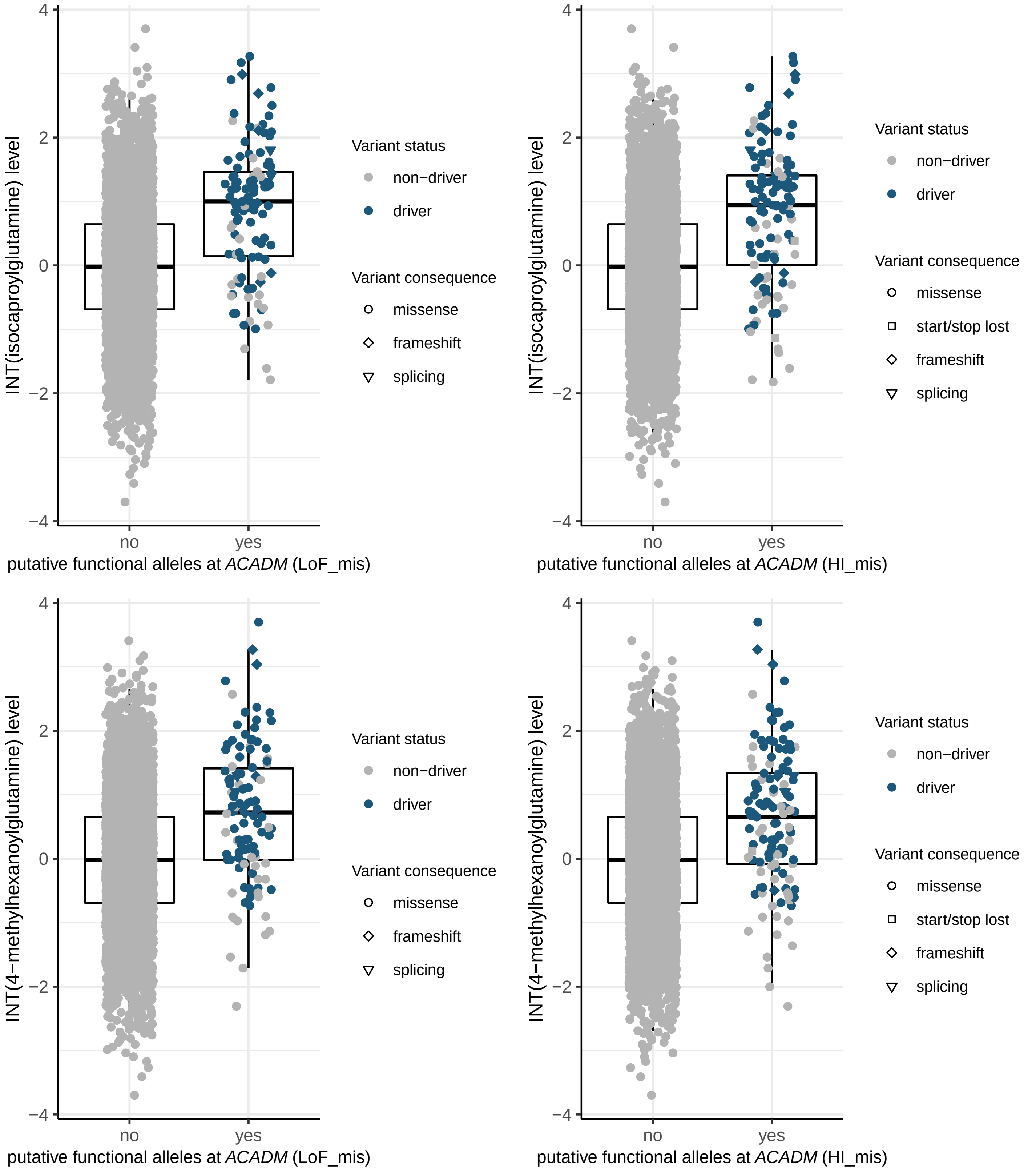
INT(isocaproylglutamine) level
putative functional alleles at ACADM (LoF_mis)
putative functional alleles at ACADM (HI_mis)
INT(4−methylhexanoylglutamine) level
no
yes
4
2
0
−2
−4
Variant status
non−driver
driver
Variant consequence
missense
start/stop lost
frameshift
splicing

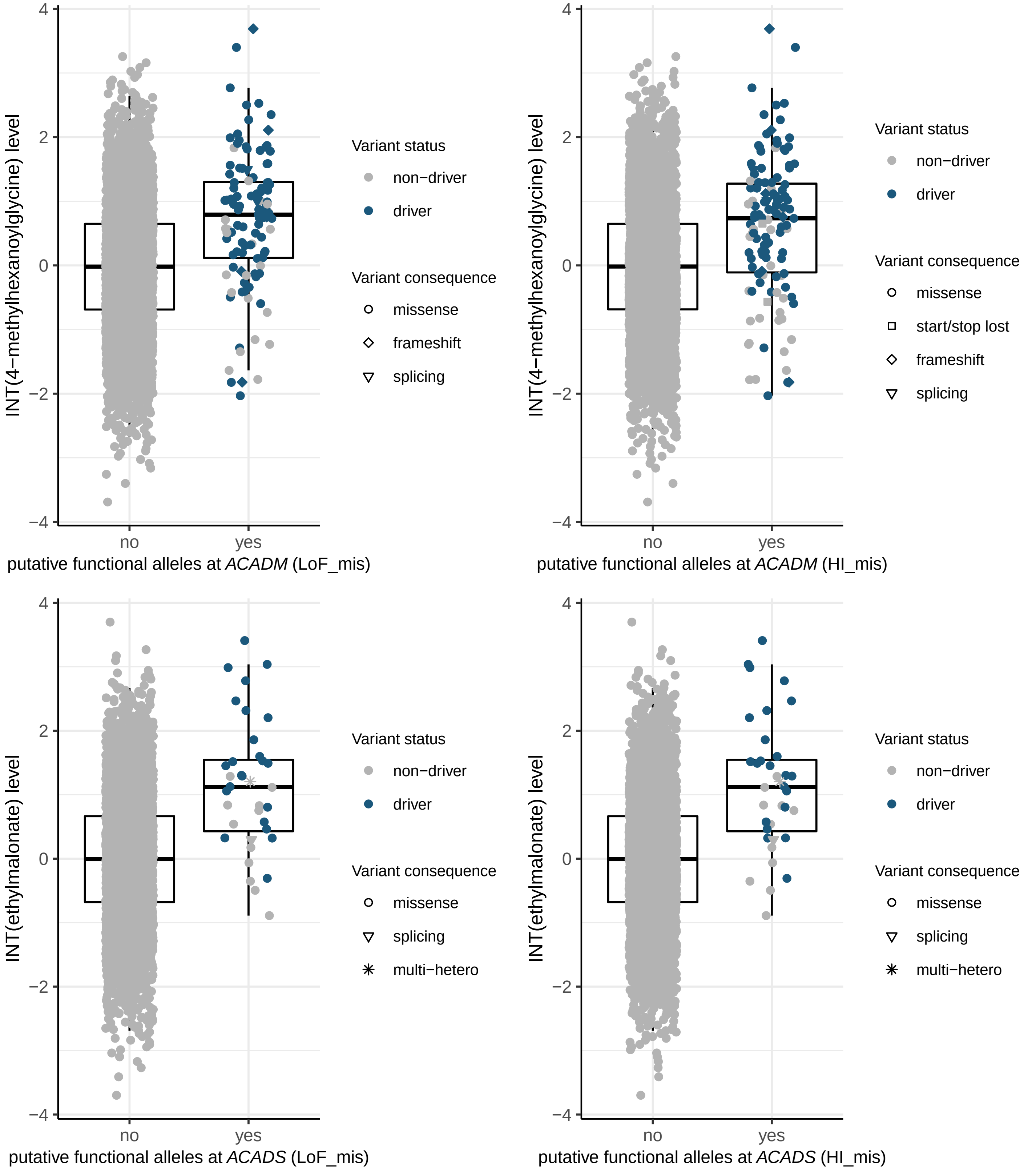
INT(4−methylhexanoylglycine) level
4
2
0
−2
−4
no
yes
putative functional alleles at ACADM (LoF_mis)
Variant status
non−driver
driver
Variant consequence
missense
frameshift
splicing
INT(4−methylhexanoylglycine) level
putative functional alleles at ACADM (HI_mis)
Variant status
non−driver
driver
Variant consequence
missense
start/stop lost
frameshift
splicing
INT(ethylmalonate) level
putative functional alleles at ACADS (LoF_mis)
Variant status
non−driver
driver
Variant consequence
missense
splicing
multi−hetero
INT(ethylmalonate) level
putative functional alleles at ACADS (HI_mis)
Variant status
non−driver
driver
Variant consequence
missense
splicing
multi−hetero

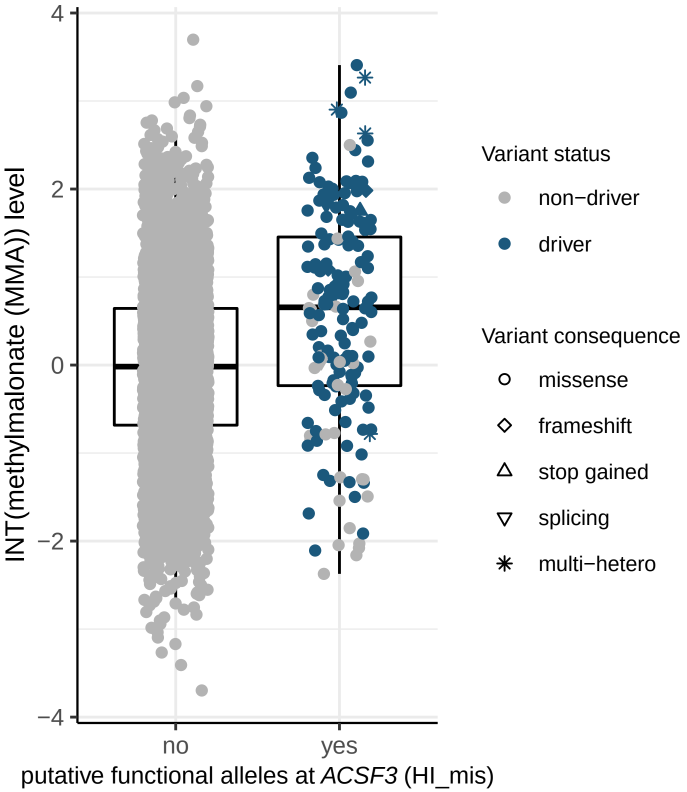

INT(methylmalonate (MMA)) level
putative functional alleles at *ACSF3* (HI_mis)
no
yes
4
2
0
−2
−4
Variant status
non−driver
driver
Variant consequence
missense
frameshift
stop gained
splicing
multi−hetero


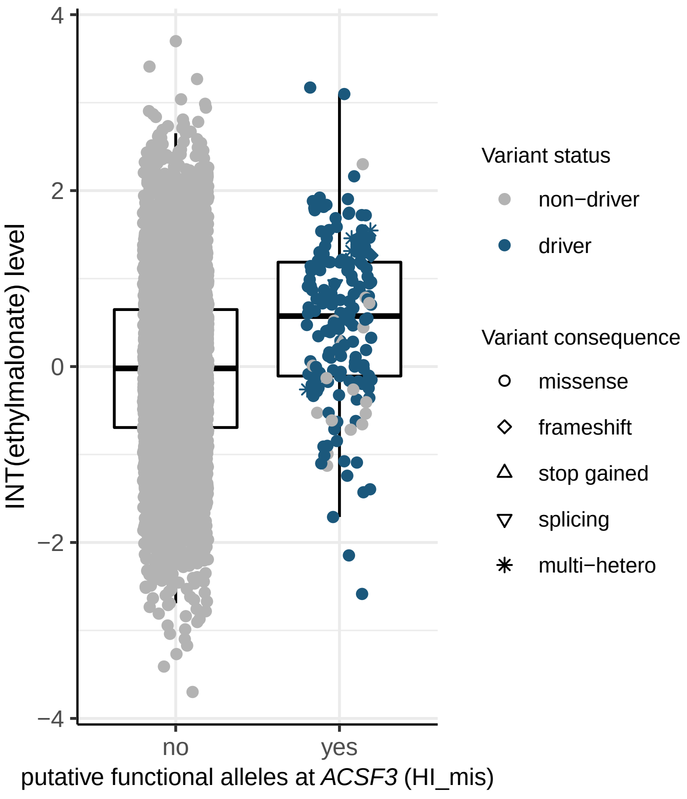

INT(ethylmalonate) level
putative functional alleles at *ACSF3* (HI_mis)
no
yes
4
2
0
−2
−4
Variant status
non−driver
driver
Variant consequence
missense
frameshift
stop gained
splicing
multi−hetero

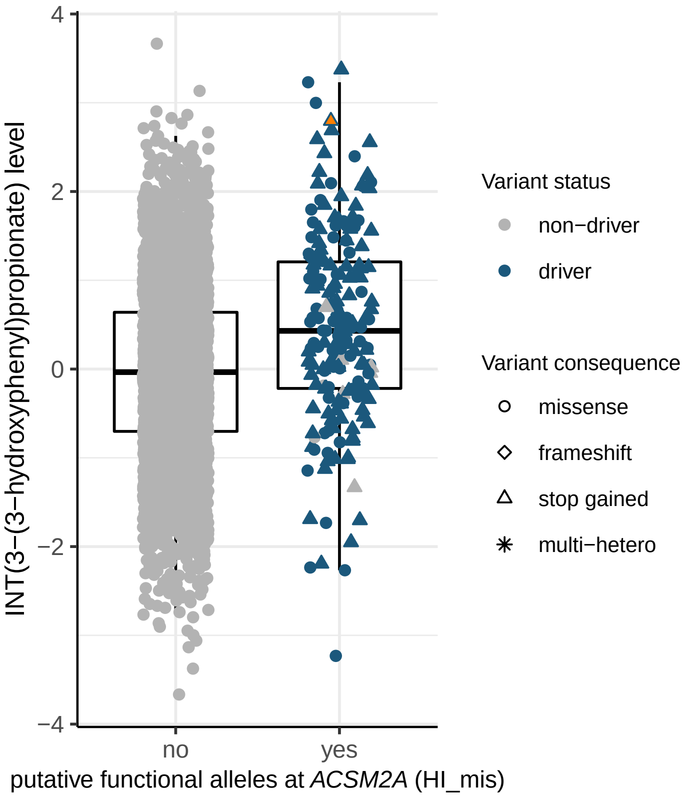
INT(3−(3−hydroxyphenyl)propionate) level
4
2
0
−2
−4
no
yes
putative functional alleles at ACSM2A (HI_mis)
Variant status
non−driver
driver
Variant consequence
missense
frameshift
stop gained
multi−hetero

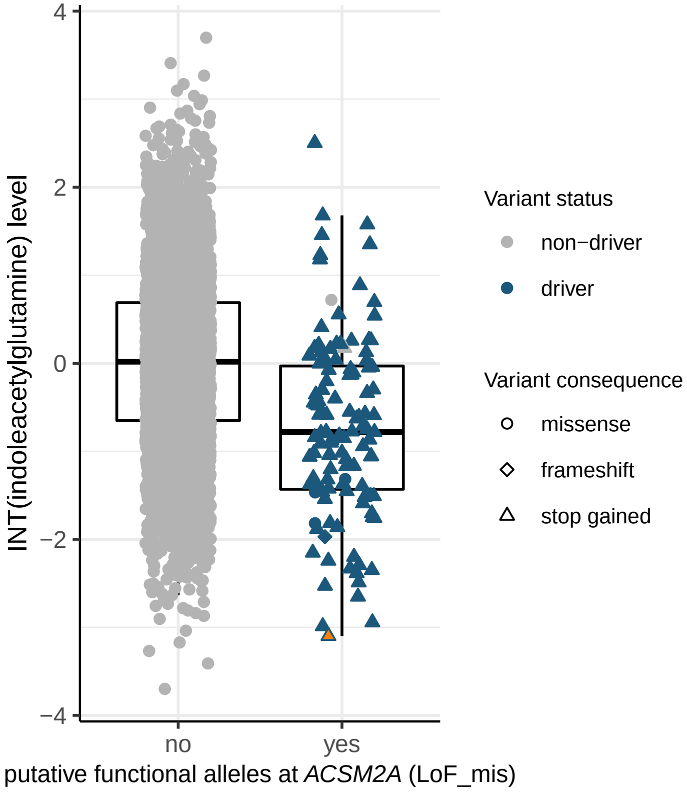
INT(indoleacetylglutamine) level
4
2
0
−2
−4
no
yes
putative functional alleles at ACSM2A (LoF_mis)
Variant status
non−driver
driver
Variant consequence
missense
frameshift
stop gained

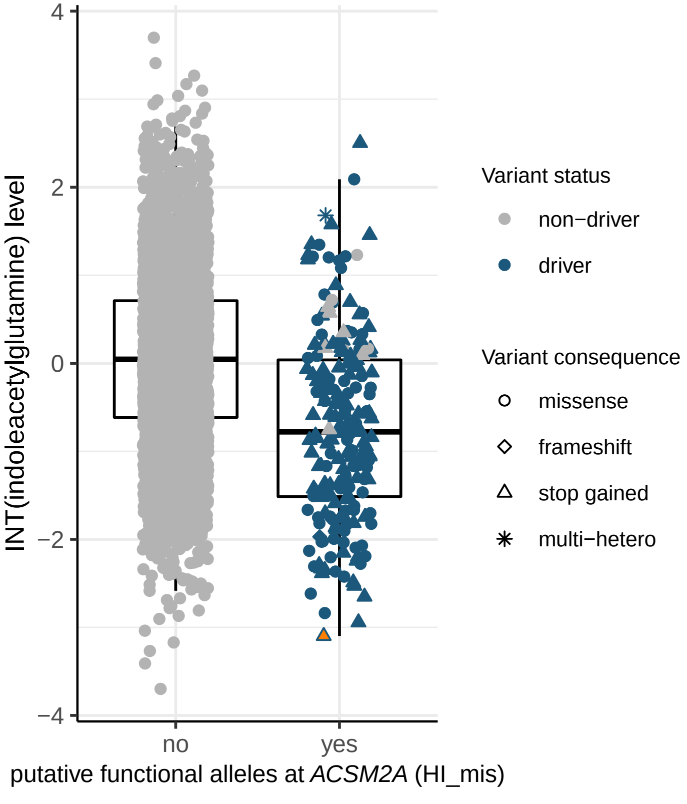
INT(indoleacetylglutamine) level
4
2
0
−2
−4
no
yes
putative functional alleles at ACSM2A (HI_mis)
Variant status
non−driver
driver
Variant consequence
missense
frameshift
stop gained
multi−hetero

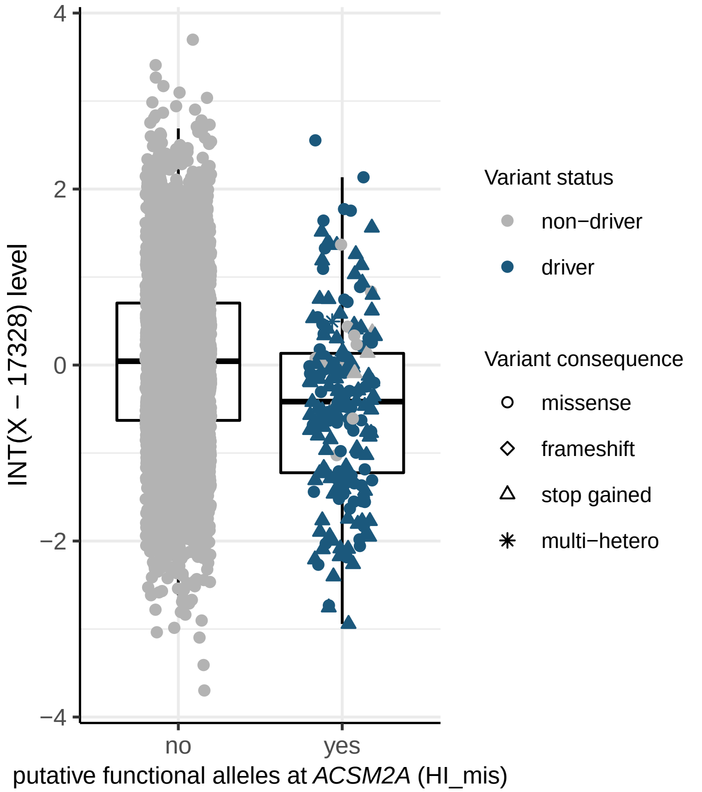

INT(X − 17328) level
4
2
0
−2
−4
no
yes
putative functional alleles at *ACSM2A* (HI_mis)
Variant status
non−driver
driver
Variant consequence
missense
frameshift
stop gained
multi−hetero


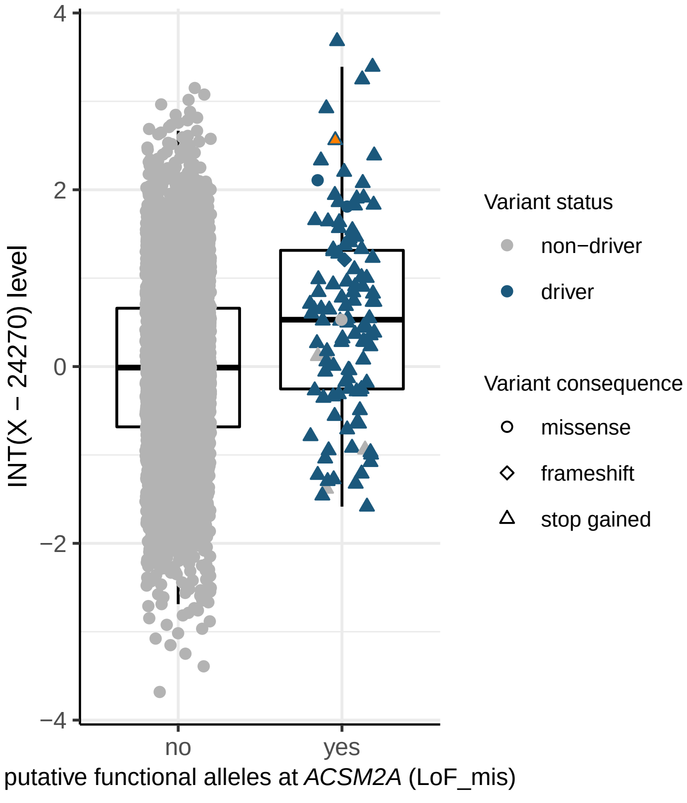

INT(X − 24270) level
4
2
0
−2
−4
no
yes
putative functional alleles at *ACSM2A* (LoF_mis)
Variant status
non−driver
driver
Variant consequence
missense
frameshift
stop gained


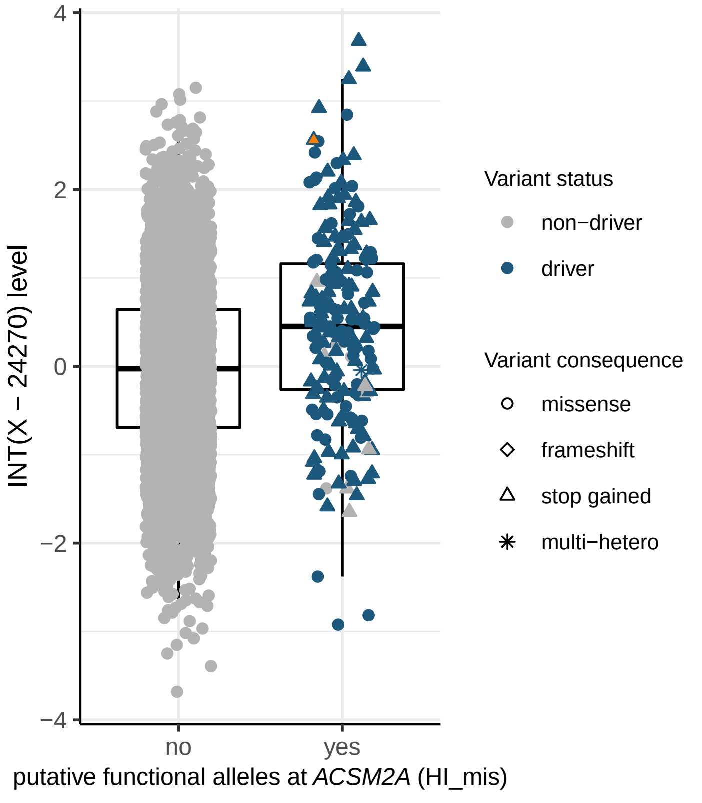

INT(X − 24270) level
4
2
0
−2
−4
no
yes
putative functional alleles at *ACSM2A* (HI_mis)
Variant status
non−driver
driver
Variant consequence
missense
frameshift
stop gained
multi−hetero

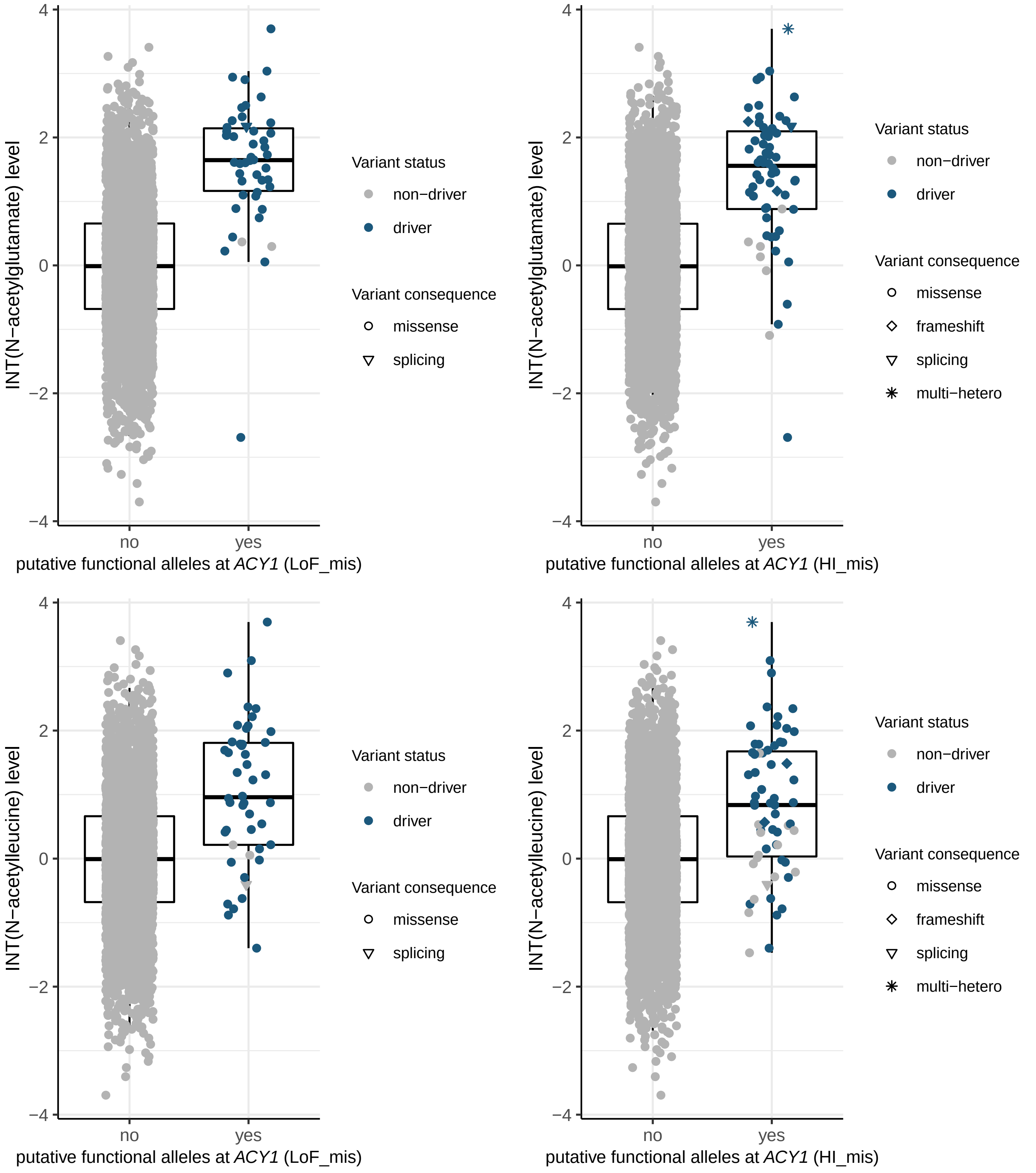
INT(N−acetylglutamate) level
putative functional alleles at ACY1 (LoF_mis)
putative functional alleles at ACY1 (HI_mis)
INT(N−acetylleucine) level
no
yes
Variant status
non−driver
driver
Variant consequence
missense
frameshift
splicing
multi−hetero

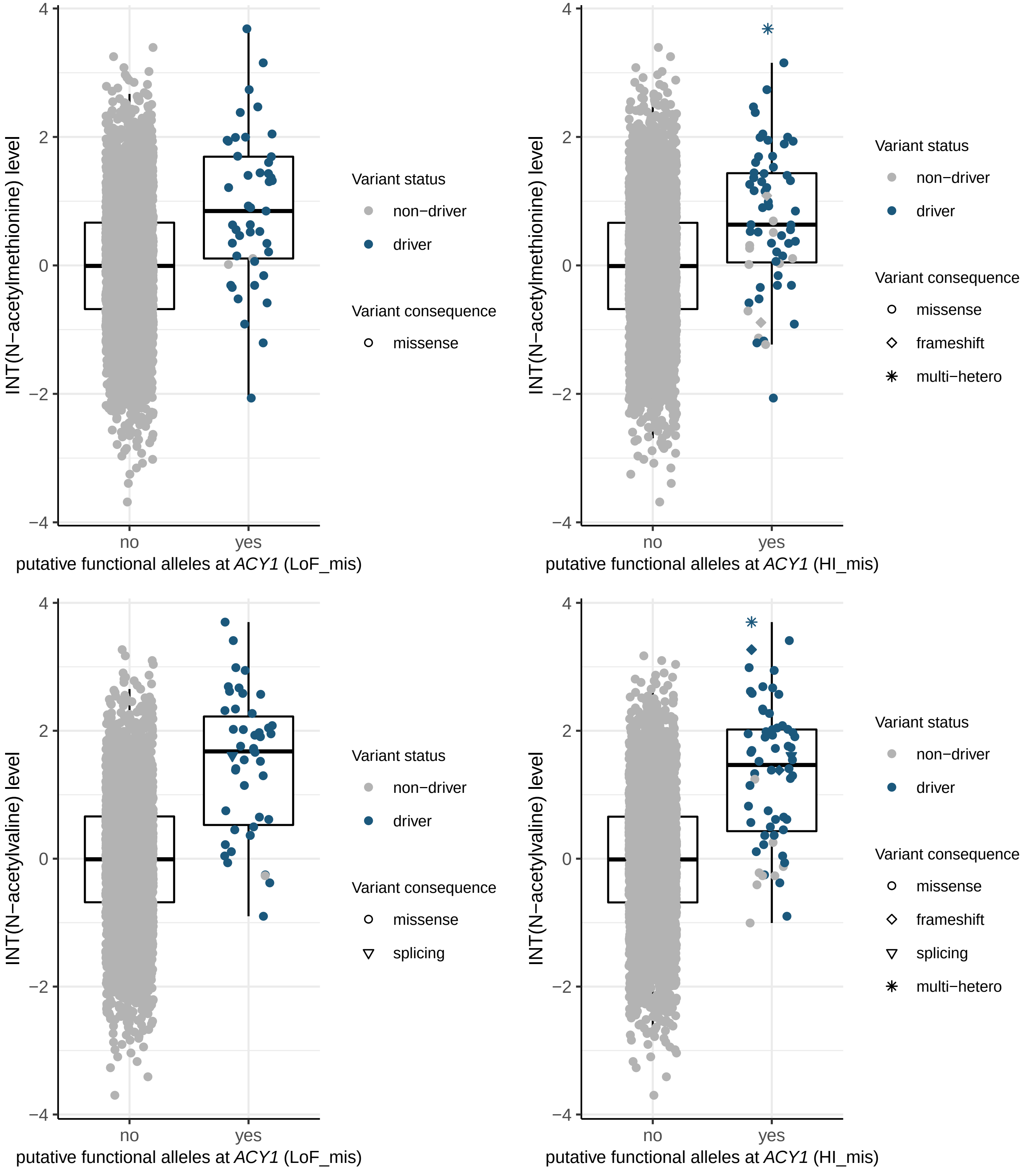
INT(N−acetylmethionine) level
putative functional alleles at ACY1 (LoF_mis)
putative functional alleles at ACY1 (HI_mis)
INT(N−acetylvaline) level
putative functional alleles at ACY1 (LoF_mis)
putative functional alleles at ACY1 (HI_mis)
4
2
0
−2
−4
no
yes
Variant status
non−driver
driver
Variant consequence
missense
frameshift
splicing
multi−hetero

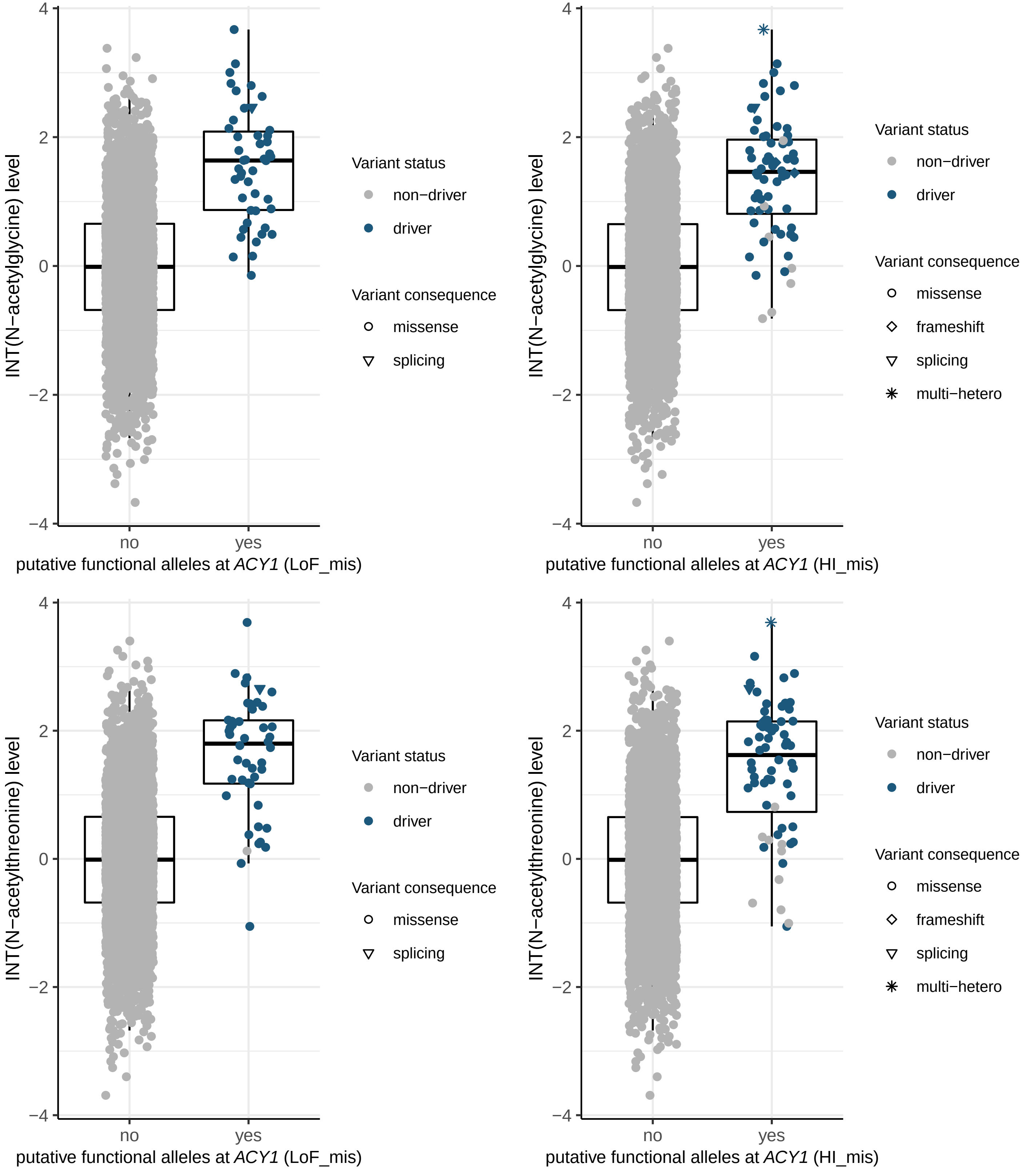
INT(N−acetylglycine) level
putative functional alleles at ACY1 (LoF_mis)
putative functional alleles at ACY1 (HI_mis)
INT(N−acetylthreonine) level
no
yes
Variant status
non−driver
driver
Variant consequence
missense
frameshift
splicing
multi−hetero

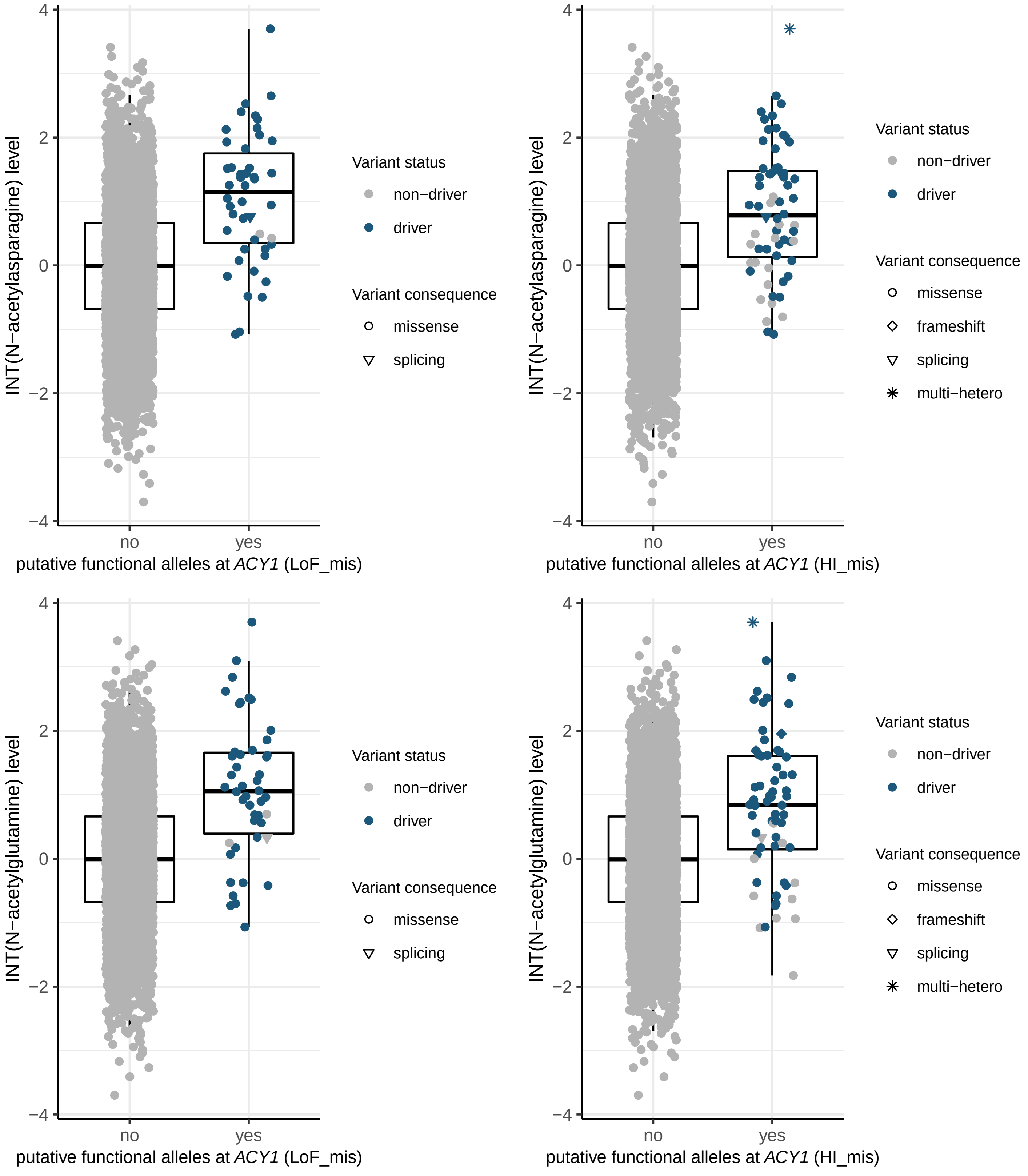

INT(N−acetylasparagine) level
putative functional alleles at ACY1 (LoF_mis)
putative functional alleles at ACY1 (HI_mis)
INT(N−acetylglutamine) level
no
yes
4
2
0
−2
−4
Variant status
non−driver
driver
Variant consequence
missense
frameshift
splicing
multi−hetero

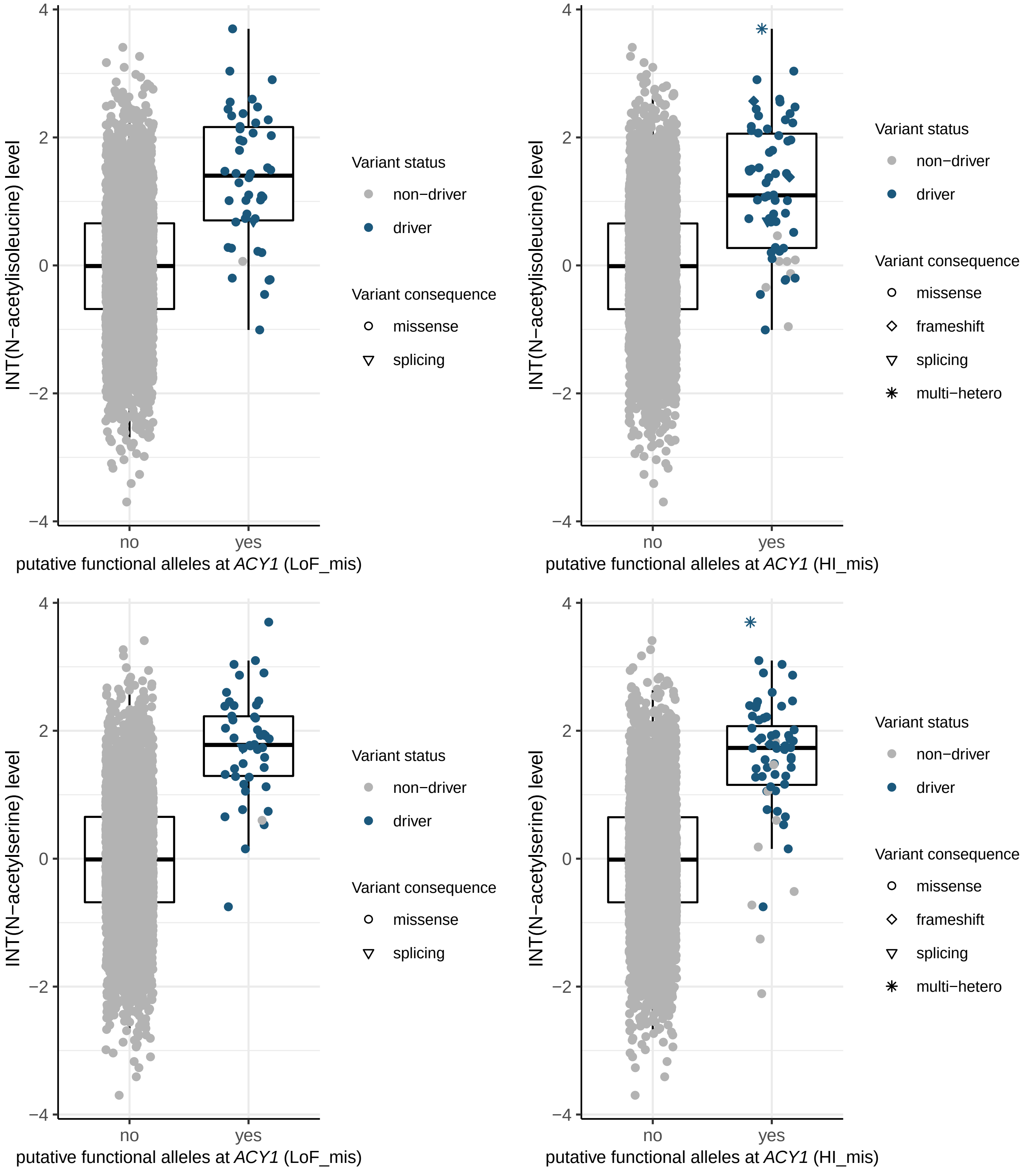

INT(N−acetylisoleucine) level
putative functional alleles at ACY1 (LoF_mis)
putative functional alleles at ACY1 (HI_mis)
INT(N−acetylserine) level
no
yes
Variant status
non−driver
driver
Variant consequence
missense
frameshift
splicing
multi−hetero

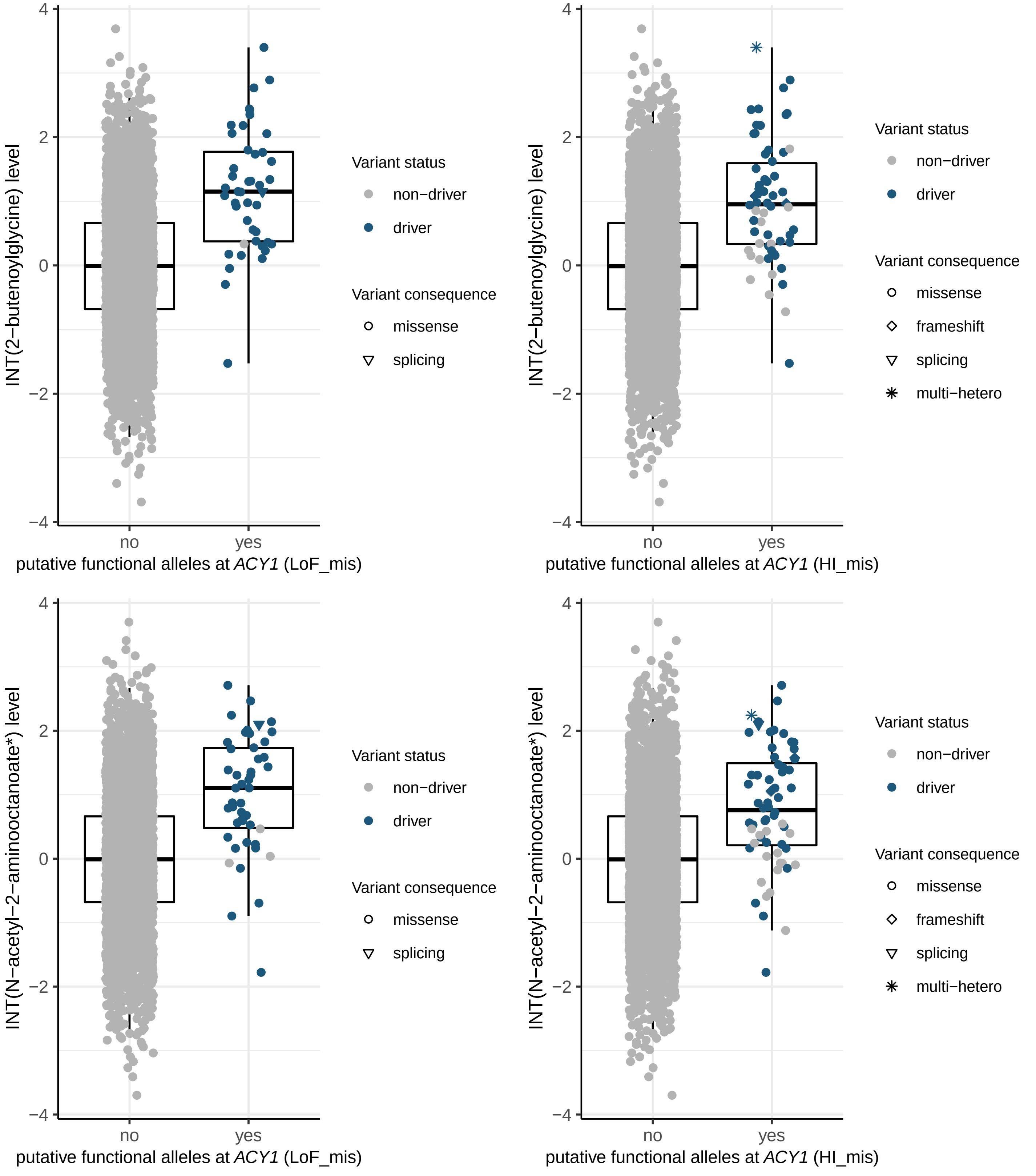

INT(2−butenoylglycine) level
putative functional alleles at ACY1 (LoF_mis)
putative functional alleles at ACY1 (HI_mis)
INT(N−acetyl−2−aminooctanoate*) level
no
yes
Variant status
non−driver
driver
Variant consequence
missense
frameshift
splicing
multi−hetero

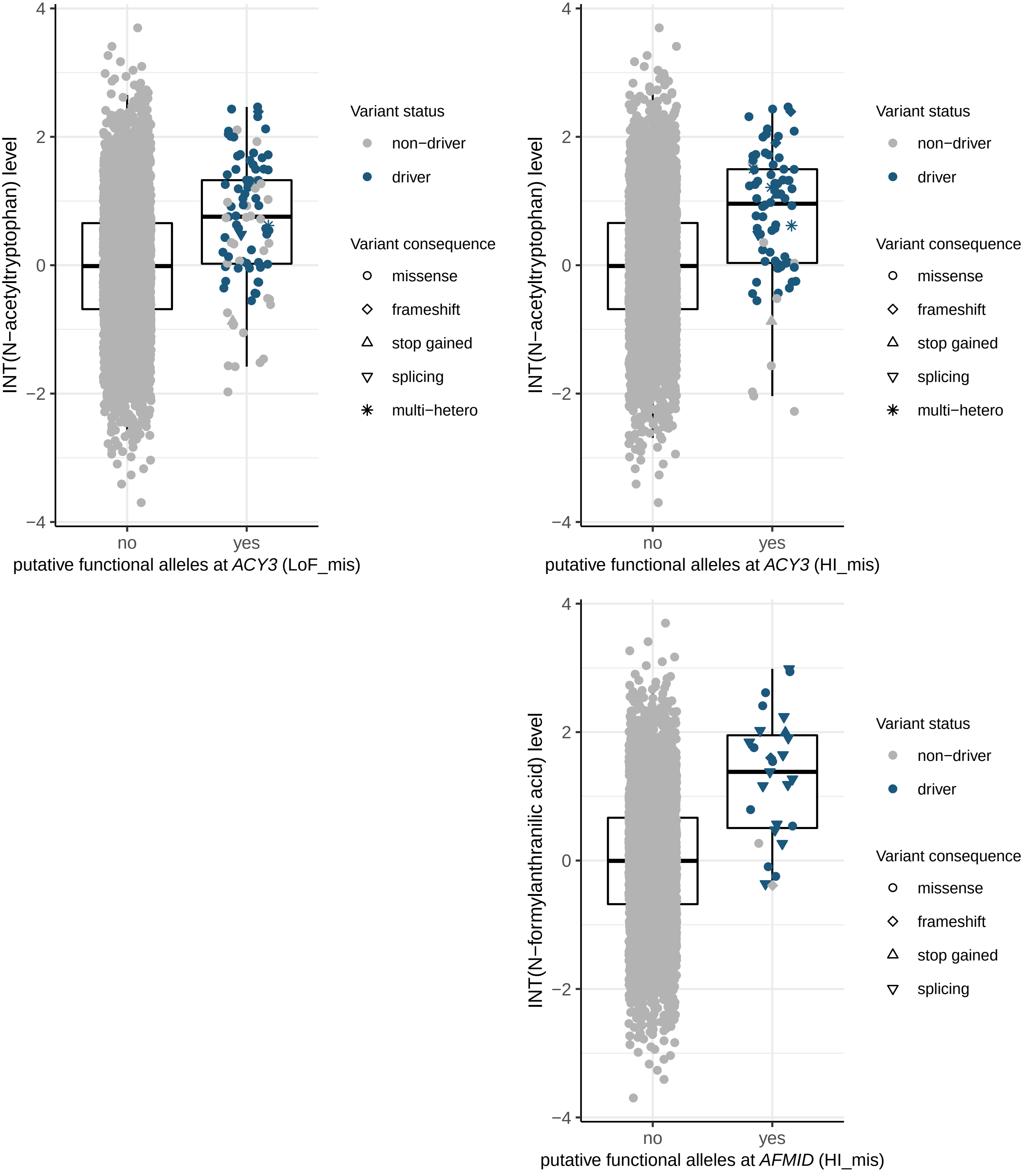

INT(N−acetyltryptophan) level
putative functional alleles at ACY3 (LoF_mis)
putative functional alleles at ACY3 (HI_mis)
INT(N−formylanthranilic acid) level
putative functional alleles at AFMID (HI_mis)
4
2
0
−2
−4
no
yes
Variant status
non−driver
driver
Variant consequence
missense
frameshift
stop gained
splicing
multi−hetero

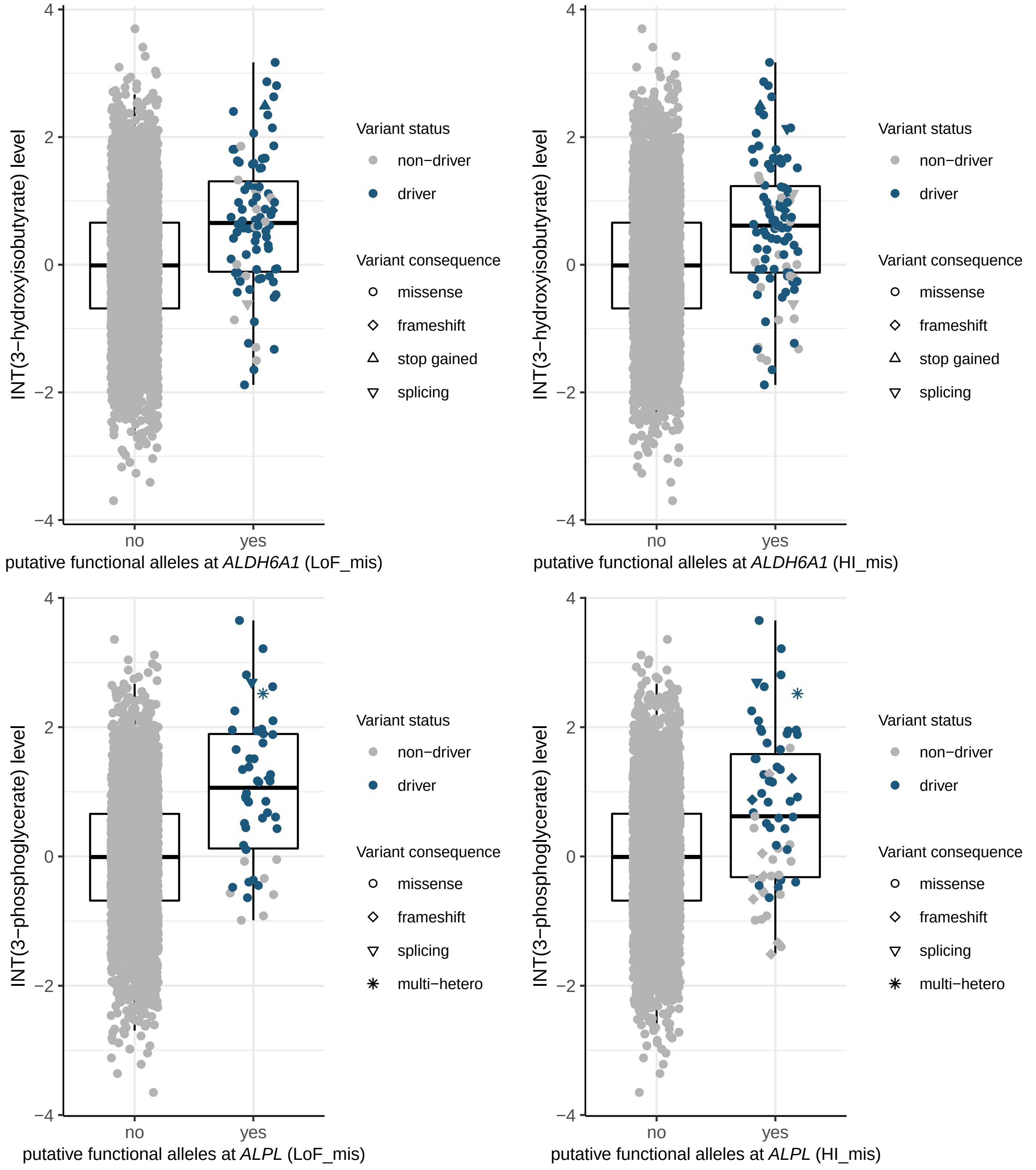
INT(3−hydroxyisobutyrate) level
putative functional alleles at ALDH6A1 (LoF_mis)
putative functional alleles at ALDH6A1 (HI_mis)
INT(3−phosphoglycerate) level
putative functional alleles at ALPL (LoF_mis)
putative functional alleles at ALPL (HI_mis)
no
yes
4
2
0
−2
−4
Variant status
non−driver
driver
Variant consequence
missense
frameshift
stop gained
splicing
multi−hetero

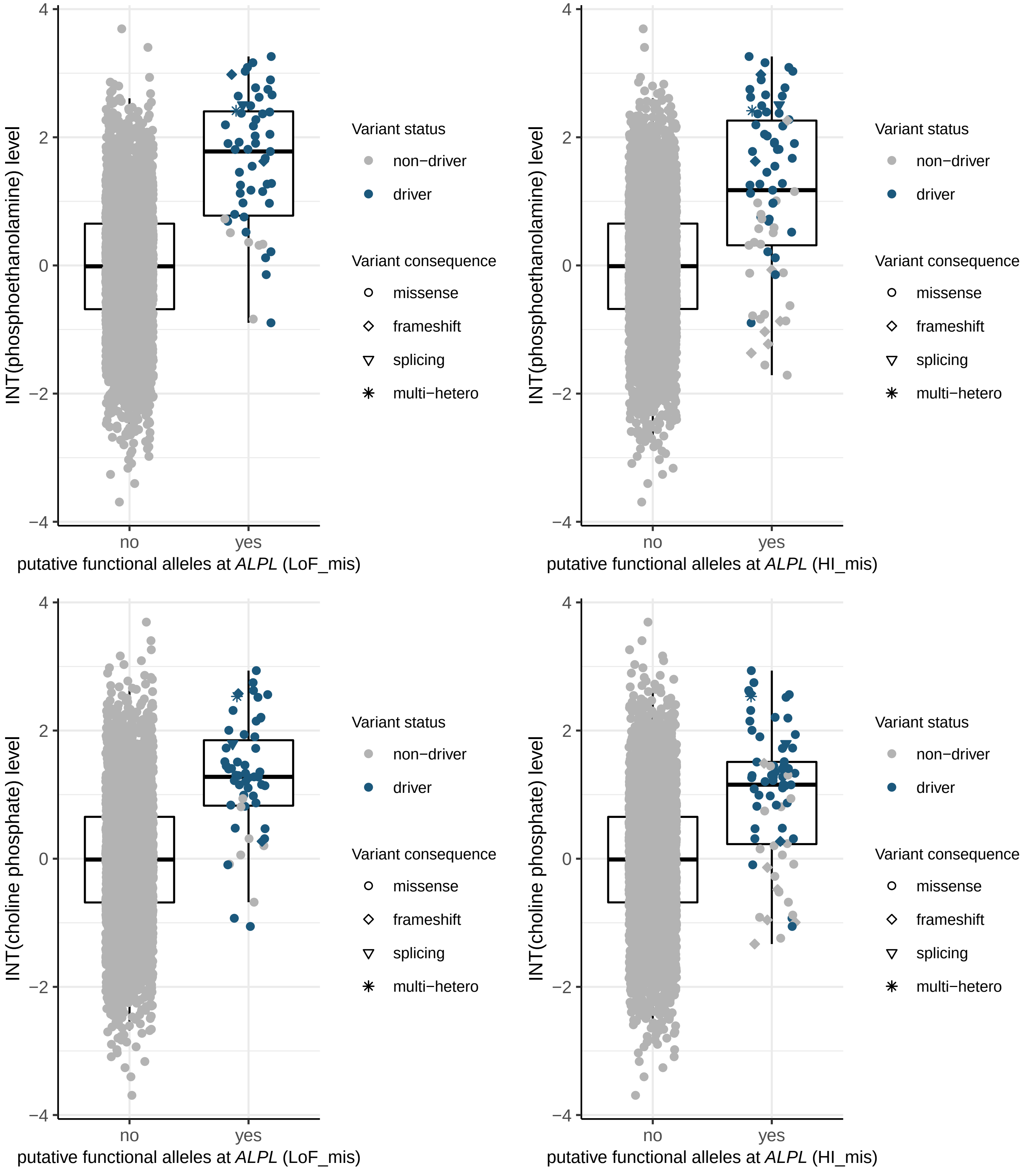

INT(phosphoethanolamine) level
putative functional alleles at ALPL (LoF_mis)
putative functional alleles at ALPL (HI_mis)
INT(choline phosphate) level
no
yes
4
2
0
−2
−4
Variant status
non−driver
driver
Variant consequence
missense
frameshift
splicing
multi−hetero

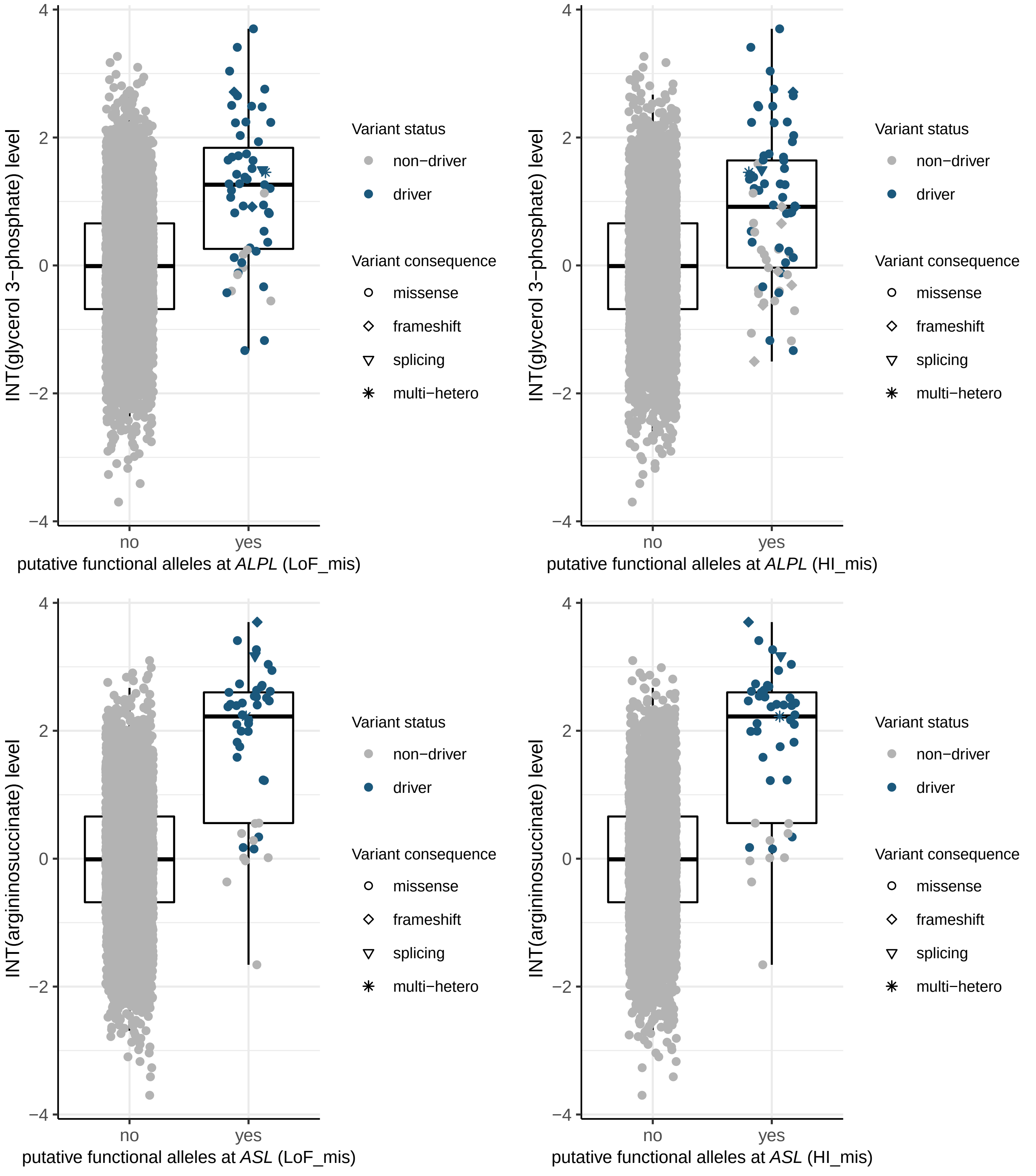

INT(glycerol 3−phosphate) level
putative functional alleles at ALPL (LoF_mis)
putative functional alleles at ALPL (HI_mis)
INT(argininosuccinate) level
putative functional alleles at ASL (LoF_mis)
putative functional alleles at ASL (HI_mis)
no
yes
Variant status
non−driver
driver
Variant consequence
missense
frameshift
splicing
multi−hetero

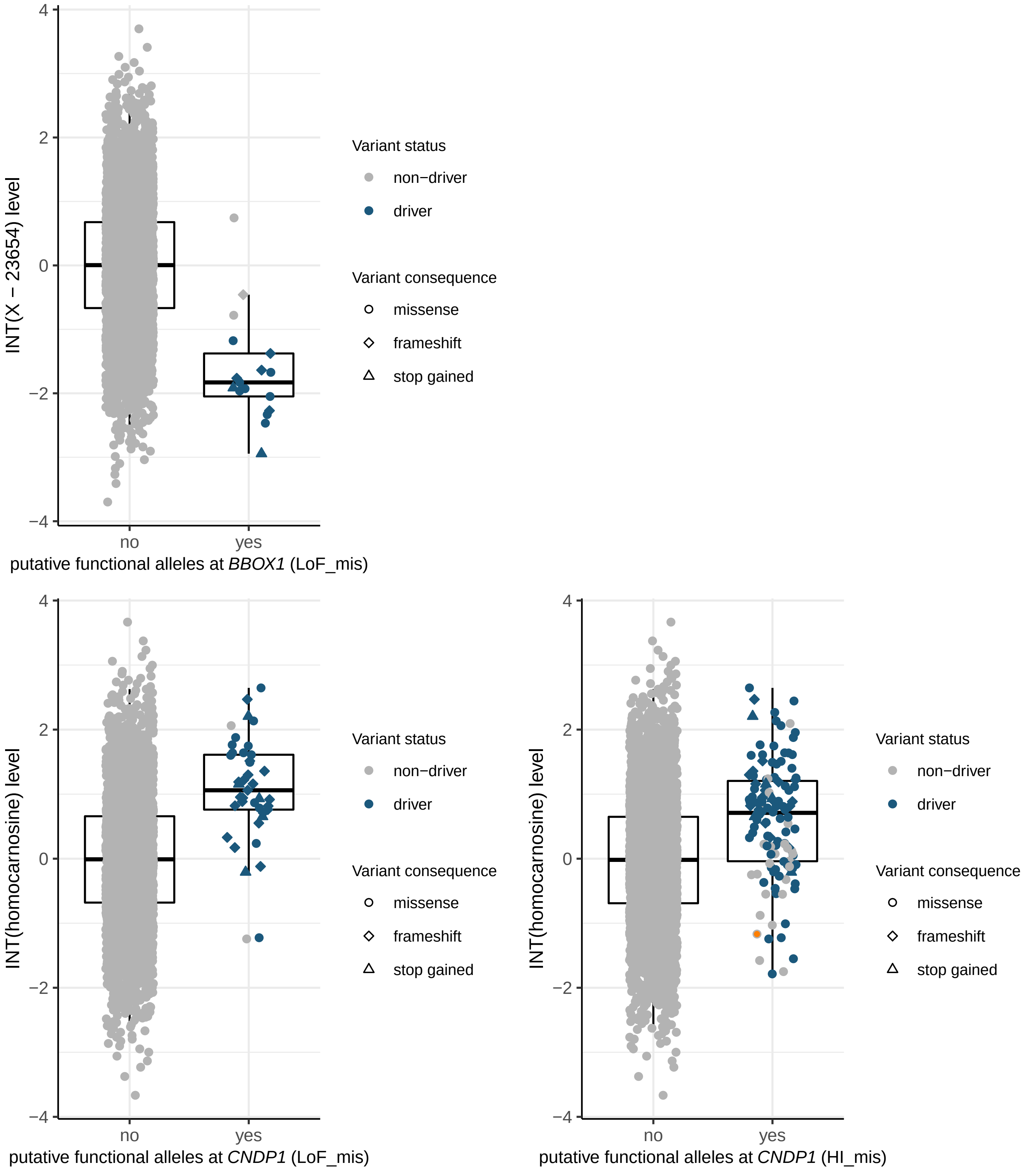
INT(X − 23654) level
putative functional alleles at BBOX1 (LoF_mis)
no
yes
4
2
0
−2
−4
Variant status
non−driver
driver
Variant consequence
missense
frameshift
stop gained
INT(homocarnosine) level
putative functional alleles at CNDP1 (LoF_mis)
putative functional alleles at CNDP1 (HI_mis)

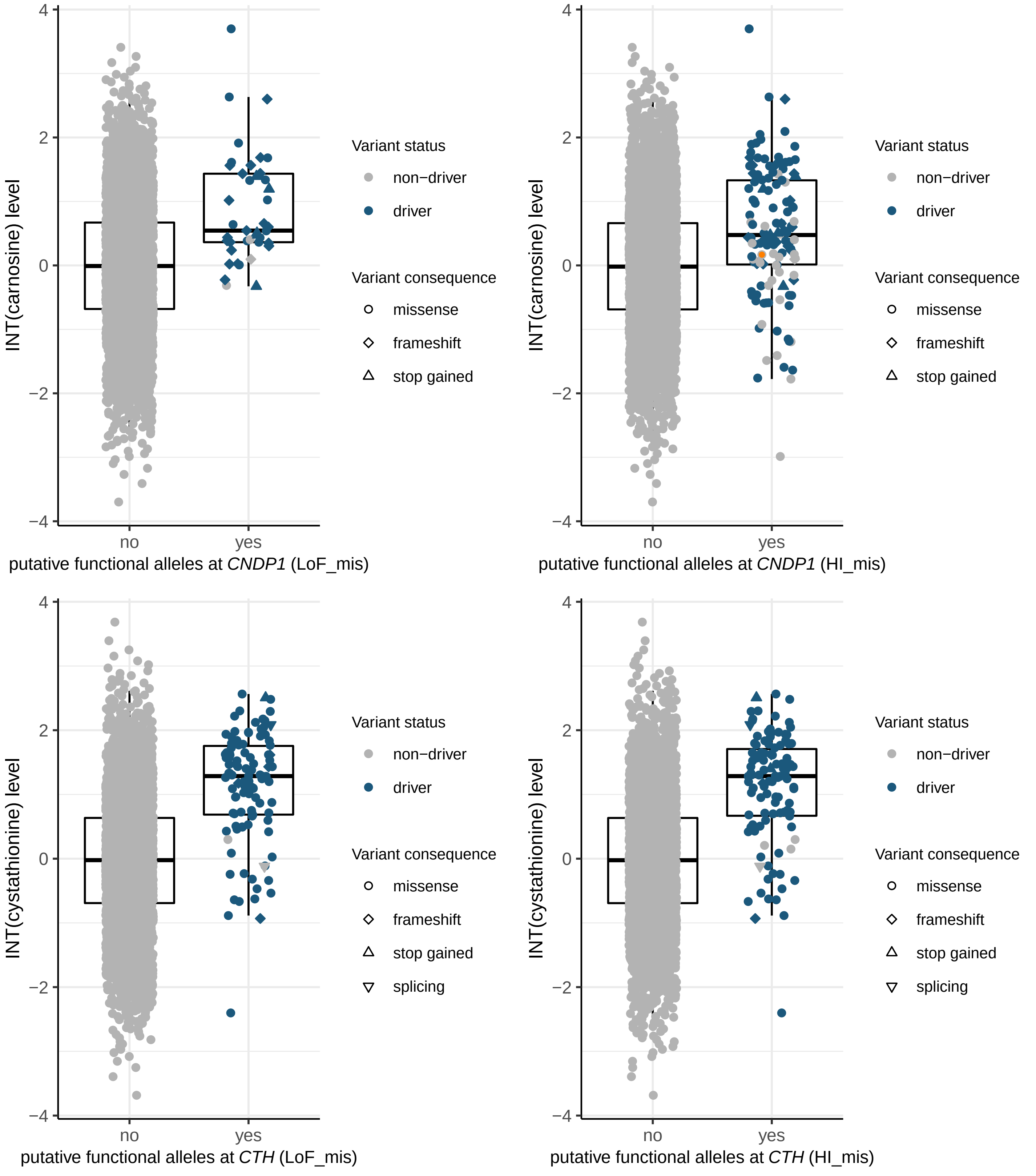

INT(carnosine) level
putative functional alleles at CNDP1 (LoF_mis)
putative functional alleles at CNDP1 (HI_mis)
INT(cystathionine) level
putative functional alleles at CTH (LoF_mis)
putative functional alleles at CTH (HI_mis)
no
yes
4
2
0
−2
−4
Variant status
non−driver
driver
Variant consequence
missense
frameshift
stop gained
splicing

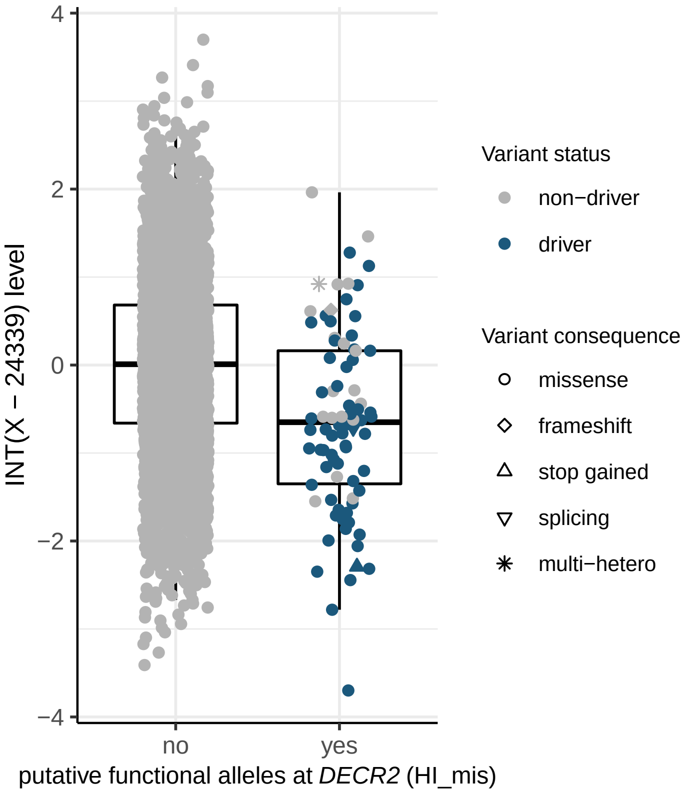

4
2
0
−2
−4
INT(X − 24339) level
no
yes
putative functional alleles at *DECR2* (HI_mis)
Variant status
non−driver
driver
Variant consequence
missense
frameshift
stop gained
splicing
multi−hetero


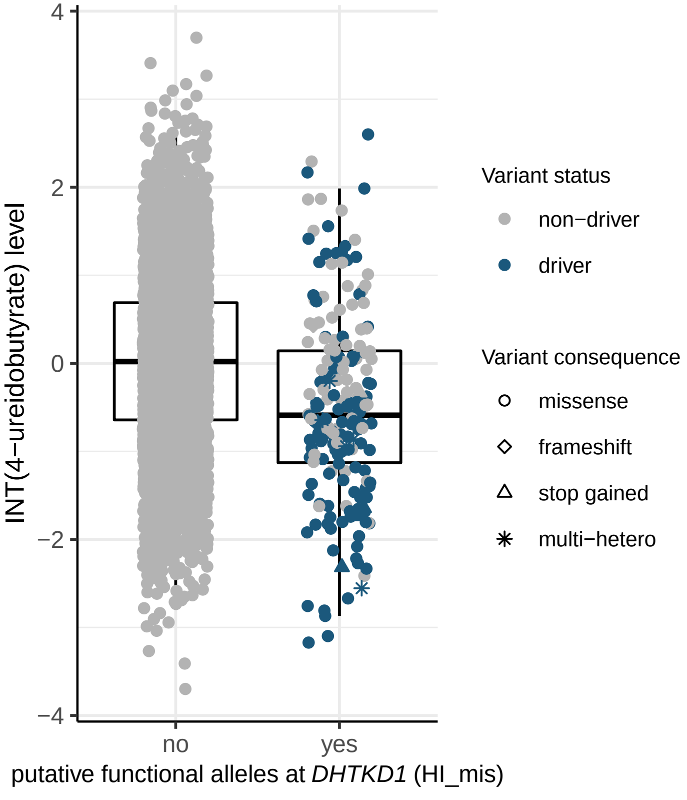

4
2
0
−2
−4
INT(4−ureidobutyrate) level
no
yes
putative functional alleles at *DHTKD1* (HI_mis)
Variant status
non−driver
driver
Variant consequence
missense
frameshift
stop gained
multi−hetero

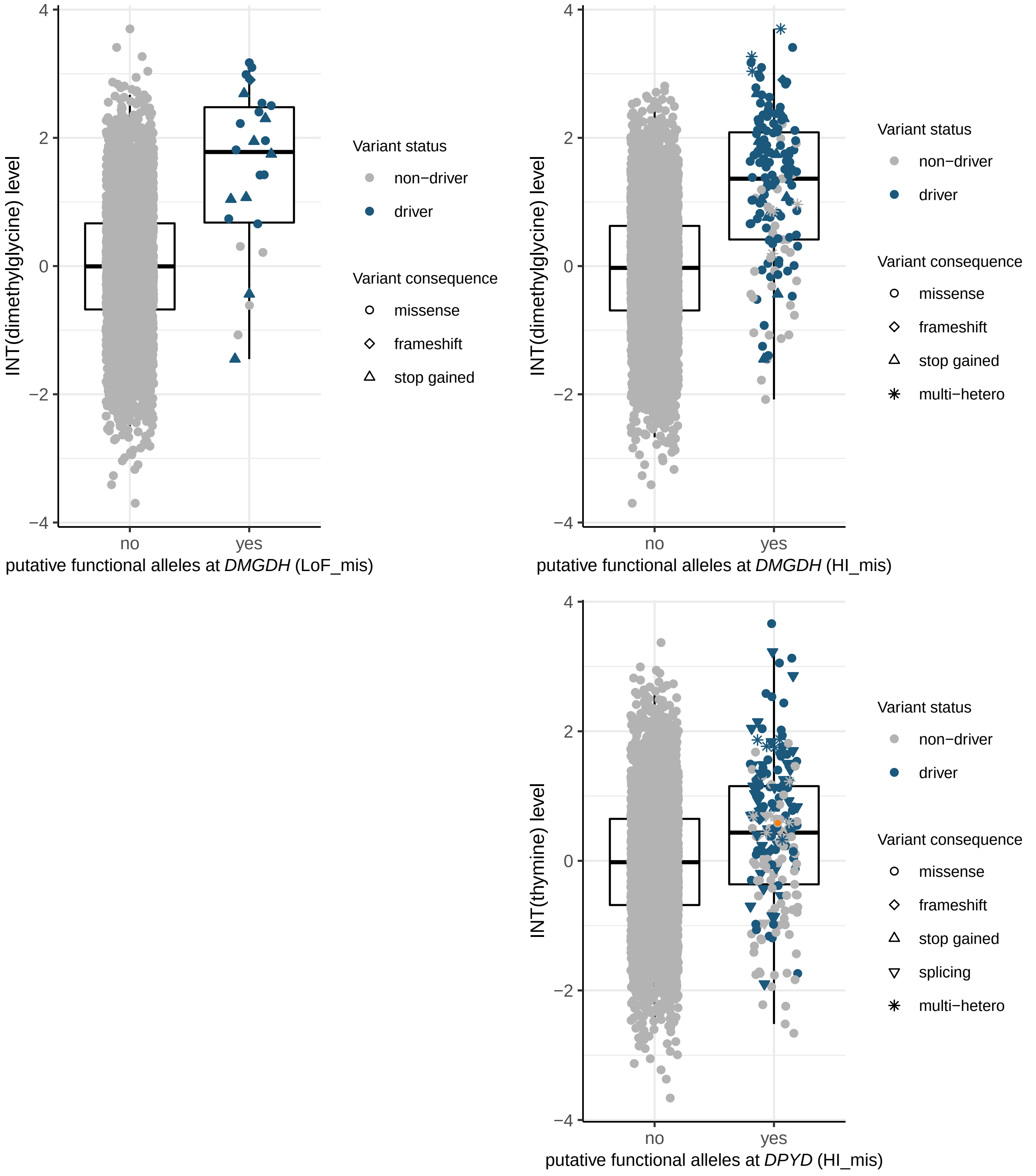

INT(dimethylglycine) level
4
2
0
−2
−4
no
yes
putative functional alleles at *DMGDH* (LoF_mis)
Variant status
non−driver
driver
Variant consequence
missense
frameshift
stop gained
INT(dimethylglycine) level
putative functional alleles at *DMGDH* (HI_mis)
Variant status
non−driver
driver
Variant consequence
missense
frameshift
stop gained
multi−hetero
INT(thymine) level
putative functional alleles at *DPYD* (HI_mis)
Variant status
non−driver
driver
Variant consequence
missense
frameshift
stop gained
splicing
multi−hetero

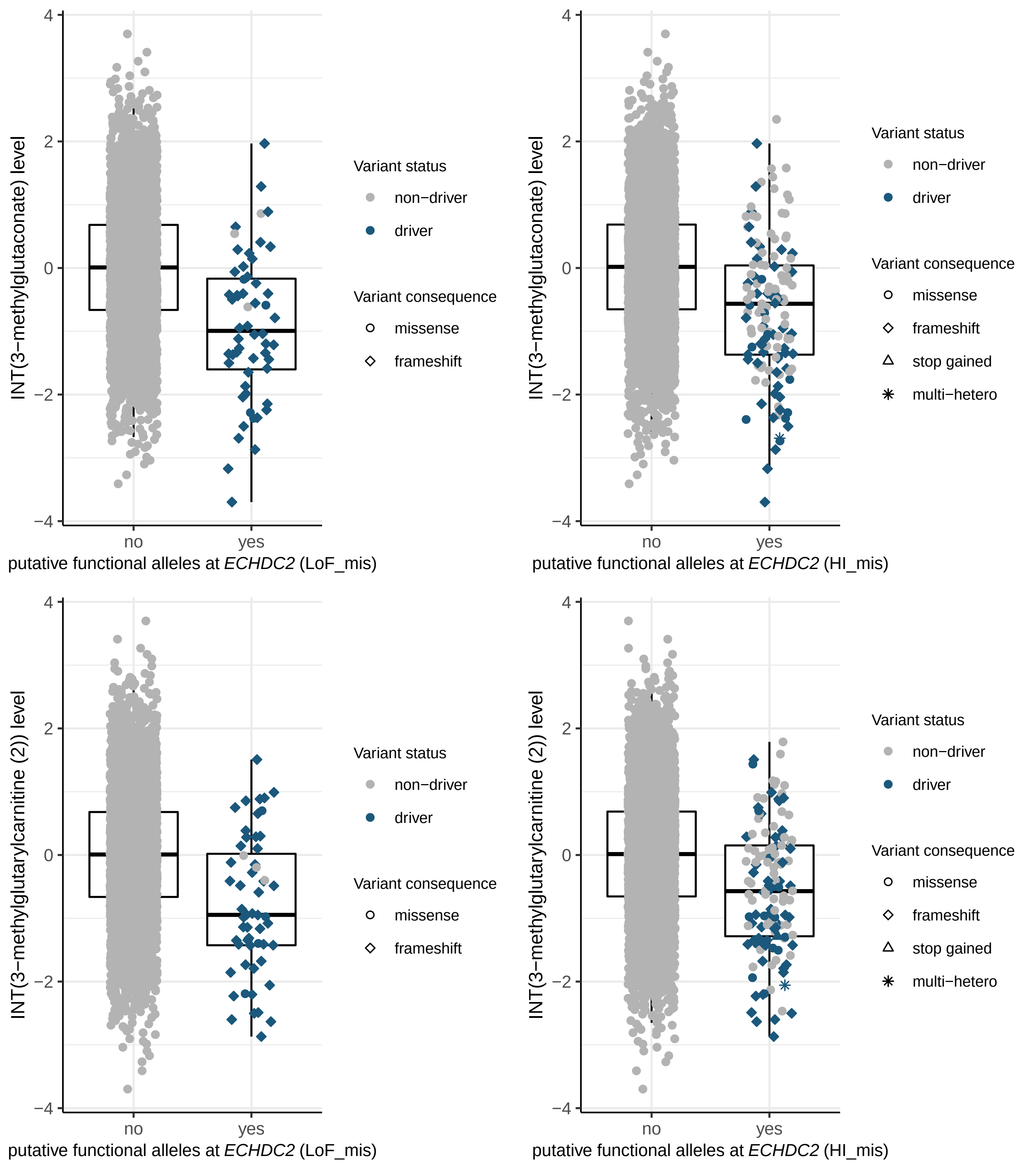
INT(3−methylglutaconate) level
4
2
0
−2
−4
no
yes
putative functional alleles at *ECHDC2* (LoF_mis)
Variant status
non−driver
driver
Variant consequence
missense
frameshift
INT(3−methylglutaconate) level
4
2
0
−2
−4
no
yes
putative functional alleles at *ECHDC2* (HI_mis)
Variant status
non−driver
driver
Variant consequence
missense
frameshift
stop gained
multi−hetero
INT(3−methylglutarylcarnitine (2)) level
4
2
0
−2
−4
no
yes
putative functional alleles at *ECHDC2* (LoF_mis)
Variant status
non−driver
driver
Variant consequence
missense
frameshift
INT(3−methylglutarylcarnitine (2)) level
4
2
0
−2
−4
no
yes
putative functional alleles at *ECHDC2* (HI_mis)
Variant status
non−driver
driver
Variant consequence
missense
frameshift
stop gained
multi−hetero

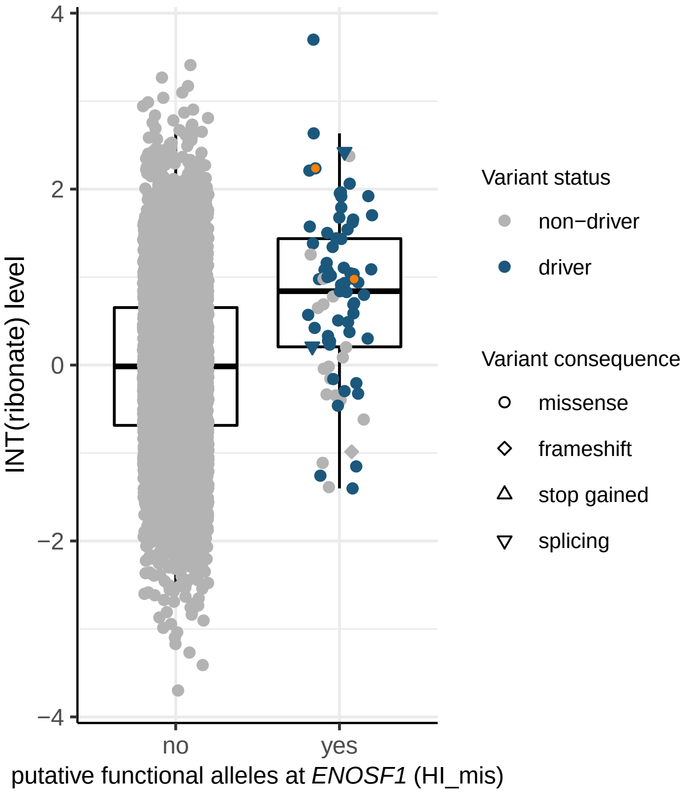
4
2
0
−2
−4
INT(ribonate) level
no
yes
putative functional alleles at ENOSF1 (HI_mis)
Variant status
non−driver
driver
Variant consequence
missense
frameshift
stop gained
splicing

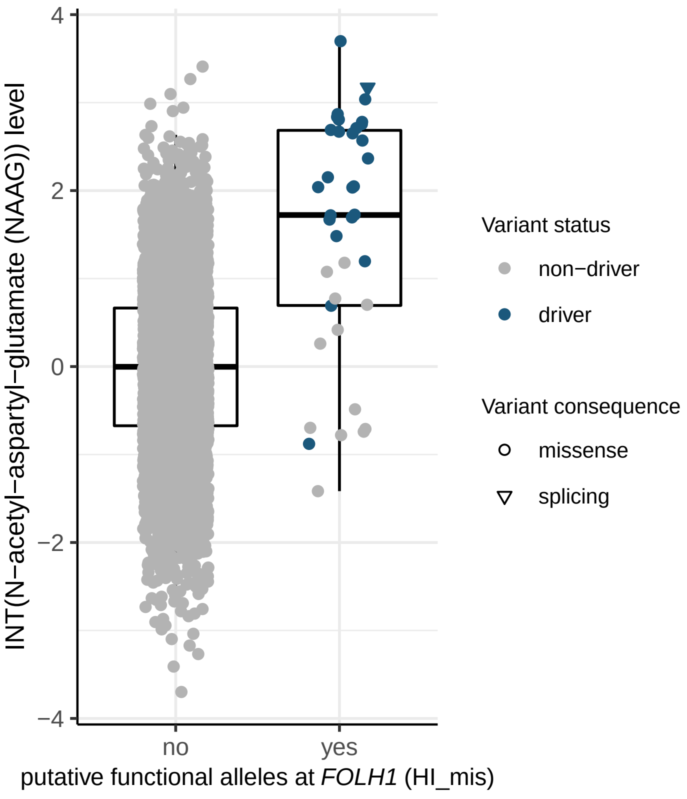
4
2
0
−2
−4
INT(N−acetyl−aspartyl−glutamate (NAAG)) level
no
yes
putative functional alleles at FOLH1 (HI_mis)
Variant status
non−driver
driver
Variant consequence
missense
splicing

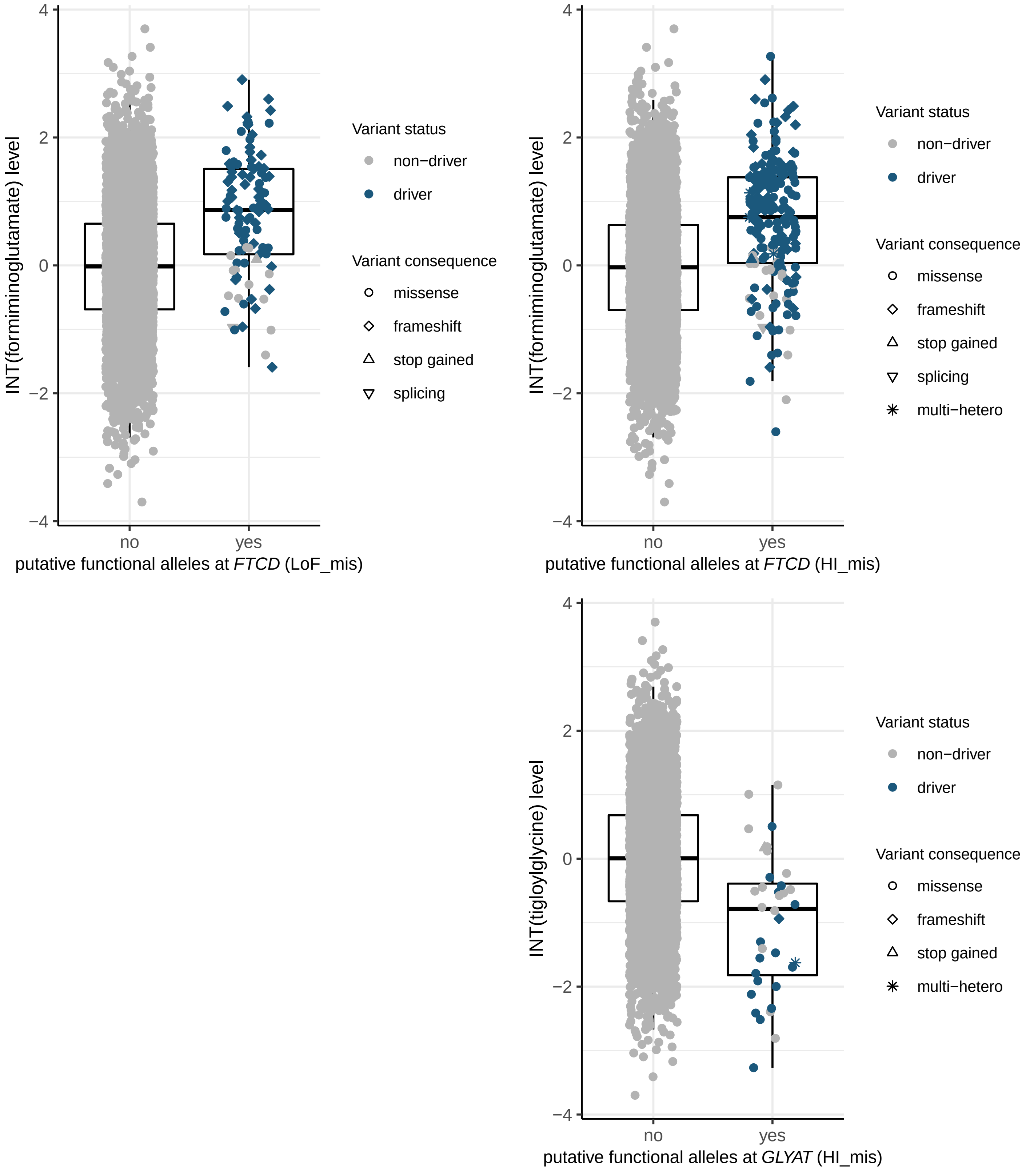
INT(formiminoglutamate) level
4
2
0
−2
−4
no
yes
putative functional alleles at FTCD (LoF_mis)
Variant status
non−driver
driver
Variant consequence
missense
frameshift
stop gained
splicing
INT(formiminoglutamate) level
4
2
0
−2
−4
no
yes
putative functional alleles at FTCD (HI_mis)
Variant status
non−driver
driver
Variant consequence
missense
frameshift
stop gained
splicing
multi−hetero
INT(tigloylglycine) level
4
2
0
−2
−4
no
yes
putative functional alleles at GLYAT (HI_mis)
Variant status
non−driver
driver
Variant consequence
missense
frameshift
stop gained
multi−hetero

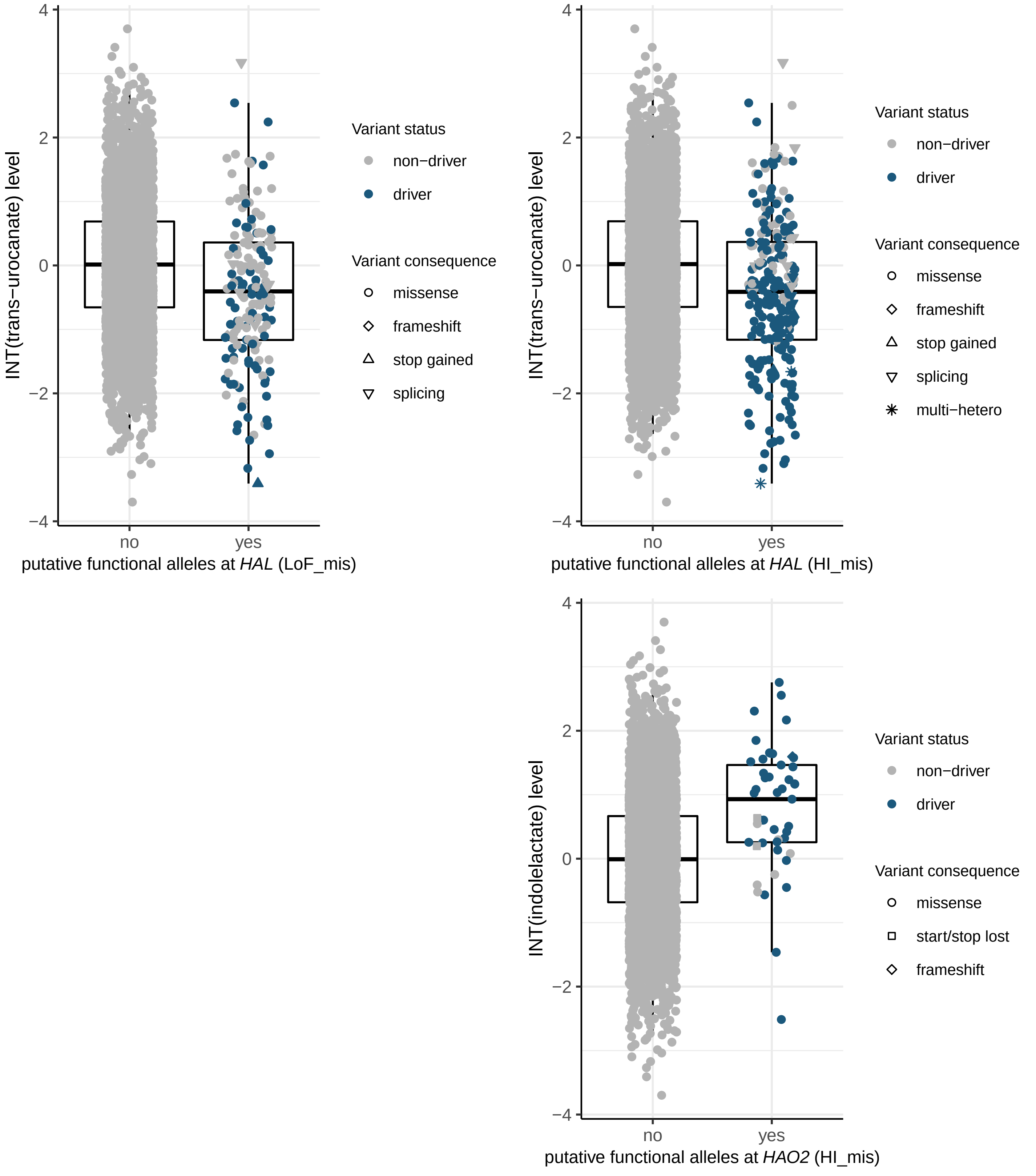

INT(trans−urocanate) level
putative functional alleles at HAL (LoF_mis)
no
yes
4
2
0
−2
−4
Variant status
non−driver
driver
Variant consequence
missense
frameshift
stop gained
splicing
INT(trans−urocanate) level
putative functional alleles at HAL (HI_mis)
Variant status
non−driver
driver
Variant consequence
missense
frameshift
stop gained
splicing
multi−hetero
INT(indolelactate) level
putative functional alleles at HAO2 (HI_mis)
Variant status
non−driver
driver
Variant consequence
missense
start/stop lost
frameshift

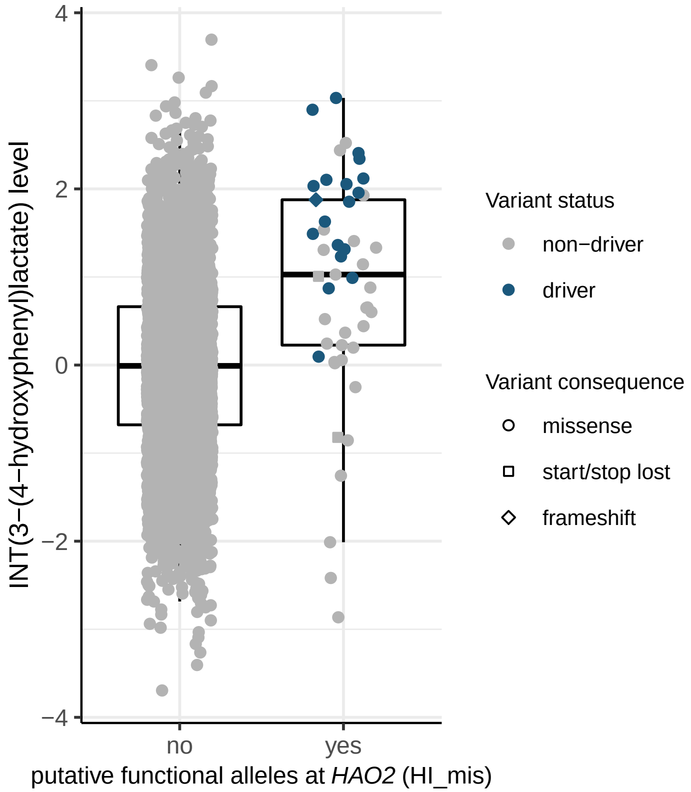

4
2
0
−2
−4
INT(3−(4−hydroxyphenyl)lactate) level
no
yes
putative functional alleles at *HAO2* (HI_mis)
Variant status
non−driver
driver
Variant consequence
missense
start/stop lost
frameshift


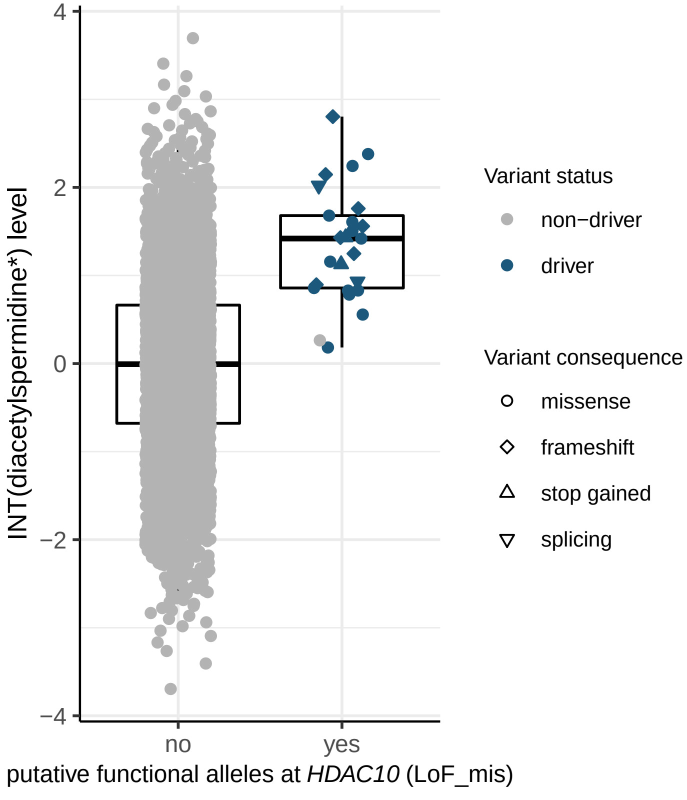

4
2
0
−2
−4
INT(diacetylspermidine*) level
no
yes
putative functional alleles at *HDAC10* (LoF_mis)
Variant status
non−driver
driver
Variant consequence
missense
frameshift
stop gained
splicing


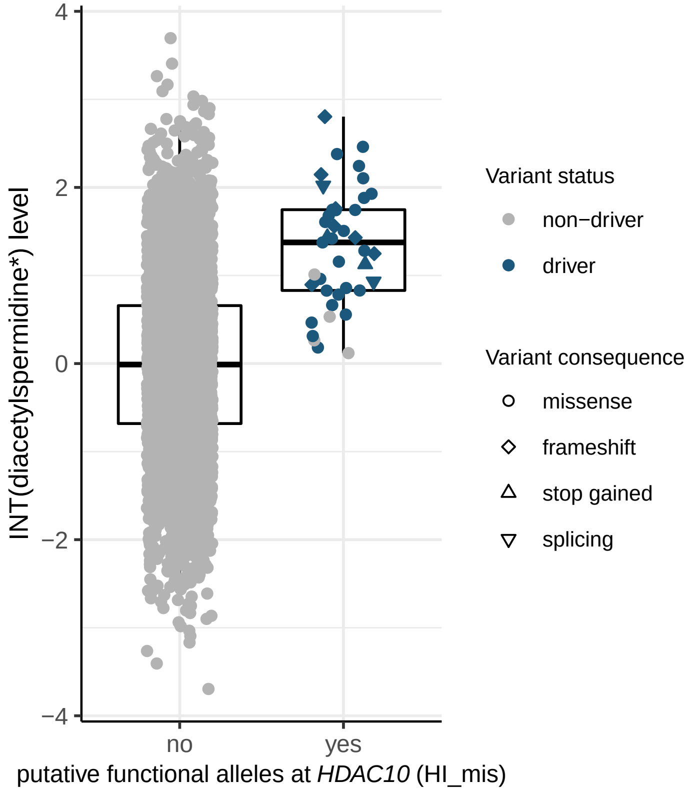

4
2
0
−2
−4
INT(diacetylspermidine*) level
no
yes
putative functional alleles at *HDAC10* (HI_mis)
Variant status
non−driver
driver
Variant consequence
missense
frameshift
stop gained
splicing

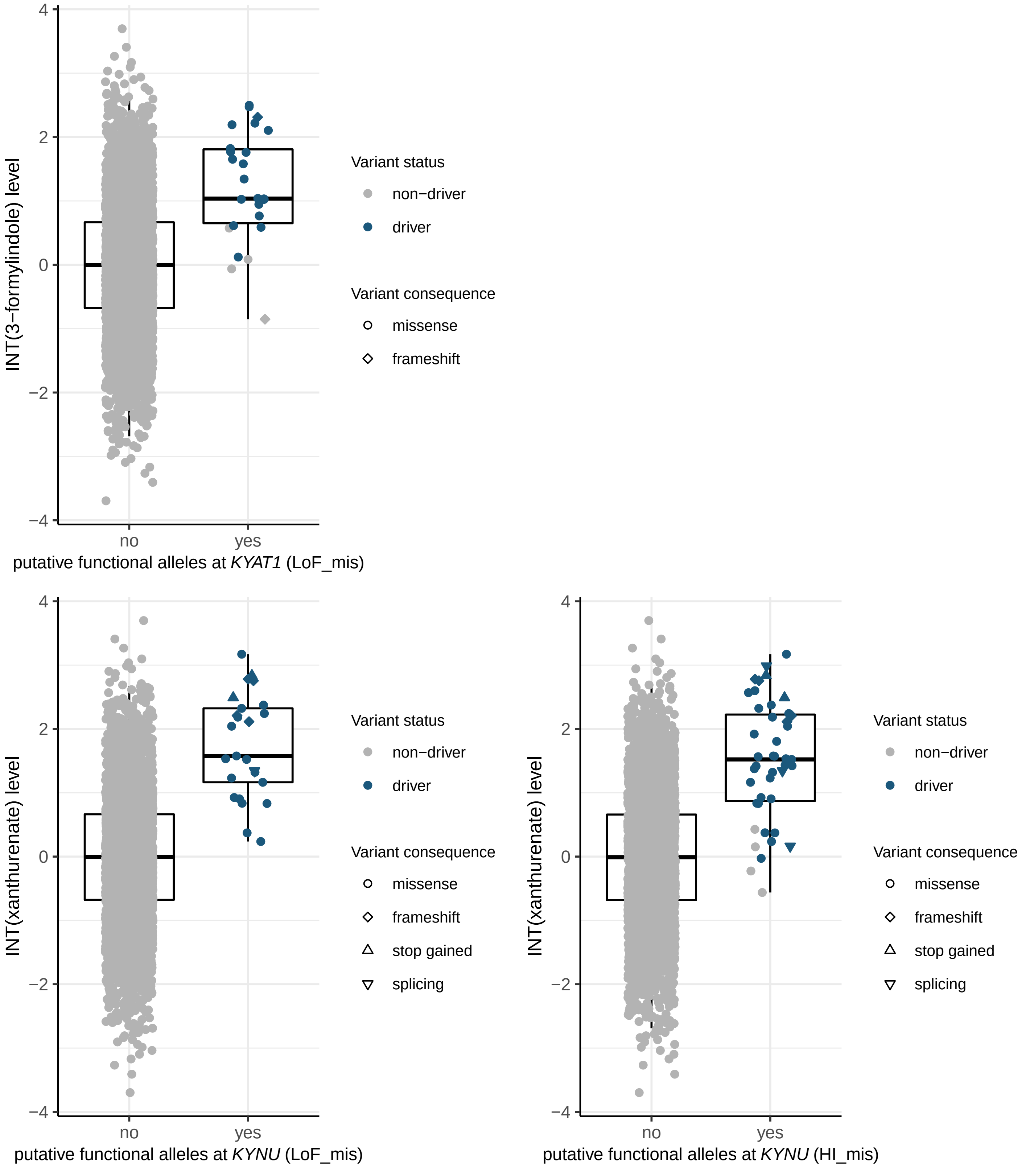
INT(3−formylindole) level
4
2
0
−2
−4
no
yes
putative functional alleles at *KYAT1* (LoF_mis)
Variant status
non−driver
driver
Variant consequence
missense
frameshift
INT(xanthurenate) level
putative functional alleles at *KYNU* (LoF_mis)
Variant status
non−driver
driver
Variant consequence
missense
frameshift
stop gained
splicing
INT(xanthurenate) level
putative functional alleles at *KYNU* (HI_mis)
Variant status
non−driver
driver
Variant consequence
missense
frameshift
stop gained
splicing

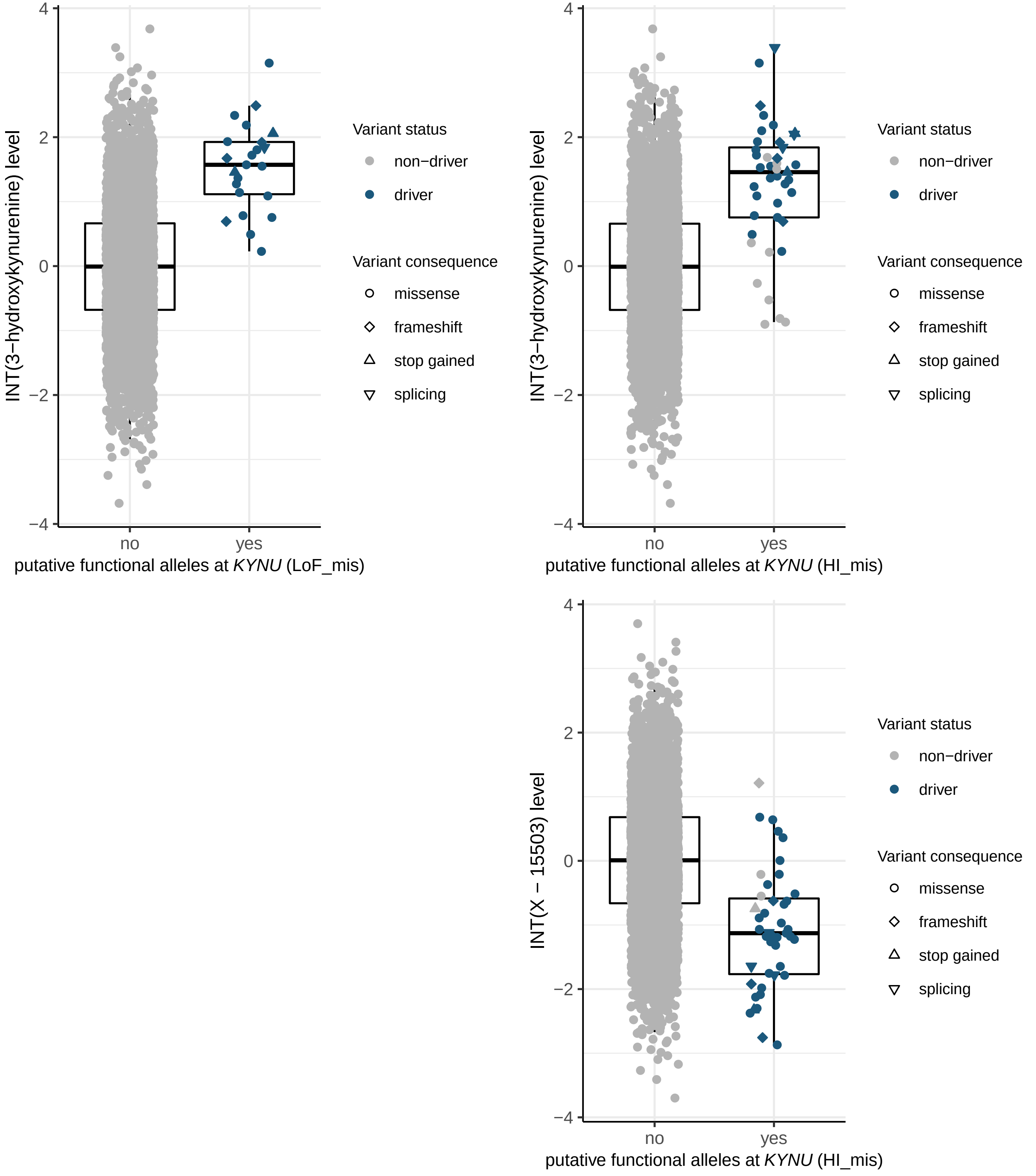

INT(3−hydroxykynurenine) level
putative functional alleles at KYNU (LoF_mis)
no
yes
Variant status
non−driver
driver
Variant consequence
missense
frameshift
stop gained
splicing
INT(3−hydroxykynurenine) level
putative functional alleles at KYNU (HI_mis)
INT(X − 15503) level
putative functional alleles at KYNU (HI_mis)

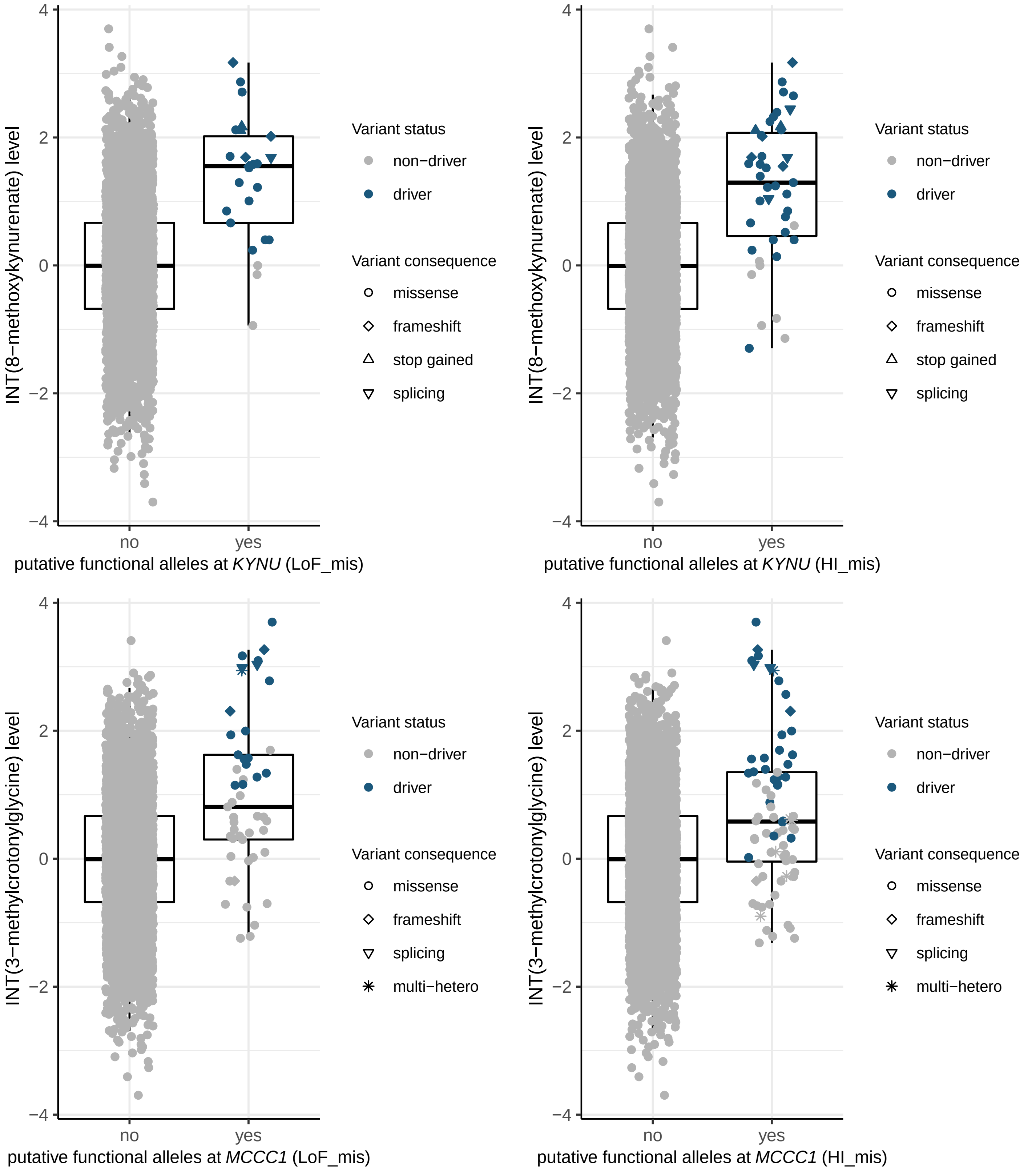

INT(8−methoxykynurenate) level
putative functional alleles at *KYNU* (LoF_mis)
putative functional alleles at *KYNU* (HI_mis)
INT(3−methylcrotonylglycine) level
putative functional alleles at *MCCC1* (LoF_mis)
putative functional alleles at *MCCC1* (HI_mis)
no
yes
4
2
0
−2
−4
Variant status
non−driver
driver
Variant consequence
missense
frameshift
stop gained
splicing
multi−hetero

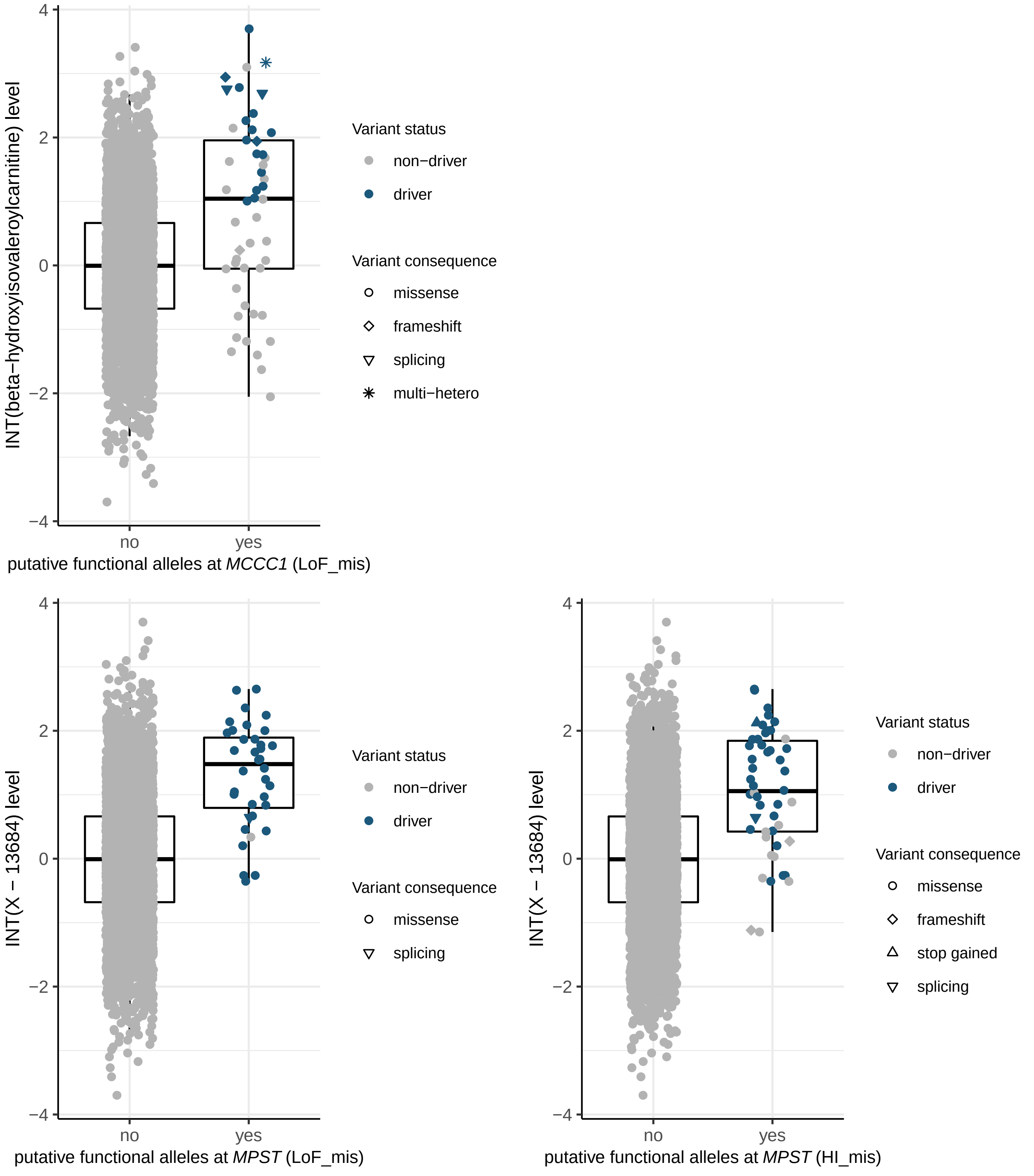
INT(beta−hydroxyisovaleroylcarnitine) level
4
2
0
−2
−4
no
yes
putative functional alleles at *MCCC1* (LoF_mis)
Variant status
non−driver
driver
Variant consequence
missense
frameshift
splicing
multi−hetero
INT(X − 13684) level
putative functional alleles at *MPST* (LoF_mis)
Variant status
non−driver
driver
Variant consequence
missense
splicing
INT(X − 13684) level
putative functional alleles at *MPST* (HI_mis)
Variant status
non−driver
driver
Variant consequence
missense
frameshift
stop gained
splicing

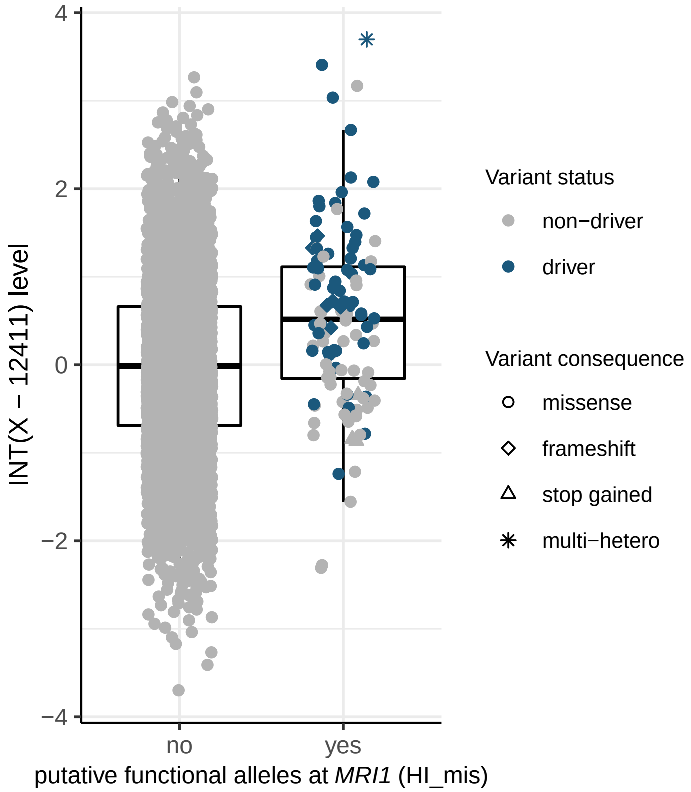

INT(X − 12411) level
4
2
0
−2
−4
no
yes
putative functional alleles at *MRI1* (HI_mis)
Variant status
non−driver
driver
Variant consequence
missense
frameshift
stop gained
multi−hetero


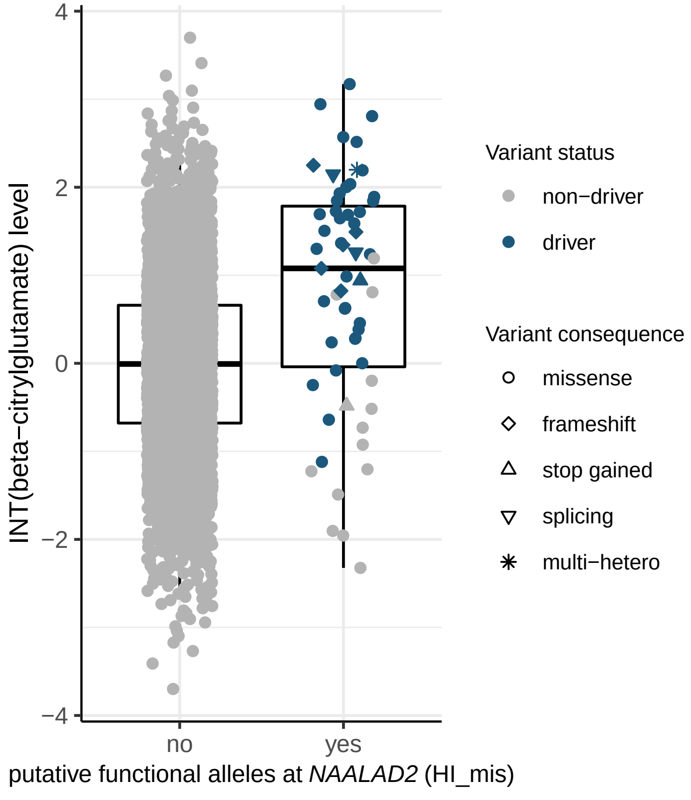

INT(beta−citrylglutamate) level
4
2
0
−2
−4
no
yes
putative functional alleles at *NAALAD2* (HI_mis)
Variant status
non−driver
driver
Variant consequence
missense
frameshift
stop gained
splicing
multi−hetero

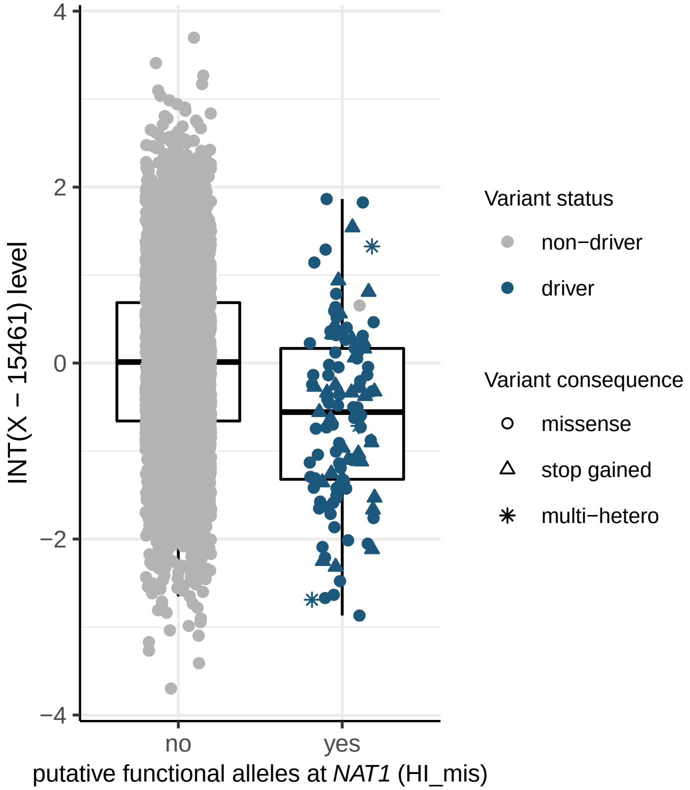
INT(X − 15461) level
4
2
0
−2
−4
no
yes
putative functional alleles at *NAT1* (HI_mis)
Variant status
non−driver
driver
Variant consequence
missense
stop gained
multi−hetero

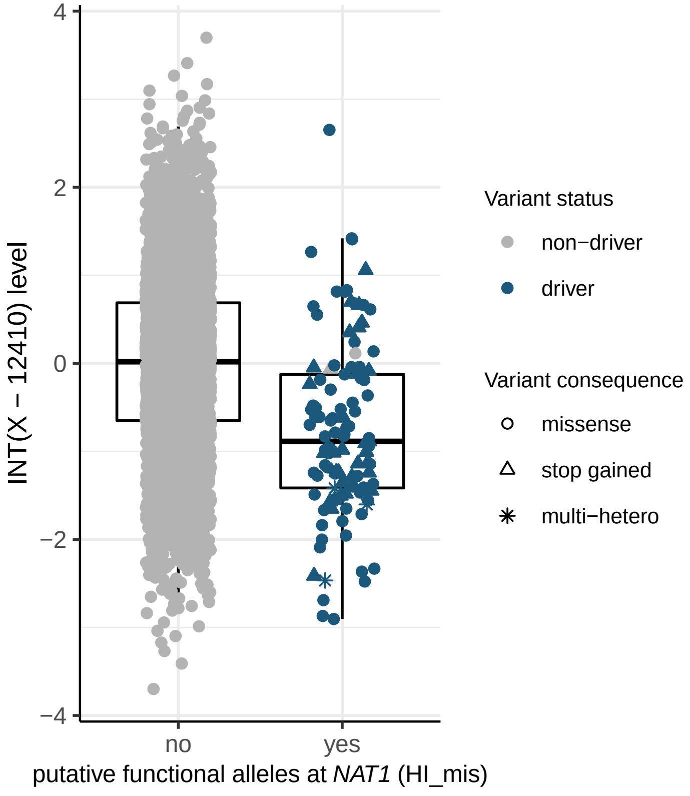
INT(X − 12410) level
4
2
0
−2
−4
no
yes
putative functional alleles at *NAT1* (HI_mis)
Variant status
non−driver
driver
Variant consequence
missense
stop gained
multi−hetero

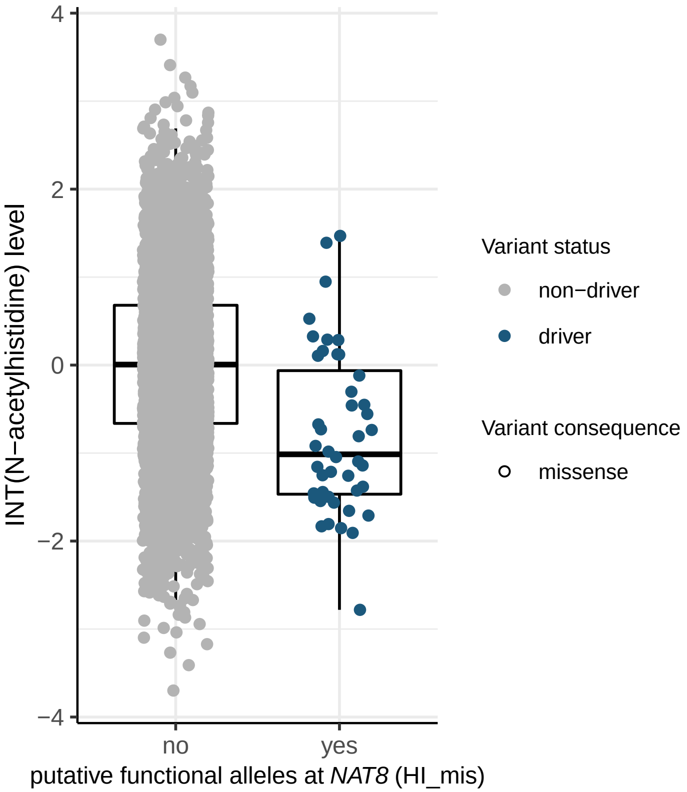
4
2
0
−2
−4
INT(N−acetylhistidine) level
no
yes
putative functional alleles at NAT8 (HI_mis)
Variant status
non−driver
driver
Variant consequence
missense

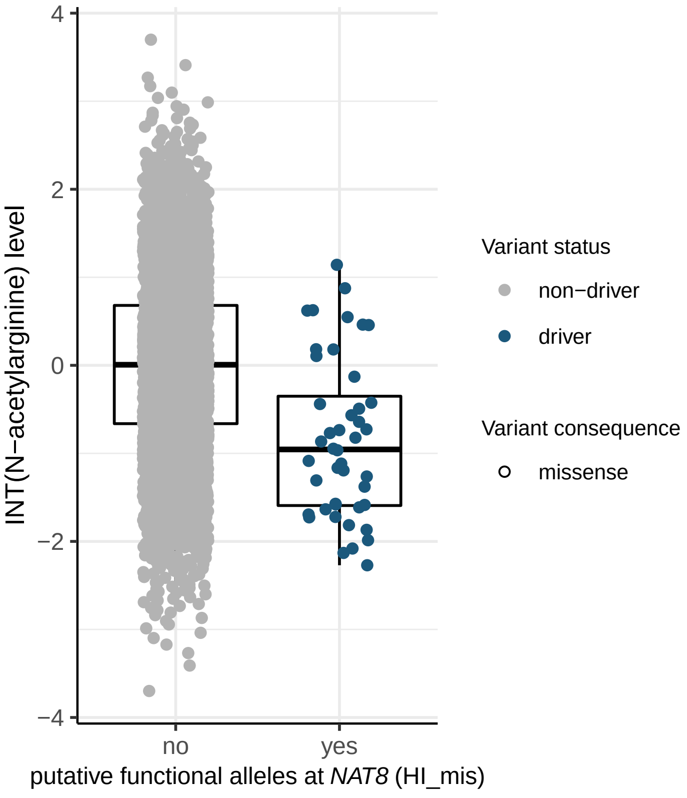
4
2
0
−2
−4
INT(N−acetylarginine) level
no
yes
putative functional alleles at NAT8 (HI_mis)
Variant status
non−driver
driver
Variant consequence
missense

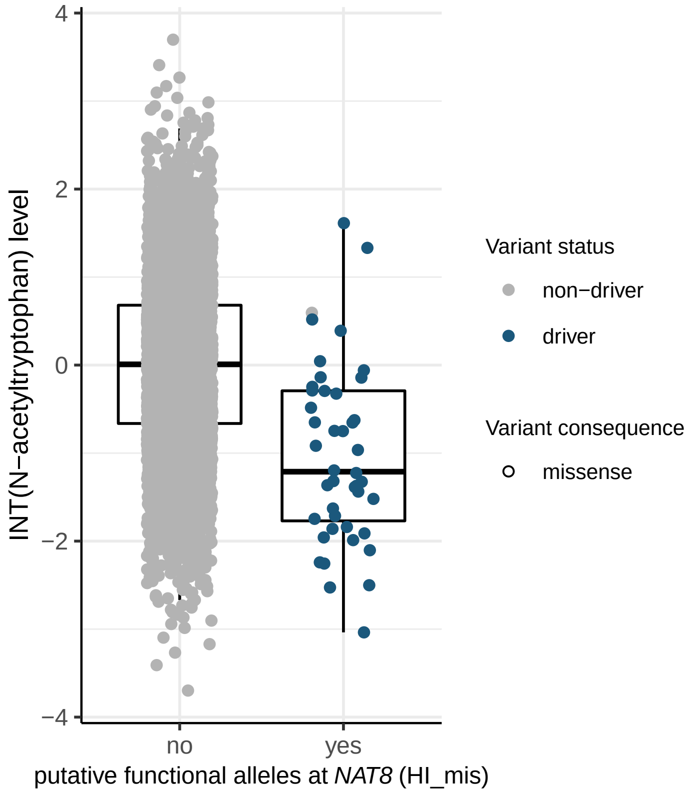

4
2
0
−2
−4
INT(N−acetyltryptophan) level
no
yes
putative functional alleles at *NAT8* (HI_mis)
Variant status
non−driver
driver
Variant consequence
missense


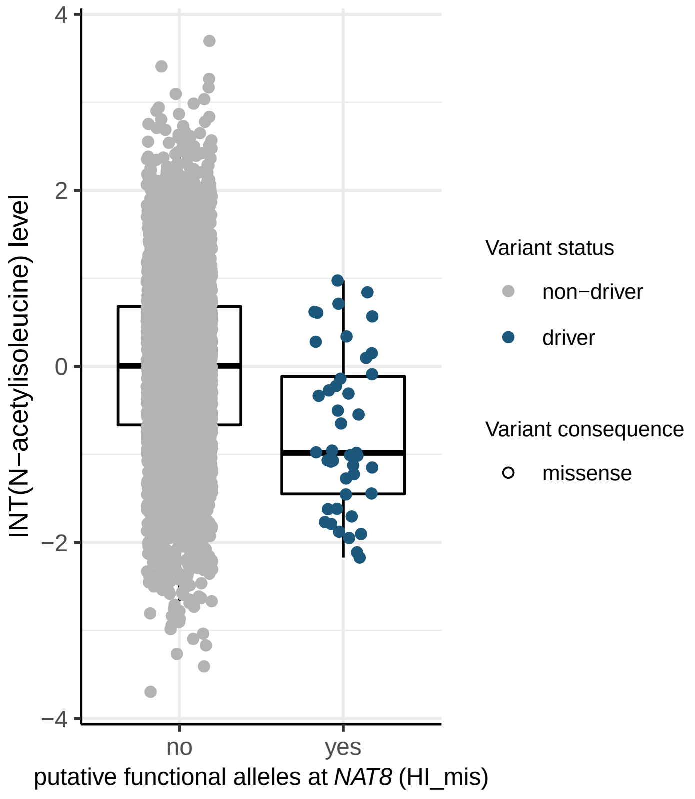

4
2
0
−2
−4
INT(N−acetylisoleucine) level
no
yes
putative functional alleles at *NAT8* (HI_mis)
Variant status
non−driver
driver
Variant consequence
missense

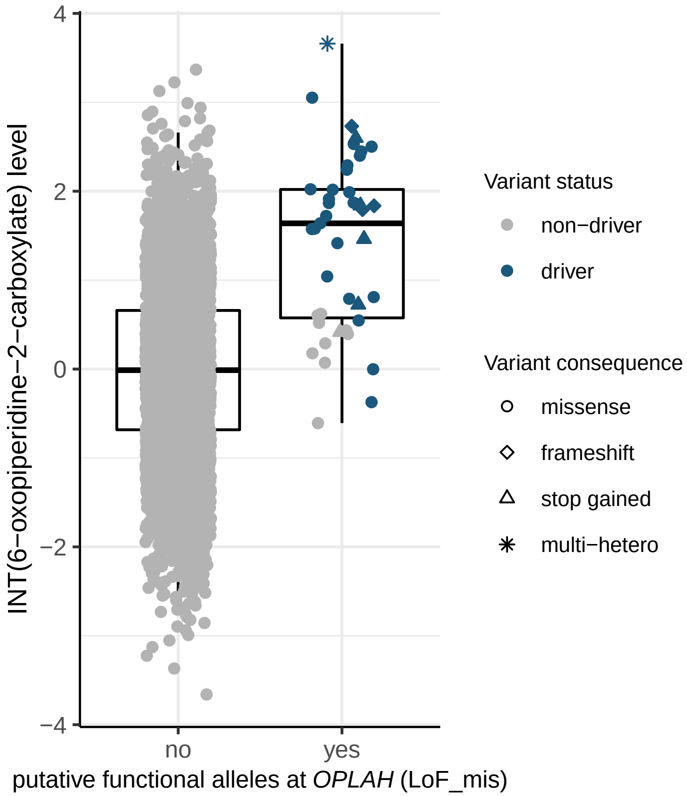

INT(6−oxopiperidine−2−carboxylate) level
4
2
0
−2
−4
no
yes
putative functional alleles at *OPLAH* (LoF_mis)
Variant status
non−driver
driver
Variant consequence
missense
frameshift
stop gained
multi−hetero


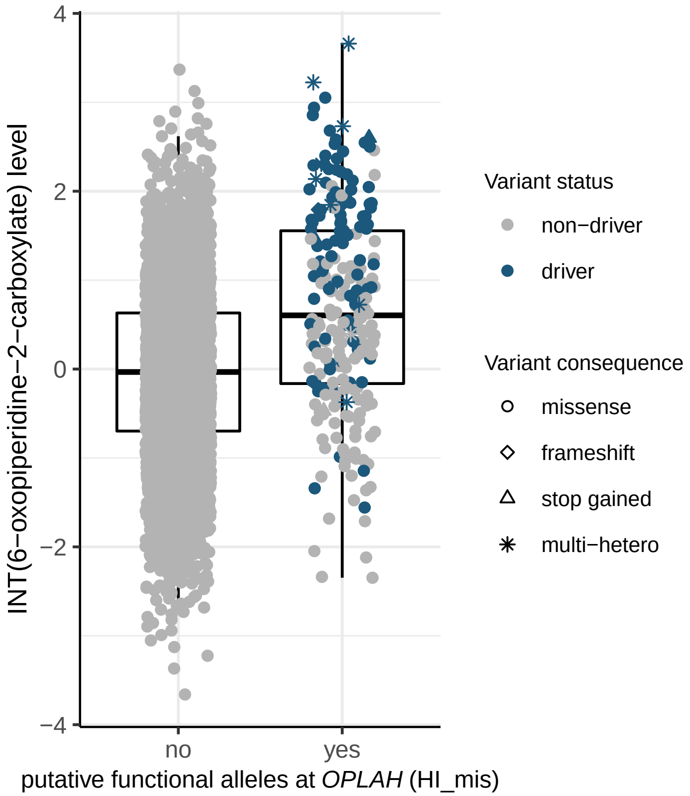

INT(6−oxopiperidine−2−carboxylate) level
4
2
0
−2
−4
no
yes
putative functional alleles at *OPLAH* (HI_mis)
Variant status
non−driver
driver
Variant consequence
missense
frameshift
stop gained
multi−hetero


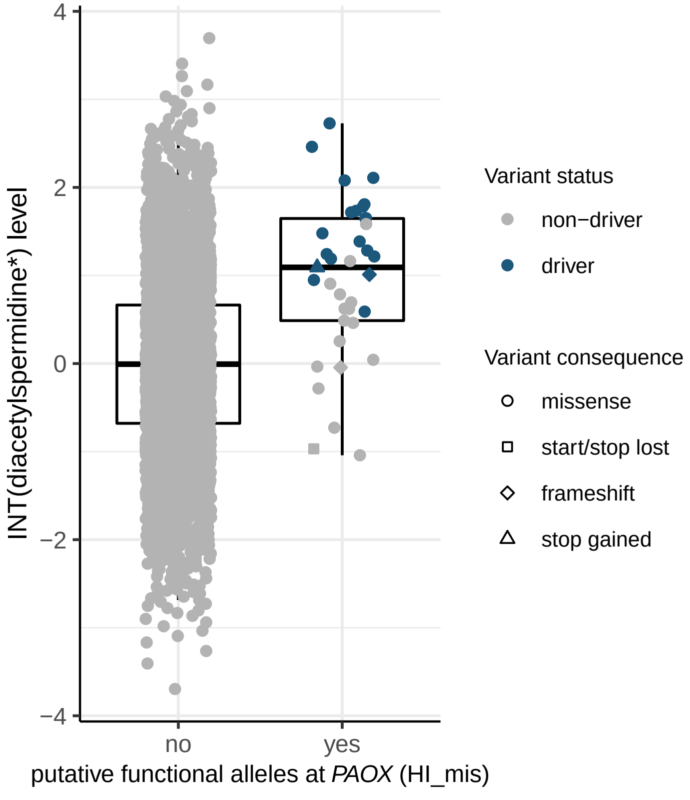

INT(diacetylspermidine*) level
4
2
0
−2
−4
no
yes
putative functional alleles at *PAOX* (HI_mis)
Variant status
non−driver
driver
Variant consequence
missense
start/stop lost
frameshift
stop gained

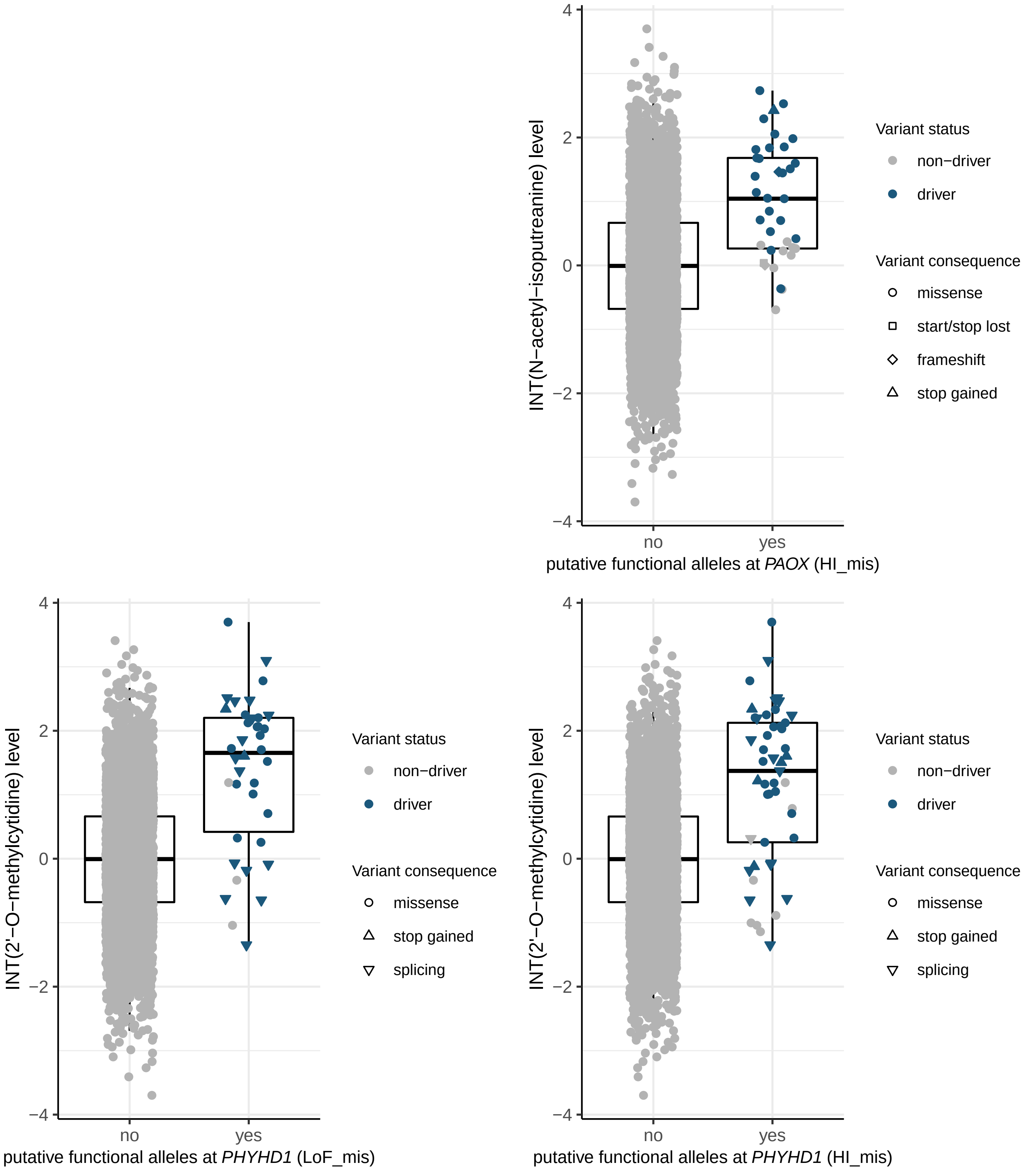
INT(N−acetyl−isoputreanine) level
4
2
0
−2
−4
no
yes
putative functional alleles at *PAOX* (HI_mis)
Variant status
non−driver
driver
Variant consequence
missense
start/stop lost
frameshift
stop gained
INT(2'−O−methylcytidine) level
4
2
0
−2
−4
no
yes
putative functional alleles at *PHYHD1* (LoF_mis)
Variant status
non−driver
driver
Variant consequence
missense
stop gained
splicing
INT(2'−O−methylcytidine) level
4
2
0
−2
−4
no
yes
putative functional alleles at *PHYHD1* (HI_mis)
Variant status
non−driver
driver
Variant consequence
missense
stop gained
splicing

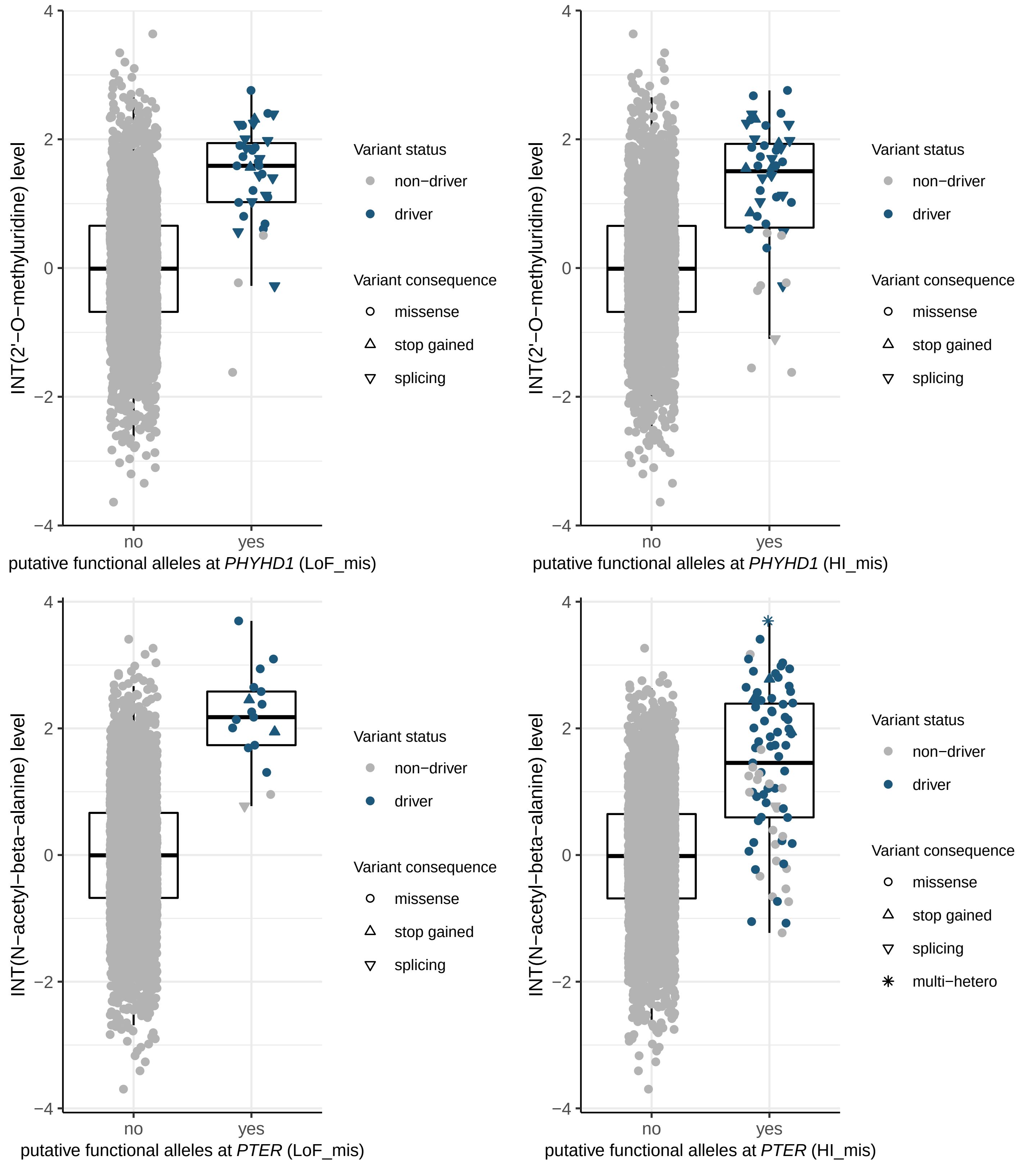
INT(2'−O−methyluridine) level
putative functional alleles at *PHYHD1* (LoF_mis)
no
yes
Variant status
non−driver
driver
Variant consequence
missense
stop gained
splicing
INT(2'−O−methyluridine) level
putative functional alleles at *PHYHD1* (HI_mis)
INT(N−acetyl−beta−alanine) level
putative functional alleles at *PTER* (LoF_mis)
INT(N−acetyl−beta−alanine) level
putative functional alleles at *PTER* (HI_mis)
multi−hetero
4
2
0
−2
−4

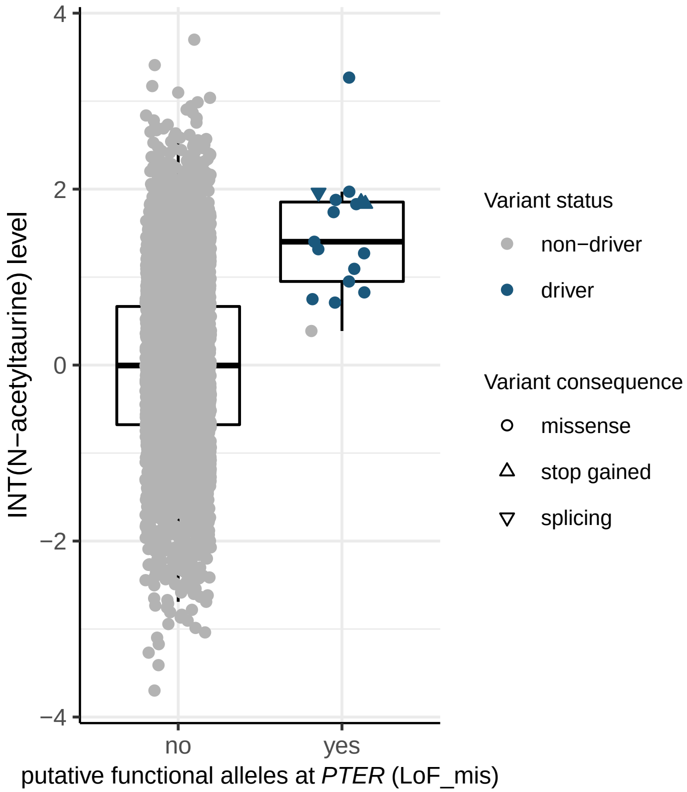
INT(N−acetyltaurine) level
putative functional alleles at PTER (LoF_mis)
no
yes
Variant status
non−driver
driver
Variant consequence
missense
stop gained
splicing

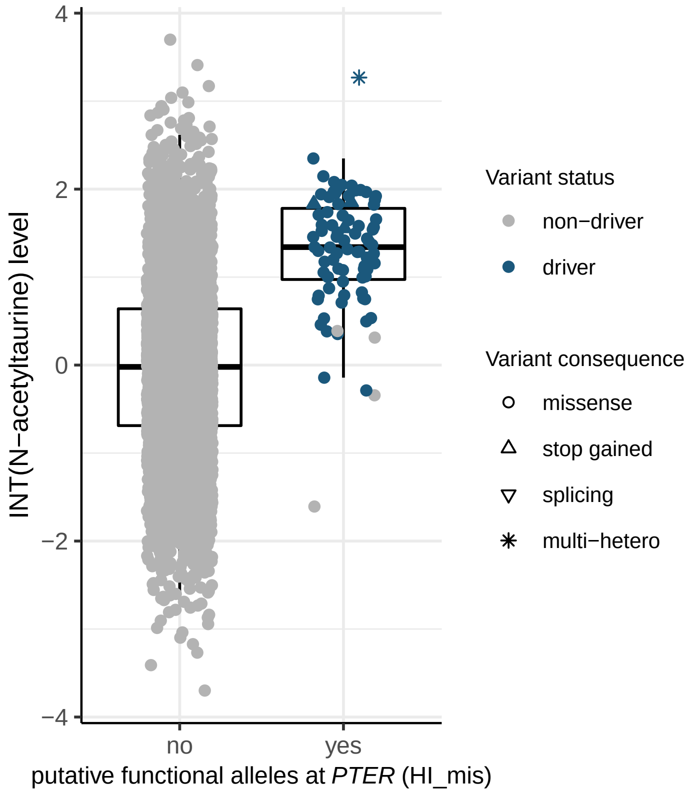
INT(N−acetyltaurine) level
putative functional alleles at PTER (HI_mis)
no
yes
Variant status
non−driver
driver
Variant consequence
missense
stop gained
splicing
multi−hetero

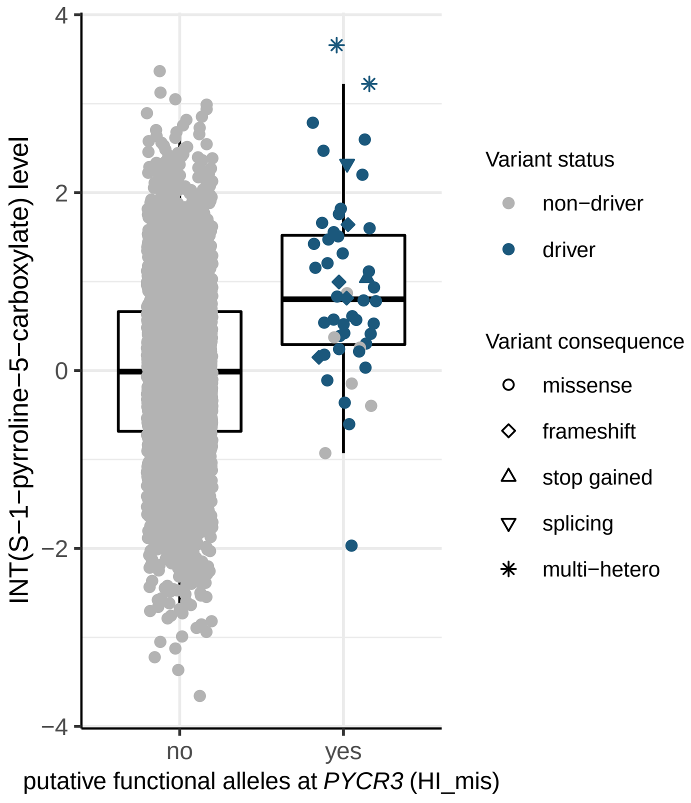
INT(S−1−pyrroline−5−carboxylate) level
putative functional alleles at PYCR3 (HI_mis)
no
yes
Variant status
non−driver
driver
Variant consequence
missense
frameshift
stop gained
splicing
multi−hetero

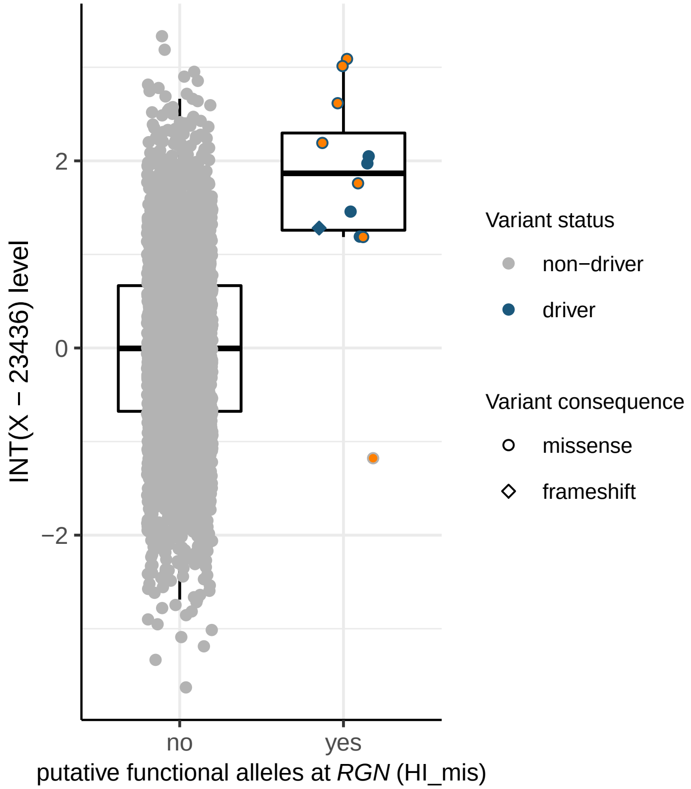
INT(X − 23436) level
2
0
−2
no
yes
putative functional alleles at *RGN* (HI_mis)
Variant status
non−driver
driver
Variant consequence
missense
frameshift

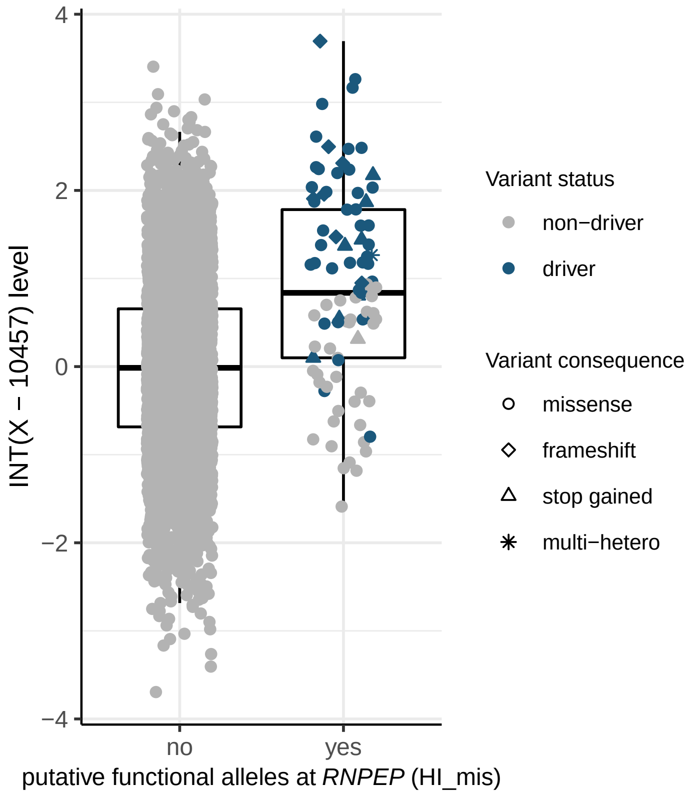
INT(X − 10457) level
4
2
0
−2
−4
no
yes
putative functional alleles at *RNPEP* (HI_mis)
Variant status
non−driver
driver
Variant consequence
missense
frameshift
stop gained
multi−hetero

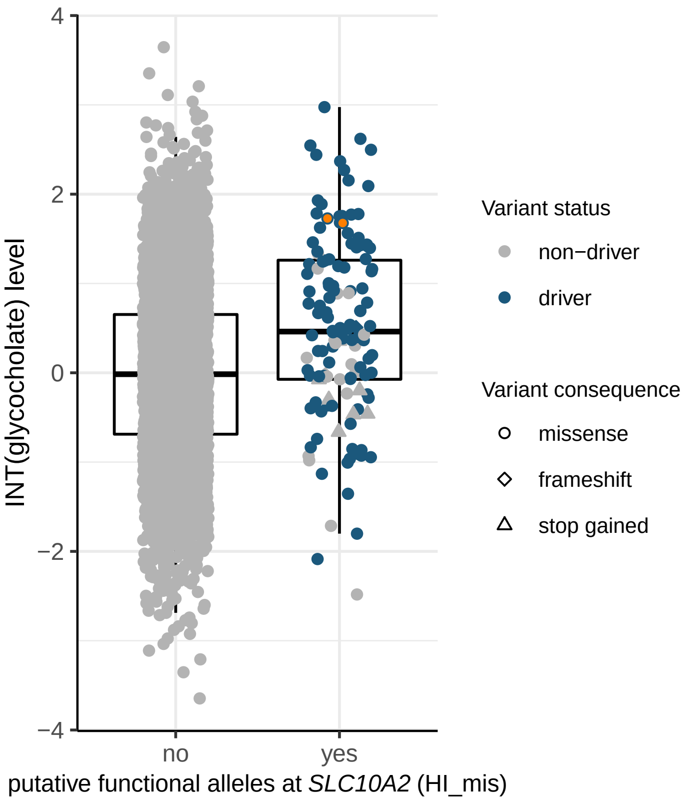
4
2
0
−2
−4
INT(glycocholate) level
no
yes
putative functional alleles at *SLC10A2* (HI_mis)
Variant status
non−driver
driver
Variant consequence
missense
frameshift
stop gained

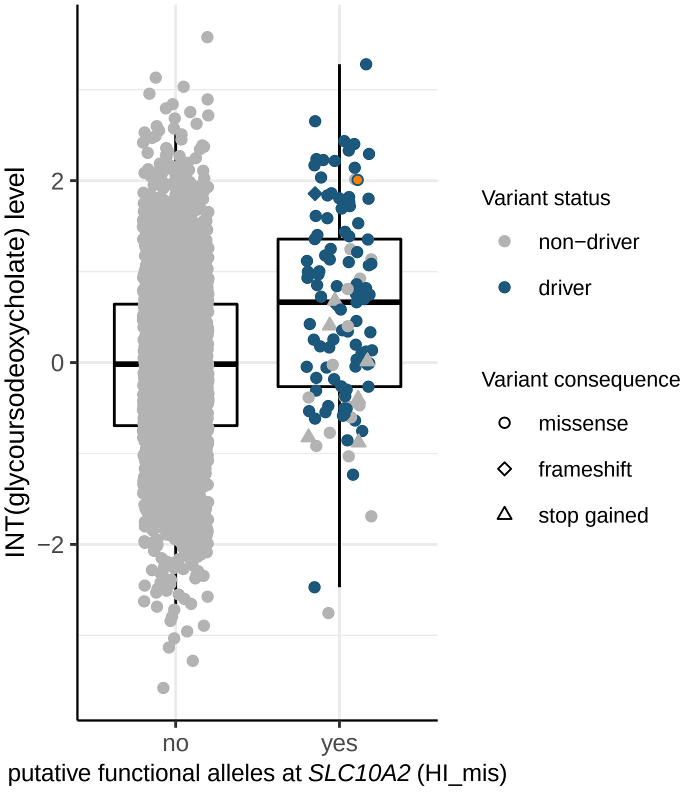
2
0
−2
INT(glycoursodeoxycholate) level
no
yes
putative functional alleles at *SLC10A2* (HI_mis)
Variant status
non−driver
driver
Variant consequence
missense
frameshift
stop gained

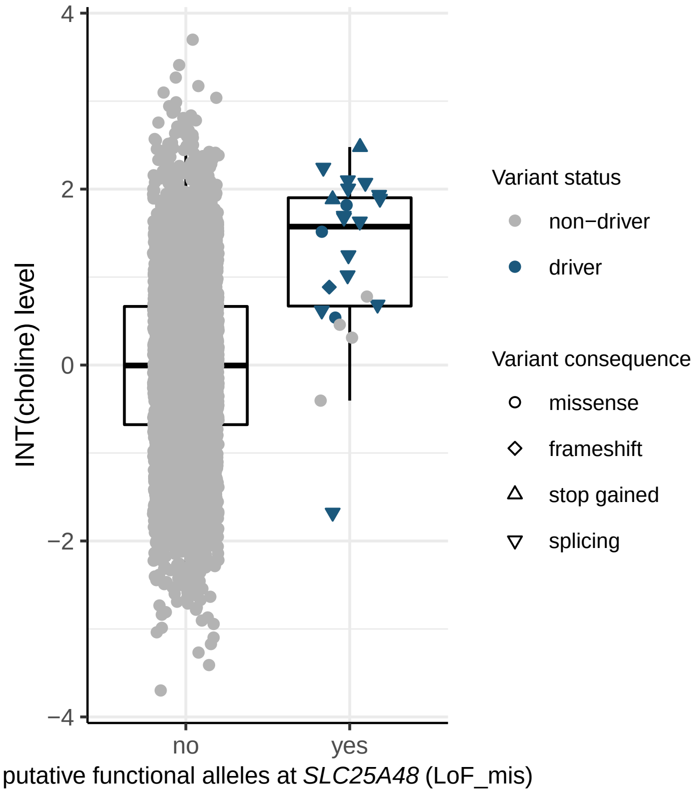

INT(choline) level
4
2
0
−2
−4
no
yes
putative functional alleles at *SLC25A48* (LoF_mis)
Variant status
non−driver
driver
Variant consequence
missense
frameshift
stop gained
splicing


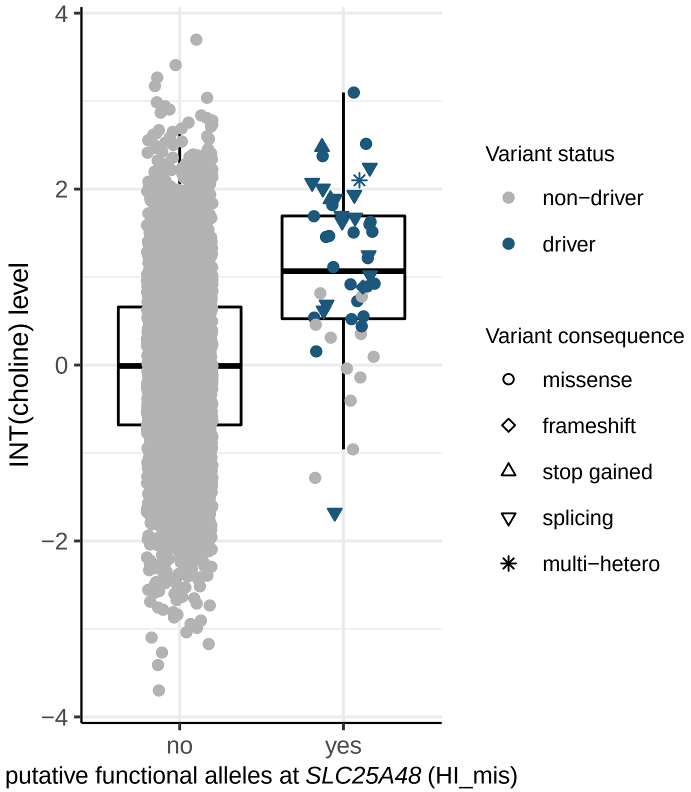

INT(choline) level
4
2
0
−2
−4
no
yes
putative functional alleles at *SLC25A48* (HI_mis)
Variant status
non−driver
driver
Variant consequence
missense
frameshift
stop gained
splicing
multi−hetero


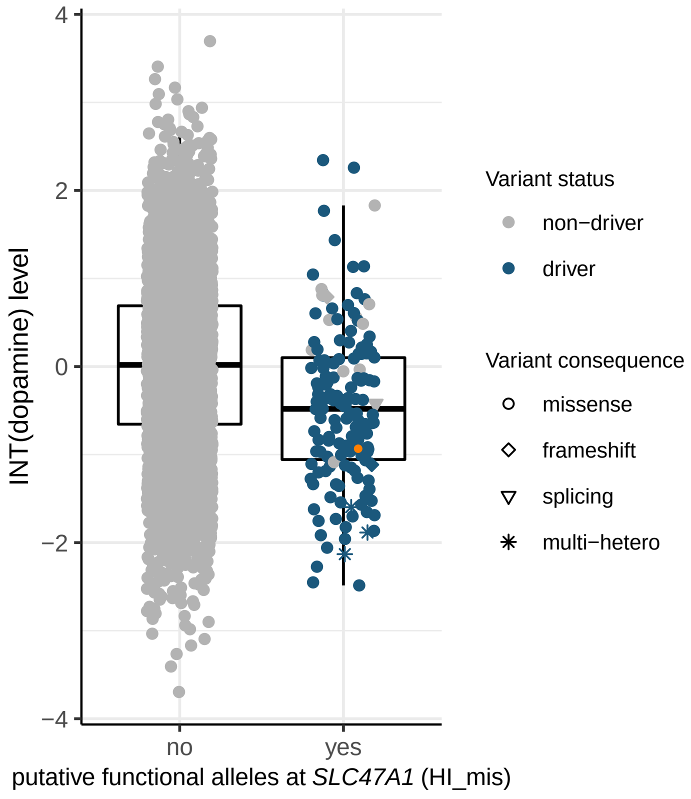

INT(dopamine) level
4
2
0
−2
−4
no
yes
putative functional alleles at *SLC47A1* (HI_mis)
Variant status
non−driver
driver
Variant consequence
missense
frameshift
splicing
multi−hetero

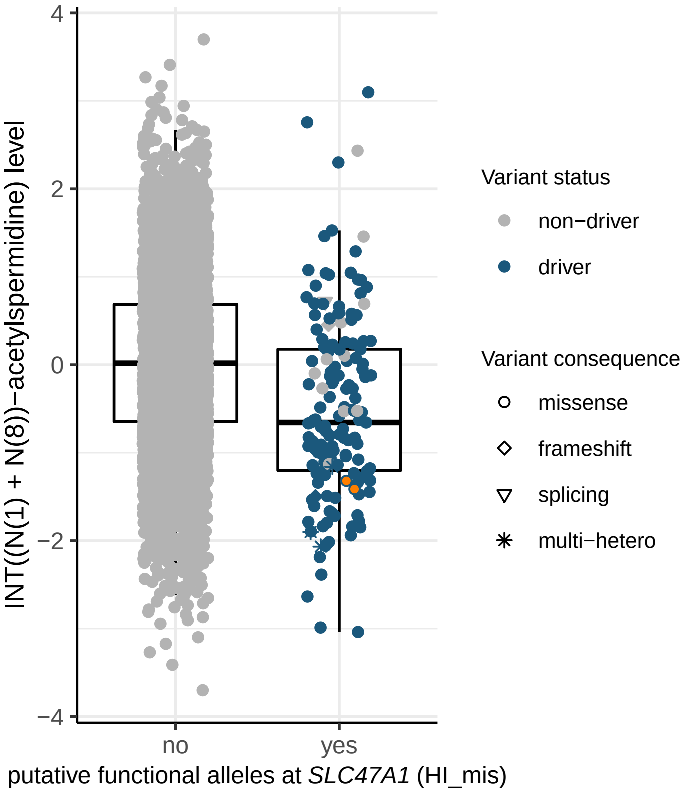

INT((N(1) + N(8))−acetylspermidine) level
4
2
0
−2
−4
no
yes
putative functional alleles at *SLC47A1* (HI_mis)
Variant status
non−driver
driver
Variant consequence
missense
frameshift
splicing
multi−hetero


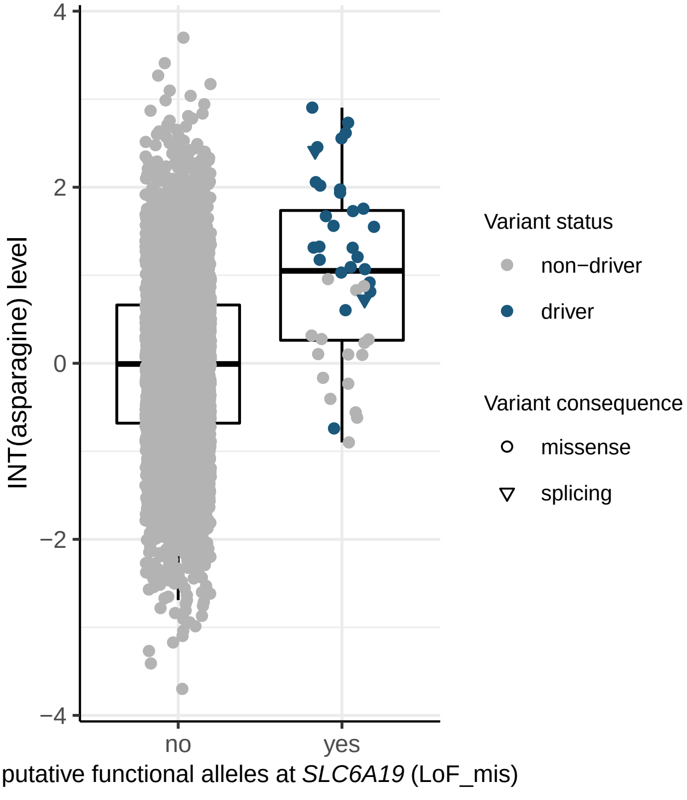

INT(asparagine) level
4
2
0
−2
−4
no
yes
putative functional alleles at *SLC6A19* (LoF_mis)
Variant status
non−driver
driver
Variant consequence
missense
splicing


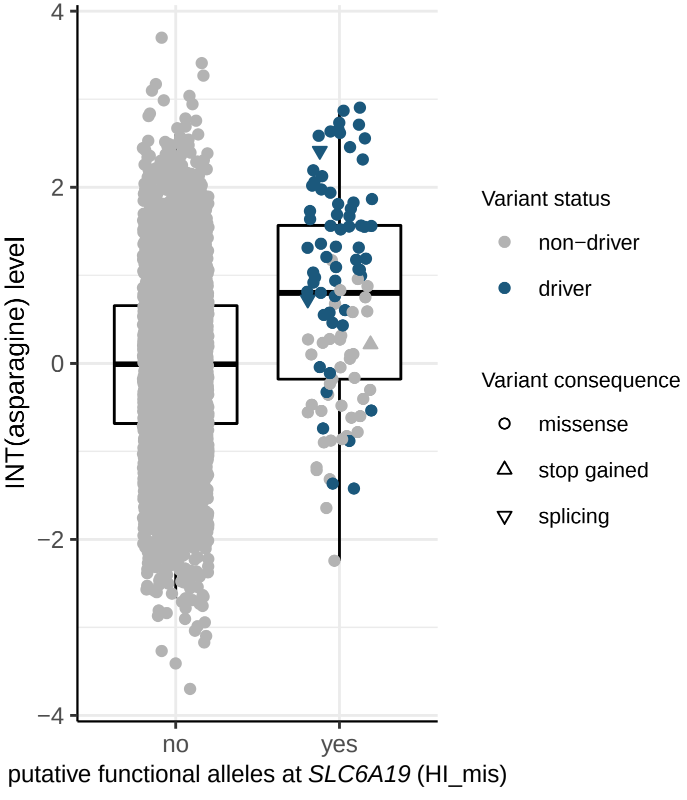

INT(asparagine) level
4
2
0
−2
−4
no
yes
putative functional alleles at *SLC6A19* (HI_mis)
Variant status
non−driver
driver
Variant consequence
missense
stop gained
splicing

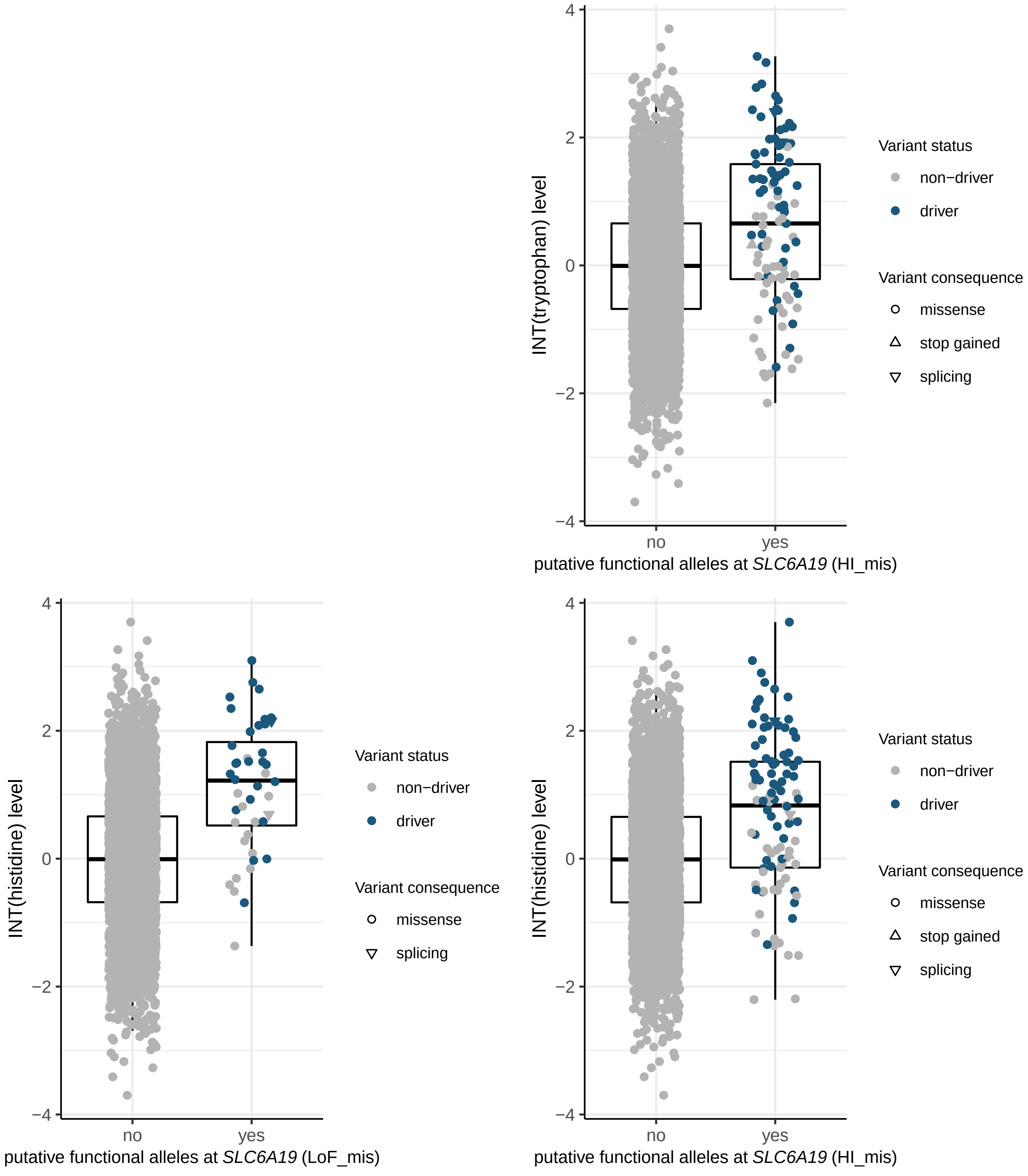
INT(tryptophan) level
putative functional alleles at SLC6A19 (HI_mis)
Variant status
non−driver
driver
Variant consequence
missense
stop gained
splicing
INT(histidine) level
putative functional alleles at SLC6A19 (LoF_mis)
Variant status
non−driver
driver
Variant consequence
missense
splicing
INT(histidine) level
putative functional alleles at SLC6A19 (HI_mis)
Variant status
non−driver
driver
Variant consequence
missense
stop gained
splicing
no
yes
4
2
0
−2
−4

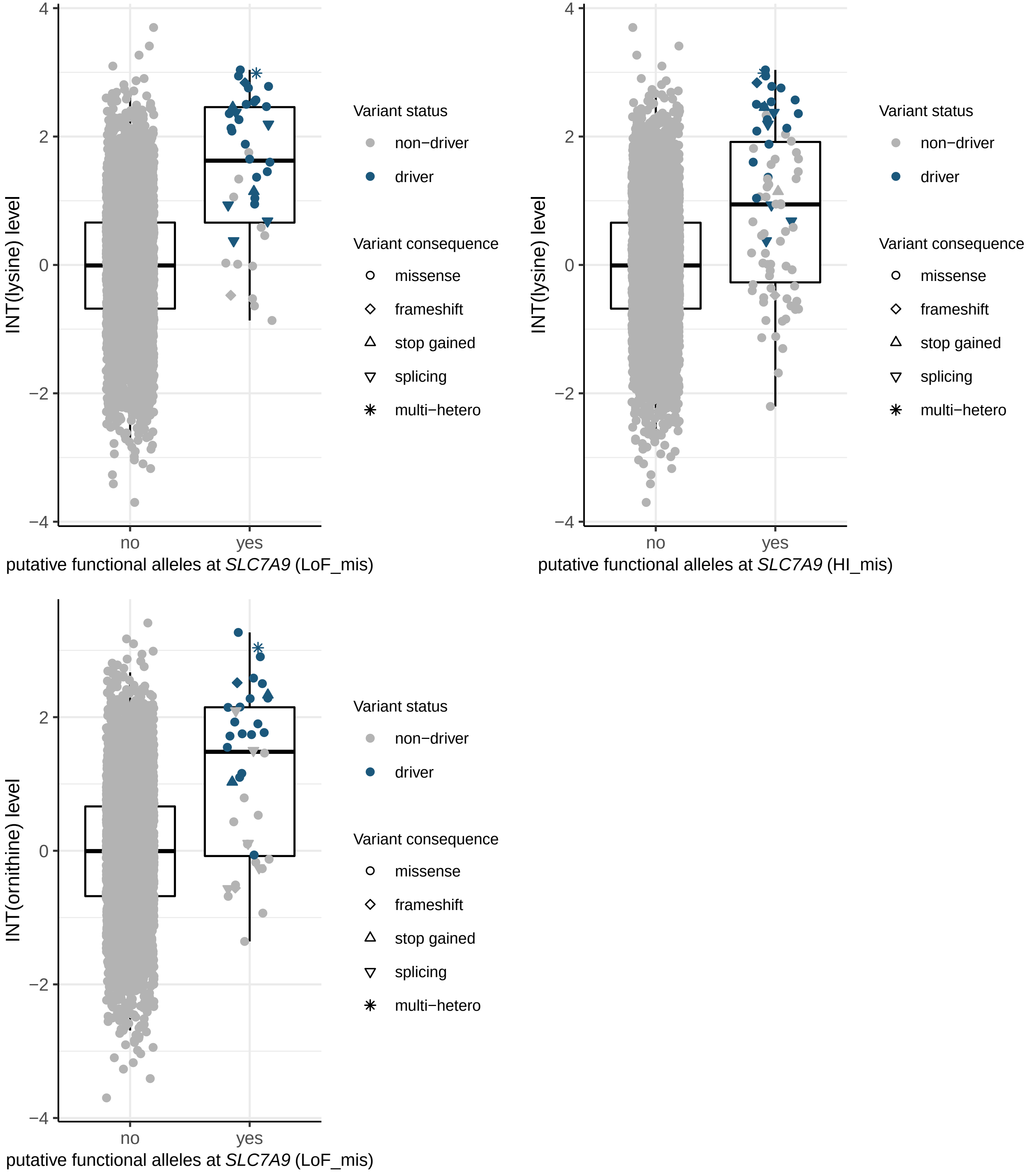

INT(lysine) level
putative functional alleles at SLC7A9 (LoF_mis)
INT(lysine) level
putative functional alleles at SLC7A9 (HI_mis)
INT(ornithine) level
putative functional alleles at SLC7A9 (LoF_mis)
no
yes
Variant status
non−driver
driver
Variant consequence
missense
frameshift
stop gained
splicing
multi−hetero
4
2
0
−2
−4

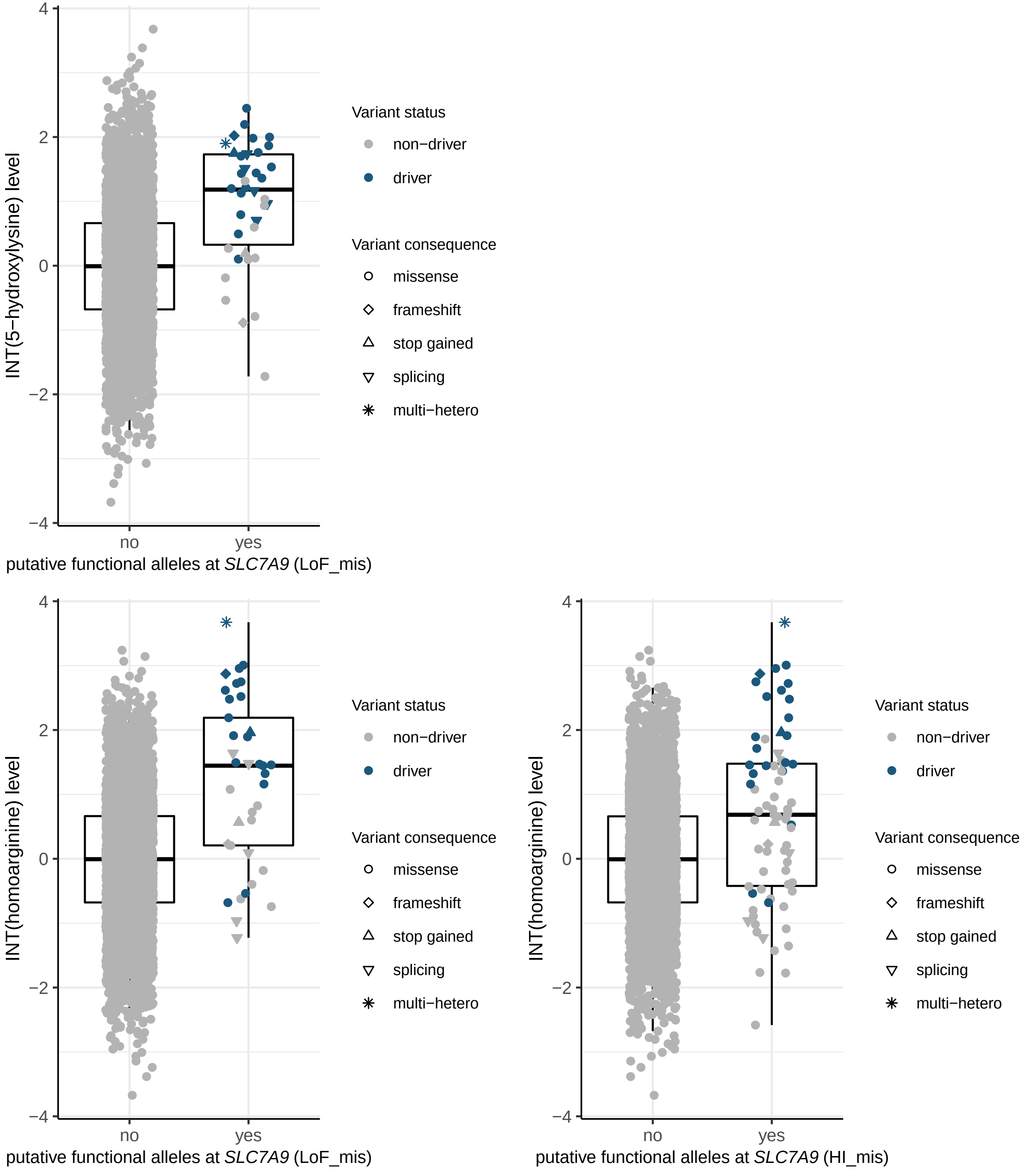

INT(5−hydroxylysine) level
putative functional alleles at SLC7A9 (LoF_mis)
INT(homoarginine) level
putative functional alleles at SLC7A9 (LoF_mis)
INT(homoarginine) level
putative functional alleles at SLC7A9 (HI_mis)
no
yes
Variant status
non−driver
driver
Variant consequence
missense
frameshift
stop gained
splicing
multi−hetero

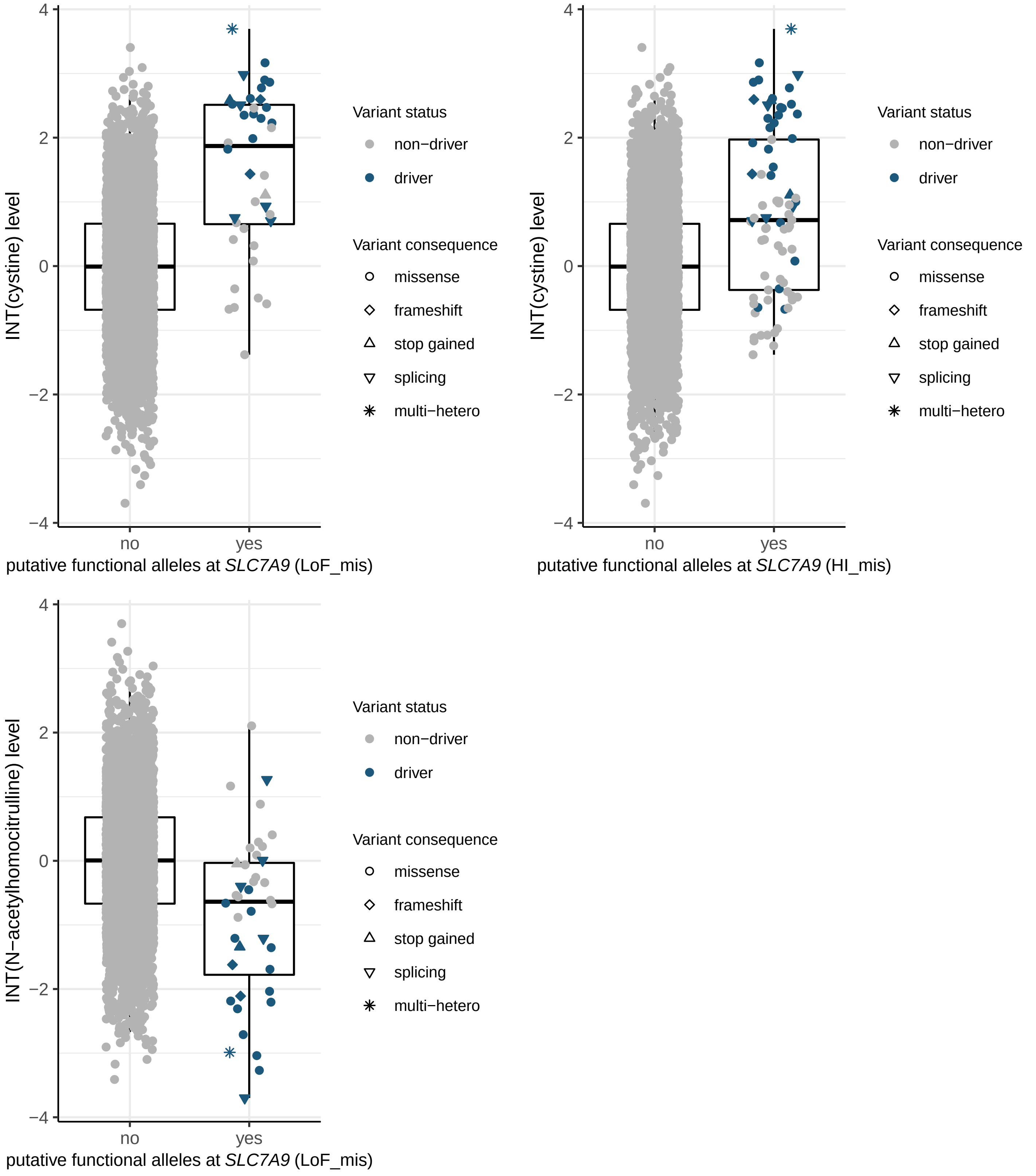

INT(cystine) level
putative functional alleles at *SLC7A9* (LoF_mis)
putative functional alleles at *SLC7A9* (HI_mis)
INT(N−acetylhomocitrulline) level
no
yes
4
2
0
−2
−4
Variant status
non−driver
driver
Variant consequence
missense
frameshift
stop gained
splicing
multi−hetero

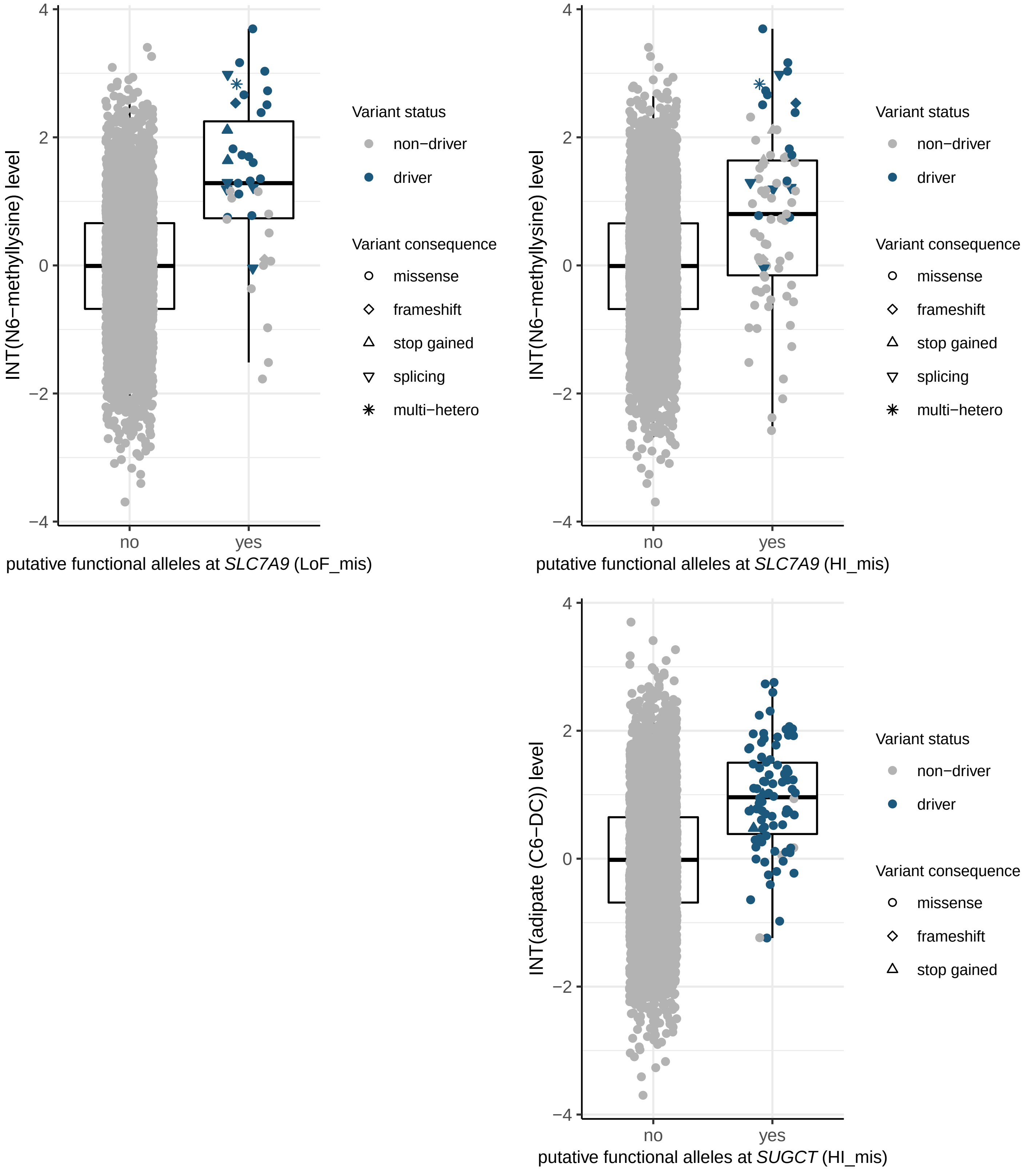

INT(N6−methyllysine) level
putative functional alleles at SLC7A9 (LoF_mis)
INT(N6−methyllysine) level
putative functional alleles at SLC7A9 (HI_mis)
INT(adipate (C6−DC)) level
putative functional alleles at SUGCT (HI_mis)
no
yes
4
2
0
−2
−4
Variant status
non−driver
driver
Variant consequence
missense
frameshift
stop gained
splicing
multi−hetero

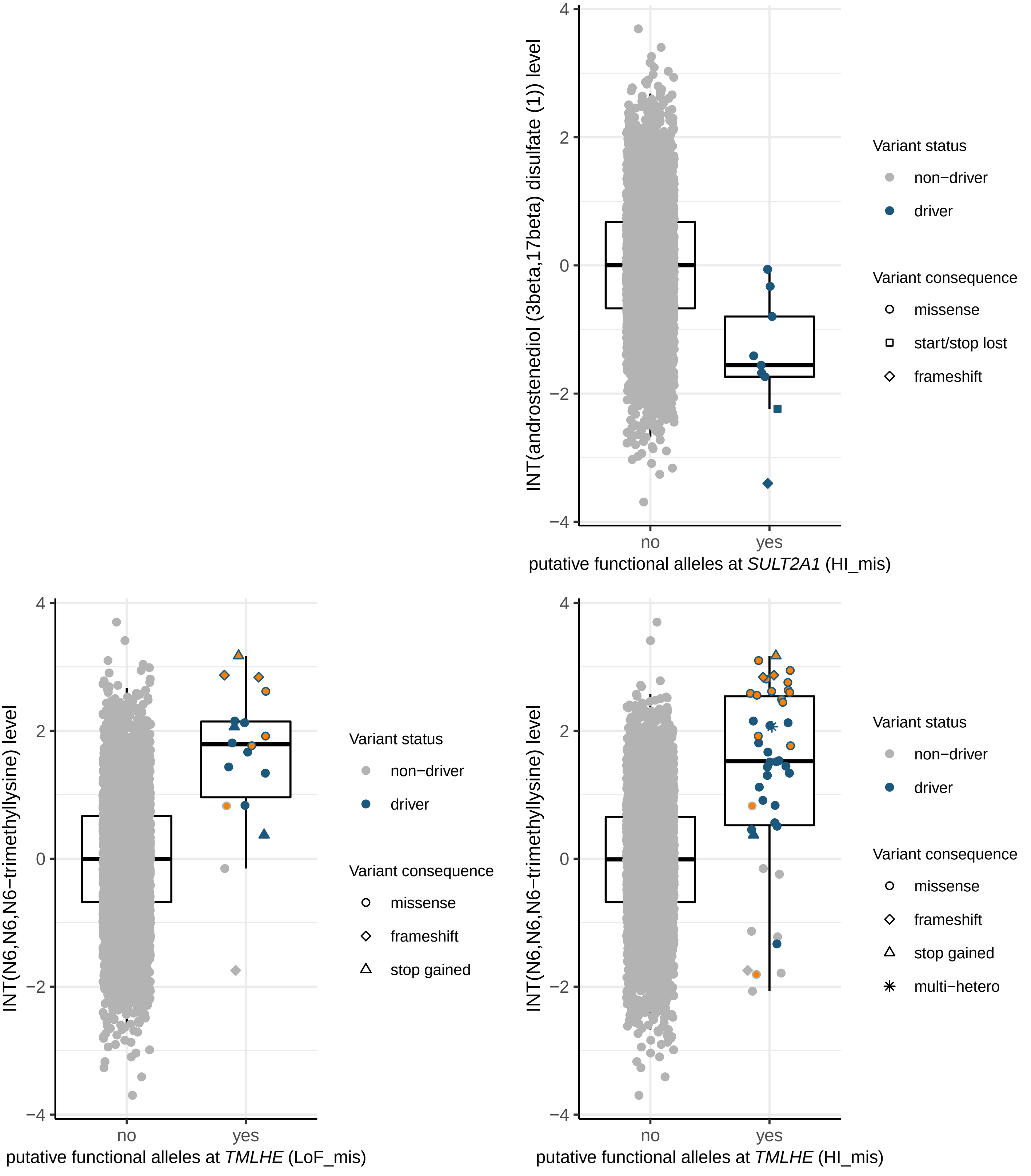

INT(androstenediol (3beta,17beta) disulfate (1)) level
4
2
0
−2
−4
no
yes
putative functional alleles at *SULT2A1* (HI_mis)
Variant status
non−driver
driver
Variant consequence
missense
start/stop lost
frameshift
INT(N6,N6,N6−trimethyllysine) level
4
2
0
−2
−4
no
yes
putative functional alleles at *TMLHE* (LoF_mis)
Variant status
non−driver
driver
Variant consequence
missense
frameshift
stop gained
INT(N6,N6,N6−trimethyllysine) level
4
2
0
−2
−4
no
yes
putative functional alleles at *TMLHE* (HI_mis)
Variant status
non−driver
driver
Variant consequence
missense
frameshift
stop gained
multi−hetero

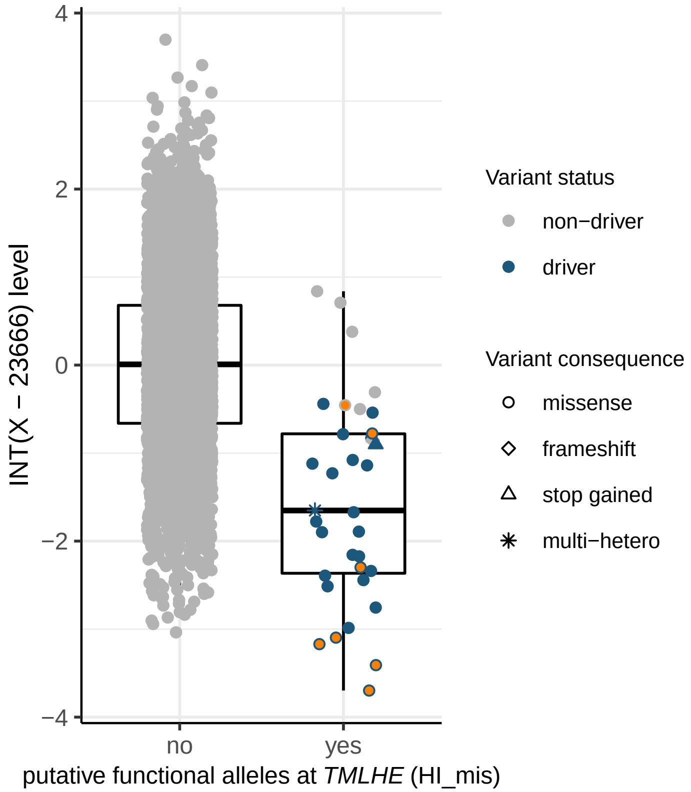

4
2
0
−2
−4
INT(X − 23666) level
no
yes
putative functional alleles at TMLHE (HI_mis)
Variant status
non−driver
driver
Variant consequence
missense
frameshift
stop gained
multi−hetero


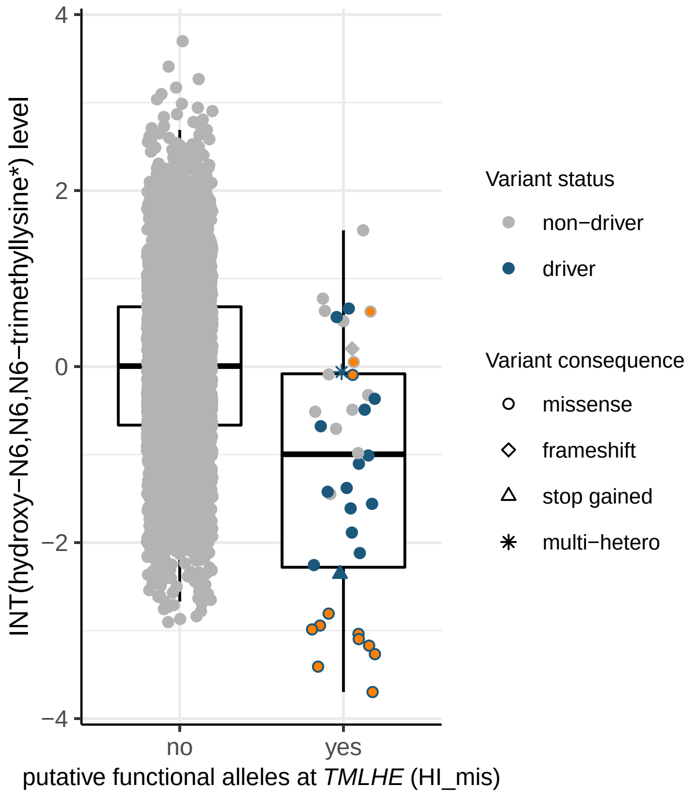

4
2
0
−2
−4
INT(hydroxy−N6,N6,N6−trimethyllysine*) level
no
yes
putative functional alleles at TMLHE (HI_mis)
Variant status
non−driver
driver
Variant consequence
missense
frameshift
stop gained
multi−hetero

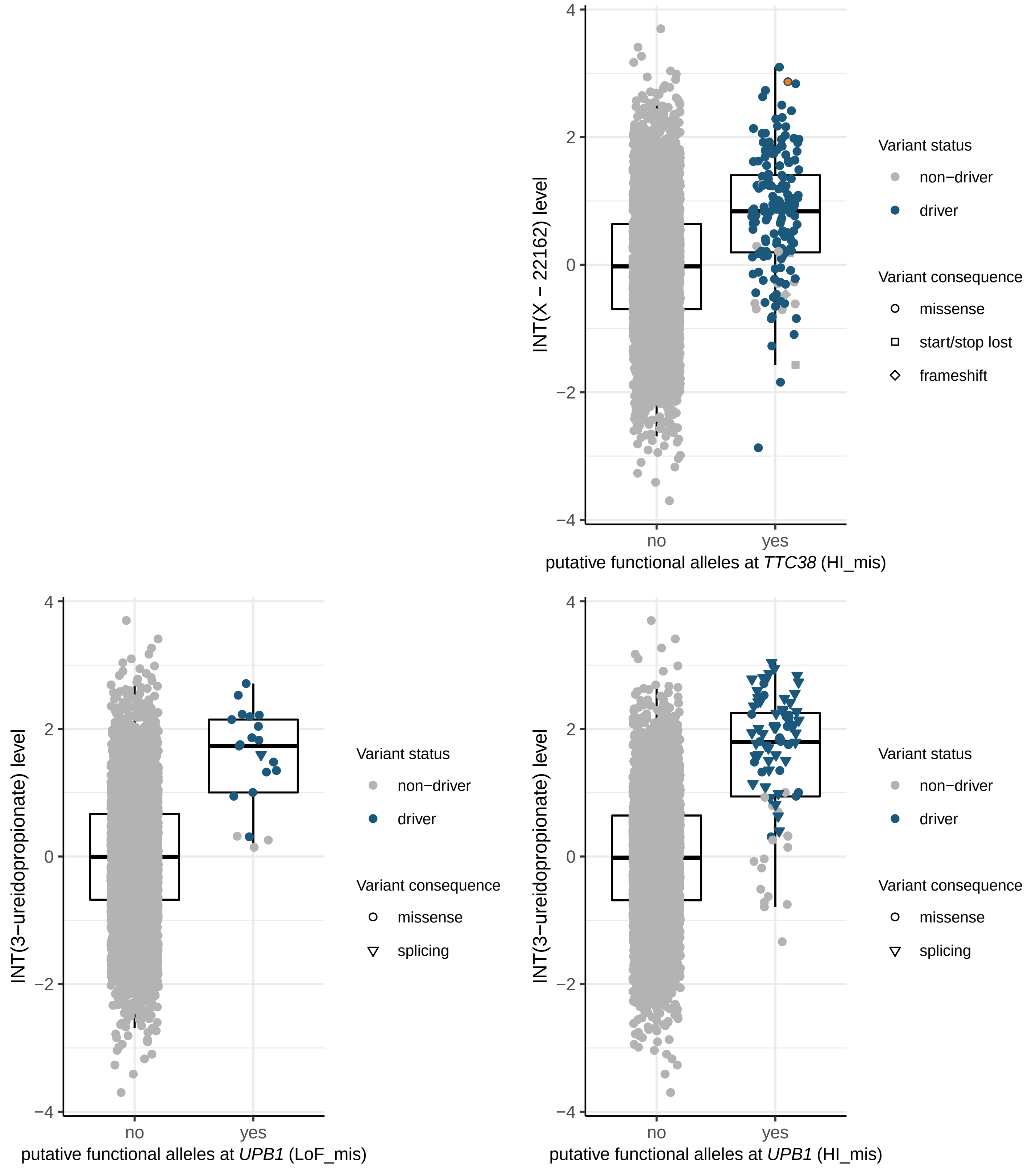

INT(X − 22162) level
putative functional alleles at TTC38 (HI_mis)
no
yes
Variant status
non−driver
driver
Variant consequence
missense
start/stop lost
frameshift
INT(3−ureidopropionate) level
putative functional alleles at UPB1 (LoF_mis)
Variant status
non−driver
driver
Variant consequence
missense
splicing
INT(3−ureidopropionate) level
putative functional alleles at UPB1 (HI_mis)
Variant status
non−driver
driver
Variant consequence
missense
splicing

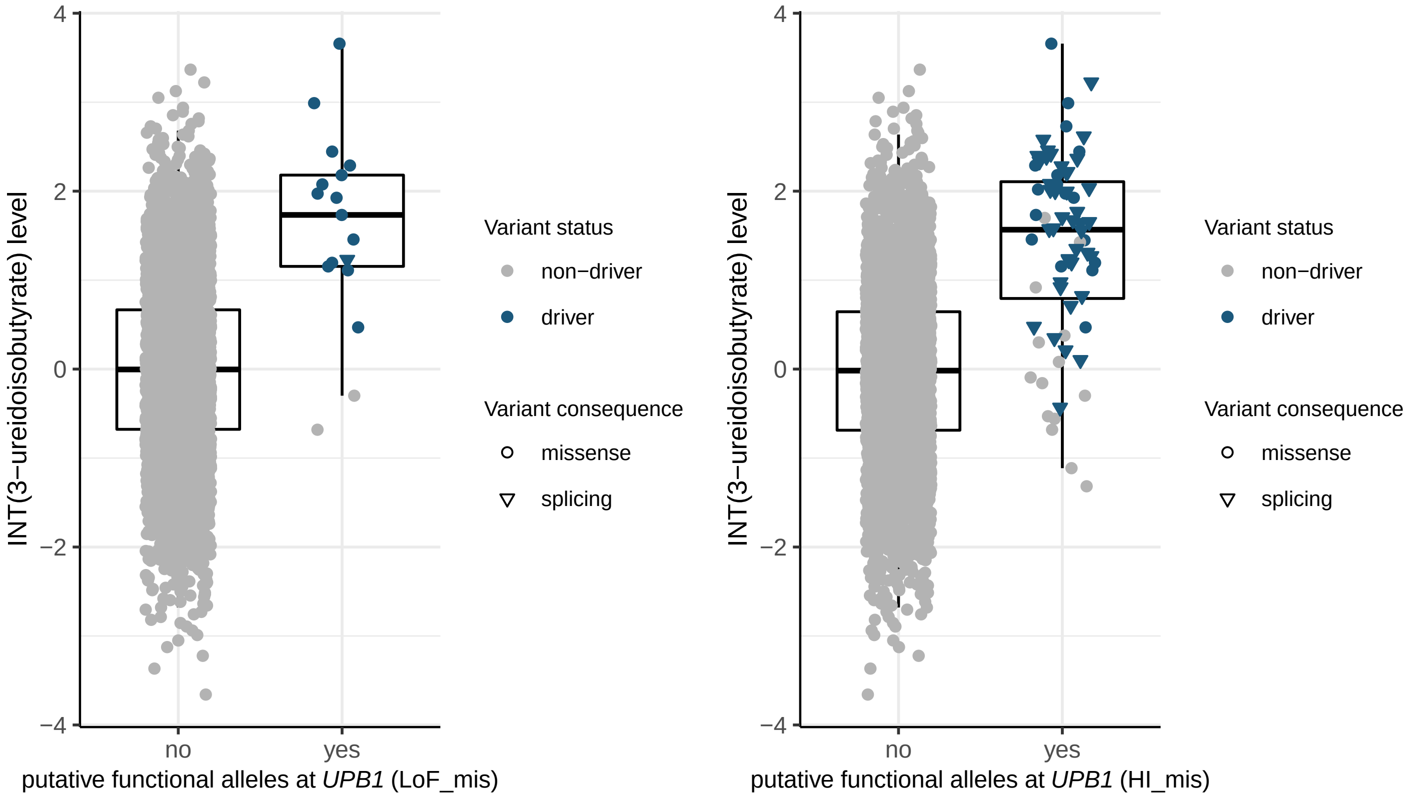

INT(3−ureidoisobutyrate) level
4
2
0
−2
−4
no
yes
putative functional alleles at UPB1 (LoF_mis)
Variant status
non−driver
driver
Variant consequence
missense
splicing
INT(3−ureidoisobutyrate) level
putative functional alleles at UPB1 (HI_mis)
